# Effects and predictive performance of multilayer environmental exposures on coccidioidomycosis: a longitudinal surveillance study

**DOI:** 10.64898/2026.09.12.26362891

**Authors:** Qianqian Li, Yue Zhan, Haiyue Li, Runqiu Wang, Jesse E. Bell

## Abstract

Coccidioidomycosis (Valley fever) is a soilborne mycosis endemic to the US Southwest whose incidence has increased markedly in recent decades. Environmental conditions are thought to influence the soil-dwelling lifecycle of *Coccidioides*; however, most prior studies have relied on above-ground meteorological conditions–primarily precipitation and air temperature (AT)–with few examining subsurface soil moisture (SM) and soil temperature (ST), which may more directly influence the fungal lifecycle. No study of coccidioidomycosis, or any other environment-sensitive soilborne mycosis, has examined how deeper-layer soil conditions relate to disease incidence, despite the prevailing soil-sterilisation hypothesis implicating deeper soil as a potential fungal refugium. Furthermore, nonlinear exposure–lag–response relationships for key dust-dispersion exposures–including PM_10_, a potential proxy for airborne spore concentration, and wind speed–remain uncharacterised. We aimed to estimate and compare the associations between coccidioidomycosis incidence and environmental exposures across multiple above- and below-ground layers, and to evaluate their independent and combined predictive performance. This ecological time-series study analysed 185,486 reported cases of coccidioidomycosis in Arizona’s hyperendemic tri-county region (Maricopa, Pima, and Pinal) during 1997–2024. We developed a mechanism-informed multilayer environmental framework comprising one dust-dispersion layer (PM_10_, wind speed) and four soil–climate layers–meteorological (precipitation, AT), topsoil (0–10 cm), midsoil (10–40 cm), and deepsoil (40–100 cm) SM and ST. We fitted distributed lag non-linear models (DLNMs) with season-specific interaction terms to estimate exposure–lag–response associations between each environmental layer and coccidioidomycosis incidence. We then developed a two-stage stacked ensemble machine learning framework to assess each layer’s independent predictive performance (stage 1) and integrate them into a unified forecast (stage 2), which was evaluated using a strictly held-out test period. At concurrent lags (1–3 months prior to reporting), coccidioidomycosis incidence was primarily associated with dustier, windier, and drier conditions, cooler air temperatures, and warmer topsoil. For each IQR increase, PM_10_ showed the most consistent concurrent associations, with significant positive incidence rate ratios (IRRs) across all four incidence seasons at lags 1–2 (ranging from 1.04 [95% CI 1.00–1.08] to 1.37 [1.26–1.48]). Across lags 1–36 months, all four soil–climate layers exhibited nonlinear, non-monotonic, and season-dependent associations with coccidioidomycosis incidence, characterised by alternating wet–dry and cool–warm oscillations. Topsoil displayed the most frequent significant associations, with moisture–temperature signals attenuating progressively from topsoil through midsoil to deepsoil. A depth-dependent lag structure was observed for both moisture and temperature, in which significant positive IRRs emerged at progressively shorter lags with increasing soil depth, accompanied by vertical divergence across depths at the same lag windows. For example, for fall incidence, positive moisture IRRs appeared at precipitation lag 15 (1.10 [1.00–1.21]), topsoil SM lag 9 (1.22 [1.15–1.30]), midsoil SM lags 8–9 (up to 1.23 [1.11–1.37]), and deepsoil SM lags 4–5 (up to 1.08 [1.01–1.16]); at these same lags, deepsoil SM was positively associated with incidence whereas topsoil SM and precipitation remained negatively associated. During the held-out test period (2021–2024), the multilayer ensemble generally captured seasonal and interannual variation well, including the timing and approximate magnitude of most peaks, outperforming all single-layer models. Although individual layers had slightly lower test RMSEs, their test gap ratios were substantially higher (0.27–0.67 vs 0.00), indicating that the ensemble generalised far more reliably. All five environmental layers contributed to the final ensemble forecast; the dust-dispersion and topsoil layers received the highest importance, with PM_10_ ranked as the most important predictor group. The best-performing of four pipeline configurations relied solely on environmental inputs available within one week, enabling the model to function as a near-real-time nowcast. This study provides the first evidence linking multilayer environmental exposures to coccidioidomycosis incidence across both temporal and vertical dimensions, offering new quantitative support for the prevailing soil-sterilisation and grow-and-blow hypotheses and demonstrating that a multilayer framework could improve both mechanistic understanding and predictive performance. The framework could be generalised to other endemic settings and readily extended with new data and methods to inform surveillance and public health preparedness. These findings support incorporating multilayer lagged environmental exposures into both effect estimation and forecasting systems to better prepare endemic regions for anticipated warming, drying, and increasingly variable climatic conditions.

**Research in context:** *Evidence before this study:* We searched PubMed, Scopus, and Google Scholar for literature published in English from database inception to July 17, 2026. The first search combined environmental exposure terms (“environment*” OR “climat*” OR “meteorolog*” OR “weather” OR “soil moisture” OR “temperature” OR “precipitation” OR “rain” OR “humidity” OR “moisture” OR “drought” OR “wind” OR “dust” OR “particulate matter” OR “PM10” OR “PM2.5”) with coccidioidomycosis terms (“coccidioidomycosis” OR “Valley fever” OR “*Coccidioides*”). The second search used the same environmental terms combined with broader soilborne mycosis terms (“soilborne mycosis” OR “soilborne mycoses” OR “endemic mycosis” OR “endemic mycoses” OR “histoplasmosis” OR “blastomycosis” OR “paracoccidioidomycosis” OR “coccidioidomycosis” OR “Valley fever” OR “*Coccidioides*”). Prior studies have generally supported the theory that antecedent alternating wet–dry and cool–warm periods increase coccidioidomycosis incidence, yet most relied on above-ground meteorological variables—primarily precipitation and air temperature—that might not adequately reflect subsurface conditions where the fungus grows. A few studies incorporated soil moisture data, but relied on correlation or univariate analyses without adjusting for potential confounders. Our recent study was the first to assess topsoil (0–10 cm) moisture and temperature effects on coccidioidomycosis incidence within a multivariable framework, but was restricted to topsoil layer and linear modelling, leaving nonlinear exposure–lag–response relationships and the potential role of deeper soil layers unexplored. Critically, no study in coccidioidomycosis—or any other environment-sensitive soilborne mycosis—has examined how deeper-layer (>10 cm) soil conditions relate to disease incidence, even though the soil-sterilisation hypothesis suggests that deeper soils might serve as fungal refugia. Nor has any study characterised the nonlinear exposure–lag–response relationship between PM_10_, a potential proxy for airborne spore concentration, and coccidioidomycosis incidence. Finally, no study has evaluated or compared the effects and predictive performance of multilayer environmental exposures for coccidioidomycosis or other environmentally sensitive soilborne mycoses.

*Added value of this study:* To our knowledge, this is the first study to use a comprehensive multilayer environmental framework—spanning one dust-dispersion and four soil–climate layers (meteorological, topsoil, midsoil, and deepsoil)—to any soilborne mycosis. Within this framework, we used DLNMs and a novel two-stage stacked ensemble approach to assess, for the first time, both the associations and predictive performance of environmental exposures across all five layers. This enabled the first characterisation of nonlinear exposure–lag–response relationships for PM_10_ and subsurface SM and ST in coccidioidomycosis research. We found that increased incidence was generally associated with concurrent dustier, windier, and drier conditions, cooler air temperatures, and warmer topsoil, preceded by alternating wet–dry and cool–warm oscillations across layers. PM_10_ exhibited consistent positive associations with incidence at concurrent lags across all four seasons and emerged as the most important predictor in the ensemble forecast, jointly providing the first support for the recently proposed dust-borne atmospheric transport hypothesis. The multilayer design extended evidence of alternating wet–dry and cool–warm cycles to midsoil and deepsoil for the first time, although this cyclical signal was most pronounced in the topsoil and attenuated progressively with depth. Across moisture and temperature variables, we identified depth-dependent lag structures in which significant positive associations appeared at progressively shorter lags with increasing soil depth, accompanied by vertical divergence across layers at the same lag windows—providing the first quantitative evidence in the vertical dimension for both the dominant soil-sterilisation and grow-and-blow hypotheses. These patterns suggest that deeper soils might function as a buffered subsurface refugium for *Coccidioides*, preserving favourable moisture and thermal conditions longer than shallower layers. Our two-stage ensemble framework demonstrated that combining multilayer environmental information achieved superior prediction over any single-layer approach. The selected pipeline—relying solely on environmental data available within one week of the target month—could serve as a near-real-time nowcast, generating incidence estimates well before finalised surveillance data become available. Our ensemble framework offers a flexible, modular architecture that can be readily extended with new predictors and candidate models to further refine forecasting performance. Collectively, these results offer the first evidence connecting multilayer environmental exposures to coccidioidomycosis incidence across both temporal and vertical dimensions, provide new quantitative support for the prevailing mechanistic hypotheses, and show that a multilayer approach could enhance both mechanistic understanding and predictive performance.

*Implications of all the available evidence:* Our results suggest that coccidioidomycosis dynamics might be associated with complex, depth-stratified hydroclimatic cycles that above-ground environmental data alone cannot fully capture, highlighting the potential value of incorporating subsurface soil data into environmental health studies of soilborne mycoses more broadly. Although the best-performing candidate models, the predictive performance of individual layers, and their relative contributions within the ensemble might vary across endemic settings, the approach used by most existing forecasting studies—relying on above-ground meteorological or dust-related variables alone—is unlikely to be sufficient. The primary contribution of this work might lie less in any particular set of candidate models or predictors than in the multilayer framework itself, which provides a modular and extensible architecture for integrating heterogeneous environmental information across vertical and temporal dimensions. The selected prediction pipeline—relying solely on environmental inputs available within one week—could function as a near-real-time nowcast, generating incidence estimates well before many contemporaneously exposed patients were diagnosed given the prolonged diagnostic pathway for coccidioidomycosis. Nowcast-identified high-incidence periods could support public health preparedness by prompting earlier clinical consideration and targeted patient counselling—particularly as patients with prior awareness of coccidioidomycosis have been shown to be diagnosed substantially earlier and to seek testing more proactively. Importantly, the modular architecture is readily extensible with forecast-derived predictors, enabling a shift from nowcasting to prospective early-warning forecasting. The evidence, including findings from this study, suggests that anticipated climatic changes in the southwestern USA—including intensifying drought, continued warming, and potentially increasing dust emissions—might escalate coccidioidomycosis burden and expand its endemic range, underscoring the need for improved surveillance and forecasting tools. Future efforts in coccidioidomycosis surveillance, effect estimation, and prediction might benefit from adopting and refining this multilayer framework and evaluating its applicability in other endemic settings and, potentially, in other environment-sensitive soilborne mycoses.

## Introduction

Coccidioidomycosis (Valley fever) is an emerging and increasingly important fungal disease caused by inhalation of airborne arthroconidia (spores) from soil-dwelling *Coccidioides* spp. Infection can cause prolonged respiratory illness and may progress to chronic (5–10% of cases) or disseminated disease (0⋅5–2%).^1,2^ Coccidioidomycosis is most frequently reported in the US Southwest, with the greatest burden concentrated in Arizona and California. From 1998 to 2023, annual reported cases in the USA increased more than ninefold, with Arizona’s cases surging from 1474 to 10,990 and accounting for 61% of nearly 300,000 nationwide cases—almost double California’s 36%.^3,4^ Approximately 95% of Arizona’s cases concentrate in the hyperendemic southcentral counties of Maricopa, Pima, and Pinal, where coccidioidomycosis has become a leading cause of community-acquired pneumonia, imposing substantial morbidity and economic burden.^1,5,6^ The absence of an effective vaccine or definitive treatment, combined with diagnostic challenges, makes coccidioidomycosis an essential public health concern in endemic regions.^1,2^ Growing evidence links this escalating burden to changing climate conditions in the Southwest, including rising temperatures, intensifying drought, and increasing dust emissions.^7–13^

Environmental drivers of coccidioidomycosis are thought to act by influencing the fungal lifecycle. Two prevailing mechanistic hypotheses guide current understanding. The dominant “grow-and-blow” hypothesis proposes that adequate moisture and favorable temperatures support *Coccidioides* hyphal growth, while subsequent hot, dry conditions promote hyphal desiccation and fragmentation into infectious arthroconidia that become airborne when soils are disturbed by wind erosion or other mechanisms, leading to dust-mediated dispersal and inhalational exposure.^14–17^ Another widely accepted “soil sterilization” hypothesis suggests that *Coccidioides*, as a filamentous organism, survives extreme hot and/or dry surface conditions by extending hyphae into deeper, cooler, and moister soil layers to evade non-filamentous microbial competitors, subsequently thriving in the topsoil with reduced competition when favorable conditions return.^15,18^

These two hypotheses imply that subsurface soil moisture (SM) and soil temperature (ST) influence *Coccidioides* lifecycle, yet most prior studies have relied on meteorological variables to evaluate climatic effects on coccidioidomycosis incidence—primarily precipitation and air temperature (AT)—that might not adequately reflect soil moisture-temperature conditions where the fungus grows.^3^ Only a few studies have incorporated SM data,^9,19^ but relied on correlation or univariate analyses without controlling for confounding from concurrent and lagged covariates. Our recent study was the first to evaluate concurrent and lagged effects of topsoil moisture and temperature on coccidioidomycosis incidence within a multivariable framework, identifying significant associations across incidence seasons.^3^ However, that analysis was confined to the 0–10 cm topsoil layer and employed linear models, leaving nonlinear exposure–response relationships and the role of deeper soil layers unexplored. Critically, no study—in coccidioidomycosis or other environment-sensitive soilborne mycosis—has investigated how deeper-layer soil conditions relate to disease incidence, despite the soil sterilization hypothesis implicating deeper layers as refugia. Additionally, existing dust–incidence analyses have used PM_10_ in linear models or applied distributed lag non-linear models (DLNMs) to PM_2.5_.^3,8,14,17^ The nonlinear exposure–lag–response relationship between PM_10_—which may be a superior proxy for airborne arthroconidia concentrations given the 2–4 µm spore diameter—and incidence remains uncharacterized. Finally, no study has compared the predictive performance of these multilayer environmental predictors or evaluated how they can be optimally combined to forecast coccidioidomycosis incidence.

To address these gaps, we developed a mechanism-informed multilayer environmental framework grounded in the two dominant hypotheses, spanning four soil–climate layers—meteorological (precipitation, AT), topsoil (0–10 cm), midsoil (10–40 cm), and deepsoil (40–100 cm) SM and ST—and one dust-dispersion layer (PM_10_, wind speed), designed to comprehensively capture above- and below-ground conditions influencing *Coccidioides* growth and dust-borne dispersal. Using this framework, we aimed to (1) estimate and compare associations between coccidioidomycosis incidence and environmental factors across five layers, providing the first nonlinear exposure–lag–response characterization for topsoil and dust-dispersion factors in coccidioidomycosis research and the first investigation of deeper-layer hydroclimatic effects in soilborne mycosis research; and (2) predict coccidioidomycosis incidence using a novel two-stage ensemble machine learning approach developed in this study that evaluates the independent predictive performance of each environmental layer (stage 1) and quantifies their relative contributions to the final ensemble prediction (stage 2). Together, these analyses aim to generate new evidence and insights that advance understanding of the two dominant hypotheses, and to inform the development of future coccidioidomycosis forecasting models.

## Methods

### Coccidioidomycosis data

This multi-county ecological time-series study used monthly county-level coccidioidomycosis surveillance data (by county of residence) in Arizona from January 1997 to December 2024, obtained from the Arizona Department of Health Services (ADHS; appendix table S1, p 2). Cases were recorded by calendar month and year of report date (ie, the date of laboratory reporting to ADHS). A confirmed case was defined by laboratory evidence alone (effective since 2008; prior to 2008, both clinical and laboratory criteria were required), with each case reported only once per person unless reinfection by a distinct *Coccidioides* strain is confirmed through whole genome sequencing.^4^ Since 1997, all healthcare providers and laboratories in Arizona have been required to report confirmed coccidioidomycosis cases to ADHS within five working days of laboratory confirmation.^4^ County-level annual mid-year population estimates for 1997–2024, obtained from the U.S. Census Bureau,^20^ were linearly interpolated to monthly values to calculate county-level monthly incidence per 100,000 population. To minimize exposure misclassification from travel-related cases, we restricted analyses to the hyperendemic tri-county region—Maricopa, Pima, and Pinal counties—which accounted for approximately 95% of Arizona’s reported cases during the 28-year study period (appendix figure S1, p 18).

### Environmental exposure data

We compiled monthly environmental exposure data from January 1994 through December 2024 to accommodate distributed lag structures of up to 36 months before the first disease observation in January 1997 (appendix table S1, p 2). We then organized these data into five layers to comprehensively capture above- and below-ground conditions influencing *Coccidioides* growth and dispersal: four soil–climate layers (meteorological, topsoil [0–10 cm], midsoil [10–40 cm], and deepsoil [40–100 cm]) and one dust-dispersion layer. Full details are provided in the appendix (text S1). For the meteorological layer, we obtained monthly, 4 km gridded precipitation and air temperature from the parameter-elevation regressions on independent slopes model (PRISM).^21^ Building on our previous work,^3^ which examined only topsoil SM and ST from the North American Land Data Assimilation System Phase 2 (NLDAS-2) gridded data, the present study extended the soil–climate exposures to three depth layers at 0.125° spatial resolution.^22^ The dust-dispersion layer included wind speed from the NLDAS-2 primary forcing data,^23^ and daily mean PM_10_ concentration from the US Environmental Protection Agency (EPA) air quality monitoring data.^24^ A dusty-day indicator, defined as the number of days per month with PM_10_ exceeding 45 µg/m³ at any monitoring site within a county, was additionally derived to enable descriptive comparison with our previous findings.^3^ All gridded data were spatially aggregated to county-level monthly means, and site-level PM_10_ measurements were averaged across monitoring sites within each county by month. The integrated multi-source environmental exposure data were then linked to the corresponding county-month case data for analysis.

### Data processing

#### Surveillance bias adjustment

Arizona’s coccidioidomycosis trends were influenced by changes in case definitions, laboratory reporting, and testing practices over the study period,^25^ creating surveillance artifacts unrelated to true disease incidence (appendix figure S2, p 19). We addressed these artifacts using a two-part approach. First, for January–March 2024, when direct quantitative evidence of data quality issues was available, we applied a numeric correction by proportionally redistributing approximately 800 false-positive cases across county-month records during this period. Second, we created a categorical surveillance regime indicator corresponding to six discrete periods to account for major surveillance changes documented in the ADHS *Coccidioidomycosis Surveillance Data Caveats*.^25^ This variable was included as a covariate to absorb abrupt, period-specific baseline shifts in reported case counts. Gradual secular trends (eg, evolving public health awareness) were additionally captured via a natural spline of year. Full details of surveillance regime definitions and transition dates are provided in the appendix (figure S2, p 19).

#### Missing data

Only one variable, PM_10_, had missing values (∼8.6% of observations, concentrated before 2002); all other variables were complete (appendix table S2, pp 3–4). Missing values were imputed using a seasonal autoregressive integrated moving average (ARIMA) model fitted separately per county, preserving the county-specific temporal structure of the series.^26^

#### Seasonal grouping

Following our earlier study,^3^ months were grouped into four seasons: winter (January–March), spring (April–June), monsoon (July–September), and fall (October–December). This grouping aligns with the distinct monsoon climatology of Arizona’s tri-county region, partitions the year into alternating wet and dry periods, and enables direct comparison of results across the two studies.

### Statistical analysis

#### Distributed lag non-linear models

To quantify the exposure–lag–response associations between multilayer environmental exposures and coccidioidomycosis incidence, we fitted DLNMs within a negative binomial regression framework across five layers.^27^ A separate model was fitted for each of five layers, each containing two exposure variables. For *k*th layer, the number of cases *Y_i_*_,*t*_ in county *i* in month *t* was modeled as:

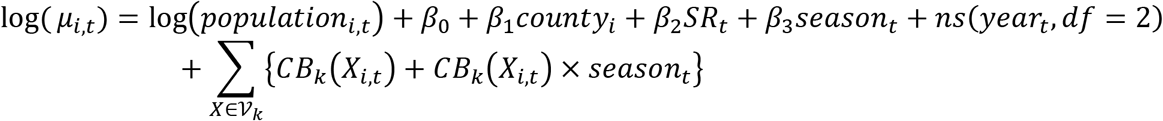

where *μ_i_*_,*t*_ = *E*(*Y_i_*_,*t*_). Monthly county-level case counts served as the outcome, with the log of county population as an offset so that coefficients are interpretable as log incidence rate ratios (IRRs). The five layers were: L_1_ (dust dispersion): V_1_ = {*PM*_10_, *wind speed*}, L_2_ (meteorological): V_2_ = {*precipitation*, *AT*}, L_3_ (topsoil 0–10 cm): V_3_ = {*SM*_0*t*10_, *ST*_0*t*10_}, L_4_ (midsoil 10–40 cm): V_4_ = {*SM*_10*t*40_, *ST*_10*t*40_}, and L_5_ (deepsoil 40–100 cm): V_5_ = {*SM*_40*t*100_, *ST*_40*t*100_}. *CB_k_*(*X_i_*_,*t*_) denotes a DLNM cross-basis for exposure *X* in layer *k*, constructed using natural cubic splines for both the exposure–response and lag–response dimensions. Lags ranged from 1 to *L_k_* months prior to incidence month (*L*_1_ = 6; *L*_2_ = ⋯ = *L*_5_ = 36). Equally spaced knots were used for all splines, with the number of knots selected via grid search minimizing the Akaike information criterion.

Models adjusted for county (absorbing between-county spatial differences), surveillance regime (SR; absorbing documented abrupt changes in surveillance practices), and a natural spline of year (capturing gradual secular trends unrelated to environmental exposures such as evolving clinical awareness and diagnostic practices). Because environmental–disease associations might vary by season, we included interaction terms between season and each exposure cross-basis, allowing season-specific exposure–lag–response estimation. Season was thus included as both a confounder (main effect) and an effect modifier.

We estimated associations in two ways: (1) lag–response IRRs for an interquartile-range (IQR) increase in each exposure at each lag, using lag-specific 25th (reference) and 75th percentiles calculated from the exposure months corresponding to each lag;^7,27^ and (2) lag-specific exposure–response curves across the exposure range, expressed as IRRs relative to the lag-specific 25th percentile.

#### Two-stage ensemble machine learning models

We developed a two-stage stacked ensemble machine learning framework to predict coccidioidomycosis incidence, designed to evaluate the independent predictive performance of each environmental layer (stage 1) and optimally combine them into a unified prediction (stage 2). Full details are provided in the appendix (figure S57, pp 70–71).

In stage 1, five base learners were trained independently—one per environmental layer (L_1_–L_5_)—using candidate model types (generalized linear model with negative binomial distribution [GLM.nb], random forest [RF], and extreme gradient boosting [XGBoost]). Environmental predictors were represented as smoothed lags (Slags): 3-month moving averages at lag positions 1, 3, …, 36 months for L_2_–L_5_ and 1, 3, 6 for L_1_. Each base learner was selected via 7-fold leave-one-year-out (LOYO) cross-validation (CV; walk-forward, validation years 2014–2020). In stage 2, a meta-learner (constrained least squares [CLS], RF, or XGBoost) combined the five out-of-fold (OOF) predictions (ŷ_L1_–ŷ_L5_) into a final ensemble forecast, selected via 3-fold expanding-window LOYO CV (validation years 2018–2020). At both stages, models were selected based on near-best recency-weighted median validation root mean square error (RMSE; within 5% of minimum), with tiebreakers prioritizing generalisation (appendix figure S57, pp 70–71).

Four pipeline configurations were compared: (1) smoothed-lag environmental features only; (2) raw monthly environmental lags, with DLNM as an additional stage 1 candidate; (3) smoothed environmental lags augmented with autoregressive incidence lags at 1, 12, 24, and 36 months; and (4) smoothed environmental lags augmented with incidence lags at 12, 24, and 36 months only. Models were trained on 1997–2020 and evaluated on held-out test data (2021–2024).The winning pipeline was reported as the primary result; the remaining configurations served as sensitivity analyses.

Feature importance was assessed for the winning pipeline at three levels—within-layer predictor group, layer, and global—using TreeSHAP and grouped permutation methods.

Analyses were done in R (version 4.5.2).

## Results

From 1997–2024, Arizona reported 195,380 coccidioidomycosis cases; the tri-county study region accounted for 185,486 (95%), including Maricopa (145,090), Pima (26,700), and Pinal (13,696). Mean annual incidence was highest in Maricopa and Pinal (both 129 cases per 100,000), followed by Pima (97 cases). Spatially, the tri-county region was characterized by warmer, drier, and dustier conditions than northern Arizona (figure 1).

**Figure 1:**
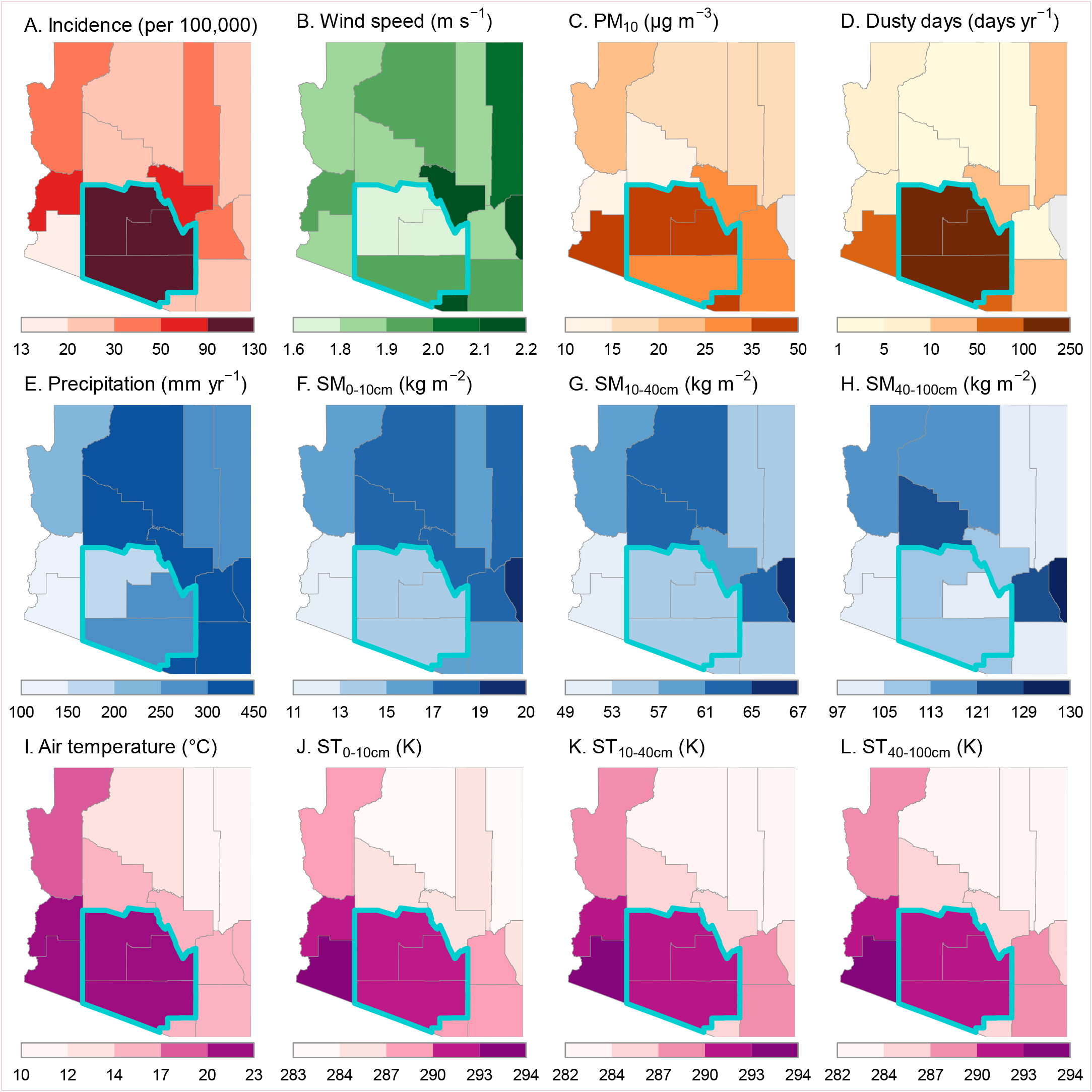
Mean annual spatial patterns of coccidioidomycosis incidence and multilayer environmental drivers in Arizona. (A) Mean annual coccidioidomycosis incidence (per 100,000 population), 1997–2024. (B–L) Mean annual multilayer environmental drivers, 1994–2024, including aboveground variables (B–E, I) and belowground variables (F–H, J–L): (B) wind speed at 10 m above ground, (C) PM_10_ concentration, (D) dusty days (annual number of days with daily mean PM_10_ > 45 µg m⁻³ at any monitoring site within the county),^3^ (E) total annual precipitation, (F–H) soil moisture at 0–10 cm, 10–40 cm, and 40–100 cm depths, (I) air temperature, and (J–L) soil temperature at 0–10 cm, 10–40 cm, and 40–100 cm depths. The tri-county study region (Maricopa, Pima, and Pinal counties) is outlined in cyan and was used for all analyses. Gray shading in panels C and D indicates missing PM_10_ monitoring data (Greenlee County).

Monthly incidence displayed a bimodal seasonal pattern, with a primary peak in November–December and a secondary peak in July–August (figure 2A). Over the study period, incidence surged during 2009–2012 and rose gradually from 2013 through 2024 (appendix figure S3A, p 20). Environmental exposures exhibited pronounced seasonality: precipitation peaked during the monsoon and was lowest in spring, air and soil temperatures peaked in the monsoon, soil moisture generally peaked in winter with topsoil and midsoil additionally showing a secondary monsoon peak, and PM_10_ (and dusty-day counts) were lowest in winter and highest in spring (figure 2; appendix table S2, pp 3–4, and figure S3, p 20).

**Figure 2:**
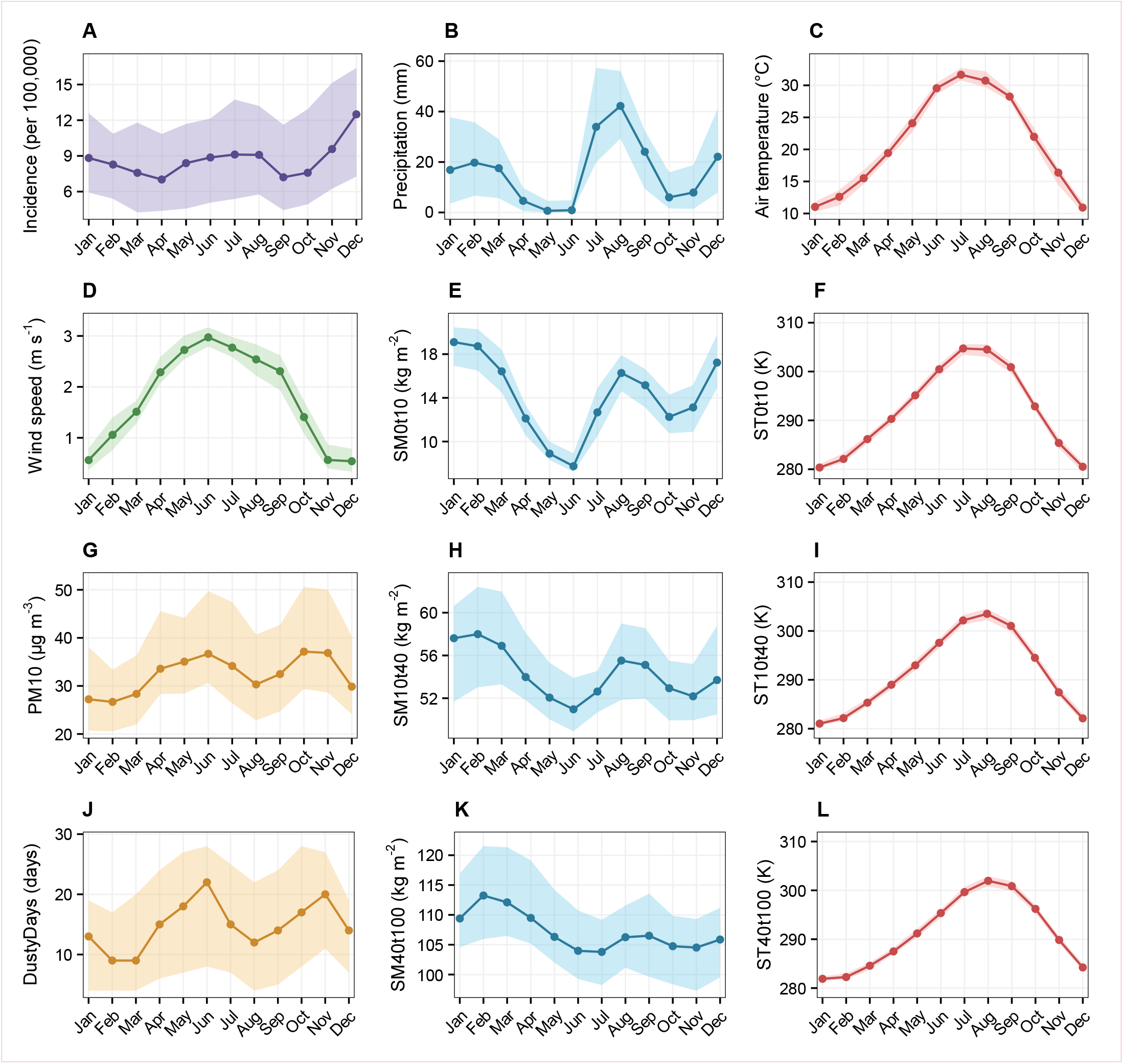
Monthly patterns of coccidioidomycosis incidence and multilayer environmental exposures. Monthly medians (lines) and interquartile ranges (IQRs: 25th–75th percentiles; shaded areas) were calculated at the county-month level in the study region. (A) Coccidioidomycosis incidence per 100,000 population (1997–2024); (B–L) environmental variables (1994–2024): precipitation (B), air temperature (C), wind speed (D), soil moisture at 0–10 cm (E), 10–40 cm (H), and 40–100 cm (K) depths, soil temperature at 0–10 cm (F), 10–40 cm (I), and 40–100 cm (L) depths, PM_10_ (G), and dusty days (J; number of days per month with PM_10_ > 45 µg m⁻³ at any monitoring site within the county).

We estimated lag–response IRRs for an IQR increase in each exposure across all five environmental layers and four incidence seasons using DLNMs (figure 3; appendix figures S4–S16, pp 21–33, and tables S3–S7, pp 5–13). Corresponding lag-specific exposure–response curves were predominantly non-linear, with shapes varying substantially across lags, seasons, and layers (appendix figures S17–S56, pp 34–69).

**Figure 3:**
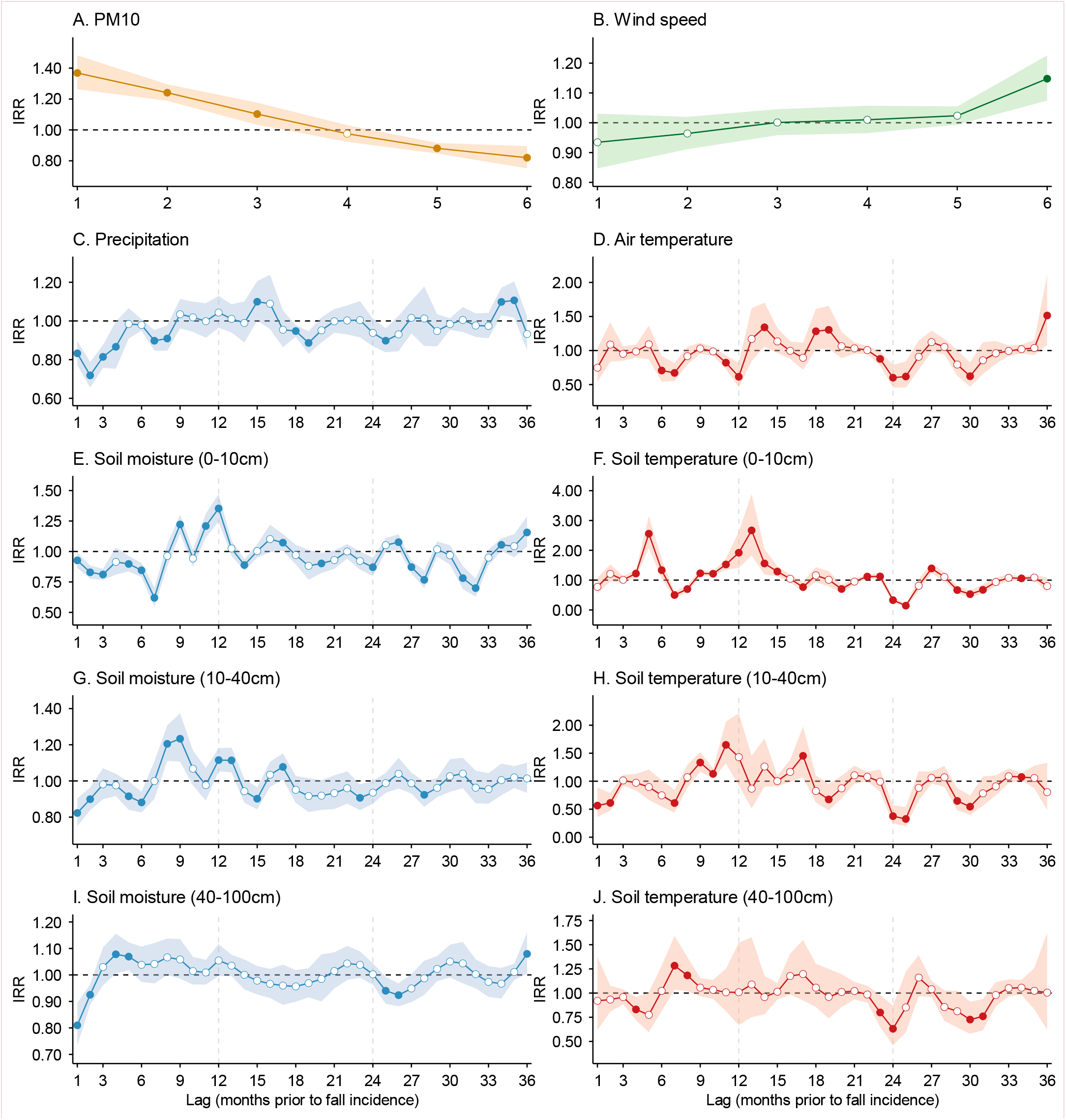
Associations between fall (October–December) coccidioidomycosis incidence and lagged multilayer environmental exposures in the study region. Incidence rate ratios (IRRs) and 95% confidence intervals (shaded areas) were estimated using distributed lag non-linear models (DLNMs) for an IQR increase in each exposure at specific lag months prior to disease incidence. Panels show above-ground exposures from the dust dispersion layer (layer 1; A: PM_10_; B: 10-m wind speed; lags 1–6 months) and meteorological layer (layer 2; C: precipitation; D: air temperature; lags 1–36 months), and below-ground exposures of soil moisture (E, G, I) and soil temperature (F, H, J) at depths of 0–10 cm (layer 3), 10–40 cm (layer 4), and 40–100 cm (layer 5), with lags of 1–36 months for layers 3–5. Models were adjusted for spatiotemporal trends and surveillance-related changes in reporting and laboratory testing. The horizontal dashed line indicates null association (IRR = 1). Solid points indicate statistically significant associations; hollow points indicate non-significant associations. Corresponding numerical data are reported in the appendix (tables S3–S7, pp 5–13). Associations for other seasons are shown in the appendix (figures S4–S6, pp 21–23).

Because delays from spore exposure to case reporting can span weeks to months,^25^ short-term lags (1–3 months prior to reporting) were considered as the concurrent period of exposure. Overall, concurrent-window increases in coccidioidomycosis incidence were predominantly associated with dustier, windier, and drier conditions, alongside cooler air temperatures and generally warmer topsoil. In contrast, associations for mid- and deep soil temperatures were heterogeneous across seasons. PM_10_ (L1) showed the most consistent associations, with significant positive IRRs across all four seasons at lags 1–2 (ranging from 1.04 [95% CI 1.00–1.08] to 1.37 [1.26–1.48]), strongest in fall (appendix table S7, p 13).

Wind speed (L1) was positively associated with incidence during spring and monsoon at lags 1–2 (1.04 [1.01–1.07] to 1.10 [1.04–1.17]). Drier conditions were associated with increased incidence for both precipitation (L2; 0.71 [0.60–0.84] to 0.89 [0.81–0.98], significant in all four seasons at lag 2) and topsoil soil moisture (L3; 0.72 [0.63–0.83] to 0.93 [0.87–0.99], significant in all four seasons at lag 1; appendix table S3, pp 5–6). Warmer topsoil temperature (L3) was predominantly associated with increased incidence during spring and monsoon (1.21 [1.10–1.33] to 1.43 [1.14–1.79]). Air temperature (L2) showed significant negative associations during winter and monsoon at lags 2–3 (0.56 [0.42–0.76] to 0.78 [0.63–0.99]).

Across lags 1–36 months, soil-climate associations (L2–L5) with coccidioidomycosis incidence were non-linear, non-monotonic and season-dependent, characterised by recurring wet–dry and cool–warm oscillations rather than a single monotonic lag structure (figure 3; appendix figures S4–S6, pp 21–23, and tables S3–S6, pp 5–12). Topsoil (L3) exhibited the most frequent significant associations, followed by the meteorological layer (L2), with moisture–temperature signals attenuating from topsoil through midsoil (L4) to deepsoil (L5). Temperature yielded more significant lags than moisture for L2–L3, whereas L4–L5 showed approximately equal numbers of significant lags for both variables. Across all eight exposures (four moisture and four temperature variables), significant negative lags generally outnumbered significant positive lags, with topsoil ST being the sole exception—showing more significant positive than negative lags across all four seasons.

For precipitation (L2), drier conditions were associated with increased incidence at short lags across all four seasons (lags 1–4; 0.71 [95% CI 0.60–0.84] in monsoon to 0.89 [0.81–0.98] in spring; appendix table S6, pp 11–12), reversing to positive associations with antecedent wetter conditions at longer lags with season-specific timing (e.g., winter lags 14–15 and 31–32; spring lags 28–29; monsoon lags 22 and 33; fall lags 15 and 34–35), forming a wet-then-dry temporal sequence preceding case reporting. Air temperature (L2) showed more negative significant lags than positive across all seasons, with the direction and timing of significant associations varying by season (appendix figure S14, p 31).

Topsoil SM (L3) displayed the clearest wet–dry oscillation among the four moisture variables, particularly in fall: negative associations at lags 1–7 (0.62 [0.54–0.70] at lag 7) reversing to positive associations at lags 9–12 (1.35 [1.24–1.47] at lag 12; appendix table S3, pp 5–6). All four seasons showed significant negative associations at lag 1, ranging from 0.72 (0.63–0.83) in monsoon to 0.93 (0.87–0.99) in fall. Topsoil ST (L3) showed the most prominent temperature associations across L3–L5, with the most pronounced alternating cool–warm cycles, the largest positive effects across all four seasons (fall lag 13: 2.67 [1.83–3.89]; spring lag 8: 2.35 [1.76–3.13]; monsoon lag 10: 2.31 [1.53–3.49]; winter lag 15: 1.70 [1.26–2.31]), and the largest negative effects in three of four seasons (fall lag 25: 0.14 [0.10–0.20]; spring lag 19: 0.14 [0.09–0.22]; winter lag 28: 0.29 [0.18–0.48]).

Midsoil SM (L4) followed a similar but attenuated dry–wet pattern: negative IRRs at short lags (lags 1–2: 0.80 [0.70–0.91] to 0.90 [0.84–0.96]), turning positive during monsoon at lags 6–7 (up to 1.31 [1.16–1.49]; appendix table S4, pp 7–8). Midsoil ST (L4) showed an attenuated cool–warm pattern with season-dependent associations, including the largest negative temperature effect in monsoon across L3–L5 (lag 22: 0.29 [0.16–0.53]).

Deepsoil (L5) associations were further attenuated. Deepsoil SM showed negative associations at short lags (lags 1–2: 0.81 [0.73–0.89] to 0.93 [0.89–0.96]) followed by brief positive reversals (monsoon lags 3–5: up to 1.19 [1.10–1.29]; fall lags 3–4: up to 1.08 [1.01–1.16]). Similarly, deepsoil ST showed the weakest cool–warm oscillations across L2–L5.

A notable finding across moisture and temperature variables (L2–L5) was a consistent depth-dependent lag structure, in which significant positive associations with coccidioidomycosis incidence appeared at progressively shorter lags with increasing depth, accompanied by vertical divergence in the direction of associations across depths at the same lag windows (figure 3; appendix figures S4–S6, pp 21–23, and tables S3–S6, pp 5–12).

For fall incidence, significant positive moisture associations emerged at progressively shorter lags with increasing depth: precipitation at lag 15 (1.10 [1.00–1.21]), topsoil SM at lag 9 (1.22 [1.15–1.30]), midsoil SM at lags 8–9 (1.21 [1.11–1.31] and 1.23 [1.11–1.37]), and deepsoil SM at lags 4–5 (1.08 [1.01–1.16] and 1.07 [1.02–1.12]). At the same lag window of 4–5 months, deepsoil SM was positively associated with incidence, whereas topsoil SM and precipitation were negatively associated. A comparable pattern was observed for monsoon incidence: precipitation at lag 8 (1.09 [1.00–1.19]), topsoil SM at lags 7–8 (up to 1.22 [1.12–1.34]), midsoil SM at lags 6–7 (up to 1.31 [1.16–1.49]), and deepsoil SM at lags 3–5 (up to 1.19 [1.10–1.29]). At lag 5, topsoil SM was significantly negative (0.87 [0.78–0.97]). For winter incidence, significant positive moisture associations appeared at precipitation at lags 14–15 (up to 1.16 [1.05–1.27]), topsoil SM at lag 12 (1.14 [1.06–1.22]), midsoil SM at lags 11–12 (up to 1.20 [1.06–1.36]), and deepsoil SM at lags 8–9 (up to 1.12 [1.04–1.22]). At lags 8–9, both topsoil and midsoil SM were significantly negative (topsoil: 0.84 [0.77–0.91] and 0.85 [0.78–0.93]; midsoil: 0.88 [0.82–0.95] and 0.79 [0.72–0.87]). These consistent patterns across three seasons revealed the depth-dependent moisture gradient with clear vertical divergence.

Temperature associations showed a comparable depth-dependent pattern. For fall incidence at lags 7–8, deepsoil ST was significantly positive (lag 7: 1.28 [1.04–1.59]; lag 8: 1.18 [1.02–1.37]), whereas midsoil ST (lag 7: 0.61 [0.44–0.84]), topsoil ST (lag 7: 0.50 [0.41–0.62]; lag 8: 0.70 [0.59–0.84]), and AT (lag 7: 0.67 [0.55–0.81]) were significantly negative—demonstrating depth-stratified divergence at the same time point. For winter incidence, significant positive temperature associations emerged at progressively shorter lags with increasing depth: AT at lags 5–6 (1.37 [1.05–1.80] and 1.22 [1.03–1.43]), topsoil ST at lags 4–5 (1.61 [1.03–2.53] and 1.35 [1.04–1.74]), midsoil ST at lag 3 (1.69 [1.08–2.63]), and deepsoil ST at lags 2–3 (1.54 [1.17–2.02] and 1.53 [1.08–2.15]). At lags 2–3, only deepsoil and midsoil ST were positively associated, whereas AT was negatively associated (lag 3: 0.59 [0.42–0.83]).

The two-stage multilayer stacked ensemble framework developed for this study (appendix figure S57, pp 70–71) was used to forecast monthly coccidioidomycosis incidence across the three study counties during a strictly held-out test period (2021–2024). Four pipeline configurations were compared. The smoothed-lag environmental-only pipeline was selected as the winning configuration based on visual inspection of test-period forecast plots—specifically peak-season timing and magnitude, trend fidelity, and county-level consistency—with test RMSE and gap ratio as secondary criteria (appendix figures S63–S65, pp 77–79, and table S10, p 16).

At stage 1 of the winning pipeline, the selected base learners were GLM.nb for the topsoil and midsoil layers, RF for the deepsoil and meteorological layers, and XGBoost for the dust dispersion layer (appendix table S8, p 14). At stage 2, an RF meta-learner combined the five base-learner out-of-fold predictions into the final ensemble forecast.

Visual inspection of the test-period forecasts suggested that the multilayer ensemble generally tracked observed monthly coccidioidomycosis incidence, capturing the seasonal cycling and interannual variation and reproducing the timing and approximate magnitude of most peaks across the four test years (figure 4). The model captured 2021 seasonal patterns well and reproduced the July 2022 peak, though it overestimated the late-2022 peak magnitude. The 2023 trend and peak-season timing were captured particularly well. For 2024, the model tracked the seasonal trend and December peak accurately but did not capture the elevated March 2024 incidence, a period affected by false-positive results from malfunctioning test kits.^25^ Although we corrected for documented false-positive cases during this period, the underestimation might reflect residual data artefacts rather than forecasting failure. At the county level, forecasts generally reproduced county-specific seasonal patterns across Maricopa, Pima, and Pinal counties, though some individual monthly peaks were over- or underpredicted (appendix figures S58–S60, pp 72–74).

**Figure 4:**
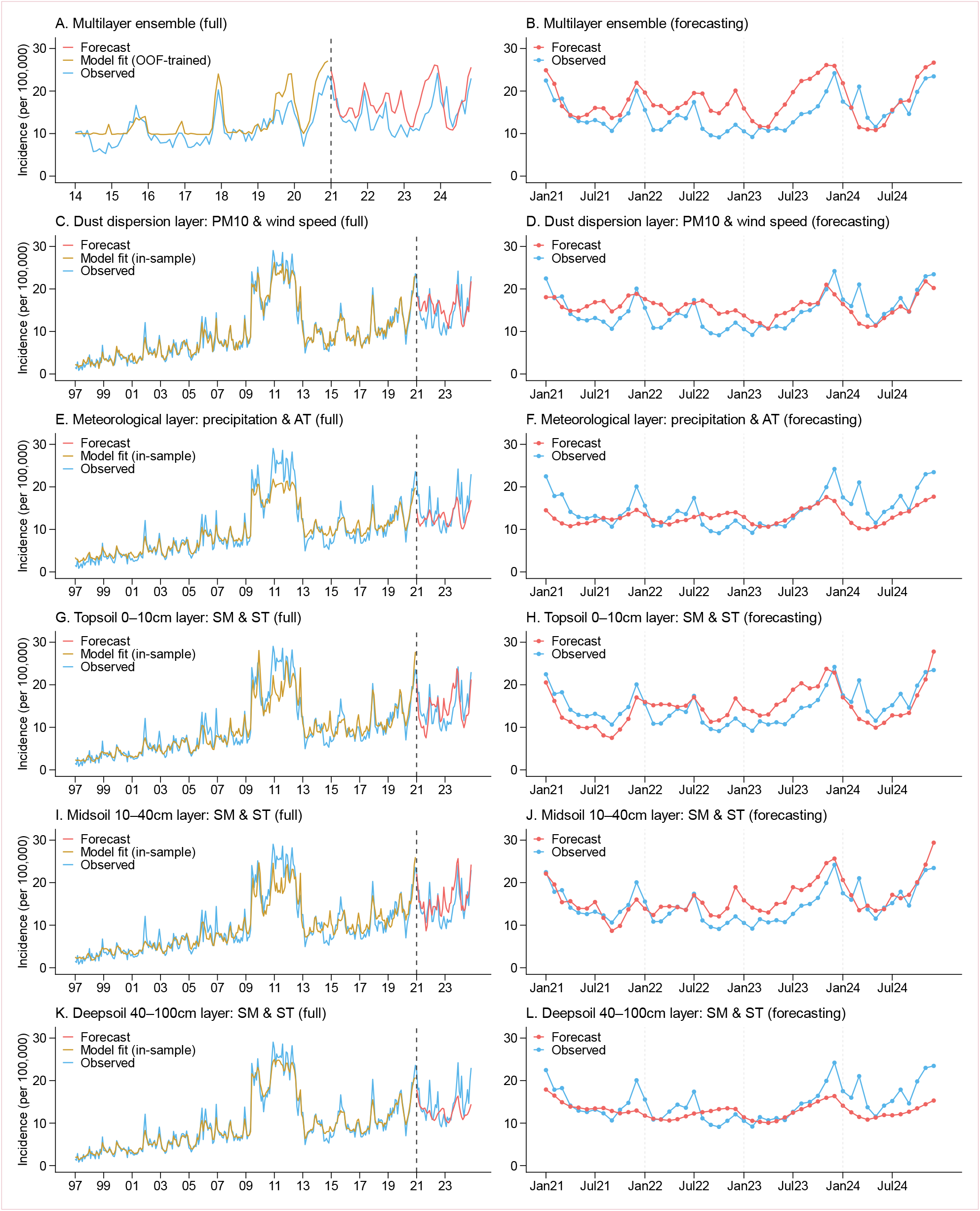
Observed and forecast monthly coccidioidomycosis incidence from the two-stage multilayer ensemble learning model in the study region (smoothed-lag environmental-only pipeline) The base learners (Stage 1) were trained on 1997–2020 using smoothed-lag environmental predictors, and the meta-learner (Stage 2) was trained on out-of-fold (OOF) predictions from the base learners during 2014–2020. The model was forecast over a strictly held-out period of 2021–2024. Left panels (A, C, E, G, I, K) show the full study period; right panels (B, D, F, H, J, L) show the forecasting period only. Panels A–B show the multilayer ensemble (meta-learner) combining OOF forecasts from all base learners. Panels C–L show individual base learners (five environmental layers): dust dispersion (PM_10_ and wind speed; C–D), meteorological conditions (precipitation and air temperature [AT]; E–F), topsoil 0–10 cm (soil moisture [SM] and soil temperature [ST]; G–H), middle soil 10–40 cm (I–J), and deep soil 40–100 cm (K–L).

Across both the combined study region and individual counties, the ensemble appeared to outperform all individual base learners on visual inspection. The topsoil and midsoil layers appeared comparable and performed most similarly to the ensemble, followed by the dust dispersion layer. The meteorological and deepsoil layers showed notably poorer peak-season capture than the ensemble. Test-period metrics supported this pattern: the ensemble achieved a test RMSE of 4.45 per 100,000 and a test gap ratio of 0.00 for the study region (appendix table S10, p 16). Although individual base learners achieved lower test RMSE values (range 3.25–3.66), their gap ratio were substantially higher (range 0.27–0.67; appendix table S11, p 17), indicating that the ensemble generalised to the test period far more reliably than any single environmental layer. Spatially, forecast performance was highest for the combined study region (test RMSE 4.45; gap ratio 0.00) and Maricopa County (4.50; 0.00), followed by Pima (4.67; 0.00) and Pinal (5.97; 0.11; appendix table S10, p 16).

Feature importance analysis was applied to the winning pipeline on the test data (2021–2024). All five environmental layers contributed to the ensemble forecast, with the dust dispersion and topsoil layers receiving the highest normalised SHAP-based importance in the stage 2 RF meta-learner, followed by the meteorological, deepsoil, and midsoil layers (appendix figure S62A, p 76). At the global level—combining layer importance with within-layer relative importance of each smoothed-lag environmental predictor group—PM_10_ was the most important predictor group, followed by topsoil SM, topsoil ST, precipitation, AT, deepsoil SM, wind speed, midsoil ST, midsoil SM, and deepsoil ST (appendix figure S62B, p 76). Within individual layers, moisture-related predictors contributed more than temperature-related predictors across all four soil–climate layers (appendix figure S61, p 75).

Three sensitivity analyses were conducted. First, adding autoregressive incidence lags (at 1, 12, 24, and 36 months) at the meta-learner stage produced the lowest test RMSE (3.02 vs 4.45 for environmental only) but a higher gap ratio (0.10 vs 0.00; appendix table S10, p 16), and systematically delayed peak-season predictions by approximately one month (appendix figure S63, p 77), making it practically inferior for near-real-time surveillance applications. Second, restricting incidence lags to annual cycles only (12, 24, and 36 months) showed no improvement in test RMSE (4.43 vs 4.45) and a worse gap ratio (0.08 vs 0.00; appendix table S10, p 16), with no improvement in peak-season capture (appendix figure S64, p 78), while adding dependence on prior surveillance data without meaningful gain. Third, replacing smoothed-lag features with raw monthly lags showed comparable peak-season timing and gap ratio for the study region but a higher test RMSE (4.67 vs 4.45), with notably poorer performance for Pima County (test RMSE 7.28 vs 4.67; gap ratio 0.11 vs 0.00; appendix figure S65, p 79, and table S10, p 16). The smoothed-lag environmental-only pipeline was therefore selected as the preferred configuration because it provided more stable performance that preserved peak season timing, addressed county-specific degradation, and eliminated dependence on prior surveillance data during the forecasting period.

## Discussion

To our knowledge, this is the first study to apply a comprehensive, multilayer environmental framework to a soilborne, environment-sensitive mycotic disease. Using DLNMs and a novel two-stage ensemble machine-learning approach across five environmental layers—dust dispersion and four soil–climate layers (meteorological, topsoil, midsoil, and deepsoil)—our analysis revealed three principal findings. First, increased coccidioidomycosis incidence was predominantly associated with concurrent dustier, windier, and drier conditions, cooler air temperatures, and warmer topsoil. Second, across lags 1–36 months, all four soil–climate layers exhibited non-monotonic wet–dry and cool–warm oscillations in their associations with incidence, alongside a depth-dependent lag structure and vertical divergence across layers—providing the first quantitative evidence in the vertical dimension for both the prevailing soil-sterilisation and grow-and-blow hypotheses. Third, the multilayer ensemble machine learning model outperformed any single-layer model in forecasting incidence, with all five layers contributing to the final prediction; because the selected model relied solely on environmental inputs available within one week, it could function as a near-real-time nowcast to inform public health preparedness. Together, these findings provide the first evidence linking multilayer environmental exposures to coccidioidomycosis incidence across both temporal and vertical dimensions, offer new support for the two prevailing hypotheses and demonstrate that a multilayer framework could improve both mechanistic understanding and predictive performance.

Our findings regarding short-term lags (1–3 months prior to case reporting) provide new evidence and insights into the concurrent conditions that facilitate spore dispersal and infection (ie, the ‘blow’ stage of the grow-and-blow hypothesis). We observed that higher incidence was predominantly associated with concurrent dustier (higher PM_10_), windier (higher wind speed), drier (lower precipitation and lower SM across topsoil, midsoil, and deepsoil), cooler AT, and warmer topsoil conditions. As the first study to characterise the exposure–lag–response relationship for PM_10_ and coccidioidomycosis incidence, we found that concurrent PM_10_ exhibited consistent positive associations across all four incidence seasons. Notably, PM_10_ also emerged as the most important environmental predictor group in our ensemble forecast, collectively reinforcing its role as a proxy for airborne arthroconidia concentration and providing empirical support for the recently proposed dust-borne atmospheric transport hypothesis.^28^ We found that concurrent drier conditions were consistently associated with increased incidence across all four soil-climate layers and all four incidence seasons, extending existing evidence from precipitation^7^ and topsoil SM^3^ to midsoil and deepsoil moisture—consistent with a recent study linking increased airborne *Coccidioides* detection to lower daily surface SM.^29^ Notably, increased incidence was associated with concurrent warmer-drier topsoil conditions, aligning with the classical mechanism of hyphal desiccation, fragmentation, and arthroconidia aerosolisation, yet unexpectedly also with concurrent cooler-drier atmospheric conditions. This suggests that beyond the traditional ‘blow’ phase (ie, arthroconidia formation and aerosolisation from topsoil following soil disturbance), the dispersal process might also involve a ‘persist’ phase in which cooler-drier atmospheric conditions prolong arthroconidia viability. Lower temperatures were associated with reduced UV exposure,^30^ which might extend arthroconidia survival in the air or on surfaces. However, direct laboratory evidence remains limited. An early laboratory study^31^ observed substantially greater *Coccidioides* viability at 4°C than at 37°C after three and six months under low relative humidity, but did not evaluate finer temperature gradients within the 8–36°C range—the monthly AT observed across the tri-county region during 1997–2024 (appendix table S2, pp 3–4)—under dry conditions. Further laboratory studies across finer temperature–moisture gradients that simulate endemic soil and atmospheric conditions are warranted to clarify the mechanisms underlying these observed associations.

Across lags 1–36 months, environmental exposures within each of the four soil–climate layers showed nonlinear, non-monotonic, and season-dependent associations with coccidioidomycosis incidence, characterised by alternating wet–dry and cool–warm oscillations rather than a single monotonic lag structure. These alternating hydroclimatic cycles within each layer are broadly consistent with the temporal ordering implied by the grow-and-blow hypothesis, in which antecedent adequate moisture and favourable temperatures support fungal growth, whereas subsequent drying promotes arthroconidia formation and dispersal.^14–17^ Similar alternating wet–dry and cool–warm patterns have been observed via monthly precipitation and AT using DLNMs in hyperendemic California counties^7^ and our earlier study identified similar cyclical patterns via topsoil SM and ST using GLM.nb at the seasonal–county level in the same tri-county region,^3^ supporting the generality of this pattern across different climate indicators, modelling approaches, and endemic settings. The present study extends this evidence by revealing these patterns at monthly resolution across multiple environmental layers. This cyclical pattern was most pronounced in the topsoil layer—which showed the greatest number of significant associations across all four incidence seasons—and attenuated progressively from topsoil through midsoil to deepsoil. One possible explanation for this attenuation is vertical buffering within the soil profile: deeper soil layers exhibit progressively dampened temporal variability in both moisture and temperature (figure 2; appendix figure S3, p 20, and table S2, pp 3–4),^32–34^ reducing exposure variability and thereby potentially limiting the statistical detectability of lag-specific associations.^35^ Alternatively, this attenuation might reflect the greater biological relevance of upper soil layers, where *Coccidioides* is thought to be most concentrated^15^ and where hydroclimatic conditions might influence the full fungal lifecycle—from hyphal growth through fragmentation to dispersal—whereas deeper-layer associations might primarily capture antecedent conditions conducive to hyphal persistence and growth. Unexpectedly, the present study also identified significant antecedent dry–cool associations (ie, significantly negative moisture and temperature IRRs at the same lag) with increased incidence at multiple lags (lags 4–36) across all four seasons and multiple layers—a finding that contrasts with our earlier seasonal-scale study, which found no such associations.^3^ The same California study also observed a significant dry–cool association at lag 4 (precipitation–AT) for fall incidence, alongside several non-significant dry–cool associations at longer lags.^7^ This discrepancy with our earlier study likely reflects methodological differences: the monthly-resolution DLNM with flexible nonlinear lag structures can detect transient dry–cool windows that seasonal aggregation and linear modelling frameworks would average out. These antecedent dry–cool windows might represent a ‘persistence’ phase, in which cooler, drier conditions prolong the environmental persistence or viability of *Coccidioides* across multiple layers before subsequent favourable conditions support renewed growth or dispersal.

A novel finding was the depth-dependent lag structure observed for moisture and temperature variables across all four soil–climate layers: significant positive associations with coccidioidomycosis incidence appeared at progressively shorter lags with increasing depth, accompanied by vertical divergence in association direction across depths at the same lag windows. For moisture, the depth-dependent gradient was most evident in fall, monsoon, and winter incidence: significant positive associations emerged at progressively shorter lags from precipitation through topsoil, midsoil, to deepsoil SM, and at some of these same lag windows where deepsoil SM showed significant positive associations, topsoil SM and/or precipitation remained negatively associated. This moisture-related pattern might reflect the soil moisture memory effect, whereby deeper layers retain antecedent moisture longer than shallower layers.^36,37^ Under this interpretation, deeper soil might continue to provide relatively favourable moisture conditions for fungal growth while the surface has already transitioned into a drier state that might sterilise microbial competitors and promote arthroconidial formation and dispersal. For temperature, a comparable depth-dependent gradient was most evident in fall and winter incidence: significant positive associations emerged at progressively shorter lags from AT through topsoil, midsoil, to deepsoil ST. At some of these same lag windows, deepsoil ST showed significant positive associations while AT—and, in fall, shallower STs—remained negatively associated. This temperature-related pattern might reflect soil temperature memory effect, whereby deeper layers retain antecedent thermal conditions longer than shallower layers.^33,38^ Under this interpretation, deeper soil might continue to provide relatively favourable thermal conditions for fungal growth while ambient conditions have already transitioned into cooler conditions that might be less favourable for growth but more favourable for *Coccidioides* persistence. Together, these moisture and temperature findings suggest that deeper soils might serve as a buffered subsurface refugium for *Coccidioides*, retaining warm, moist conditions longer than shallower layers. These patterns are undetectable in single-layer analyses and provide the first quantitative evidence in the vertical dimension for both prevailing mechanistic hypotheses: that deeper soils might act as a refugium from surface sterilisation, as posited by the soil-sterilisation hypothesis, and might sustain favourable growth conditions at depth even after the surface has transitioned to dispersal- or persistence-phase conditions, consistent with the grow-and-blow hypothesis.

Our two-stage multilayer ensemble results demonstrated that integrating environmental predictors across multilayers achieved superior forecasting performance compared with any single layer alone. Although individual base learners yielded lower test RMSE values, the ensemble more faithfully captured observed incidence patterns while achieving substantially lower gap ratios, indicating stronger generalisation to the held-out test period. Among single-layer models, the topsoil and midsoil layers most closely resembled the ensemble in capturing observed patterns, followed by the dust-dispersion layer, whereas the meteorological and deepsoil layers showed notably weaker performance. Spatially, forecast performance was highest for the combined tri-county region and Maricopa County, followed by Pima then Pinal—mirroring the gradient in case counts across counties and likely reflecting greater statistical stability in settings with larger case counts. Feature importance analysis revealed that all five layers contributed to the final ensemble forecast, with the dust-dispersion and topsoil layers receiving the highest importance, followed by the meteorological, deepsoil, and midsoil layers. The relatively lower importance assigned to midsoil compared with deepsoil might not reflect a genuine difference in ecological relevance but rather the high correlation between topsoil and midsoil conditions: once the ensemble had captured much of the overlapping information from topsoil, the marginal contribution from midsoil was limited, whereas deepsoil—being more distinct from topsoil—could provide additional predictive information. Although we used aggregated TreeSHAP and grouped permutation at the predictor-group level rather than individual lags to mitigate correlation-driven importance bias, the meta-learner might still not fully disentangle contributions from highly correlated layers, potentially underestimating midsoil’s true importance. Nevertheless, although interpretation of relative importance across correlated layers requires caution, the finding that all five layers contributed to the ensemble—which outperformed any single layer—underscores the value of multilayer environmental information for improving coccidioidomycosis prediction.

The smoothed-lag environmental-only pipeline achieved the best forecasting performance and was therefore selected as the primary model; the remaining three configurations were reported as sensitivity analyses. Adding autoregressive incidence lags at the meta-learner stage lowered test RMSE but increased the gap ratio and systematically delayed peak-season predictions by about one month. Because this configuration requires the previous month’s reported case count, forecasts are limited to at most one month ahead; the observed peak delay therefore means that peak-incidence periods would be identified only after they have already occurred, offering little prediction value over routine surveillance. Restricting incidence lags to annual cycles only offered no meaningful improvement while still introducing dependence on prior surveillance data. Smoothed-lag features also outperformed raw monthly lags, likely because the three-month moving averages provided beneficial noise reduction and implicit dimensionality reduction that improved prediction accuracy. Across both primary and sensitivity analyses, GLM.nb was consistently selected as the best base learner for the topsoil layer, aligning with the strong performance of GLM.nb for topsoil reported in our earlier seasonal-scale study.^3^

The selected model structure also has practical and methodological implications. Because the winning pipeline avoided dependence on prior surveillance counts and all environmental predictors are available within one week of each target month, the model can function as a near-real-time nowcast—generating incidence estimates well before finalised surveillance data become available. Although these estimates are produced shortly after each target month, the prolonged diagnostic pathway for coccidioidomycosis means that many contemporaneously exposed individuals remain undiagnosed when the nowcast becomes available: symptoms typically appear 1–4 weeks after exposure,^39^ yet patients waited an average of 44 days before seeking healthcare, and the interval from symptomatic onset to diagnosis averaged 209 days.^25,39^ Importantly, patients with prior awareness of coccidioidomycosis were diagnosed substantially earlier than those unfamiliar with the disease (mean 79 vs 282 days from symptomatic onset) and were twice as likely to request testing from their healthcare provider.^25,39^ Nowcast-identified high-incidence periods could therefore inform public health preparedness and heighten clinical and public awareness, potentially prompting earlier diagnostic consideration and targeted patient counselling for individuals still progressing through the diagnostic pathway. Future studies might further extend lead time by incorporating forecast-derived or projected covariates, enabling estimates before the target month rather than shortly after. Our two-stage framework is intentionally modular and flexible: future studies could retain the multilayer architecture while substituting or adding candidate models (e.g., deep learning or other machine learning approaches), incorporating additional environmental predictors—either as new layers or integrated into existing ones (e.g., drought indicators, soil texture, soil chemistry, elevation, land use, vegetation)—as well as non-environmental factors (e.g., small-mammal host distributions, sociodemographic changes, construction and agricultural activity, occupational and recreational exposure) to further refine forecasting performance. Accordingly, the main contribution of this predictive framework might lie less in any particular set of candidate models or predictors than in demonstrating that a flexible multilayer ensemble architecture can evaluate and integrate heterogeneous, multisource data into a more robust forecasting system for coccidioidomycosis.

This study has several strengths. First, the mechanism-informed multilayer, multiyear lagged environmental framework via DLNMs advances mechanistic understanding of coccidioidomycosis. This is the first study to comprehensively evaluate and compare coccidioidomycosis incidence in relation to environmental exposures across both above-ground (atmospheric and dust-dispersion) and below-ground (topsoil, midsoil, and deepsoil) multilayers, providing the first evidence linking multilayer environmental exposures to coccidioidomycosis incidence across both temporal and vertical dimensions. The depth-dependent lag structures and vertical divergence observed across layers provide the first quantitative evidence in the vertical dimension for both the prevailing soil-sterilisation and grow-and-blow hypotheses. The multilayer design also offers new evidence in the temporal dimension across multiple environmental layers, particularly extending the wet–dry and cool–warm cyclical pattern evidence to midsoil and deepsoil layers. Second, the DLNM framework enabled the first characterisation of nonlinear exposure–lag–response relationships for subsurface SM and ST at three depth intervals, and for the dust-dispersion layer (PM_10_ and wind speed), in coccidioidomycosis research. Notably, the consistent concurrent positive PM_10_–incidence associations across all four seasons, reinforced by PM_10_ ranking as the most important ensemble predictor, jointly provide the first evidence for the dust-borne atmospheric transport hypothesis.^28^ Third, the 28-year surveillance period, combined with 36-month distributed lags, provides substantial temporal depth for detecting multiyear antecedent hydroclimatic signals. Fourth, the season-specific DLNM interaction terms revealed not only season-varying exposure–lag–response associations but also general patterns consistent across seasons.

The two-stage multilayer ensemble framework offers several additional strengths. First, integrating predictors from all five environmental layers outperformed any single-layer model, demonstrating that multilayer information improves forecasting performance. Second, evaluating diverse candidate model types—including linear (GLM.nb), nonlinear (DLNM), and tree-based (RF, XGBoost) approaches—at each layer enhances the robustness of model selection beyond reliance on any single model class. Third, the framework is modular, flexible, and extensible, allowing future studies to retain the multilayer architecture while substituting or adding candidate models and incorporating additional environmental and non-environmental predictors; accordingly, the main contribution might lie less in any particular set of candidate models or predictors than in the multilayer ensemble architecture itself. Fourth, the framework incorporates progressive walk-forward cross-validation, strict temporal separation between training and test periods, recency-weighted model selection with tiebreak criteria, and multiple sensitivity analyses. Finally, the winning pipeline relied solely on environmental covariates available within one week of the target month, enabling the model to function as a near-real-time nowcast for public health preparedness.

Several limitations should be noted. First, two sources of exposure misclassification might have biased coccidioidomycosis case counts: (a) cases were assigned to the county of residence rather than the true location of spore exposure, and (b) delays from spore exposure to case notification, which typically span weeks to months,^25^ introduced temporal misalignment. To mitigate (a)—spatial misclassification arising from travel- or work-related exposures outside the county of residence—we restricted analyses to the hyperendemic tri-county area, but some misclassification might remain, likely attenuating exposure–outcome associations. To mitigate (b), we treated the first three lag months as the concurrent exposure period, but residual temporal misalignment might persist. Future studies with individual-level case data—where multiple date fields including symptom onset, diagnosis, and report dates are available—might further reduce this bias by estimating exposure dates from the known temporal relationships among these dates.^14,25^ Such data would also permit analysis at finer spatial resolution, such as census-tract-level counts derived from geocoded residential addresses, where larger sample sizes might further improve predictive performance and the precision of effect estimation. We encourage such studies to adopt the multilayer framework proposed here to help validate our findings. Second, as a county-level ecological time-series, the study cannot establish causality and is subject to ecological fallacy: estimated associations captured population-level patterns that might not reflect individual-level risk. Third, case counts could be biased by changes in case definitions, laboratory reporting, and testing practices over the study period,^25^ creating surveillance artifacts unrelated to true incidence. We addressed these through (a) numeric correction where quantitative evidence of data quality issues was available and (b) a categorical surveillance regime indicator to account for documented major transitions (appendix figure S2, p 19), but residual bias could not be entirely excluded. Fourth, reanalysis-derived SM and ST products were subject to parametrisation uncertainty, might not represent fine-scale soil heterogeneity, and could diverge from in-situ observations. However, county-level aggregation diminished the influence of localised discrepancies, and these products have been extensively validated and shown to reflect observed patterns well.^3,40,41^ Lastly, our study could not disentangle direct environmental effects on *Coccidioides* growth from indirect effects mediated through vegetation that influenced small-mammal abundance, as posited by the “endozoan, small-mammal reservoir” hypothesis^42^—a pathway that might contribute to delays in environmental effects on infection. Nonetheless, our findings—including multiyear antecedent wet–dry and cool–warm oscillations, depth-dependent lag structures for moisture and temperature across layers, and the superior predictive performance of the multilayer framework—suggest that future research related to not only the two prevailing mechanistic hypotheses but also the endozoan hypothesis might benefit from adopting a multilayer environmental framework.

Our study provides new evidence that multilayer environmental framework proposed here can advance both mechanistic understanding and predictive performance for coccidioidomycosis. By combining a multiyear, multilayer lagged environmental framework for effect estimation with a two-stage ensemble architecture for prediction, we linked multilayer environmental exposures to coccidioidomycosis incidence across both temporal and vertical dimensions and demonstrated that multilayer ensemble prediction outperformed single-layer approaches. Although specific environmental associations, best-performing models, and predictor importance may vary across endemic settings, the multilayer framework itself has the potential to be generalised to other endemic regions. We therefore encourage future efforts in coccidioidomycosis surveillance, effect estimation, and prediction to adopt and refine this multilayer framework, adapting covariates and models as needed.

## Data Availability

Coccidioidomycosis case-count data were provided by the Arizona Department of Health Services and are available from the Arizona Department of Health Services upon reasonable request, subject to their data-sharing policies. All other data used in this study are publicly available; sources are listed in table S1 of the supplementary appendix.

## Contributor Roles

Qianqian Li conceived and designed the study; conducted conceptualization, data curation, methodology development, formal analysis, project administration, validation, visualization, and original draft writing. Yue Zhan and Runqiu Wang assisted with statistical methods. Haiyue Li assisted with the curation of partial climate data. Jesse E. Bell acquired funding, supervised the study, and approved the final submission for publication. All authors reviewed and edited the final manuscript.

## Data sharing

Coccidioidomycosis case-count data were provided by the Arizona Department of Health Services (ADHS) and are available from ADHS upon reasonable request, subject to their data-sharing policies. All other data used in this study are publicly available; sources are listed in the appendix (table S1, p 2).

## Acknowledgments

We thank Thomas Williamson (Arizona Department of Health Services) for assistance in obtaining coccidioidomycosis case data. We also thank JoEllyn McMillan, Hongying Dai and Yiqun Jiang (University of Nebraska Medical Center) for their valuable suggestions or comments on the research. This work was supported in part by the National Oceanic and Atmospheric Administration (NOAA) National Integrated Drought Information System (NIDIS) program through the project “ Evaluation of Drought Indicators for Improved Decision-Making in Public Health and Emergency Preparedness: Reducing Drought’s Burden on Health” (grant NA20OAR4310368), and by the National Aeronautics and Space Administration (NASA) Research Opportunities in Space and Earth Sciences (ROSES) program through the project “ Identifying Public Health Applications of Satellite-derived Drought Indicators: Improved Monitoring for Respiratory Health” (grant 80NSSC22K1050).

## IRB

Ethics committee/IRB of the University of Nebraska Medical Center (UNMC) waived ethical approval for this work. The work relied exclusively on de-identified, county-level coccidioidomycosis case-count data provided by the Arizona Department of Health Services (ADHS), which are available upon request from the ADHS. These case-count data contain no individual identifiable information or protected health information. And we did not interact or intervene with individuals to obtain information about them. Accordingly, the UNMC IRB Office of Regulatory Affairs (ORA) has determined that this project does not constitute human subject research as defined at 45CFR46.102 and is therefore not subject to the federal regulations.

