## Supplementary Appendix for "Effects and predictive performance of multilayer environmental exposures on coccidioidomycosis: a longitudinal surveillance study"

Figure S35. Lag-specific exposure–response relationships between spring (April–June) coccidioidomycosis incidence and above-ground $PM_{10}$ (Layer 1: dust dispersion layer) at lags 1–36 months. 51

Figure S36. Lag-specific exposure–response relationships between spring (April–June) coccidioidomycosis incidence and above-ground wind speed (Layer 1: dust dispersion layer) at lags 1–36 months. 51

Figure S37. Lag-specific exposure–response relationships between monsoon (July–September) coccidioidomycosis incidence and below-ground soil moisture (Layer 3: 0–10 cm; topsoil; SM0t10) at lags 1–36 months. 52

Figure S38. Lag-specific exposure–response relationships between monsoon (July–September) coccidioidomycosis incidence and below-ground soil temperature (Layer 3: 0–10 cm; topsoil; ST0t10) at lags 1–36 months. 53

Figure S39. Lag-specific exposure–response relationships between monsoon (July–September) coccidioidomycosis incidence and below-ground soil moisture (Layer 4: 10–40 cm; SM10t40) at lags 1–36 months. 54

Figure S40. Lag-specific exposure–response relationships between monsoon (July–September) coccidioidomycosis incidence and below-ground soil temperature (Layer 4: 10–40 cm; ST10t40) at lags 1–36 months. 55

Figure S41. Lag-specific exposure–response relationships between monsoon (July–September) coccidioidomycosis incidence and below-ground soil moisture (Layer 5: 40–100 cm; SM40t100) at lags 1–36 months. 56

Figure S42. Lag-specific exposure–response relationships between monsoon (July–September) coccidioidomycosis incidence and below-ground soil temperature (Layer 5: 40–100 cm; ST40t100) at lags 1–36 months. 57

Figure S43. Lag-specific exposure–response relationships between monsoon (July–September) coccidioidomycosis incidence and above-ground precipitation (Layer 2: meteorological layer) at lags 1–36 months. 58

Figure S44. Lag-specific exposure–response relationships between monsoon (July–September) coccidioidomycosis incidence and above-ground air temperature (Layer 2: meteorological layer; AT) at lags 1–36 months. 59

Figure S45. Lag-specific exposure–response relationships between monsoon (July–September) coccidioidomycosis incidence and above-ground $PM_{10}$ (Layer 1: dust dispersion layer) at lags 1–36 months. 60

Figure S46. Lag-specific exposure–response relationships between monsoon (July–September) coccidioidomycosis incidence and above-ground wind speed (Layer 1: dust dispersion layer) at lags 1–36 months. 60

Figure S47. Lag-specific exposure–response relationships between fall (October–December) coccidioidomycosis incidence and below-ground soil moisture (Layer 3: 0–10 cm; topsoil; SM0t10) at lags 1–36 months. 61

Figure S48. Lag-specific exposure–response relationships between fall (October–December) coccidioidomycosis incidence and below-ground soil temperature (Layer 3: 0–10 cm; topsoil; ST0t10) at lags 1–36 months. 62

Figure S49. Lag-specific exposure–response relationships between fall (October–December) coccidioidomycosis incidence and below-ground soil moisture (Layer 4: 10–40 cm; SM10t40) at lags 1–36 months. 63

Figure S50. Lag-specific exposure–response relationships between fall (October–December) coccidioidomycosis incidence and below-ground soil temperature (Layer 4: 10–40 cm; ST10t40) at lags 1–36 months. 64

Figure S51. Lag-specific exposure–response relationships between fall (October–December) coccidioidomycosis incidence and below-ground soil moisture (Layer 5: 40–100 cm; SM40t100) at lags 1–36 months. 65

Figure S52. Lag-specific exposure–response relationships between fall (October–December) coccidioidomycosis incidence and below-ground soil temperature (Layer 5: 40–100 cm; ST40t100) at lags 1–36 months. 66

Figure S53. Lag-specific exposure–response relationships between fall (October–December) coccidioidomycosis incidence and above-ground precipitation (Layer 2: meteorological layer) at lags 1–36 months. 67

Figure S54. Lag-specific exposure–response relationships between fall (October–December) coccidioidomycosis incidence and above-ground air temperature (Layer 2: meteorological layer; AT) at lags 1–36 months. 68

Figure S55. Lag-specific exposure–response relationships between fall (October–December) coccidioidomycosis incidence and above-ground $PM_{10}$ (Layer 1: dust dispersion layer) at lags 1–36 months. 69

Figure S56. Lag-specific exposure–response relationships between fall (October–December) coccidioidomycosis incidence and above-ground wind speed (Layer 1: dust dispersion layer) at lags 1–36 months. 69

Figure S57. Two-stage multilayer stacked ensemble machine learning framework for forecasting coccidioidomycosis incidence. 70

### Supplemental Texts

#### Text S1. Environmental exposure data

Environmental exposure data spanning January 1994 to December 2024 were compiled to enable distributed lag non-linear modelling (DLNM) with lag structures of up to 36 months prior to the first disease observation month (January 1997). Data sources, variables, spatial and temporal resolutions, and coverage periods are summarized in table S1.

We organized predictors into a five-layer environmental framework designed to capture the above- and below-ground conditions hypothesized to influence *Coccidioides* growth and dust-borne dispersal: (1) an meteorological layer (precipitation and air temperature); (2) a topsoil layer (0–10 cm soil moisture [SM] and soil temperature [ST]); (3) a midsoil layer (10–40 cm SM and ST); (4) a deepsoil layer (40–100 cm SM and ST); and (5) a dust-dispersion layer ($PM_{10}$ concentration and wind speed). The first four layers were classified as soil-climate layers, and the fifth represented conditions related to fungal transport and potential human exposure.

For the meteorological layer, monthly total precipitation (mm) and mean air temperature (°C) were obtained from the parameter-elevation regressions on independent slopes model (PRISM), at 4 km grid resolution.[1] PRISM uses climatologically aided interpolation to generate estimates from over 25,000 precipitation and temperature stations daily, accounting for elevation, coastal effects, temperature inversions, and terrain barriers.

For the three subsurface layers, soil moisture (SM; kg $m^{-2}$) and soil temperature (ST; K) at 0–10, 10–40, and 40–100 cm were extracted from a single gridded product: the North American Land Data Assimilation System Phase 2 (NLDAS-2) Noah land surface model (LSM), with a spatial resolution of 0.125° × 0.125°.[2] Within NLDAS-2, the Noah LSM simulates SM and ST by assimilating observed precipitation, air temperature, surface pressure, and solar radiation, together with observation- and reanalysis-derived soil and vegetation parameters.[3–5] These SM and ST estimates are produced through a coupled, iterative modelling framework in which water and energy states dynamically interact and feedback on one another through multiple linear and non-linear physical processes.[5] As a result, the SM and ST represent integrated moisture-temperature conditions shaped by interacting drivers such as precipitation, air temperature, evapotranspiration, runoff, infiltration, radiation, and soil and vegetation properties.[5] The performance of these products has been extensively evaluated against in situ observations across soil depths (0–10 cm, 10–40 cm, 40–100 cm) and temporal scales (daily, monthly, and annual).[6,7] Specifically, Xia and colleagues demonstrated that the Noah LSM effectively captures broad patterns of observed SM variability—including seasonal cycles, interannual trends, and wet–dry events—across depths and timescales examined.[6] This capability is especially relevant here, given the central importance of soil wet–dry transitions in the prevailing "grow and blow" hypothesis of coccidioidomycosis ecology. Likewise, Xia and colleagues reported good agreement between Noah-simulated and observed ST.[7] Our earlier study used only topsoil (0–10 cm) SM and ST,[8] the present study extends this framework to include midsoil (10–40 cm) and deepsoil (40–100 cm) layers to more fully characterize subsurface moisture and thermal dynamics hypothesized to influence *Coccidioides* ecology.

For the dust-dispersion layers, mean wind speed (m $s^{-1}$) was obtained from the NLDAS-2 primary forcing dataset,[9] which provides10-m above-ground zonal (west–east; $u$) and meridional (south–north; $v$) wind components derived from the North American Regional Reanalysis (NARR).[3] Total wind speed was calculated as $\sqrt{(u^2 + v^2)}$. Mean $PM_{10}$ concentration (μg $m^{-3}$) was derived from daily measurements at outdoor monitoring sites recorded in the US Environmental Protection Agency (EPA) Air Quality System.[10] Daily site-level concentrations were first aggregated to monthly means per site, then averaged across all sites within each county to produce county-level monthly mean $PM_{10}$. For descriptive purposes and comparability with our earlier work,[8] we additionally computed a dusty-day indicator—defined as the number of days per month on which daily mean $PM_{10}$ exceeded 45 μg $m^{-3}$ at any site within a county—consistent with the World Health Organization (WHO) global air quality guideline.[11] This indicator was used for characterization only and was not entered into inferential or predictive models. $PM_{10}$ concentration was chosen for statistical modelling because, as a continuous measure, it is directly suited to exposure–lag–response estimation via distributed lag non-linear models (DLNMs) and offers greater information content than threshold-based counts.

All gridded datasets (PRISM and NLDAS-2) were spatially aggregated to county-level monthly means using zonal statistics based on county boundary shapefiles. Site-level $PM_{10}$ measurements were averaged across monitoring sites within each county-month. The integrated multi-source environmental exposure dataset was then linked to surveillance case counts by county and calendar month for analysis.

#### Supplementary Tables

**Table S1.** List of data used in this study.

| Category | Variables | Spatial/Temporal resolution | Period | Source |
|---|---|---|---|---|
| Above-ground exposures | Total Precipitation (mm) | 4km/Monthly | 1994–2024 | PRISM Group, Oregon State University[1] |
| | Air temperature (°C) | 4km/Monthly | 1994–2024 | PRISM Group, Oregon State University[1] |
| | Wind speed at 10 m above ground (m $s^{-1}$) | 0.125°×0.125°/Monthly | 1994–2024 | NLDAS, Primary forcing data, Version 2.0[9] |
| | Daily mean $PM_{10}$ concentration (μg $m^{-3}$) | Monitoring site/Daily | 1994–2024 | U.S. EPA, Outdoor air quality data[10] |
| Below-ground exposures | Soil moisture (0–10 cm, 10–40 cm, 40–100 cm) (kg $m^{-2}$) | 0.125°×0.125°/Monthly | 1994–2024 | NLDAS, Noah LSM, Version 2.0[2] |
| | Soil temperature (0–10 cm, 10–40 cm, 40–100 cm) (K) | 0.125°×0.125°/Monthly | 1994–2024 | NLDAS, Noah LSM, Version 2.0[2] |
| Outcome | Valley fever case count | County/Monthly | 1997–2024 | ADHS |

**Abbreviations**: PRISM Group = Parameter-elevation Regressions on Independent Slopes Model Group; NLDAS = North American Land Data Assimilation System; LSM = Land Surface Model; EPA = U.S. Environmental Protection Agency; ADHS = Arizona Department of Health Services.

**Table S2.** Descriptive statistics of coccidioidomycosis and environmental variables across seasons in the tri-county study region (Maricopa, Pima, and Pinal counties), Arizona.

| Variable by season | Mean (SD) | Min | Max | 5th | 25th | 50th | 75th | 95th |
|---|---|---|---|---|---|---|---|---|
| Fall | | | | | | | | |
| CM | 218 (291) | 3 | 1251 | 9 | 47 | 94 | 227 | 953 |
| CM incidence | 11.45 (7.32) | 0.87 | 34.47 | 2.64 | 6.1 | 10.03 | 14.83 | 26.43 |
| SM0t10, kg m-2 | 14.30 (3.79) | 7.46 | 22.62 | 8.64 | 11.26 | 13.85 | 17.13 | 20.99 |
| SM10t40, kg m-2 | 53.55 (4.70) | 44.85 | 68.74 | 47.21 | 49.99 | 53.07 | 55.95 | 63.2 |
| SM40t100, kg m-2 | 104.38 (9.02) | 79.71 | 129.11 | 90.01 | 98.35 | 105.07 | 110.47 | 118.66 |
| ST0t10, K | 286.25 (5.36) | 277.67 | 296.79 | 279.01 | 281.28 | 285.38 | 291.99 | 294.51 |
| ST10t40, K | 288.14 (5.26) | 279.89 | 297.92 | 281.18 | 283.05 | 287.44 | 293.77 | 295.88 |
| ST40t100, K | 290.13 (4.96) | 282.29 | 299.08 | 283.4 | 284.99 | 289.84 | 295.34 | 297.41 |
| Prep, mm | 18.14 (22.14) | 0 | 116.83 | 0.01 | 2.23 | 9.69 | 25.76 | 64.7 |
| AT, °C | 16.28 (4.87) | 8.13 | 26.33 | 9.61 | 11.71 | 16.37 | 20.89 | 23.75 |
| Wind, m s-1 | 0.90 (0.56) | 0.06 | 2.8 | 0.19 | 0.45 | 0.8 | 1.28 | 1.93 |
| PM10, µg m-3 | 39.25 (18.65) | 14.46 | 136.17 | 18.78 | 26.61 | 34.06 | 46.83 | 71.86 |
| DustyDays | 16 (10) | 0 | 31 | 3 | 8 | 16 | 25 | 31 |
| Monsoon | | | | | | | | |
| CM | 178 (237) | 2 | 1272 | 7 | 39 | 86 | 175 | 728 |
| CM incidence | 9.58 (5.89) | 0.93 | 32.93 | 2.19 | 5.34 | 8.26 | 13.01 | 20.33 |
| SM0t10, kg m-2 | 14.44 (3.02) | 6.6 | 21.05 | 9.13 | 12.46 | 14.87 | 16.66 | 18.99 |
| SM10t40, kg m-2 | 54.53 (4.45) | 42.33 | 68 | 48.24 | 51.38 | 53.75 | 57.38 | 62.44 |
| SM40t100, kg m-2 | 105.16 (9.13) | 82.04 | 133.68 | 91.18 | 99.38 | 105 | 110.92 | 120.28 |
| ST0t10, K | 303.22 (2.29) | 297.67 | 307.69 | 299.3 | 301.67 | 303.31 | 305.13 | 306.55 |
| ST10t40, K | 302.17 (1.66) | 298.18 | 305.76 | 299.5 | 301.01 | 302.08 | 303.4 | 304.91 |
| ST40t100, K | 300.83 (1.50) | 296.84 | 304.29 | 298.33 | 299.65 | 300.81 | 301.95 | 303.34 |
| Prep, mm | 36.06 (23.82) | 0.01 | 149.84 | 4.27 | 18.52 | 32.47 | 50.53 | 82.52 |
| AT, °C | 30.34 (2.04) | 25.08 | 35.71 | 27.16 | 28.91 | 30.41 | 31.8 | 33.5 |
| Wind, m s-1 | 2.51 (0.48) | 0.99 | 3.54 | 1.65 | 2.22 | 2.55 | 2.86 | 3.2 |
| PM10, µg m-3 | 36.07 (16.74) | 15.39 | 133.2 | 18.6 | 23.91 | 32.34 | 44.25 | 68.96 |
| DustyDays | 14 (10) | 0 | 31 | 1 | 5 | 14 | 23 | 29 |
| Spring | | | | | | | | |
| CM | 165 (229) | 1 | 1239 | 6 | 37 | 76 | 164 | 655 |
| CM incidence | 8.89 (5.76) | 0.49 | 31.75 | 1.79 | 4.62 | 8.1 | 11.52 | 19.04 |
| SM0t10, kg m-2 | 9.83 (2.37) | 6.38 | 17.66 | 7.05 | 7.85 | 9.29 | 11.25 | 14.52 |
| SM10t40, kg m-2 | 52.70 (4.05) | 41.94 | 65.43 | 45.79 | 50.01 | 52.73 | 55.23 | 59.06 |
| SM40t100, kg m-2 | 107.68 (9.13) | 83.33 | 134.59 | 93.43 | 101.54 | 106.54 | 113.92 | 123.02 |
| ST0t10, K | 295.21 (4.45) | 286.79 | 303.49 | 288.8 | 291.08 | 295.14 | 299.24 | 302.24 |
| ST10t40, K | 293.11 (3.77) | 286.02 | 300.71 | 287.63 | 289.75 | 292.96 | 296.55 | 298.96 |
| ST40t100, K | 291.35 (3.32) | 285.16 | 297.83 | 286.61 | 288.29 | 291.19 | 294.45 | 296.49 |
| Prep, mm | 4.99 (7.90) | 0 | 44.77 | 0 | 0.09 | 1.4 | 6.16 | 20.96 |
| AT, °C | 24.28 (4.41) | 15.16 | 32.26 | 18.1 | 20.33 | 24.09 | 28.54 | 30.93 |
| Wind, m s-1 | 2.69 (0.46) | 1.34 | 3.77 | 1.94 | 2.39 | 2.72 | 2.99 | 3.44 |

| | | | | | | | | |
|---|---|---|---|---|---|---|---|---|
| PM10, µg m-3 | 40.06 (16.75) | 17.47 | 122.98 | 23.19 | 29.35 | 35.53 | 46.2 | 71.81 |
| DustyDays | 17 (10) | 1 | 31 | 2 | 8 | 18 | 27 | 30 |
| Winter | | | | | | | | |
| CM | 176 (241) | 3 | 1178 | 8 | 37 | 78 | 183 | 736 |
| CM incidence | 9.47 (6.05) | 1.09 | 30.68 | 2.24 | 5.15 | 8.19 | 11.74 | 21.34 |
| SM0t10, kg m-2 | 17.71 (2.87) | 10.87 | 23.9 | 12.63 | 15.65 | 18.13 | 19.86 | 21.97 |
| SM10t40, kg m-2 | 57.26 (6.17) | 42.02 | 72.19 | 47.2 | 52.69 | 57.29 | 61.6 | 67.62 |
| SM40t100, kg m-2 | 113.24 (11.18) | 85.59 | 143.62 | 96.21 | 105.77 | 111.66 | 120.69 | 132.71 |
| ST0t10, K | 282.99 (2.75) | 278.03 | 290.21 | 279.38 | 280.67 | 282.24 | 285.43 | 287.63 |
| ST10t40, K | 282.97 (2.13) | 278.98 | 288.44 | 279.98 | 281.21 | 282.46 | 284.79 | 286.59 |
| ST40t100, K | 282.99 (1.48) | 280.21 | 287.05 | 280.91 | 281.84 | 282.68 | 284.04 | 285.78 |
| Prep, mm | 23.42 (22.64) | 0 | 123.91 | 0.14 | 5.24 | 18.1 | 33.77 | 66.33 |
| AT, °C | 13.10 (2.48) | 8.17 | 20.11 | 9.58 | 11.12 | 12.85 | 14.82 | 17.42 |
| Wind, m s-1 | 1.08 (0.61) | 0.06 | 2.82 | 0.21 | 0.6 | 1 | 1.5 | 2.24 |
| PM10, µg m-3 | 30.61 (14.49) | 13.05 | 118.81 | 15.71 | 21.03 | 27.18 | 35.6 | 55.36 |
| DustyDays | 12 (9) | 0 | 31 | 1 | 4 | 10 | 19 | 27 |

**Abbreviations**: CM = coccidioidomycosis monthly case count; CM incidence = coccidioidomycosis incidence per 100,000 population; SM0t10 = soil moisture at 0–10 cm depth; SM10t40 = soil moisture at 10–40 cm depth; SM40t100 = soil moisture at 40–100 cm depth; ST0t10 = soil temperature at 0–10 cm depth; ST10t40 = soil temperature at 10–40 cm depth; ST40t100 = soil temperature at 40–100 cm depth; Prep = precipitation; AT = air temperature; Wind = wind speed at 10 m above ground; $PM_{10}$ = particulate matter ≤10 µm concentration; DustyDays = number of days per month with PM10 > 45 µg $m^{-3}$ at any monitoring site within the county;[8] SD = standard deviation; N = number of valid (non-missing) county-month observations; % missing = percentage of county-month observations with missing values.

**Note:** All statistics were calculated at the county-month level. For CM and CM incidence, the study period was 1997–2024; for environmental variables, the study period was 1994–2024. For example, the mean CM represents the average monthly case count per county across the tri-county region during 1997–2024, and the mean Prep represents the average monthly precipitation per county across the tri-county region during 1994–2024. The 5th, 25th, 50th, 75th, and 95th columns represent the corresponding percentiles of the distribution. Following the seasonal classification established in our earlier study,[8] months were grouped into winter (January–March), spring (April–June), monsoon (July–September), and fall (October–December) to reflect region-specific disease incidence patterns and climatic conditions.

**Table S3.** Season-specific lagged associations between coccidioidomycosis incidence and below-ground (Layer 3: 0–10 cm; topsoil) soil moisture (SM0t10) and soil temperature (ST0t10).

| Variable | Lag | Winter_IRR_CI | Spring_IRR_CI | Monsoon_IRR_CI | Fall_IRR_CI | Winter_IQR | Spring_IQR | Monsoon_IQR | Fall_IQR |
|---|---|---|---|---|---|---|---|---|---|
| SM0t10, kg m-2 | 1 | 0.80 (0.74, 0.87) | 0.79 (0.69, 0.91) | 0.72 (0.63, 0.83) | 0.93 (0.87, 0.99) | [16.20, 20.23] | [9.32, 15.00] | [8.18, 15.59] | [11.10, 15.72] |
| | 2 | 0.99 (0.91, 1.08) | 0.74 (0.66, 0.82) | 1.04 (0.96, 1.14) | 0.83 (0.78, 0.88) | [13.03, 19.49] | [12.45, 18.48] | [7.80, 11.15] | [12.04, 16.89] |
| | 3 | 1.06 (0.98, 1.16) | 0.95 (0.89, 1.01) | 1.07 (0.99, 1.15) | 0.81 (0.77, 0.86) | [11.07, 17.12] | [15.37, 19.83] | [7.88, 11.22] | [12.47, 16.64] |
| | 4 | 0.84 (0.78, 0.90) | 1.01 (0.94, 1.08) | 1.03 (0.90, 1.19) | 0.91 (0.81, 1.03) | [11.10, 15.72] | [16.20, 20.23] | [9.32, 15.00] | [8.18, 15.59] |
| | 5 | 1.01 (0.95, 1.08) | 1.20 (1.09, 1.32) | 0.87 (0.78, 0.97) | 0.90 (0.84, 0.96) | [12.04, 16.89] | [13.03, 19.49] | [12.45, 18.48] | [7.80, 11.15] |
| | 6 | 1.05 (0.99, 1.12) | 1.09 (1.01, 1.18) | 1.04 (0.97, 1.12) | 0.85 (0.79, 0.91) | [12.47, 16.64] | [11.07, 17.12] | [15.37, 19.83] | [7.88, 11.22] |
| | 7 | 0.88 (0.76, 1.01) | 0.92 (0.85, 0.99) | 1.13 (1.05, 1.21) | 0.62 (0.54, 0.70) | [8.18, 15.59] | [11.10, 15.72] | [16.20, 20.23] | [9.32, 15.00] |
| | 8 | 0.84 (0.77, 0.91) | 0.94 (0.88, 1.00) | 1.22 (1.12, 1.34) | 0.96 (0.88, 1.06) | [7.80, 11.15] | [12.04, 16.89] | [13.03, 19.49] | [12.45, 18.48] |
| | 9 | 0.85 (0.78, 0.93) | 0.99 (0.93, 1.05) | 1.03 (0.95, 1.11) | 1.22 (1.15, 1.30) | [7.88, 11.22] | [12.47, 16.64] | [11.07, 17.12] | [15.37, 19.83] |
| | 10 | 0.87 (0.75, 1.00) | 1.03 (0.90, 1.18) | 0.95 (0.89, 1.01) | 0.94 (0.88, 1.01) | [9.32, 15.00] | [8.18, 15.59] | [11.10, 15.72] | [16.20, 20.23] |
| | 11 | 1.06 (0.94, 1.19) | 0.90 (0.84, 0.97) | 0.97 (0.92, 1.03) | 1.21 (1.12, 1.31) | [12.45, 18.48] | [7.80, 11.15] | [12.04, 16.89] | [13.03, 19.49] |
| | 12 | 1.14 (1.06, 1.22) | 0.86 (0.79, 0.94) | 1.03 (0.97, 1.09) | 1.35 (1.24, 1.47) | [15.37, 19.83] | [7.88, 11.22] | [12.47, 16.64] | [11.07, 17.12] |
| | 13 | 1.04 (0.95, 1.12) | 0.87 (0.76, 1.00) | 0.85 (0.76, 0.96) | 1.02 (0.96, 1.09) | [16.20, 20.23] | [9.32, 15.00] | [8.18, 15.59] | [11.10, 15.72] |
| | 14 | 1.07 (0.98, 1.18) | 1.04 (0.93, 1.16) | 0.92 (0.86, 0.99) | 0.89 (0.84, 0.94) | [13.03, 19.49] | [12.45, 18.48] | [7.80, 11.15] | [12.04, 16.89] |
| | 15 | 1.07 (0.98, 1.18) | 1.10 (1.02, 1.20) | 0.90 (0.83, 0.98) | 1.00 (0.95, 1.06) | [11.07, 17.12] | [15.37, 19.83] | [7.88, 11.22] | [12.47, 16.64] |
| | 16 | 0.91 (0.85, 0.97) | 1.02 (0.95, 1.10) | 0.76 (0.66, 0.87) | 1.10 (0.99, 1.22) | [11.10, 15.72] | [16.20, 20.23] | [9.32, 15.00] | [8.18, 15.59] |
| | 17 | 0.94 (0.89, 1.00) | 0.91 (0.83, 1.00) | 0.90 (0.81, 1.00) | 1.07 (1.01, 1.14) | [12.04, 16.89] | [13.03, 19.49] | [12.45, 18.48] | [7.80, 11.15] |
| | 18 | 1.09 (1.03, 1.16) | 0.96 (0.88, 1.05) | 1.09 (1.01, 1.18) | 0.97 (0.89, 1.05) | [12.47, 16.64] | [11.07, 17.12] | [15.37, 19.83] | [7.88, 11.22] |
| | 19 | 1.29 (1.15, 1.45) | 0.94 (0.88, 1.00) | 1.03 (0.96, 1.10) | 0.88 (0.77, 1.01) | [8.18, 15.59] | [11.10, 15.72] | [16.20, 20.23] | [9.32, 15.00] |
| | 20 | 1.03 (0.97, 1.11) | 1.01 (0.95, 1.08) | 1.04 (0.96, 1.14) | 0.90 (0.82, 0.99) | [7.80, 11.15] | [12.04, 16.89] | [13.03, 19.49] | [12.45, 18.48] |
| | 21 | 0.87 (0.80, 0.95) | 1.01 (0.95, 1.08) | 1.03 (0.93, 1.13) | 0.93 (0.86, 1.01) | [7.88, 11.22] | [12.47, 16.64] | [11.07, 17.12] | [15.37, 19.83] |
| | 22 | 0.92 (0.81, 1.05) | 0.98 (0.87, 1.11) | 0.93 (0.87, 0.99) | 1.00 (0.94, 1.06) | [9.32, 15.00] | [8.18, 15.59] | [11.10, 15.72] | [16.20, 20.23] |
| | 23 | 0.85 (0.77, 0.94) | 0.97 (0.91, 1.04) | 0.99 (0.93, 1.05) | 0.92 (0.85, 1.00) | [12.45, 18.48] | [7.80, 11.15] | [12.04, 16.89] | [13.03, 19.49] |
| | 24 | 0.89 (0.82, 0.98) | 0.89 (0.81, 0.98) | 0.98 (0.92, 1.04) | 0.87 (0.80, 0.95) | [15.37, 19.83] | [7.88, 11.22] | [12.47, 16.64] | [11.07, 17.12] |
| | 25 | 0.99 (0.93, 1.06) | 0.95 (0.84, 1.07) | 0.76 (0.69, 0.85) | 1.05 (1.00, 1.11) | [16.20, 20.23] | [9.32, 15.00] | [8.18, 15.59] | [11.10, 15.72] |
| | 26 | 1.08 (0.98, 1.18) | 1.08 (0.97, 1.19) | 1.09 (1.02, 1.17) | 1.08 (1.01, 1.14) | [13.03, 19.49] | [12.45, 18.48] | [7.80, 11.15] | [12.04, 16.89] |
| | 27 | 1.10 (1.01, 1.21) | 0.94 (0.87, 1.02) | 1.07 (0.99, 1.16) | 0.87 (0.82, 0.92) | [11.07, 17.12] | [15.37, 19.83] | [7.88, 11.22] | [12.47, 16.64] |
| | 28 | 0.99 (0.93, 1.05) | 0.92 (0.87, 0.98) | 0.84 (0.74, 0.95) | 0.77 (0.70, 0.84) | [11.10, 15.72] | [16.20, 20.23] | [9.32, 15.00] | [8.18, 15.59] |
| | 29 | 1.01 (0.95, 1.09) | 0.98 (0.89, 1.08) | 0.91 (0.82, 1.00) | 1.02 (0.96, 1.08) | [12.04, 16.89] | [13.03, 19.49] | [12.45, 18.48] | [7.80, 11.15] |
| | 30 | 0.94 (0.88, 1.01) | 0.96 (0.87, 1.06) | 0.87 (0.81, 0.94) | 0.97 (0.89, 1.05) | [12.47, 16.64] | [11.07, 17.12] | [15.37, 19.83] | [7.88, 11.22] |
| | 31 | 0.90 (0.81, 1.00) | 0.97 (0.92, 1.03) | 0.91 (0.86, 0.96) | 0.78 (0.69, 0.88) | [8.18, 15.59] | [11.10, 15.72] | [16.20, 20.23] | [9.32, 15.00] |
| | 32 | 1.04 (0.97, 1.11) | 0.98 (0.91, 1.06) | 0.93 (0.85, 1.02) | 0.70 (0.63, 0.78) | [7.80, 11.15] | [12.04, 16.89] | [13.03, 19.49] | [12.45, 18.48] |
| | 33 | 1.10 (0.99, 1.21) | 0.97 (0.90, 1.04) | 0.91 (0.83, 1.00) | 0.95 (0.89, 1.02) | [7.88, 11.22] | [12.47, 16.64] | [11.07, 17.12] | [15.37, 19.83] |
| | 34 | 0.90 (0.81, 1.01) | 0.88 (0.80, 0.97) | 1.06 (1.01, 1.12) | 1.05 (1.00, 1.11) | [9.32, 15.00] | [8.18, 15.59] | [11.10, 15.72] | [16.20, 20.23] |
| | 35 | 0.98 (0.85, 1.12) | 0.99 (0.93, 1.06) | 1.12 (1.04, 1.20) | 1.04 (0.95, 1.14) | [12.45, 18.48] | [7.80, 11.15] | [12.04, 16.89] | [13.03, 19.49] |
| | 36 | 1.01 (0.91, 1.11) | 1.06 (0.96, 1.16) | 0.90 (0.84, 0.96) | 1.16 (1.04, 1.29) | [15.37, 19.83] | [7.88, 11.22] | [12.47, 16.64] | [11.07, 17.12] |
| ST0t10, K | 1 | 0.82 (0.77, 0.88) | 0.99 (0.75, 1.29) | 0.96 (0.84, 1.10) | 0.78 (0.56, 1.08) | [279.94, 281.82] | [286.92, 293.92] | [301.32, 305.13] | [286.46, 299.49] |
| | 2 | 0.94 (0.82, 1.08) | 1.03 (0.88, 1.21) | 1.43 (1.14, 1.79) | 1.21 (0.96, 1.53) | [279.98, 284.33] | [283.05, 289.36] | [296.05, 303.49] | [294.05, 303.31] |
| | 3 | 1.33 (0.95, 1.87) | 1.21 (1.10, 1.33) | 1.19 (0.94, 1.51) | 1.01 (0.90, 1.14) | [281.23, 291.92] | [280.65, 285.20] | [291.08, 299.30] | [301.56, 305.13] |
| | 4 | 1.61 (1.03, 2.53) | 1.12 (1.06, 1.18) | 0.73 (0.62, 0.86) | 1.22 (1.10, 1.35) | [286.46, 299.49] | [279.94, 281.82] | [286.92, 293.92] | [301.32, 305.13] |

| | | | | | | | | |
|---|---|---|---|---|---|---|---|---|
| 5 | 1.35 (1.04, 1.74) | 1.24 (1.09, 1.42) | 0.75 (0.64, 0.88) | 2.56 (2.08, 3.15) | [294.05, 303.31] | [279.98, 284.33] | [283.05, 289.36] | [296.05, 303.49] |
| 6 | 0.94 (0.81, 1.09) | 1.40 (0.98, 1.99) | 0.94 (0.85, 1.05) | 1.34 (1.05, 1.70) | [301.56, 305.13] | [281.23, 291.92] | [280.65, 285.20] | [291.08, 299.30] |
| 7 | 1.06 (0.92, 1.23) | 2.17 (1.41, 3.32) | 1.07 (1.02, 1.12) | 0.50 (0.41, 0.62) | [301.32, 305.13] | [286.46, 299.49] | [279.94, 281.82] | [286.92, 293.92] |
| 8 | 1.43 (1.09, 1.87) | 2.35 (1.76, 3.13) | 1.33 (1.18, 1.49) | 0.70 (0.59, 0.84) | [296.05, 303.49] | [294.05, 303.31] | [279.98, 284.33] | [283.05, 289.36] |
| 9 | 0.75 (0.58, 0.97) | 1.38 (1.21, 1.58) | 1.91 (1.37, 2.65) | 1.23 (1.12, 1.35) | [291.08, 299.30] | [301.56, 305.13] | [281.23, 291.92] | [280.65, 285.20] |
| 10 | 0.47 (0.35, 0.63) | 1.39 (1.19, 1.62) | 2.31 (1.53, 3.49) | 1.22 (1.14, 1.29) | [286.92, 293.92] | [301.32, 305.13] | [286.46, 299.49] | [279.94, 281.82] |
| 11 | 0.75 (0.61, 0.91) | 1.54 (1.22, 1.94) | 1.54 (1.14, 2.08) | 1.53 (1.35, 1.73) | [283.05, 289.36] | [296.05, 303.49] | [294.05, 303.31] | [279.98, 284.33] |
| 12 | 0.95 (0.84, 1.06) | 0.79 (0.60, 1.04) | 1.20 (1.07, 1.36) | 1.92 (1.41, 2.62) | [280.65, 285.20] | [291.08, 299.30] | [301.56, 305.13] | [281.23, 291.92] |
| 13 | 0.96 (0.91, 1.02) | 0.53 (0.41, 0.69) | 1.11 (0.96, 1.28) | 2.67 (1.83, 3.89) | [279.94, 281.82] | [286.92, 293.92] | [301.32, 305.13] | [286.46, 299.49] |
| 14 | 1.08 (0.97, 1.20) | 0.65 (0.53, 0.79) | 1.00 (0.79, 1.27) | 1.56 (1.19, 2.05) | [279.98, 284.33] | [283.05, 289.36] | [296.05, 303.49] | [294.05, 303.31] |
| 15 | 1.70 (1.26, 2.31) | 0.88 (0.78, 0.99) | 1.13 (0.88, 1.44) | 1.29 (1.13, 1.46) | [281.23, 291.92] | [280.65, 285.20] | [291.08, 299.30] | [301.56, 305.13] |
| 16 | 0.93 (0.63, 1.38) | 1.01 (0.96, 1.06) | 0.68 (0.55, 0.85) | 1.05 (0.93, 1.18) | [286.46, 299.49] | [279.94, 281.82] | [286.92, 293.92] | [301.32, 305.13] |
| 17 | 0.91 (0.67, 1.24) | 1.14 (1.01, 1.28) | 0.67 (0.55, 0.82) | 0.77 (0.62, 0.94) | [294.05, 303.31] | [279.98, 284.33] | [283.05, 289.36] | [296.05, 303.49] |
| 18 | 1.28 (1.12, 1.47) | 0.86 (0.65, 1.15) | 1.11 (0.99, 1.25) | 1.16 (0.94, 1.44) | [301.56, 305.13] | [281.23, 291.92] | [280.65, 285.20] | [291.08, 299.30] |
| 19 | 1.16 (1.00, 1.34) | 0.14 (0.09, 0.22) | 1.10 (1.04, 1.15) | 1.01 (0.79, 1.30) | [301.32, 305.13] | [286.46, 299.49] | [279.94, 281.82] | [286.92, 293.92] |
| 20 | 1.22 (0.97, 1.53) | 0.47 (0.35, 0.62) | 1.13 (1.01, 1.27) | 0.70 (0.59, 0.84) | [296.05, 303.49] | [294.05, 303.31] | [279.98, 284.33] | [283.05, 289.36] |
| 21 | 1.24 (0.97, 1.57) | 1.28 (1.13, 1.45) | 0.73 (0.55, 0.96) | 0.95 (0.86, 1.06) | [291.08, 299.30] | [301.56, 305.13] | [281.23, 291.92] | [280.65, 285.20] |
| 22 | 1.21 (0.88, 1.67) | 1.41 (1.21, 1.63) | 0.34 (0.21, 0.57) | 1.12 (1.06, 1.18) | [286.92, 293.92] | [301.32, 305.13] | [286.46, 299.49] | [279.94, 281.82] |
| 23 | 1.24 (0.99, 1.55) | 1.46 (1.19, 1.80) | 0.71 (0.51, 0.98) | 1.12 (1.01, 1.25) | [283.05, 289.36] | [296.05, 303.49] | [294.05, 303.31] | [279.98, 284.33] |
| 24 | 1.19 (1.05, 1.35) | 0.62 (0.49, 0.80) | 1.31 (1.18, 1.47) | 0.33 (0.25, 0.44) | [280.65, 285.20] | [291.08, 299.30] | [301.56, 305.13] | [281.23, 291.92] |
| 25 | 1.14 (1.08, 1.21) | 0.58 (0.45, 0.75) | 1.40 (1.24, 1.59) | 0.14 (0.10, 0.20) | [279.94, 281.82] | [286.92, 293.92] | [301.32, 305.13] | [286.46, 299.49] |
| 26 | 1.42 (1.26, 1.60) | 1.01 (0.82, 1.24) | 0.98 (0.79, 1.20) | 0.81 (0.64, 1.04) | [279.98, 284.33] | [283.05, 289.36] | [296.05, 303.49] | [294.05, 303.31] |
| 27 | 0.71 (0.50, 1.00) | 1.24 (1.10, 1.39) | 0.52 (0.40, 0.68) | 1.39 (1.24, 1.57) | [281.23, 291.92] | [280.65, 285.20] | [291.08, 299.30] | [301.56, 305.13] |
| 28 | 0.29 (0.18, 0.48) | 1.18 (1.12, 1.24) | 0.85 (0.64, 1.11) | 1.11 (0.97, 1.26) | [286.46, 299.49] | [279.94, 281.82] | [286.92, 293.92] | [301.32, 305.13] |
| 29 | 0.84 (0.60, 1.16) | 1.34 (1.19, 1.52) | 1.10 (0.89, 1.37) | 0.67 (0.55, 0.80) | [294.05, 303.31] | [279.98, 284.33] | [283.05, 289.36] | [296.05, 303.49] |
| 30 | 1.31 (1.15, 1.49) | 0.62 (0.44, 0.89) | 1.08 (0.97, 1.22) | 0.53 (0.43, 0.65) | [301.56, 305.13] | [281.23, 291.92] | [280.65, 285.20] | [291.08, 299.30] |
| 31 | 1.26 (1.09, 1.45) | 0.59 (0.37, 0.95) | 1.08 (1.02, 1.14) | 0.68 (0.53, 0.86) | [301.32, 305.13] | [286.46, 299.49] | [279.94, 281.82] | [286.92, 293.92] |
| 32 | 0.75 (0.62, 0.91) | 0.99 (0.73, 1.35) | 1.04 (0.94, 1.15) | 0.94 (0.81, 1.09) | [296.05, 303.49] | [294.05, 303.31] | [279.98, 284.33] | [283.05, 289.36] |
| 33 | 0.91 (0.75, 1.11) | 1.09 (0.94, 1.26) | 0.46 (0.35, 0.61) | 1.08 (0.98, 1.19) | [291.08, 299.30] | [301.56, 305.13] | [281.23, 291.92] | [280.65, 285.20] |
| 34 | 1.13 (0.89, 1.42) | 1.00 (0.86, 1.16) | 0.35 (0.22, 0.58) | 1.06 (1.01, 1.11) | [286.92, 293.92] | [301.32, 305.13] | [286.46, 299.49] | [279.94, 281.82] |
| 35 | 0.76 (0.64, 0.91) | 0.79 (0.66, 0.94) | 1.11 (0.77, 1.59) | 1.09 (1.00, 1.18) | [283.05, 289.36] | [296.05, 303.49] | [294.05, 303.31] | [279.98, 284.33] |
| 36 | 1.18 (1.00, 1.38) | 0.64 (0.51, 0.80) | 0.96 (0.84, 1.09) | 0.80 (0.60, 1.08) | [280.65, 285.20] | [291.08, 299.30] | [301.56, 305.13] | [281.23, 291.92] |

Incidence rate ratios (IRRs) and 95% confidence intervals (CIs) were estimated in the study region using distributed lag non-linear models (DLNMs) for an interquartile range (IQR) increase in each exposure across lags 1–36 months prior to disease incidence, stratified by season. Column notation [Season]_IRR_CI (e.g., Fall_IRR_CI) indicates IRRs (95% CIs) for that season's incidence per IQR increase in exposures at corresponding lags; IQRs were calculated separately for each season-lag combination (e.g., Fall_IQR reflects the exposure range at lag months preceding fall incidence); this format applies to all DLNM results. Models were adjusted for spatiotemporal trends and surveillance-related changes in reporting and laboratory testing. Layer 3 soil moisture and soil temperature were modeled simultaneously in a single DLNM with season-specific effects.

**Table S4.** Season-specific lagged associations between coccidioidomycosis incidence and below-ground (Layer 4: 10–40 cm) soil moisture (SM10t40) and soil temperature (ST10t40).

| Variable | Lag | Winter_IRR_CI | Spring_IRR_CI | Monsoon_IRR_CI | Fall_IRR_CI | Winter_IQR | Spring_IQR | Monsoon_IQR | Fall_IQR |
|---|---|---|---|---|---|---|---|---|---|
| SM10t40, kg m-2 | 1 | 0.84 (0.75, 0.94) | 0.80 (0.70, 0.91) | 0.91 (0.83, 1.01) | 0.82 (0.75, 0.90) | [51.43, 60.43] | [51.16, 57.61] | [50.25, 55.53] | [50.53, 56.20] |
| | 2 | 0.98 (0.90, 1.06) | 0.88 (0.82, 0.95) | 0.98 (0.91, 1.05) | 0.90 (0.84, 0.96) | [50.51, 58.59] | [52.36, 59.91] | [49.53, 54.20] | [51.29, 57.90] |
| | 3 | 1.04 (0.96, 1.12) | 1.01 (0.91, 1.12) | 0.98 (0.88, 1.08) | 0.98 (0.90, 1.07) | [49.94, 55.71] | [52.63, 61.19] | [49.92, 54.97] | [51.17, 57.40] |
| | 4 | 0.95 (0.90, 1.01) | 1.09 (1.00, 1.18) | 0.97 (0.91, 1.05) | 0.98 (0.92, 1.04) | [50.53, 56.20] | [51.43, 60.43] | [51.16, 57.61] | [50.25, 55.53] |
| | 5 | 0.92 (0.84, 1.01) | 1.12 (1.00, 1.25) | 1.04 (0.93, 1.17) | 0.92 (0.84, 1.00) | [51.29, 57.90] | [50.51, 58.59] | [52.36, 59.91] | [49.53, 54.20] |
| | 6 | 1.05 (0.97, 1.12) | 1.05 (0.99, 1.12) | 1.22 (1.12, 1.33) | 0.88 (0.83, 0.94) | [51.17, 57.40] | [49.94, 55.71] | [52.63, 61.19] | [49.92, 54.97] |
| | 7 | 1.07 (0.99, 1.16) | 0.98 (0.91, 1.06) | 1.31 (1.16, 1.49) | 1.00 (0.90, 1.11) | [50.25, 55.53] | [50.53, 56.20] | [51.43, 60.43] | [51.16, 57.61] |
| | 8 | 0.88 (0.82, 0.95) | 0.94 (0.87, 1.01) | 1.07 (0.99, 1.16) | 1.21 (1.11, 1.31) | [49.53, 54.20] | [51.29, 57.90] | [50.51, 58.59] | [52.36, 59.91] |
| | 9 | 0.79 (0.72, 0.87) | 0.98 (0.90, 1.06) | 0.94 (0.87, 1.01) | 1.23 (1.11, 1.37) | [49.92, 54.97] | [51.17, 57.40] | [49.94, 55.71] | [52.63, 61.19] |
| | 10 | 0.99 (0.91, 1.06) | 1.03 (0.97, 1.11) | 0.98 (0.92, 1.04) | 1.07 (0.98, 1.17) | [51.16, 57.61] | [50.25, 55.53] | [50.53, 56.20] | [51.43, 60.43] |
| | 11 | 1.20 (1.06, 1.36) | 0.97 (0.90, 1.05) | 1.04 (0.96, 1.11) | 0.98 (0.89, 1.07) | [52.36, 59.91] | [49.53, 54.20] | [51.29, 57.90] | [50.51, 58.59] |
| | 12 | 1.10 (1.01, 1.19) | 0.85 (0.78, 0.91) | 1.08 (1.01, 1.16) | 1.12 (1.05, 1.18) | [52.63, 61.19] | [49.92, 54.97] | [51.17, 57.40] | [49.94, 55.71] |
| | 13 | 0.98 (0.89, 1.09) | 0.85 (0.76, 0.95) | 1.00 (0.93, 1.06) | 1.11 (1.05, 1.18) | [51.43, 60.43] | [51.16, 57.61] | [50.25, 55.53] | [50.53, 56.20] |
| | 14 | 0.94 (0.86, 1.02) | 1.12 (1.02, 1.22) | 0.91 (0.85, 0.98) | 0.94 (0.88, 1.01) | [50.51, 58.59] | [52.36, 59.91] | [49.53, 54.20] | [51.29, 57.90] |
| | 15 | 0.95 (0.89, 1.01) | 1.19 (1.06, 1.32) | 0.92 (0.84, 1.01) | 0.90 (0.84, 0.96) | [49.94, 55.71] | [52.63, 61.19] | [49.92, 54.97] | [51.17, 57.40] |
| | 16 | 1.03 (0.96, 1.10) | 0.91 (0.83, 0.99) | 1.08 (0.99, 1.18) | 1.03 (0.97, 1.11) | [50.53, 56.20] | [51.43, 60.43] | [51.16, 57.61] | [50.25, 55.53] |
| | 17 | 1.06 (0.98, 1.15) | 0.81 (0.74, 0.88) | 1.19 (1.06, 1.34) | 1.08 (1.01, 1.15) | [51.29, 57.90] | [50.51, 58.59] | [52.36, 59.91] | [49.53, 54.20] |
| | 18 | 1.09 (1.01, 1.17) | 0.98 (0.91, 1.06) | 1.00 (0.91, 1.10) | 0.95 (0.88, 1.03) | [51.17, 57.40] | [49.94, 55.71] | [52.63, 61.19] | [49.92, 54.97] |
| | 19 | 0.96 (0.90, 1.02) | 1.09 (1.02, 1.16) | 0.95 (0.87, 1.05) | 0.92 (0.83, 1.01) | [50.25, 55.53] | [50.53, 56.20] | [51.43, 60.43] | [51.16, 57.61] |
| | 20 | 0.89 (0.81, 0.97) | 1.04 (0.95, 1.13) | 1.04 (0.95, 1.14) | 0.92 (0.84, 1.00) | [49.53, 54.20] | [51.29, 57.90] | [50.51, 58.59] | [52.36, 59.91] |
| | 21 | 0.89 (0.82, 0.96) | 1.01 (0.94, 1.08) | 1.02 (0.96, 1.08) | 0.93 (0.84, 1.03) | [49.92, 54.97] | [51.17, 57.40] | [49.94, 55.71] | [52.63, 61.19] |
| | 22 | 0.96 (0.88, 1.05) | 1.05 (0.96, 1.14) | 0.98 (0.91, 1.06) | 0.96 (0.87, 1.05) | [51.16, 57.61] | [50.25, 55.53] | [50.53, 56.20] | [51.43, 60.43] |
| | 23 | 1.03 (0.92, 1.14) | 1.04 (0.97, 1.11) | 0.92 (0.86, 0.99) | 0.91 (0.84, 0.98) | [52.36, 59.91] | [49.53, 54.20] | [51.29, 57.90] | [50.51, 58.59] |
| | 24 | 0.96 (0.88, 1.06) | 1.00 (0.92, 1.08) | 1.00 (0.92, 1.08) | 0.94 (0.87, 1.00) | [52.63, 61.19] | [49.92, 54.97] | [51.17, 57.40] | [49.94, 55.71] |
| | 25 | 0.98 (0.90, 1.07) | 0.98 (0.90, 1.08) | 0.98 (0.93, 1.05) | 0.99 (0.93, 1.04) | [51.43, 60.43] | [51.16, 57.61] | [50.25, 55.53] | [50.53, 56.20] |
| | 26 | 1.06 (0.96, 1.17) | 0.97 (0.88, 1.06) | 0.98 (0.90, 1.07) | 1.04 (0.96, 1.13) | [50.51, 58.59] | [52.36, 59.91] | [49.53, 54.20] | [51.29, 57.90] |
| | 27 | 1.06 (1.00, 1.13) | 0.93 (0.85, 1.01) | 0.91 (0.85, 0.97) | 0.99 (0.93, 1.05) | [49.94, 55.71] | [52.63, 61.19] | [49.92, 54.97] | [51.17, 57.40] |
| | 28 | 1.03 (0.95, 1.11) | 0.91 (0.82, 1.01) | 0.89 (0.82, 0.98) | 0.92 (0.86, 1.00) | [50.53, 56.20] | [51.43, 60.43] | [51.16, 57.61] | [50.25, 55.53] |
| | 29 | 0.94 (0.88, 1.01) | 0.92 (0.84, 1.00) | 0.99 (0.91, 1.08) | 0.96 (0.91, 1.02) | [51.29, 57.90] | [50.51, 58.59] | [52.36, 59.91] | [49.53, 54.20] |
| | 30 | 0.95 (0.87, 1.04) | 0.97 (0.90, 1.04) | 0.99 (0.90, 1.08) | 1.03 (0.94, 1.12) | [51.17, 57.40] | [49.94, 55.71] | [52.63, 61.19] | [49.92, 54.97] |
| | 31 | 0.99 (0.94, 1.06) | 1.03 (0.98, 1.10) | 1.02 (0.94, 1.11) | 1.04 (0.96, 1.13) | [50.25, 55.53] | [50.53, 56.20] | [51.43, 60.43] | [51.16, 57.61] |
| | 32 | 1.06 (0.97, 1.16) | 1.09 (1.00, 1.19) | 1.04 (0.95, 1.14) | 0.96 (0.88, 1.06) | [49.53, 54.20] | [51.29, 57.90] | [50.51, 58.59] | [52.36, 59.91] |
| | 33 | 0.96 (0.91, 1.02) | 0.99 (0.92, 1.06) | 1.06 (1.01, 1.11) | 0.95 (0.87, 1.04) | [49.92, 54.97] | [51.17, 57.40] | [49.94, 55.71] | [52.63, 61.19] |
| | 34 | 0.87 (0.79, 0.97) | 0.91 (0.84, 0.99) | 1.14 (1.07, 1.22) | 1.00 (0.92, 1.10) | [51.16, 57.61] | [50.25, 55.53] | [50.53, 56.20] | [51.43, 60.43] |
| | 35 | 0.94 (0.86, 1.03) | 0.93 (0.88, 0.97) | 1.08 (1.01, 1.14) | 1.02 (0.96, 1.08) | [52.36, 59.91] | [49.53, 54.20] | [51.29, 57.90] | [50.51, 58.59] |
| | 36 | 1.01 (0.87, 1.17) | 0.96 (0.85, 1.07) | 0.94 (0.86, 1.03) | 1.01 (0.93, 1.10) | [52.63, 61.19] | [49.92, 54.97] | [51.17, 57.40] | [49.94, 55.71] |
| ST10t40, K | 1 | 0.89 (0.82, 0.97) | 1.22 (0.87, 1.71) | 0.83 (0.65, 1.06) | 0.56 (0.36, 0.89) | [281.00, 282.56] | [286.04, 291.96] | [298.40, 303.36] | [288.55, 299.73] |
| | 2 | 0.99 (0.81, 1.22) | 1.25 (1.00, 1.56) | 1.15 (0.85, 1.57) | 0.61 (0.48, 0.79) | [281.18, 286.66] | [283.01, 288.18] | [293.96, 301.41] | [295.51, 302.55] |
| | 3 | 1.69 (1.08, 2.63) | 1.02 (0.89, 1.18) | 1.35 (0.99, 1.83) | 1.01 (0.92, 1.11) | [283.03, 293.76] | [281.18, 284.60] | [289.76, 296.57] | [301.01, 303.40] |
| | 4 | 1.01 (0.64, 1.59) | 0.98 (0.92, 1.04) | 1.02 (0.78, 1.33) | 0.97 (0.84, 1.13) | [288.55, 299.73] | [281.00, 282.56] | [286.04, 291.96] | [298.40, 303.36] |

| | | | | | | | | |
|---|---|---|---|---|---|---|---|---|
| 5 | 0.81 (0.61, 1.08) | 1.10 (0.88, 1.38) | 1.13 (0.92, 1.38) | 0.90 (0.67, 1.21) | [295.51, 302.55] | [281.18, 286.66] | [283.01, 288.18] | [293.96, 301.41] |
| 6 | 0.91 (0.82, 1.01) | 1.65 (1.06, 2.58) | 1.18 (1.03, 1.35) | 0.75 (0.55, 1.02) | [301.01, 303.40] | [283.03, 293.76] | [281.18, 284.60] | [289.76, 296.57] |
| 7 | 0.86 (0.70, 1.05) | 0.92 (0.54, 1.55) | 1.00 (0.93, 1.07) | 0.61 (0.44, 0.84) | [298.40, 303.36] | [288.55, 299.73] | [281.00, 282.56] | [286.04, 291.96] |
| 8 | 1.21 (0.88, 1.67) | 0.86 (0.62, 1.21) | 0.94 (0.75, 1.17) | 1.07 (0.89, 1.29) | [293.96, 301.41] | [295.51, 302.55] | [281.18, 286.66] | [283.01, 288.18] |
| 9 | 0.96 (0.71, 1.31) | 1.03 (0.93, 1.14) | 1.56 (1.02, 2.39) | 1.33 (1.17, 1.52) | [289.76, 296.57] | [301.01, 303.40] | [283.03, 293.76] | [281.18, 284.60] |
| 10 | 0.65 (0.46, 0.90) | 1.13 (0.90, 1.41) | 1.07 (0.63, 1.84) | 1.13 (1.05, 1.22) | [286.04, 291.96] | [298.40, 303.36] | [288.55, 299.73] | [281.00, 282.56] |
| 11 | 0.84 (0.65, 1.09) | 1.24 (0.89, 1.74) | 1.09 (0.80, 1.50) | 1.65 (1.32, 2.06) | [283.01, 288.18] | [293.96, 301.41] | [295.51, 302.55] | [281.18, 286.66] |
| 12 | 0.91 (0.78, 1.08) | 0.92 (0.65, 1.29) | 1.02 (0.92, 1.12) | 1.43 (0.92, 2.21) | [281.18, 284.60] | [289.76, 296.57] | [301.01, 303.40] | [283.03, 293.76] |
| 13 | 0.97 (0.89, 1.05) | 0.50 (0.35, 0.69) | 0.92 (0.75, 1.11) | 0.87 (0.52, 1.46) | [281.00, 282.56] | [286.04, 291.96] | [298.40, 303.36] | [288.55, 299.73] |
| 14 | 1.20 (0.93, 1.55) | 0.54 (0.41, 0.71) | 0.84 (0.61, 1.15) | 1.26 (0.90, 1.76) | [281.18, 286.66] | [283.01, 288.18] | [293.96, 301.41] | [295.51, 302.55] |
| 15 | 1.23 (0.80, 1.90) | 0.99 (0.84, 1.17) | 0.65 (0.46, 0.92) | 1.00 (0.91, 1.10) | [283.03, 293.76] | [281.18, 284.60] | [289.76, 296.57] | [301.01, 303.40] |
| 16 | 0.44 (0.26, 0.75) | 1.11 (1.02, 1.21) | 0.56 (0.40, 0.78) | 1.17 (0.99, 1.38) | [288.55, 299.73] | [281.00, 282.56] | [286.04, 291.96] | [298.40, 303.36] |
| 17 | 0.72 (0.54, 0.96) | 1.39 (1.10, 1.77) | 0.95 (0.73, 1.23) | 1.45 (1.07, 1.98) | [295.51, 302.55] | [281.18, 286.66] | [283.01, 288.18] | [293.96, 301.41] |
| 18 | 1.06 (0.96, 1.18) | 0.98 (0.64, 1.51) | 1.25 (1.06, 1.48) | 0.83 (0.62, 1.10) | [301.01, 303.40] | [283.03, 293.76] | [281.18, 284.60] | [289.76, 296.57] |
| 19 | 1.02 (0.84, 1.23) | 0.27 (0.14, 0.49) | 1.05 (0.97, 1.14) | 0.67 (0.48, 0.95) | [298.40, 303.36] | [288.55, 299.73] | [281.00, 282.56] | [286.04, 291.96] |
| 20 | 0.84 (0.63, 1.12) | 0.59 (0.43, 0.81) | 0.94 (0.76, 1.15) | 0.87 (0.69, 1.10) | [293.96, 301.41] | [295.51, 302.55] | [281.18, 286.66] | [283.01, 288.18] |
| 21 | 0.87 (0.63, 1.20) | 1.07 (0.98, 1.17) | 0.64 (0.43, 0.97) | 1.11 (0.96, 1.28) | [289.76, 296.57] | [301.01, 303.40] | [283.03, 293.76] | [281.18, 284.60] |
| 22 | 1.05 (0.73, 1.51) | 1.38 (1.12, 1.70) | 0.29 (0.16, 0.53) | 1.08 (1.00, 1.17) | [286.04, 291.96] | [298.40, 303.36] | [288.55, 299.73] | [281.00, 282.56] |
| 23 | 1.05 (0.80, 1.38) | 1.15 (0.84, 1.58) | 0.64 (0.47, 0.88) | 0.99 (0.81, 1.21) | [283.01, 288.18] | [293.96, 301.41] | [295.51, 302.55] | [281.18, 286.66] |
| 24 | 1.07 (0.92, 1.26) | 0.56 (0.39, 0.81) | 1.04 (0.94, 1.14) | 0.38 (0.25, 0.57) | [281.18, 284.60] | [289.76, 296.57] | [301.01, 303.40] | [283.03, 293.76] |
| 25 | 1.08 (1.00, 1.17) | 0.69 (0.49, 0.98) | 1.05 (0.87, 1.27) | 0.33 (0.20, 0.54) | [281.00, 282.56] | [286.04, 291.96] | [298.40, 303.36] | [288.55, 299.73] |
| 26 | 1.32 (1.06, 1.65) | 1.03 (0.81, 1.29) | 0.75 (0.55, 1.03) | 0.88 (0.66, 1.18) | [281.18, 286.66] | [283.01, 288.18] | [293.96, 301.41] | [295.51, 302.55] |
| 27 | 0.70 (0.46, 1.06) | 1.09 (0.95, 1.26) | 0.67 (0.47, 0.96) | 1.06 (0.96, 1.17) | [283.03, 293.76] | [281.18, 284.60] | [289.76, 296.57] | [301.01, 303.40] |
| 28 | 0.62 (0.35, 1.10) | 1.09 (1.02, 1.17) | 0.85 (0.62, 1.16) | 1.07 (0.90, 1.27) | [288.55, 299.73] | [281.00, 282.56] | [286.04, 291.96] | [298.40, 303.36] |
| 29 | 0.93 (0.69, 1.25) | 1.23 (1.00, 1.51) | 0.89 (0.71, 1.11) | 0.65 (0.48, 0.87) | [295.51, 302.55] | [281.18, 286.66] | [283.01, 288.18] | [293.96, 301.41] |
| 30 | 1.06 (0.96, 1.17) | 0.65 (0.41, 1.05) | 1.11 (0.97, 1.28) | 0.54 (0.40, 0.75) | [301.01, 303.40] | [283.03, 293.76] | [281.18, 284.60] | [289.76, 296.57] |
| 31 | 1.13 (0.94, 1.36) | 0.63 (0.34, 1.16) | 1.13 (1.04, 1.22) | 0.78 (0.56, 1.09) | [298.40, 303.36] | [288.55, 299.73] | [281.00, 282.56] | [286.04, 291.96] |
| 32 | 0.76 (0.56, 1.04) | 0.93 (0.68, 1.28) | 0.86 (0.70, 1.05) | 0.91 (0.75, 1.10) | [293.96, 301.41] | [295.51, 302.55] | [281.18, 286.66] | [283.01, 288.18] |
| 33 | 0.90 (0.66, 1.23) | 1.02 (0.93, 1.11) | 0.62 (0.39, 0.98) | 1.09 (0.96, 1.23) | [289.76, 296.57] | [301.01, 303.40] | [283.03, 293.76] | [281.18, 284.60] |
| 34 | 0.96 (0.71, 1.30) | 0.88 (0.74, 1.05) | 2.21 (1.19, 4.10) | 1.07 (1.01, 1.14) | [286.04, 291.96] | [298.40, 303.36] | [288.55, 299.73] | [281.00, 282.56] |
| 35 | 0.87 (0.69, 1.09) | 0.75 (0.58, 0.98) | 1.17 (0.81, 1.68) | 1.06 (0.87, 1.28) | [283.01, 288.18] | [293.96, 301.41] | [295.51, 302.55] | [281.18, 286.66] |
| 36 | 1.16 (0.98, 1.37) | 0.83 (0.60, 1.15) | 0.93 (0.82, 1.05) | 0.80 (0.49, 1.33) | [281.18, 284.60] | [289.76, 296.57] | [301.01, 303.40] | [283.03, 293.76] |

IRRs and 95% CIs were estimated in the study region using DLNMs for an IQR increase in each exposure across lags 1–36 months prior to disease incidence, stratified by season. Models were adjusted for spatiotemporal trends and surveillance-related changes in reporting and laboratory testing. Layer 4 soil moisture and soil temperature were modeled simultaneously in a single DLNM with season-specific effects.

**Table S5.** Season-specific lagged associations between coccidioidomycosis incidence and below-ground (Layer 5: 40–100 cm) soil moisture (SM40t100) and soil temperature (ST40t100).

| Variable | Lag | Winter_IRR_CI | Spring_IRR_CI | Monsoon_IRR_CI | Fall_IRR_CI | Winter_IQR | Spring_IQR | Monsoon_IQR | Fall_IQR |
|---|---|---|---|---|---|---|---|---|---|
| SM40t100, kg m-2 | 1 | 1.01 (0.90, 1.13) | 0.85 (0.77, 0.94) | 0.82 (0.73, 0.93) | 0.81 (0.73, 0.89) | [102.39, 116.27] | [103.66, 117.04] | [98.80, 110.00] | [98.02, 110.87] |
| | 2 | 1.02 (0.98, 1.06) | 0.95 (0.91, 1.00) | 0.98 (0.93, 1.04) | 0.93 (0.89, 0.96) | [100.38, 111.50] | [105.23, 119.92] | [99.18, 110.25] | [98.70, 111.52] |
| | 3 | 1.04 (0.98, 1.10) | 1.03 (0.96, 1.10) | 1.13 (1.06, 1.20) | 1.03 (0.96, 1.10) | [97.85, 109.93] | [105.18, 119.28] | [101.05, 112.71] | [98.92, 110.87] |
| | 4 | 1.02 (0.96, 1.08) | 1.07 (1.00, 1.15) | 1.19 (1.10, 1.29) | 1.08 (1.01, 1.16) | [98.02, 110.87] | [102.39, 116.27] | [103.66, 117.04] | [98.80, 110.00] |
| | 5 | 0.98 (0.93, 1.02) | 1.02 (0.98, 1.06) | 1.10 (1.03, 1.17) | 1.07 (1.02, 1.12) | [98.70, 111.52] | [100.38, 111.50] | [105.23, 119.92] | [99.18, 110.25] |
| | 6 | 0.96 (0.89, 1.03) | 0.97 (0.92, 1.03) | 0.99 (0.93, 1.06) | 1.04 (0.97, 1.11) | [98.92, 110.87] | [97.85, 109.93] | [105.18, 119.28] | [101.05, 112.71] |
| | 7 | 1.00 (0.94, 1.07) | 0.98 (0.93, 1.04) | 0.96 (0.90, 1.02) | 1.04 (0.97, 1.12) | [98.80, 110.00] | [98.02, 110.87] | [102.39, 116.27] | [103.66, 117.04] |
| | 8 | 1.08 (1.02, 1.15) | 1.06 (1.00, 1.11) | 1.01 (0.96, 1.05) | 1.07 (1.00, 1.14) | [99.18, 110.25] | [98.70, 111.52] | [100.38, 111.50] | [105.23, 119.92] |
| | 9 | 1.12 (1.04, 1.22) | 1.09 (1.01, 1.17) | 1.06 (1.00, 1.13) | 1.06 (0.99, 1.14) | [101.05, 112.71] | [98.92, 110.87] | [97.85, 109.93] | [105.18, 119.28] |
| | 10 | 1.07 (0.99, 1.14) | 1.02 (0.96, 1.08) | 1.05 (1.00, 1.09) | 1.01 (0.96, 1.07) | [103.66, 117.04] | [98.80, 110.00] | [98.02, 110.87] | [102.39, 116.27] |
| | 11 | 0.97 (0.90, 1.04) | 0.93 (0.86, 1.00) | 0.99 (0.94, 1.04) | 1.01 (0.96, 1.06) | [105.23, 119.92] | [99.18, 110.25] | [98.70, 111.52] | [100.38, 111.50] |
| | 12 | 0.93 (0.86, 1.00) | 0.89 (0.82, 0.97) | 0.97 (0.91, 1.03) | 1.05 (1.00, 1.12) | [105.18, 119.28] | [101.05, 112.71] | [98.92, 110.87] | [97.85, 109.93] |
| | 13 | 0.92 (0.87, 0.98) | 0.95 (0.89, 1.01) | 0.99 (0.94, 1.05) | 1.04 (1.00, 1.08) | [102.39, 116.27] | [103.66, 117.04] | [98.80, 110.00] | [98.02, 110.87] |
| | 14 | 0.96 (0.91, 1.01) | 1.02 (0.95, 1.11) | 1.03 (0.95, 1.12) | 1.00 (0.95, 1.06) | [100.38, 111.50] | [105.23, 119.92] | [99.18, 110.25] | [98.70, 111.52] |
| | 15 | 1.00 (0.95, 1.06) | 1.06 (0.98, 1.15) | 1.06 (0.98, 1.14) | 0.98 (0.93, 1.03) | [97.85, 109.93] | [105.18, 119.28] | [101.05, 112.71] | [98.92, 110.87] |
| | 16 | 1.03 (0.99, 1.08) | 1.03 (0.97, 1.09) | 1.00 (0.95, 1.06) | 0.97 (0.91, 1.02) | [98.02, 110.87] | [102.39, 116.27] | [103.66, 117.04] | [98.80, 110.00] |
| | 17 | 1.05 (0.98, 1.12) | 0.98 (0.92, 1.03) | 0.93 (0.85, 1.02) | 0.96 (0.89, 1.04) | [98.70, 111.52] | [100.38, 111.50] | [105.23, 119.92] | [99.18, 110.25] |
| | 18 | 1.04 (0.99, 1.10) | 0.98 (0.94, 1.03) | 0.97 (0.90, 1.05) | 0.96 (0.90, 1.02) | [98.92, 110.87] | [97.85, 109.93] | [105.18, 119.28] | [101.05, 112.71] |
| | 19 | 1.01 (0.95, 1.08) | 1.02 (0.98, 1.07) | 1.06 (0.99, 1.13) | 0.97 (0.92, 1.03) | [98.80, 110.00] | [98.02, 110.87] | [102.39, 116.27] | [103.66, 117.04] |
| | 20 | 0.98 (0.91, 1.06) | 1.03 (0.97, 1.09) | 1.06 (1.00, 1.12) | 0.98 (0.90, 1.08) | [99.18, 110.25] | [98.70, 111.52] | [100.38, 111.50] | [105.23, 119.92] |
| | 21 | 0.96 (0.91, 1.01) | 0.99 (0.93, 1.05) | 1.03 (0.99, 1.07) | 1.01 (0.95, 1.09) | [101.05, 112.71] | [98.92, 110.87] | [97.85, 109.93] | [105.18, 119.28] |
| | 22 | 0.95 (0.88, 1.02) | 0.95 (0.88, 1.02) | 0.97 (0.92, 1.02) | 1.04 (0.98, 1.11) | [103.66, 117.04] | [98.80, 110.00] | [98.02, 110.87] | [102.39, 116.27] |
| | 23 | 0.98 (0.89, 1.07) | 0.98 (0.91, 1.05) | 0.94 (0.89, 0.99) | 1.04 (0.99, 1.09) | [105.23, 119.92] | [99.18, 110.25] | [98.70, 111.52] | [100.38, 111.50] |
| | 24 | 1.04 (0.98, 1.11) | 1.08 (1.02, 1.13) | 0.96 (0.91, 1.01) | 1.00 (0.96, 1.04) | [105.18, 119.28] | [101.05, 112.71] | [98.92, 110.87] | [97.85, 109.93] |
| | 25 | 1.10 (1.03, 1.18) | 1.13 (1.04, 1.23) | 0.99 (0.92, 1.07) | 0.94 (0.89, 0.99) | [102.39, 116.27] | [103.66, 117.04] | [98.80, 110.00] | [98.02, 110.87] |
| | 26 | 1.04 (0.99, 1.09) | 1.05 (0.97, 1.14) | 1.03 (0.98, 1.09) | 0.92 (0.88, 0.97) | [100.38, 111.50] | [105.23, 119.92] | [99.18, 110.25] | [98.70, 111.52] |
| | 27 | 0.95 (0.90, 0.99) | 0.95 (0.89, 1.01) | 1.05 (0.99, 1.10) | 0.95 (0.90, 1.00) | [97.85, 109.93] | [105.18, 119.28] | [101.05, 112.71] | [98.92, 110.87] |
| | 28 | 0.87 (0.82, 0.93) | 0.91 (0.85, 0.98) | 0.99 (0.91, 1.07) | 0.99 (0.92, 1.05) | [98.02, 110.87] | [102.39, 116.27] | [103.66, 117.04] | [98.80, 110.00] |
| | 29 | 0.90 (0.85, 0.95) | 0.95 (0.91, 1.00) | 0.93 (0.87, 1.00) | 1.02 (0.98, 1.07) | [98.70, 111.52] | [100.38, 111.50] | [105.23, 119.92] | [99.18, 110.25] |
| | 30 | 0.97 (0.90, 1.05) | 0.99 (0.93, 1.04) | 0.93 (0.88, 0.99) | 1.05 (0.99, 1.11) | [98.92, 110.87] | [97.85, 109.93] | [105.18, 119.28] | [101.05, 112.71] |
| | 31 | 1.02 (0.95, 1.10) | 1.03 (0.97, 1.09) | 0.95 (0.89, 1.02) | 1.04 (0.97, 1.12) | [98.80, 110.00] | [98.02, 110.87] | [102.39, 116.27] | [103.66, 117.04] |
| | 32 | 1.03 (0.97, 1.08) | 1.05 (0.99, 1.11) | 1.01 (0.97, 1.05) | 1.00 (0.95, 1.06) | [99.18, 110.25] | [98.70, 111.52] | [100.38, 111.50] | [105.23, 119.92] |
| | 33 | 1.02 (0.94, 1.10) | 1.04 (0.96, 1.12) | 1.08 (1.02, 1.14) | 0.97 (0.92, 1.03) | [101.05, 112.71] | [98.92, 110.87] | [97.85, 109.93] | [105.18, 119.28] |
| | 34 | 1.00 (0.92, 1.09) | 1.00 (0.94, 1.07) | 1.08 (1.03, 1.14) | 0.97 (0.91, 1.03) | [103.66, 117.04] | [98.80, 110.00] | [98.02, 110.87] | [102.39, 116.27] |
| | 35 | 0.97 (0.92, 1.03) | 0.94 (0.90, 0.98) | 1.01 (0.97, 1.05) | 1.01 (0.98, 1.04) | [105.23, 119.92] | [99.18, 110.25] | [98.70, 111.52] | [100.38, 111.50] |
| | 36 | 0.94 (0.83, 1.06) | 0.87 (0.78, 0.97) | 0.92 (0.84, 1.01) | 1.08 (1.00, 1.16) | [105.18, 119.28] | [101.05, 112.71] | [98.92, 110.87] | [97.85, 109.93] |
| ST40t100, K | 1 | 1.00 (0.89, 1.12) | 1.15 (0.88, 1.51) | 0.64 (0.49, 0.84) | 0.92 (0.62, 1.37) | [281.78, 283.84] | [285.15, 290.37] | [296.08, 301.33] | [290.61, 299.70] |
| | 2 | 1.54 (1.17, 2.02) | 1.07 (0.94, 1.23) | 1.07 (0.81, 1.41) | 0.93 (0.81, 1.08) | [282.31, 289.04] | [282.94, 287.01] | [292.16, 298.89] | [296.90, 301.92] |
| | 3 | 1.53 (1.08, 2.15) | 0.95 (0.88, 1.03) | 1.15 (0.89, 1.48) | 0.96 (0.88, 1.04) | [284.94, 295.34] | [281.78, 283.89] | [288.31, 294.44] | [299.65, 301.94] |
| | 4 | 1.04 (0.72, 1.48) | 0.94 (0.86, 1.02) | 1.09 (0.88, 1.35) | 0.83 (0.72, 0.96) | [290.61, 299.70] | [281.78, 283.84] | [285.15, 290.37] | [296.08, 301.33] |

| | | | | | | | | |
|---|---|---|---|---|---|---|---|---|
| 5 | 1.14 (0.94, 1.38) | 1.08 (0.84, 1.38) | 1.04 (0.90, 1.22) | 0.77 (0.60, 1.01) | [296.90, 301.92] | [282.31, 289.04] | [282.94, 287.01] | [292.16, 298.89] |
| 6 | 1.00 (0.92, 1.09) | 0.89 (0.60, 1.33) | 1.00 (0.92, 1.09) | 1.02 (0.81, 1.28) | [299.65, 301.94] | [284.94, 295.34] | [281.78, 283.89] | [288.31, 294.44] |
| 7 | 0.96 (0.78, 1.19) | 0.59 (0.39, 0.87) | 1.00 (0.92, 1.10) | 1.28 (1.04, 1.59) | [296.08, 301.33] | [290.61, 299.70] | [281.78, 283.84] | [285.15, 290.37] |
| 8 | 1.20 (0.91, 1.59) | 1.00 (0.81, 1.23) | 1.21 (0.95, 1.54) | 1.18 (1.02, 1.37) | [292.16, 298.89] | [296.90, 301.92] | [282.31, 289.04] | [282.94, 287.01] |
| 9 | 0.96 (0.76, 1.22) | 1.03 (0.94, 1.12) | 1.35 (0.91, 2.01) | 1.06 (0.97, 1.15) | [288.31, 294.44] | [299.65, 301.94] | [284.94, 295.34] | [281.78, 283.89] |
| 10 | 0.73 (0.57, 0.94) | 1.01 (0.83, 1.22) | 1.21 (0.84, 1.77) | 1.03 (0.94, 1.14) | [285.15, 290.37] | [296.08, 301.33] | [290.61, 299.70] | [281.78, 283.84] |
| 11 | 0.79 (0.67, 0.94) | 0.99 (0.72, 1.35) | 1.30 (1.05, 1.60) | 1.01 (0.81, 1.25) | [282.94, 287.01] | [292.16, 298.89] | [296.90, 301.92] | [282.31, 289.04] |
| 12 | 0.94 (0.85, 1.04) | 0.86 (0.69, 1.06) | 1.03 (0.94, 1.12) | 1.01 (0.67, 1.52) | [281.78, 283.89] | [288.31, 294.44] | [299.65, 301.94] | [284.94, 295.34] |
| 13 | 1.00 (0.90, 1.11) | 0.83 (0.62, 1.10) | 0.78 (0.64, 0.94) | 1.09 (0.75, 1.58) | [281.78, 283.84] | [285.15, 290.37] | [296.08, 301.33] | [290.61, 299.70] |
| 14 | 1.13 (0.93, 1.38) | 0.94 (0.78, 1.13) | 0.69 (0.52, 0.91) | 0.96 (0.78, 1.18) | [282.31, 289.04] | [282.94, 287.01] | [292.16, 298.89] | [296.90, 301.92] |
| 15 | 0.63 (0.45, 0.89) | 1.06 (0.96, 1.17) | 0.77 (0.63, 0.95) | 1.01 (0.93, 1.10) | [284.94, 295.34] | [281.78, 283.89] | [288.31, 294.44] | [299.65, 301.94] |
| 16 | 0.41 (0.28, 0.61) | 1.12 (1.01, 1.23) | 0.88 (0.68, 1.14) | 1.18 (0.98, 1.41) | [290.61, 299.70] | [281.78, 283.84] | [285.15, 290.37] | [296.08, 301.33] |
| 17 | 1.02 (0.82, 1.26) | 1.14 (0.93, 1.39) | 1.04 (0.86, 1.24) | 1.20 (0.92, 1.55) | [296.90, 301.92] | [282.31, 289.04] | [282.94, 287.01] | [292.16, 298.89] |
| 18 | 1.05 (0.96, 1.15) | 0.63 (0.45, 0.90) | 1.04 (0.94, 1.14) | 1.06 (0.86, 1.30) | [299.65, 301.94] | [284.94, 295.34] | [281.78, 283.89] | [288.31, 294.44] |
| 19 | 0.84 (0.70, 1.01) | 0.70 (0.45, 1.07) | 1.00 (0.91, 1.11) | 0.96 (0.74, 1.25) | [296.08, 301.33] | [290.61, 299.70] | [281.78, 283.84] | [285.15, 290.37] |
| 20 | 0.81 (0.63, 1.05) | 1.04 (0.85, 1.28) | 0.90 (0.73, 1.11) | 1.01 (0.86, 1.20) | [292.16, 298.89] | [296.90, 301.92] | [282.31, 289.04] | [282.94, 287.01] |
| 21 | 1.07 (0.85, 1.33) | 1.06 (0.97, 1.16) | 0.74 (0.52, 1.05) | 1.02 (0.93, 1.12) | [288.31, 294.44] | [299.65, 301.94] | [284.94, 295.34] | [281.78, 283.89] |
| 22 | 1.07 (0.79, 1.44) | 1.12 (0.94, 1.35) | 0.62 (0.41, 0.93) | 0.98 (0.90, 1.08) | [285.15, 290.37] | [296.08, 301.33] | [290.61, 299.70] | [281.78, 283.84] |
| 23 | 0.99 (0.82, 1.20) | 1.13 (0.88, 1.45) | 0.78 (0.64, 0.96) | 0.80 (0.65, 0.98) | [282.94, 287.01] | [292.16, 298.89] | [296.90, 301.92] | [282.31, 289.04] |
| 24 | 1.02 (0.91, 1.13) | 0.93 (0.74, 1.16) | 0.98 (0.90, 1.07) | 0.63 (0.46, 0.87) | [281.78, 283.89] | [288.31, 294.44] | [299.65, 301.94] | [284.94, 295.34] |
| 25 | 1.06 (0.97, 1.17) | 0.82 (0.61, 1.09) | 1.16 (0.97, 1.39) | 0.85 (0.59, 1.22) | [281.78, 283.84] | [285.15, 290.37] | [296.08, 301.33] | [290.61, 299.70] |
| 26 | 1.00 (0.79, 1.27) | 0.97 (0.83, 1.14) | 1.24 (0.96, 1.61) | 1.16 (0.97, 1.39) | [282.31, 289.04] | [282.94, 287.01] | [292.16, 298.89] | [296.90, 301.92] |
| 27 | 0.59 (0.38, 0.92) | 1.04 (0.94, 1.15) | 0.92 (0.73, 1.16) | 1.04 (0.96, 1.13) | [284.94, 295.34] | [281.78, 283.89] | [288.31, 294.44] | [299.65, 301.94] |
| 28 | 0.83 (0.58, 1.17) | 1.02 (0.93, 1.11) | 0.91 (0.71, 1.17) | 0.86 (0.71, 1.03) | [290.61, 299.70] | [281.78, 283.84] | [285.15, 290.37] | [296.08, 301.33] |
| 29 | 0.93 (0.77, 1.11) | 1.01 (0.79, 1.30) | 1.10 (0.94, 1.29) | 0.81 (0.64, 1.02) | [296.90, 301.92] | [282.31, 289.04] | [282.94, 287.01] | [292.16, 298.89] |
| 30 | 0.97 (0.89, 1.07) | 1.23 (0.79, 1.93) | 1.06 (0.96, 1.18) | 0.73 (0.58, 0.91) | [299.65, 301.94] | [284.94, 295.34] | [281.78, 283.89] | [288.31, 294.44] |
| 31 | 1.11 (0.91, 1.34) | 1.01 (0.73, 1.39) | 0.96 (0.88, 1.05) | 0.76 (0.62, 0.94) | [296.08, 301.33] | [290.61, 299.70] | [281.78, 283.84] | [285.15, 290.37] |
| 32 | 0.89 (0.70, 1.13) | 0.95 (0.80, 1.13) | 0.71 (0.56, 0.90) | 0.98 (0.86, 1.11) | [292.16, 298.89] | [296.90, 301.92] | [282.31, 289.04] | [282.94, 287.01] |
| 33 | 0.62 (0.47, 0.81) | 1.01 (0.92, 1.11) | 0.84 (0.53, 1.33) | 1.05 (0.97, 1.14) | [288.31, 294.44] | [299.65, 301.94] | [284.94, 295.34] | [281.78, 283.89] |
| 34 | 0.90 (0.73, 1.11) | 1.01 (0.87, 1.17) | 0.95 (0.65, 1.39) | 1.06 (0.98, 1.14) | [285.15, 290.37] | [296.08, 301.33] | [290.61, 299.70] | [281.78, 283.84] |
| 35 | 1.20 (1.03, 1.40) | 0.81 (0.64, 1.03) | 0.97 (0.79, 1.19) | 1.02 (0.83, 1.26) | [282.94, 287.01] | [292.16, 298.89] | [296.90, 301.92] | [282.31, 289.04] |
| 36 | 1.16 (1.03, 1.32) | 0.63 (0.47, 0.83) | 0.92 (0.82, 1.04) | 1.00 (0.62, 1.63) | [281.78, 283.89] | [288.31, 294.44] | [299.65, 301.94] | [284.94, 295.34] |

IRRs and 95% CIs were estimated in the study region using DLNMs for an IQR increase in each exposure across lags 1–36 months prior to disease incidence, stratified by season. Models were adjusted for spatiotemporal trends and surveillance-related changes in reporting and laboratory testing. Layer 5 soil moisture and soil temperature were modeled simultaneously in a single DLNM with season-specific effects.

**Table S6.** Season-specific lagged associations between coccidioidomycosis incidence and above-ground (Layer 2: meteorological layer) precipitation and air temperature (AT).

| Variable | Lag | Winter_IRR_CI | Spring_IRR_CI | Monsoon_IRR_CI | Fall_IRR_CI | Winter_IQR | Spring_IQR | Monsoon_IQR | Fall_IQR |
|---|---|---|---|---|---|---|---|---|---|
| Precipitation, mm | 1 | 0.99 (0.89, 1.10) | 0.94 (0.86, 1.02) | 0.71 (0.60, 0.84) | 0.83 (0.77, 0.90) | [6.01, 38.81] | [0.55, 14.21] | [3.60, 48.84] | [2.57, 24.43] |
| | 2 | 0.85 (0.77, 0.93) | 0.86 (0.80, 0.93) | 0.83 (0.76, 0.92) | 0.72 (0.66, 0.79) | [3.36, 34.73] | [2.73, 24.66] | [0.21, 23.52] | [6.46, 41.02] |
| | 3 | 0.80 (0.74, 0.87) | 0.89 (0.81, 0.98) | 0.98 (0.95, 1.01) | 0.81 (0.76, 0.88) | [2.20, 25.95] | [5.26, 33.71] | [0.09, 6.15] | [18.30, 50.63] |
| | 4 | 0.82 (0.75, 0.89) | 1.06 (0.97, 1.16) | 0.99 (0.92, 1.06) | 0.87 (0.75, 1.00) | [2.57, 24.43] | [6.01, 38.81] | [0.55, 14.21] | [3.60, 48.84] |
| | 5 | 0.92 (0.84, 1.02) | 1.12 (1.02, 1.23) | 0.95 (0.89, 1.02) | 0.98 (0.91, 1.07) | [6.46, 41.02] | [3.36, 34.73] | [2.73, 24.66] | [0.21, 23.52] |
| | 6 | 1.11 (1.01, 1.21) | 1.02 (0.94, 1.11) | 0.91 (0.84, 0.99) | 0.98 (0.95, 1.01) | [18.30, 50.63] | [2.20, 25.95] | [5.26, 33.71] | [0.09, 6.15] |
| | 7 | 1.11 (0.95, 1.29) | 0.94 (0.86, 1.02) | 1.01 (0.92, 1.10) | 0.90 (0.85, 0.95) | [3.60, 48.84] | [2.57, 24.43] | [6.01, 38.81] | [0.55, 14.21] |
| | 8 | 0.96 (0.87, 1.06) | 0.86 (0.77, 0.95) | 1.09 (1.00, 1.19) | 0.91 (0.85, 0.98) | [0.21, 23.52] | [6.46, 41.02] | [3.36, 34.73] | [2.73, 24.66] |
| | 9 | 0.97 (0.93, 1.01) | 0.89 (0.81, 0.98) | 1.04 (0.95, 1.12) | 1.03 (0.96, 1.11) | [0.09, 6.15] | [18.30, 50.63] | [2.20, 25.95] | [5.26, 33.71] |
| | 10 | 0.93 (0.88, 0.99) | 0.86 (0.74, 1.00) | 0.92 (0.85, 1.00) | 1.02 (0.94, 1.10) | [0.55, 14.21] | [3.60, 48.84] | [2.57, 24.43] | [6.01, 38.81] |
| | 11 | 0.91 (0.84, 0.99) | 1.04 (0.94, 1.15) | 0.84 (0.76, 0.94) | 1.00 (0.91, 1.09) | [2.73, 24.66] | [0.21, 23.52] | [6.46, 41.02] | [3.36, 34.73] |
| | 12 | 0.94 (0.87, 1.01) | 1.04 (1.00, 1.08) | 0.88 (0.80, 0.97) | 1.04 (0.96, 1.13) | [5.26, 33.71] | [0.09, 6.15] | [18.30, 50.63] | [2.20, 25.95] |
| | 13 | 1.04 (0.97, 1.13) | 1.01 (0.95, 1.08) | 0.94 (0.82, 1.07) | 1.01 (0.94, 1.09) | [6.01, 38.81] | [0.55, 14.21] | [3.60, 48.84] | [2.57, 24.43] |
| | 14 | 1.16 (1.05, 1.27) | 0.92 (0.86, 1.00) | 0.96 (0.88, 1.06) | 0.99 (0.89, 1.10) | [3.36, 34.73] | [2.73, 24.66] | [0.21, 23.52] | [6.46, 41.02] |
| | 15 | 1.14 (1.05, 1.24) | 1.00 (0.93, 1.08) | 0.97 (0.94, 1.01) | 1.10 (1.00, 1.21) | [2.20, 25.95] | [5.26, 33.71] | [0.09, 6.15] | [18.30, 50.63] |
| | 16 | 1.03 (0.96, 1.11) | 1.05 (0.97, 1.13) | 0.92 (0.87, 0.99) | 1.09 (0.96, 1.24) | [2.57, 24.43] | [6.01, 38.81] | [0.55, 14.21] | [3.60, 48.84] |
| | 17 | 0.92 (0.82, 1.03) | 1.04 (0.94, 1.15) | 0.94 (0.88, 1.01) | 0.95 (0.86, 1.05) | [6.46, 41.02] | [3.36, 34.73] | [2.73, 24.66] | [0.21, 23.52] |
| | 18 | 0.95 (0.87, 1.03) | 1.09 (1.00, 1.18) | 1.06 (0.99, 1.13) | 0.95 (0.92, 0.98) | [18.30, 50.63] | [2.20, 25.95] | [5.26, 33.71] | [0.09, 6.15] |
| | 19 | 0.94 (0.82, 1.08) | 0.98 (0.91, 1.06) | 1.06 (0.99, 1.15) | 0.89 (0.83, 0.95) | [3.60, 48.84] | [2.57, 24.43] | [6.01, 38.81] | [0.55, 14.21] |
| | 20 | 0.94 (0.84, 1.05) | 0.79 (0.70, 0.89) | 0.98 (0.89, 1.07) | 0.95 (0.89, 1.02) | [0.21, 23.52] | [6.46, 41.02] | [3.36, 34.73] | [2.73, 24.66] |
| | 21 | 0.98 (0.95, 1.01) | 1.00 (0.92, 1.07) | 1.07 (0.99, 1.16) | 1.00 (0.94, 1.06) | [0.09, 6.15] | [18.30, 50.63] | [2.20, 25.95] | [5.26, 33.71] |
| | 22 | 0.95 (0.89, 1.01) | 1.20 (1.06, 1.35) | 1.11 (1.03, 1.20) | 1.00 (0.93, 1.08) | [0.55, 14.21] | [3.60, 48.84] | [2.57, 24.43] | [6.01, 38.81] |
| | 23 | 0.93 (0.87, 1.00) | 1.02 (0.91, 1.14) | 1.01 (0.90, 1.12) | 1.00 (0.92, 1.10) | [2.73, 24.66] | [0.21, 23.52] | [6.46, 41.02] | [3.36, 34.73] |
| | 24 | 0.92 (0.86, 0.99) | 0.99 (0.96, 1.02) | 1.01 (0.93, 1.10) | 0.94 (0.87, 1.01) | [5.26, 33.71] | [0.09, 6.15] | [18.30, 50.63] | [2.20, 25.95] |
| | 25 | 0.95 (0.88, 1.02) | 0.96 (0.90, 1.03) | 1.06 (0.92, 1.22) | 0.90 (0.84, 0.96) | [6.01, 38.81] | [0.55, 14.21] | [3.60, 48.84] | [2.57, 24.43] |
| | 26 | 1.06 (0.97, 1.16) | 0.97 (0.90, 1.04) | 1.06 (0.95, 1.17) | 0.93 (0.84, 1.03) | [3.36, 34.73] | [2.73, 24.66] | [0.21, 23.52] | [6.46, 41.02] |
| | 27 | 1.10 (1.02, 1.19) | 1.04 (0.96, 1.12) | 0.98 (0.95, 1.02) | 1.02 (0.94, 1.10) | [2.20, 25.95] | [5.26, 33.71] | [0.09, 6.15] | [18.30, 50.63] |
| | 28 | 1.04 (0.97, 1.13) | 1.12 (1.04, 1.21) | 0.91 (0.85, 0.97) | 1.01 (0.87, 1.18) | [2.57, 24.43] | [6.01, 38.81] | [0.55, 14.21] | [3.60, 48.84] |
| | 29 | 0.93 (0.84, 1.02) | 1.14 (1.04, 1.24) | 1.00 (0.94, 1.06) | 0.95 (0.87, 1.03) | [6.46, 41.02] | [3.36, 34.73] | [2.73, 24.66] | [0.21, 23.52] |
| | 30 | 0.96 (0.88, 1.05) | 1.07 (0.98, 1.17) | 1.08 (1.01, 1.15) | 0.98 (0.95, 1.01) | [18.30, 50.63] | [2.20, 25.95] | [5.26, 33.71] | [0.09, 6.15] |
| | 31 | 1.18 (1.02, 1.38) | 1.01 (0.93, 1.10) | 0.98 (0.91, 1.06) | 1.01 (0.94, 1.07) | [3.60, 48.84] | [2.57, 24.43] | [6.01, 38.81] | [0.55, 14.21] |
| | 32 | 1.13 (1.03, 1.25) | 0.94 (0.85, 1.03) | 1.01 (0.93, 1.11) | 0.98 (0.92, 1.04) | [0.21, 23.52] | [6.46, 41.02] | [3.36, 34.73] | [2.73, 24.66] |
| | 33 | 1.02 (0.99, 1.06) | 0.94 (0.86, 1.02) | 1.10 (1.00, 1.20) | 0.97 (0.91, 1.04) | [0.09, 6.15] | [18.30, 50.63] | [2.20, 25.95] | [5.26, 33.71] |
| | 34 | 1.04 (0.98, 1.11) | 0.97 (0.84, 1.13) | 1.03 (0.96, 1.11) | 1.10 (1.03, 1.17) | [0.55, 14.21] | [3.60, 48.84] | [2.57, 24.43] | [6.01, 38.81] |
| | 35 | 1.01 (0.94, 1.09) | 1.00 (0.91, 1.10) | 0.96 (0.87, 1.05) | 1.11 (1.02, 1.20) | [2.73, 24.66] | [0.21, 23.52] | [6.46, 41.02] | [3.36, 34.73] |
| | 36 | 0.92 (0.84, 1.00) | 0.98 (0.95, 1.02) | 0.95 (0.86, 1.05) | 0.93 (0.85, 1.03) | [5.26, 33.71] | [0.09, 6.15] | [18.30, 50.63] | [2.20, 25.95] |
| AT, °C | 1 | 1.02 (0.92, 1.12) | 0.92 (0.69, 1.22) | 1.13 (1.00, 1.27) | 0.75 (0.54, 1.03) | [10.29, 12.48] | [16.51, 22.86] | [29.55, 31.91] | [17.18, 27.57] |
| | 2 | 0.90 (0.79, 1.03) | 1.00 (0.84, 1.17) | 0.78 (0.63, 0.99) | 1.09 (0.84, 1.41) | [10.49, 14.50] | [13.30, 18.72] | [25.29, 31.14] | [23.54, 29.98] |
| | 3 | 0.59 (0.42, 0.83) | 0.98 (0.89, 1.09) | 0.56 (0.42, 0.76) | 0.95 (0.85, 1.06) | [11.60, 20.95] | [11.12, 14.73] | [20.34, 28.61] | [28.92, 31.80] |
| | 4 | 0.81 (0.54, 1.21) | 1.02 (0.96, 1.10) | 0.64 (0.50, 0.82) | 0.99 (0.90, 1.08) | [17.18, 27.57] | [10.29, 12.48] | [16.51, 22.86] | [29.55, 31.91] |

| | | | | | | | | |
|---|---|---|---|---|---|---|---|---|
| 5 | 1.37 (1.05, 1.80) | 1.11 (0.96, 1.29) | 0.73 (0.63, 0.85) | 1.09 (0.87, 1.37) | [23.54, 29.98] | [10.49, 14.50] | [13.30, 18.72] | [25.29, 31.14] |
| 6 | 1.22 (1.03, 1.43) | 0.78 (0.56, 1.08) | 0.88 (0.80, 0.97) | 0.71 (0.54, 0.92) | [28.92, 31.80] | [11.60, 20.95] | [11.12, 14.73] | [20.34, 28.61] |
| 7 | 1.10 (0.97, 1.25) | 0.88 (0.60, 1.28) | 1.00 (0.93, 1.06) | 0.67 (0.55, 0.81) | [29.55, 31.91] | [17.18, 27.57] | [10.29, 12.48] | [16.51, 22.86] |
| 8 | 0.92 (0.74, 1.16) | 1.36 (1.03, 1.80) | 1.02 (0.90, 1.16) | 0.92 (0.80, 1.06) | [25.29, 31.14] | [23.54, 29.98] | [10.49, 14.50] | [13.30, 18.72] |
| 9 | 0.61 (0.46, 0.81) | 1.07 (0.92, 1.24) | 1.02 (0.76, 1.35) | 1.03 (0.95, 1.11) | [20.34, 28.61] | [28.92, 31.80] | [11.60, 20.95] | [11.12, 14.73] |
| 10 | 0.79 (0.62, 1.00) | 0.95 (0.84, 1.07) | 1.48 (0.99, 2.22) | 0.99 (0.93, 1.05) | [16.51, 22.86] | [29.55, 31.91] | [17.18, 27.57] | [10.29, 12.48] |
| 11 | 0.97 (0.82, 1.15) | 0.68 (0.56, 0.83) | 0.87 (0.66, 1.15) | 0.82 (0.73, 0.93) | [13.30, 18.72] | [25.29, 31.14] | [23.54, 29.98] | [10.49, 14.50] |
| 12 | 0.92 (0.84, 1.00) | 0.55 (0.42, 0.73) | 0.85 (0.73, 0.98) | 0.61 (0.46, 0.82) | [11.12, 14.73] | [20.34, 28.61] | [28.92, 31.80] | [11.60, 20.95] |
| 13 | 0.89 (0.84, 0.95) | 0.75 (0.59, 0.94) | 0.93 (0.81, 1.05) | 1.17 (0.84, 1.62) | [10.29, 12.48] | [16.51, 22.86] | [29.55, 31.91] | [17.18, 27.57] |
| 14 | 0.83 (0.72, 0.94) | 0.85 (0.72, 0.99) | 0.92 (0.74, 1.15) | 1.34 (1.06, 1.70) | [10.49, 14.50] | [13.30, 18.72] | [25.29, 31.14] | [23.54, 29.98] |
| 15 | 0.79 (0.60, 1.04) | 0.91 (0.83, 1.01) | 0.77 (0.60, 0.99) | 1.13 (0.97, 1.33) | [11.60, 20.95] | [11.12, 14.73] | [20.34, 28.61] | [28.92, 31.80] |
| 16 | 0.55 (0.42, 0.73) | 0.95 (0.90, 1.01) | 0.88 (0.70, 1.10) | 1.00 (0.88, 1.13) | [17.18, 27.57] | [10.29, 12.48] | [16.51, 22.86] | [29.55, 31.91] |
| 17 | 0.72 (0.56, 0.91) | 0.93 (0.82, 1.06) | 1.15 (0.96, 1.39) | 0.89 (0.72, 1.11) | [23.54, 29.98] | [10.49, 14.50] | [13.30, 18.72] | [25.29, 31.14] |
| 18 | 1.02 (0.87, 1.19) | 0.81 (0.62, 1.08) | 1.21 (1.11, 1.32) | 1.28 (1.02, 1.61) | [28.92, 31.80] | [11.60, 20.95] | [11.12, 14.73] | [20.34, 28.61] |
| 19 | 1.05 (0.91, 1.21) | 0.55 (0.40, 0.75) | 1.08 (1.03, 1.14) | 1.30 (1.03, 1.66) | [29.55, 31.91] | [17.18, 27.57] | [10.29, 12.48] | [16.51, 22.86] |
| 20 | 1.19 (0.95, 1.48) | 0.64 (0.51, 0.81) | 1.00 (0.89, 1.12) | 1.06 (0.89, 1.27) | [25.29, 31.14] | [23.54, 29.98] | [10.49, 14.50] | [13.30, 18.72] |
| 21 | 1.01 (0.79, 1.29) | 1.07 (0.93, 1.24) | 0.62 (0.46, 0.83) | 1.03 (0.95, 1.12) | [20.34, 28.61] | [28.92, 31.80] | [11.60, 20.95] | [11.12, 14.73] |
| 22 | 0.68 (0.52, 0.89) | 1.23 (1.07, 1.41) | 0.47 (0.32, 0.68) | 1.01 (0.95, 1.07) | [16.51, 22.86] | [29.55, 31.91] | [17.18, 27.57] | [10.29, 12.48] |
| 23 | 0.77 (0.62, 0.96) | 1.29 (1.03, 1.62) | 1.26 (0.96, 1.64) | 0.88 (0.78, 0.98) | [13.30, 18.72] | [25.29, 31.14] | [23.54, 29.98] | [10.49, 14.50] |
| 24 | 1.01 (0.90, 1.12) | 1.12 (0.86, 1.45) | 1.39 (1.19, 1.63) | 0.60 (0.46, 0.79) | [11.12, 14.73] | [20.34, 28.61] | [28.92, 31.80] | [11.60, 20.95] |
| 25 | 1.09 (1.03, 1.16) | 1.14 (0.89, 1.47) | 1.13 (0.99, 1.28) | 0.62 (0.46, 0.84) | [10.29, 12.48] | [16.51, 22.86] | [29.55, 31.91] | [17.18, 27.57] |
| 26 | 1.19 (1.03, 1.37) | 1.08 (0.90, 1.28) | 0.98 (0.78, 1.22) | 0.91 (0.72, 1.15) | [10.49, 14.50] | [13.30, 18.72] | [25.29, 31.14] | [23.54, 29.98] |
| 27 | 1.05 (0.77, 1.43) | 1.02 (0.93, 1.11) | 1.13 (0.80, 1.58) | 1.12 (0.98, 1.29) | [11.60, 20.95] | [11.12, 14.73] | [20.34, 28.61] | [28.92, 31.80] |
| 28 | 1.10 (0.76, 1.61) | 0.98 (0.93, 1.04) | 1.02 (0.80, 1.30) | 1.05 (0.95, 1.17) | [17.18, 27.57] | [10.29, 12.48] | [16.51, 22.86] | [29.55, 31.91] |
| 29 | 0.92 (0.71, 1.19) | 0.96 (0.84, 1.09) | 0.89 (0.74, 1.07) | 0.79 (0.63, 1.00) | [23.54, 29.98] | [10.49, 14.50] | [13.30, 18.72] | [25.29, 31.14] |
| 30 | 1.01 (0.88, 1.16) | 1.07 (0.75, 1.53) | 0.86 (0.79, 0.94) | 0.62 (0.47, 0.83) | [28.92, 31.80] | [11.60, 20.95] | [11.12, 14.73] | [20.34, 28.61] |
| 31 | 1.09 (0.97, 1.23) | 0.86 (0.59, 1.24) | 0.87 (0.82, 0.93) | 0.86 (0.65, 1.12) | [29.55, 31.91] | [17.18, 27.57] | [10.29, 12.48] | [16.51, 22.86] |
| 32 | 0.77 (0.62, 0.95) | 0.88 (0.68, 1.14) | 0.79 (0.70, 0.89) | 0.96 (0.81, 1.14) | [25.29, 31.14] | [23.54, 29.98] | [10.49, 14.50] | [13.30, 18.72] |
| 33 | 0.68 (0.52, 0.89) | 1.03 (0.90, 1.19) | 0.78 (0.54, 1.13) | 0.99 (0.92, 1.07) | [20.34, 28.61] | [28.92, 31.80] | [11.60, 20.95] | [11.12, 14.73] |
| 34 | 1.22 (0.97, 1.53) | 0.96 (0.86, 1.07) | 0.59 (0.41, 0.85) | 1.02 (0.96, 1.09) | [16.51, 22.86] | [29.55, 31.91] | [17.18, 27.57] | [10.29, 12.48] |
| 35 | 0.95 (0.79, 1.14) | 0.71 (0.59, 0.85) | 0.68 (0.54, 0.86) | 1.04 (0.93, 1.15) | [13.30, 18.72] | [25.29, 31.14] | [23.54, 29.98] | [10.49, 14.50] |
| 36 | 0.92 (0.81, 1.05) | 0.67 (0.50, 0.88) | 1.02 (0.88, 1.17) | 1.51 (1.08, 2.11) | [11.12, 14.73] | [20.34, 28.61] | [28.92, 31.80] | [11.60, 20.95] |

IRRs and 95% CIs were estimated in the study region using DLNMs for an IQR increase in each exposure across lags 1–36 months prior to disease incidence, stratified by season. Models were adjusted for spatiotemporal trends and surveillance-related changes in reporting and laboratory testing. Layer 2 meteorological conditions (precipitation and AT) were modeled simultaneously in a single DLNM with season-specific effects.

**Table S7.** Season-specific lagged associations between coccidioidomycosis incidence and above-ground (Layer 1: dust dispersion layer) $PM_{10}$ and wind speed.

| Variable | Lag | Winter_IRR_CI | Spring_IRR_CI | Monsoon_IRR_CI | Fall_IRR_CI | Winter_IQR | Spring_IQR | Monsoon_IQR | Fall_IQR |
|---|---|---|---|---|---|---|---|---|---|
| PM10, µg m-3 | 1 | 1.08 (1.00, 1.16) | 1.01 (0.94, 1.08) | 1.15 (1.05, 1.26) | 1.37 (1.26, 1.48) | [21.20, 36.63] | [26.49, 42.97] | [26.46, 47.04] | [27.22, 47.35] |
| | 2 | 1.07 (1.03, 1.11) | 1.04 (1.00, 1.07) | 1.04 (1.00, 1.08) | 1.24 (1.19, 1.30) | [23.66, 42.36] | [23.61, 38.45] | [28.97, 48.71] | [24.43, 45.10] |
| | 3 | 1.05 (0.99, 1.11) | 1.04 (1.00, 1.09) | 0.97 (0.92, 1.02) | 1.10 (1.03, 1.18) | [26.70, 46.84] | [21.04, 35.97] | [29.57, 47.39] | [24.03, 44.98] |
| | 4 | 1.04 (0.98, 1.09) | 1.04 (0.99, 1.09) | 0.94 (0.90, 0.99) | 0.98 (0.92, 1.03) | [27.22, 47.35] | [21.20, 36.63] | [26.49, 42.97] | [26.46, 47.04] |
| | 5 | 1.05 (1.01, 1.09) | 1.02 (0.98, 1.06) | 0.94 (0.91, 0.97) | 0.88 (0.85, 0.92) | [24.43, 45.10] | [23.66, 42.36] | [23.61, 38.45] | [28.97, 48.71] |
| | 6 | 1.06 (0.97, 1.16) | 0.99 (0.92, 1.07) | 0.93 (0.86, 1.01) | 0.82 (0.75, 0.90) | [24.03, 44.98] | [26.70, 46.84] | [21.04, 35.97] | [29.57, 47.39] |
| Wind speed, m s-1 | 1 | 0.98 (0.93, 1.04) | 1.06 (0.98, 1.16) | 1.10 (1.04, 1.17) | 0.93 (0.85, 1.03) | [0.44, 1.01] | [1.71, 2.69] | [2.46, 2.99] | [0.79, 2.00] |
| | 2 | 1.00 (0.97, 1.04) | 1.09 (1.03, 1.16) | 1.04 (1.01, 1.07) | 0.96 (0.91, 1.02) | [0.37, 0.81] | [1.15, 2.23] | [2.59, 3.02] | [1.50, 2.53] |
| | 3 | 1.03 (0.98, 1.09) | 0.97 (0.93, 1.02) | 1.04 (1.00, 1.09) | 1.00 (0.96, 1.05) | [0.47, 1.26] | [0.61, 1.51] | [2.34, 2.97] | [2.20, 2.81] |
| | 4 | 0.97 (0.90, 1.04) | 0.92 (0.88, 0.96) | 1.10 (1.04, 1.17) | 1.01 (0.97, 1.06) | [0.79, 2.00] | [0.44, 1.01] | [1.71, 2.69] | [2.46, 2.99] |
| | 5 | 1.00 (0.94, 1.06) | 0.96 (0.93, 0.99) | 1.06 (1.00, 1.13) | 1.02 (0.99, 1.05) | [1.50, 2.53] | [0.37, 0.81] | [1.15, 2.23] | [2.59, 3.02] |
| | 6 | 1.01 (0.95, 1.08) | 1.00 (0.93, 1.06) | 0.94 (0.88, 1.01) | 1.15 (1.07, 1.23) | [2.20, 2.81] | [0.47, 1.26] | [0.61, 1.51] | [2.34, 2.97] |

IRRs and 95% CIs were estimated in the study region using DLNMs for an IQR increase in each exposure across lags 1–36 months prior to disease incidence, stratified by season. Models were adjusted for spatiotemporal trends and surveillance-related changes in reporting and laboratory testing. Layer 1 dust dispersion conditions ($PM_{10}$ and wind speed) were modeled simultaneously in a single DLNM with season-specific effects.

**Table S8.** Selected models and performance metrics for the two-stage stacked ensemble prediction models using smoothed lag (Slag) environmental predictors.

| Stage | Model component (predictor layer) | Winner model | WtdMed validation RMSE | WtdMed GapRatio | IQR validation RMSE | WorstFold validation RMSE |
|---|---|---|---|---|---|---|
| Stage 1 (base learner: Slags) | Topsoil (0–10 cm): SM & ST | GLM.nb | 2.99 | 0.13 | 1.99 | 6.86 |
| | Midsoil (10–40 cm): SM & ST | GLM.nb | 3.27 | 0.24 | 2.78 | 6.45 |
| | Deepsoil (40–100 cm):SM & ST | RF | 3.09 | 0.58 | 2.35 | 6.75 |
| | Meteorological layer: precipitation & AT | RF | 3.23 | 0.40 | 2.65 | 6.59 |
| | Dust dispersion layer: PM10 & wind speed | XGB | 2.62 | 0.51 | 2.81 | 6.61 |
| Stage 2 (meta-learner) | Multilayer ensemble (stacked across all stage-1 base learners) | RF | 3.85 | 0.20 | 1.68 | 5.79 |

Abbreviations: SM=soil moisture; ST=soil temperature; AT=air temperature; GLM.nb=negative binomial generalized linear model; RF=random forest; XGB=extreme gradient boosting (XGBoost); RMSE=root mean square error; WtdMed=recency-weighted median.

For each environmental predictor layer at stage 1, three candidate model types (GLM.nb, RF, and XGB) were evaluated by progressive (walk-forward) time-series cross-validation (CV) and the best-performing configuration was selected. Stage 1 used 7-fold expanding-window, one-year-ahead CV with validation years 2014–2020 (recency weights 1, 1, 1, 2, 2, 2, 2). Stage 2 used 3-fold expanding-window, one-year-ahead CV with validation years 2018–2020 (recency weights 1, 1, 2). Candidate meta-learner families for stage 2 were constrained least squares (CLS), RF, and XGB.

WtdMed validation RMSE is the recency-weighted median of the validation RMSE across CV folds (incidence per 100,000 population); for RF and XGB, fold-level RMSE is the median across five random seeds. WtdMed GapRatio is the recency-weighted median of the generalisation gap ratio, defined as max(0, (validation RMSE − training RMSE) / validation RMSE); values closer 0 indicate better generalisation, whereas values approaching 1 indicate overfitting. WorstFold validation RMSE is the maximum validation RMSE across all folds, representing the worst single-year prediction error. IQR validation RMSE is the unweighted interquartile range of fold-level validation RMSE, measuring prediction stability over time.

Among configurations within 5% of the minimum validation RMSE (near-best set), the final model was selected independently for each Stage 1 layer and for the Stage 2 meta-learner by sequential tiebreaking: (1) lowest gap ratio, (2) lowest IQR, (3) lowest worst-fold RMSE.

**Table S9.** Sensitivity analysis with raw environmental lag predictors: selected models and performance metrics for the two-stage stacked ensemble prediction models.

| Stage | Model component (predictor layer) | Winner model | WtdMed validation RMSE | WtdMed GapRatio | IQR validation RMSE | WorstFold validation RMSE |
|---|---|---|---|---|---|---|
| Stage 1 (base learner: raw lags) | Topsoil (0–10 cm): SM & ST | GLM.nb | 3.21 | 0.22 | 2.09 | 6.86 |
| | Midsoil (10–40 cm): SM & ST | RF | 3.14 | 0.49 | 2.70 | 6.99 |
| | Deepsoil (40–100 cm):SM & ST | RF | 3.25 | 0.43 | 2.58 | 7.01 |
| | Meteorological layer: precipitation & AT | RF | 3.21 | 0.45 | 2.68 | 6.89 |
| | Dust dispersion layer: PM10 & wind speed | DLNM | 2.77 | 0.00 | 3.04 | 5.95 |
| Stage 2 (meta-learner) | Multilayer ensemble (stacked across all stage-1 base learners) | RF | 3.92 | 0.28 | 1.39 | 5.60 |

Abbreviations: SM=soil moisture; ST=soil temperature; AT=air temperature; GLM.nb=negative binomial generalized linear model; DLNM=distributed lag non-linear model; RF=random forest; XGB=extreme gradient boosting (XGBoost); RMSE=root mean square error; WtdMed=recency-weighted median.

The model structure is identical to Table S8 but uses raw individual monthly lag values in place of smoothed environmental lag features. Because DLNM internally estimates the lag-response function through cross-basis terms, it requires raw lag inputs and was therefore included as a Stage 1 candidate only in this sensitivity analysis (four candidates: GLM.nb, DLNM, RF, and XGB) but not in the main analysis using pre-smoothed features (three candidates: GLM.nb, RF, and XGB). Stage 1 used seven-fold expanding-window, one-year-ahead CV with validation years 2014–2020 (recency weights 1, 1, 1, 2, 2, 2, 2). Stage 2 used three-fold expanding-window, one-year-ahead CV with validation years 2018–2020 (recency weights 1, 1, 2). Candidate meta-learner families for Stage 2 were CLS, RF, and XGB.

WtdMed validation RMSE is the recency-weighted median of the validation RMSE across CV folds (incidence per 100,000 population); for RF and XGB, fold-level RMSE is the median across five random seeds. WtdMed GapRatio is the recency-weighted median of the generalisation gap ratio, defined as max(0, (validation RMSE − training RMSE) / validation RMSE); values closer 0 indicate better generalisation, whereas values approaching 1 indicate overfitting. WorstFold validation RMSE is the maximum validation RMSE across all folds, representing the worst single-year prediction error. IQR validation RMSE is the unweighted interquartile range of fold-level validation RMSE, measuring prediction stability over time.

Among configurations within 5% of the minimum validation RMSE (near-best set), the final model was selected independently for each Stage 1 layer and for the Stage 2 meta-learner by sequential tiebreaking: (1) lowest gap ratio, (2) lowest IQR, (3) lowest worst-fold RMSE.

**Table S10.** Test-period multilayer ensemble performance across four pipeline configurations (2021–2024).

| Pipeline configuration | Stage 2 winner | Spatial range | Test RMSE | Test GapRatio |
|---|---|---|---|---|
| Smoothed lags, environmental only* | RF | Study region | 4.45 | 0.00 |
| | | Maricopa | 4.50 | 0.00 |
| | | Pima | 4.67 | 0.00 |
| | | Pinal | 5.97 | 0.11 |
| Smoothed lags, env + incidence (1, 12, 24, 36) | RF | Study region | 3.02 | 0.10 |
| | | Maricopa | 3.23 | 0.08 |
| | | Pima | 2.82 | 0.00 |
| | | Pinal | 4.36 | 0.20 |
| ASmoothed lags, env + incidence (12, 24, 36) | RF | Study region | 4.43 | 0.08 |
| | | Maricopa | 4.49 | 0.05 |
| | | Pima | 4.57 | 0.00 |
| | | Pinal | 6.00 | 0.19 |
| Raw lags, environmental only | RF | Study region | 4.67 | 0.00 |
| | | Maricopa | 4.34 | 0.00 |
| | | Pima | 7.28 | 0.11 |
| | | Pinal | 6.52 | 0.00 |

Abbreviations: RF = random forest; Test RMSE = root mean square error on the held-out test period (2021–2024); study region values are population-weighted across three counties (n = 48 county-months per county; n = 144 total).

Test GapRatio = max(0, (Test RMSE − Train RMSE) / Test RMSE); 0 indicates no generalisation gap (test performance equal to or better than training), with higher values indicating poorer generalisation. Values displayed as 0.00 represent either an exact zero (Test RMSE ≤ Train RMSE) or a small positive value <0.005 rounded to two decimal places.

All four pipeline configurations selected RF as the Stage 2 meta-learner via cross-validation (tables S8–S9; figure S57).

For the two incidence lag configurations, stage 1 base learners are identical to the smoothed-lag environmental-only pipeline; only the stage 2 meta-learner differs.

* Winning pipeline configuration, selected based on primary assessment (visual inspection of test-period forecast plots).

**Table S11.** Test-period individual base learner performance: smoothed lags versus raw lags (2021–2024)

| Pipeline configuration | Stage 1 layer | Stage 1 winner | Test RMSE | Test GapRatio |
|---|---|---|---|---|
| Smoothed lags, environmental only* | Topsoil (0–10 cm): SM & ST | GLM.nb | 3.53 | 0.31 |
| | Midsoil (10–40 cm): SM & ST | GLM.nb | 3.25 | 0.27 |
| | Deepsoil (40–100 cm):SM & ST | RF | 3.66 | 0.67 |
| | Meteorological layer: precipitation & AT | RF | 3.63 | 0.47 |
| | Dust dispersion layer: PM10 & wind speed | XGB | 3.30 | 0.65 |
| Raw lags, environmental only | Topsoil (0–10 cm): SM & ST | GLM.nb | 3.59 | 0.33 |
| | Midsoil (10–40 cm): SM & ST | RF | 3.66 | 0.58 |
| | Deepsoil (40–100 cm):SM & ST | RF | 4.35 | 0.58 |
| | Meteorological layer: precipitation & AT | RF | 3.63 | 0.49 |
| | Dust dispersion layer: PM10 & wind speed | DLNM | 4.05 | 0.34 |

Abbreviations: GLM.nb = negative binomial generalised linear model; RF = random forest; XGB = extreme gradient boosting (XGBoost); DLNM = distributed lag non-linear model; RMSE = root mean square error.

Test RMSE and Test GapRatio are defined as in table S10. Values are for the study region (population-weighted, n = 144 county-months). Only the smoothed-lag and raw-lag pipelines are compared because the two incidence lag sensitivity analyses share identical stage 1 base learners with the smoothed-lag pipeline. Winner model was selected independently per layer via cross-validation (table S8 for smoothed lags; table S9 for raw lags). * Winning pipeline configuration.

#### Supplementary Figures

**Figure S1.** Spatial trends in coccidioidomycosis incidence and environmental exposures.

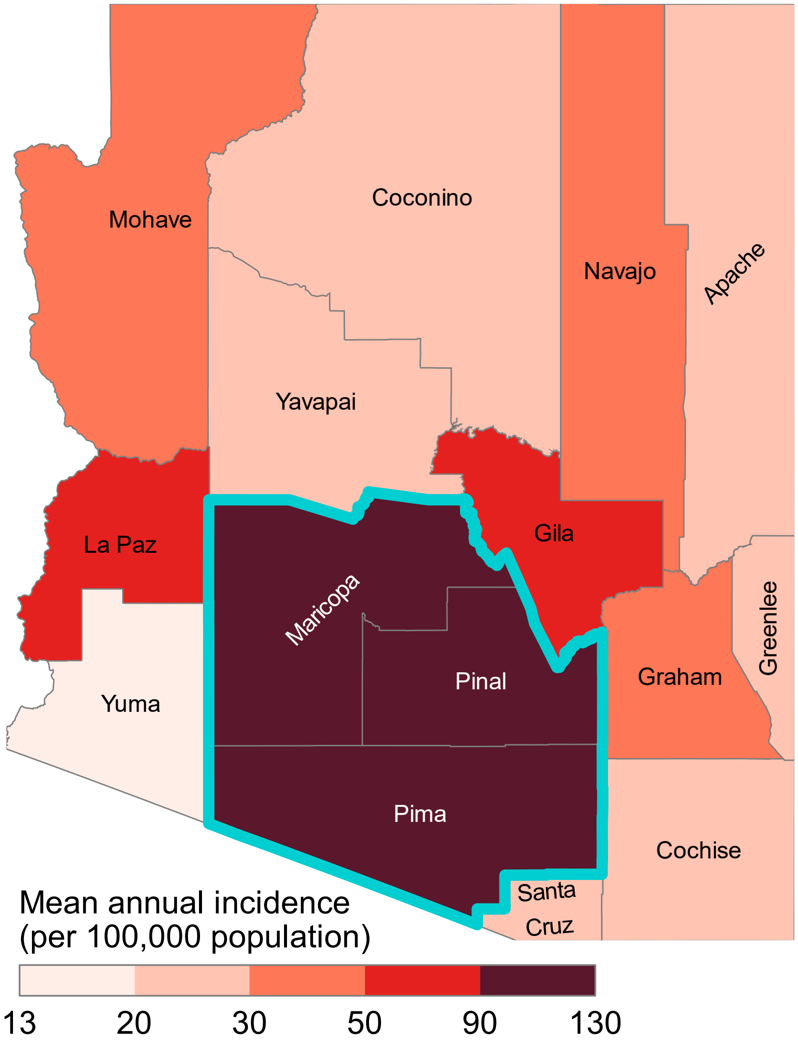

**Note:** Counties are shaded by mean annual coccidioidomycosis incidence (per 100,000 population) across Arizona for 1997–2024. The study region—Maricopa, Pima, and Pinal counties—is outlined in cyan and was used for all analyses.

**Figure S2.** Total monthly coccidioidomycosis case counts across surveillance regimes in the study region.

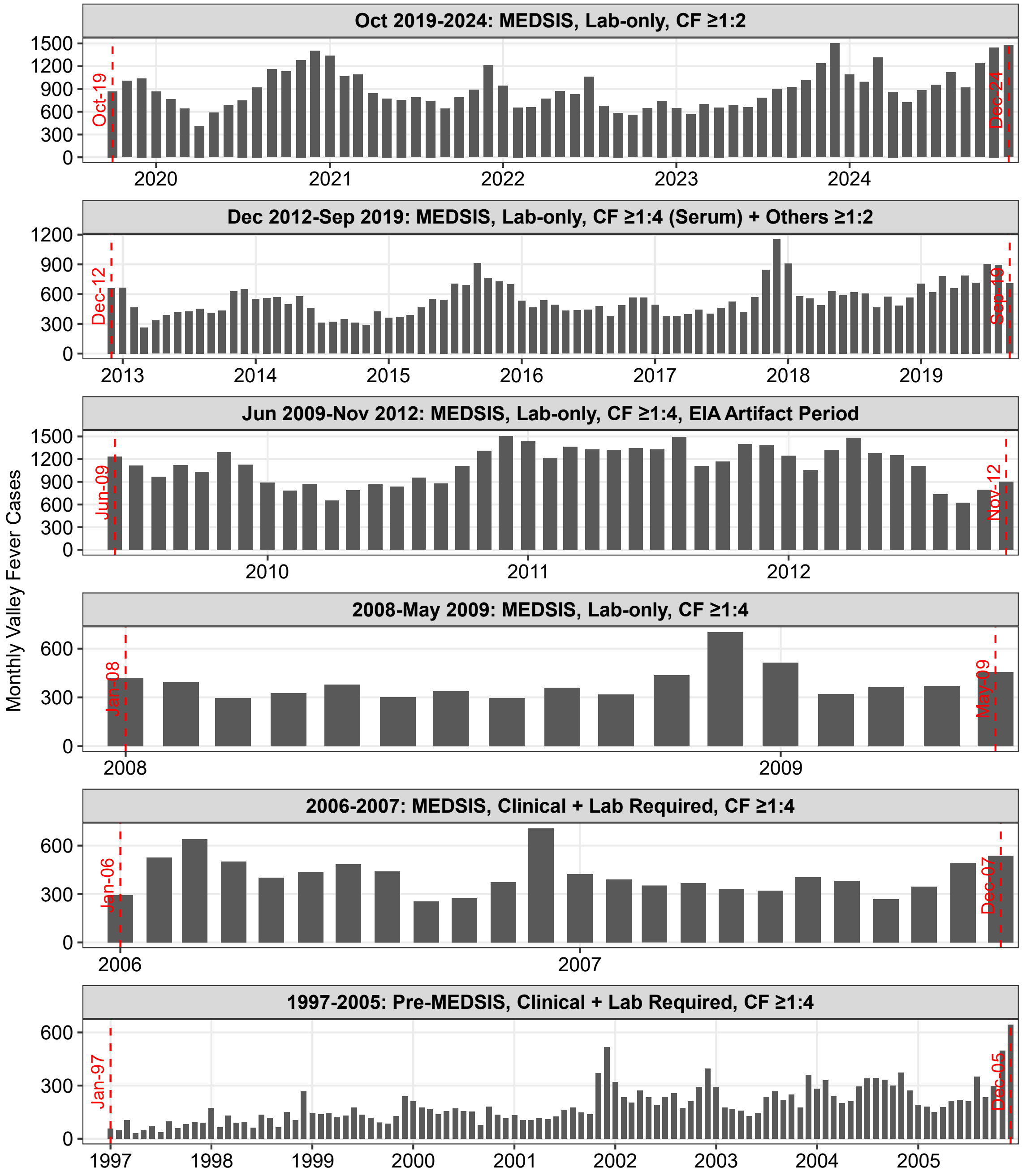

**Note**: The study region is the hyperendemic tri-county area comprising Maricopa, Pima, and Pinal counties, Arizona. Red dashed lines indicate transitions between surveillance regimes. We defined a surveillance regime indicator to capture major surveillance changes in reporting systems and laboratory testing practices that influenced coccidioidomycosis trends. These changes were documented in the ADHS coccidioidomycosis surveillance data caveats.[12] The Pre-MEDSIS regime (prior to 2006) reflects the period before implementation of the Medical Electronic Disease Surveillance Intelligence System (MEDSIS) in 2006, when case reports were primarily received by fax or mail and manually reviewed against the state's disease registry. In 2008, the case definition changed from requiring both clinical and laboratory criteria to a lab-only definition. From June 2009 to November 2012, a major laboratory changed its reporting practices to include all positive enzyme immunoassay (EIA) results, resulting in a pronounced increase in reported cases. In October 2019, ADHS began entering and classifying cases with 1:2 complement fixation (CF) titers as confirmed across all laboratory tests; previously (2011–2019), serum CF titers required ≥1:4 for confirmation, while other laboratory tests (e.g., IgM EIA and IgG EIA) could be confirmatory at ≥1:2. We created a categorical variable (surv_regime) to account for these discrete surveillance-related artifacts in our analyses, with categories corresponding to the regimes shown in this figure. For January–March 2024, we applied a numeric correction by proportionally redistributing about 800 false-positive cases across all county-month records during this period; this was the only interval where direct quantitative evidence of data quality issues was available.

**Figure S3.** Monthly time series of coccidioidomycosis incidence and multilayer environmental drivers in the study region.

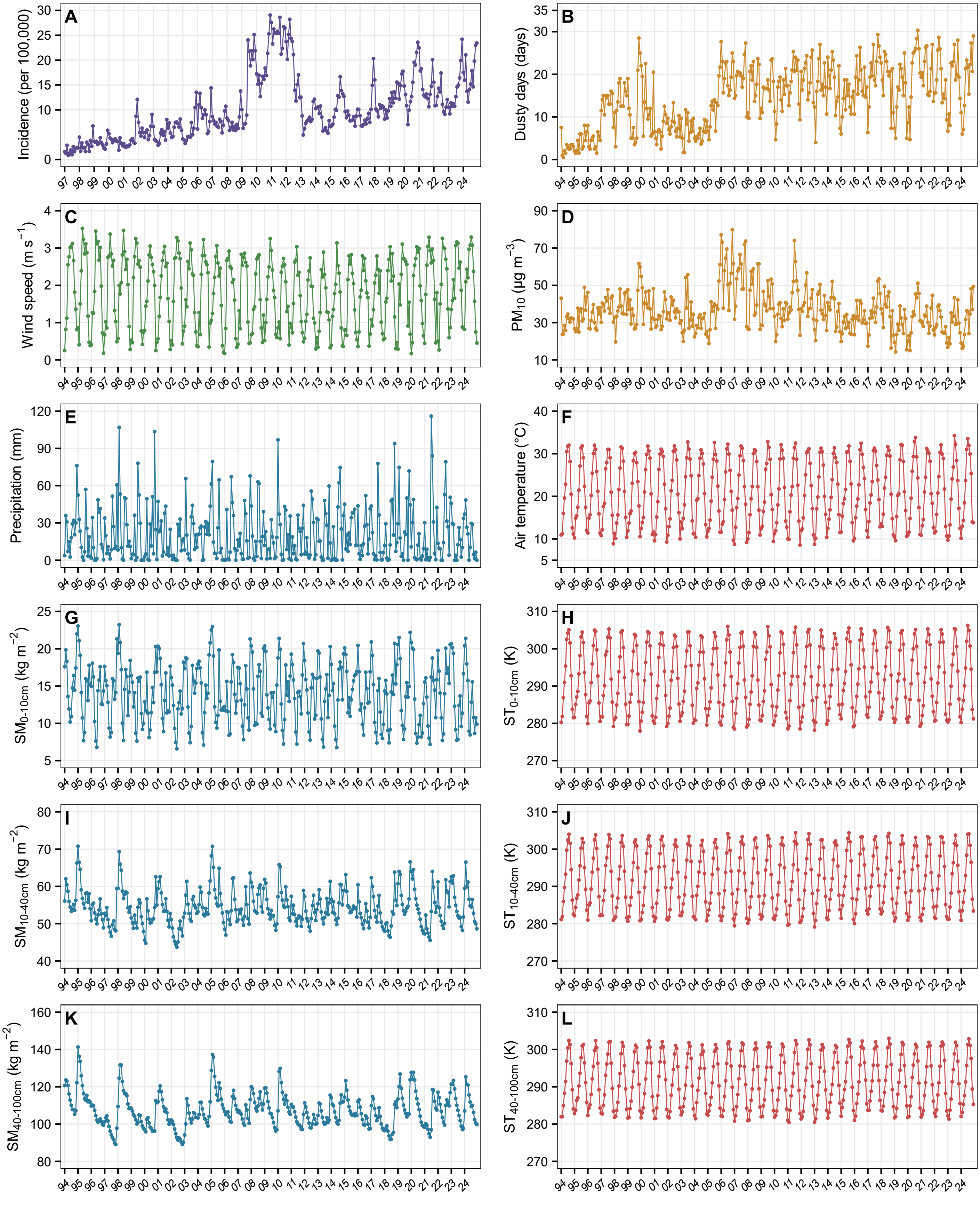

**Note:** Panels show monthly coccidioidomycosis incidence (A; cases per 100,000 population; 1997–2024) and monthly environmental exposures (B–L; 1994–2024): (B) mean monthly number of dusty days, (C) 10-m wind speed, (D) $PM_{10}$ concentration, (E) precipitation, (F) air temperature, (G, I, K) soil moisture at 0–10 cm, 10–40 cm, and 40–100 cm depths, and (H, J, L) soil temperature at 0–10 cm, 10–40 cm, and 40–100 cm depths. Points represent monthly values and lines connect consecutive months. Two-digit year labels are used on the x-axis (e.g., 94 = 1994).

**Figure S4.** Associations between winter (January–March) coccidioidomycosis incidence and lagged multilayer environmental exposures in the study region.

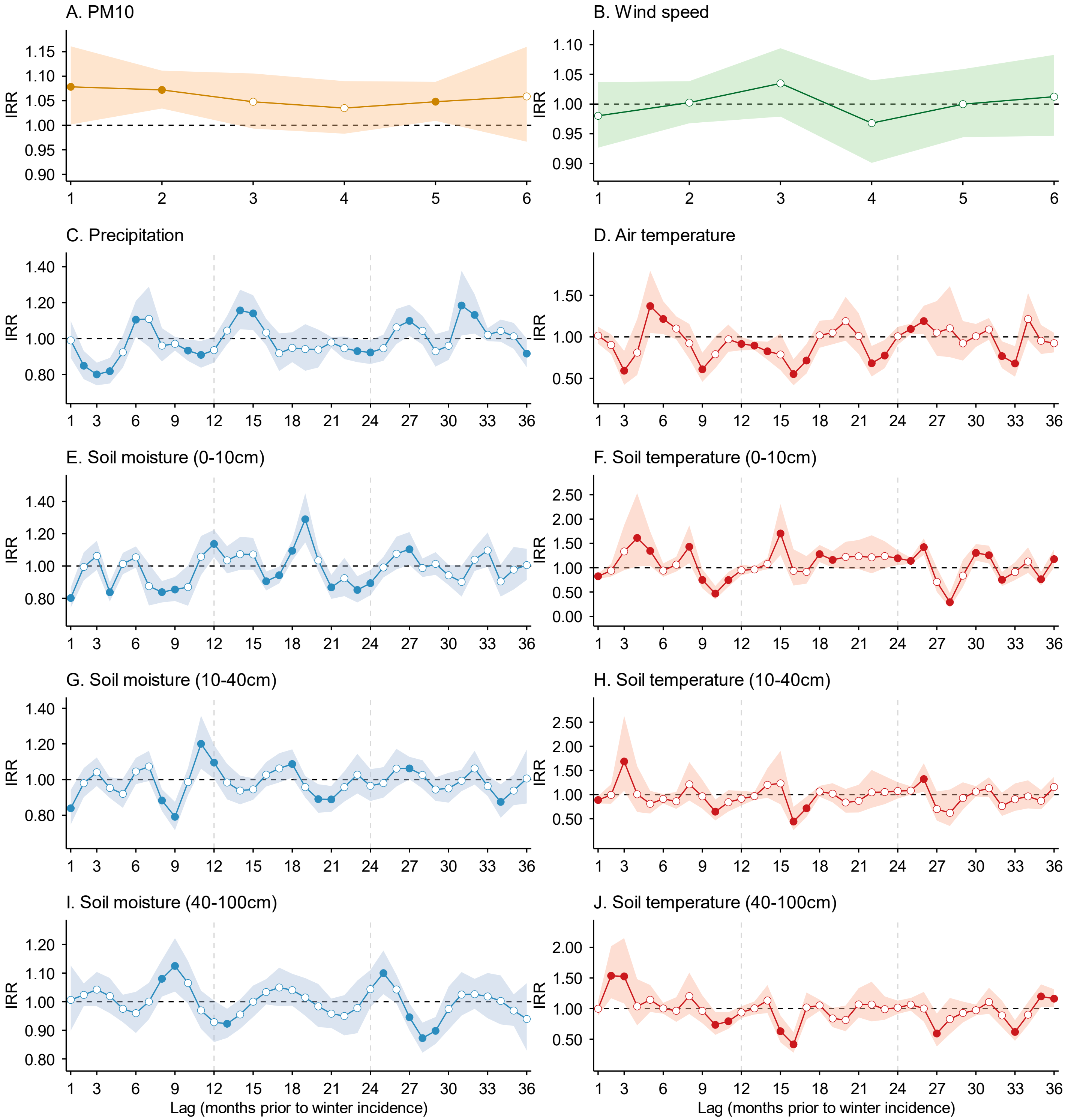

Incidence rate ratios (IRRs) and 95% confidence intervals (CIs; shaded areas) were estimated using distributed lag non-linear models (DLNMs) for an interquartile range (IQR) increase in each exposure at specific lag months prior to disease incidence. Panels show above-ground exposures from the dust dispersion layer (A: $PM_{10}$; B: 10-m wind speed; lags 1–6 months) and meteorological layer (C: precipitation; D: air temperature; lags 1–36 months), and below-ground exposures of soil moisture (E, G, I) and soil temperature (F, H, J) at 0–10 cm, 10–40 cm, and 40–100 cm depths (lags 1–36 months). Models were adjusted for spatiotemporal trends and surveillance-related changes in reporting and laboratory testing. The horizontal dashed line indicates null association (IRR = 1). Solid points indicate statistically significant associations; hollow points indicate non-significant associations. Corresponding numerical data are reported in Tables S3–S7.

**Figure S5.** Associations between spring (April–June) coccidioidomycosis incidence and lagged multilayer environmental exposures in the study region.

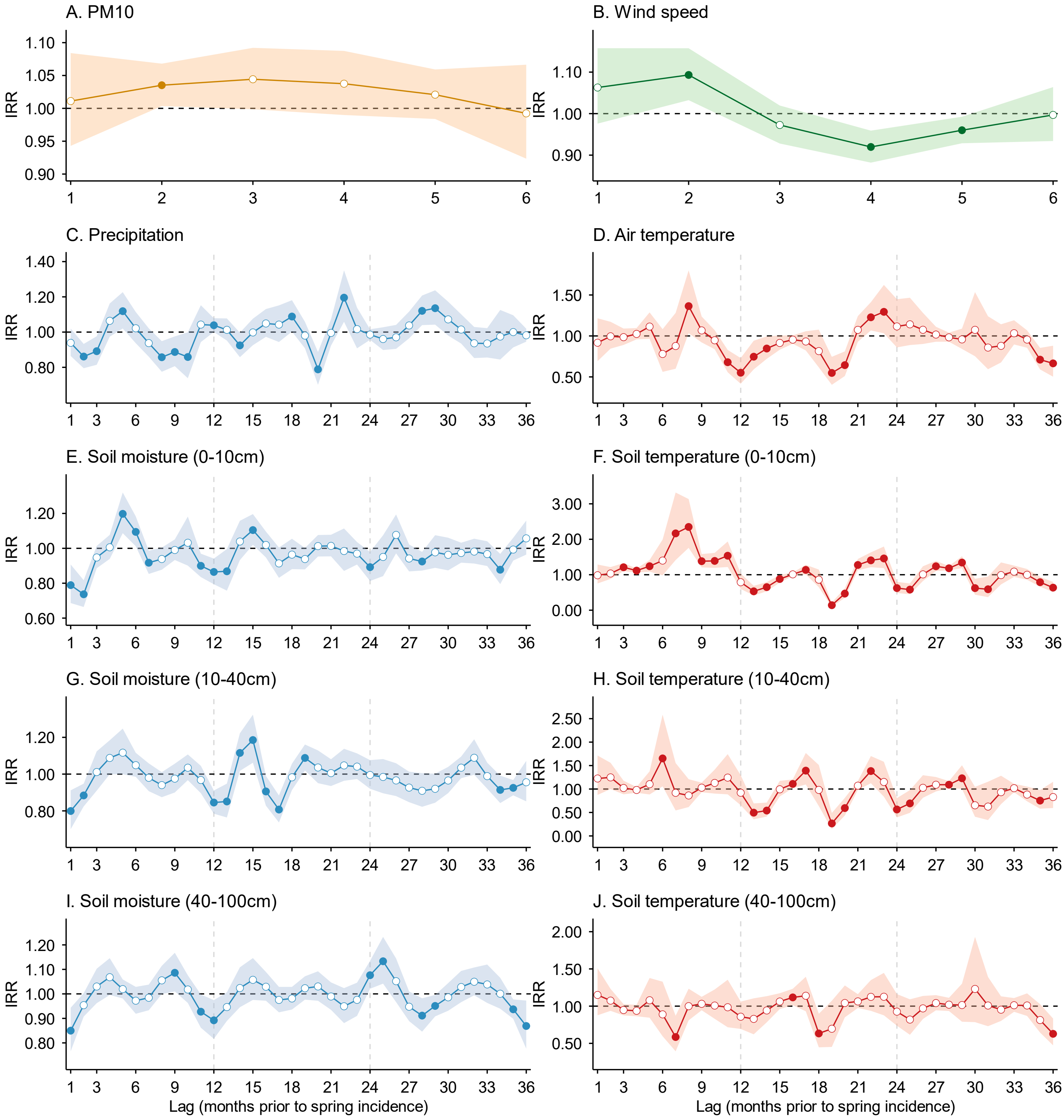

IRRs and 95% CIs (shaded areas) were estimated using DLNMs for an IQR increase in each exposure across lag months prior to disease incidence. Panels show above-ground exposures from the dust dispersion layer (A: $PM_{10}$; B: 10-m wind speed; lags 1–6 months) and meteorological layer (C: precipitation; D: air temperature; lags 1–36 months), and below-ground exposures of soil moisture (E, G, I) and soil temperature (F, H, J) at 0–10 cm, 10–40 cm, and 40–100 cm depths (lags 1–36 months). Models were adjusted for spatiotemporal trends and surveillance-related changes in reporting and laboratory testing. The horizontal dashed line indicates null association (IRR = 1). Solid points indicate statistically significant associations; hollow points indicate non-significant associations. Corresponding numerical data are reported in Tables S3–S7.

**Figure S6.** Associations between monsoon (July–September) coccidioidomycosis incidence and lagged multilayer environmental exposures in the study region.

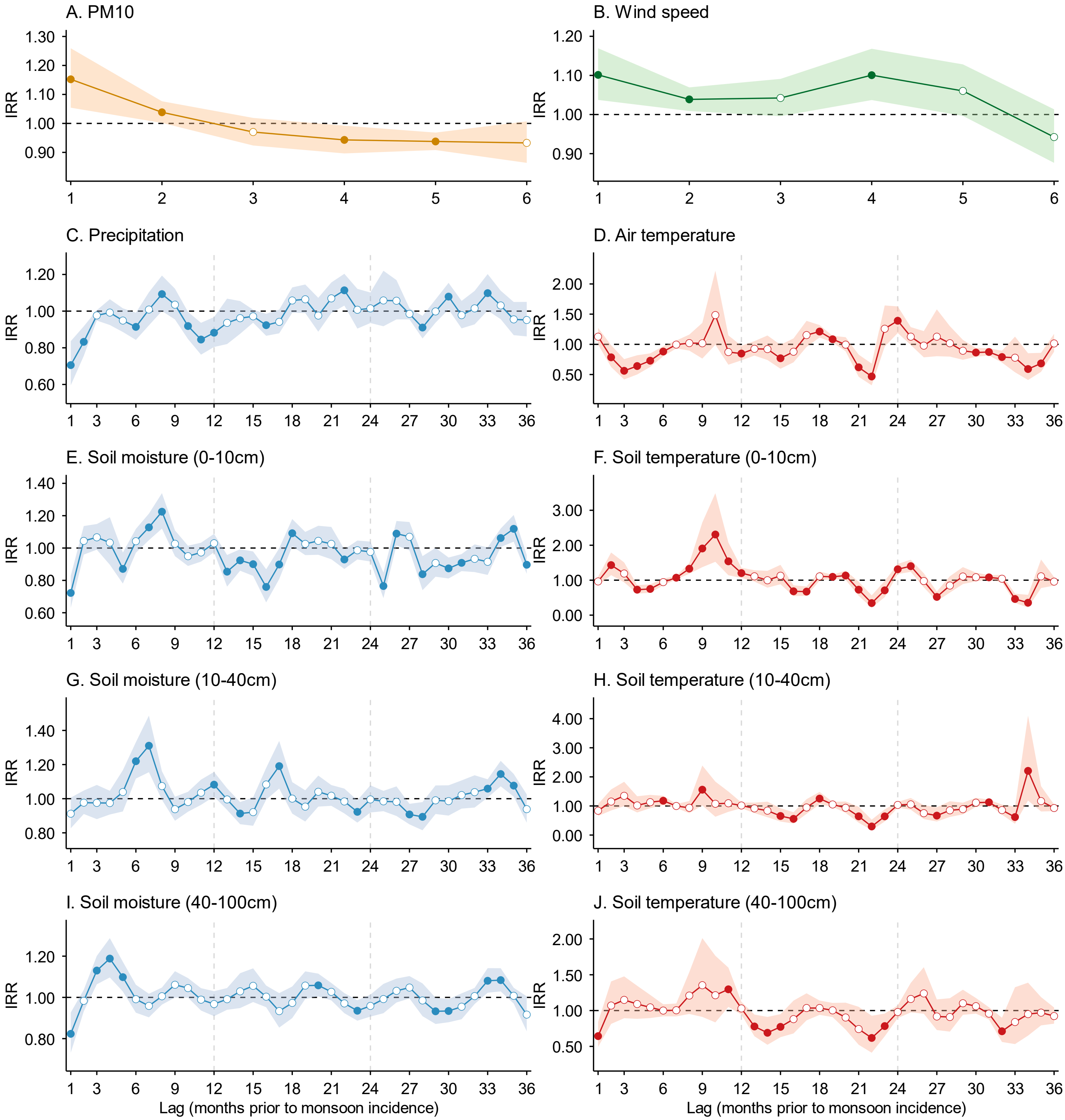

IRRs and 95% CIs (shaded areas) were estimated using DLNMs for an IQR increase in each exposure across lag months prior to disease incidence. Panels show above-ground exposures from the dust dispersion layer (A: $PM_{10}$; B: 10-m wind speed; lags 1–6 months) and meteorological layer (C: precipitation; D: air temperature; lags 1–36 months), and below-ground exposures of soil moisture (E, G, I) and soil temperature (F, H, J) at 0–10 cm, 10–40 cm, and 40–100 cm depths (lags 1–36 months). Models were adjusted for spatiotemporal trends and surveillance-related changes in reporting and laboratory testing. The horizontal dashed line indicates null association (IRR = 1). Solid points indicate statistically significant associations; hollow points indicate non-significant associations. Corresponding numerical data are reported in Tables S3–S7.

**Figure S7.** Associations between coccidioidomycosis incidence and lagged below-ground soil moisture (Layer 3: 0–10 cm; topsoil; SM0t10) across seasons.

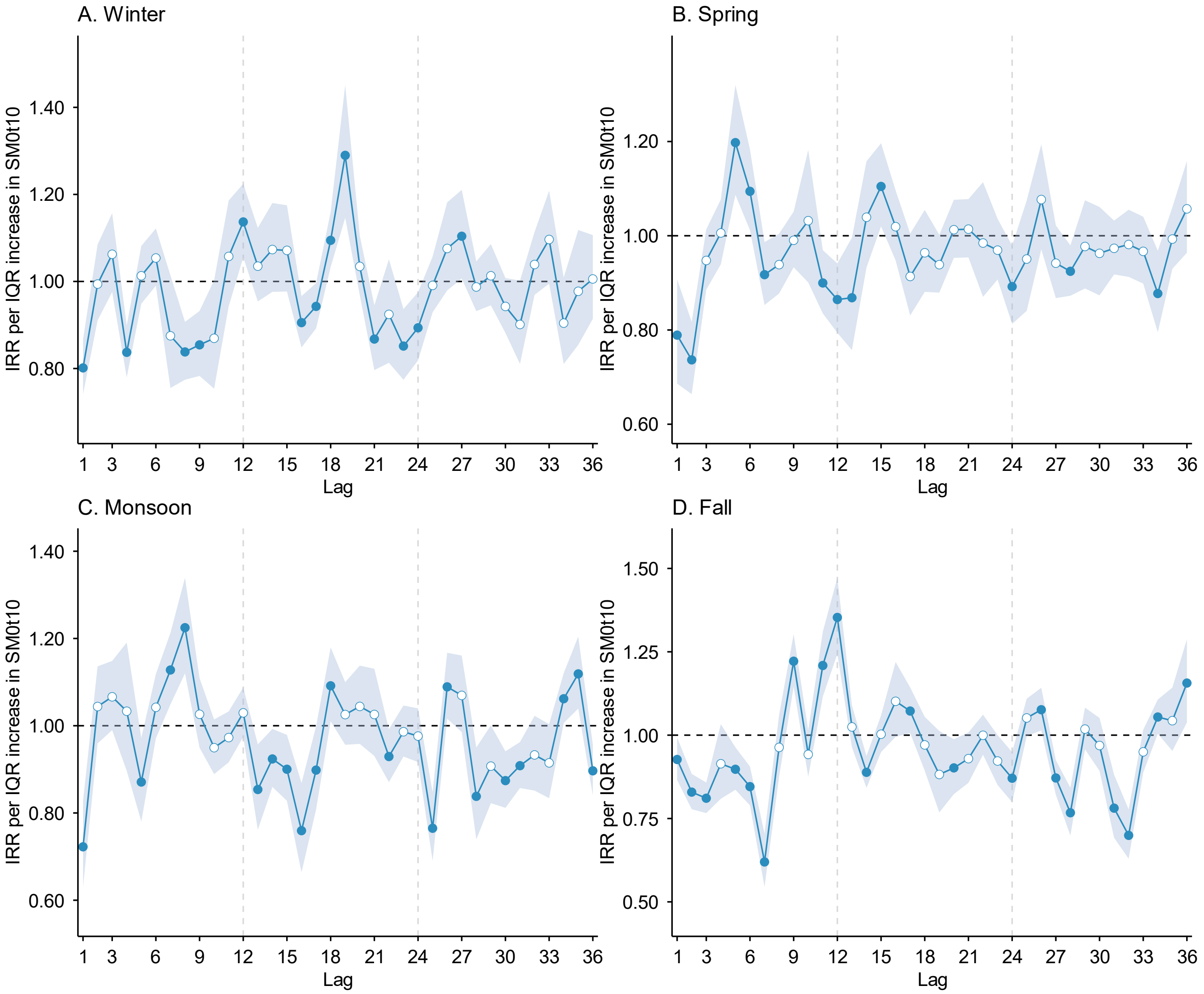

IRRs and 95% CIs (shaded areas) per IQR increase were estimated using DLNMs across lag months 1–36 preceding disease incidence, stratified by season (A: Winter; B: Spring; C: Monsoon; D: Fall). Models were adjusted for spatiotemporal trends and surveillance-related changes in reporting and laboratory testing. Layer 3 soil moisture and soil temperature were modeled simultaneously in a single DLNM with season-specific effects. The horizontal dashed line indicates null association (IRR = 1). Solid points indicate statistically significant associations; hollow points indicate non-significant associations. Corresponding numerical data are reported in Table S3.

**Figure S8.** Associations between coccidioidomycosis incidence and lagged below-ground soil temperature (Layer 3: 0–10 cm; topsoil; ST0t10) across seasons.

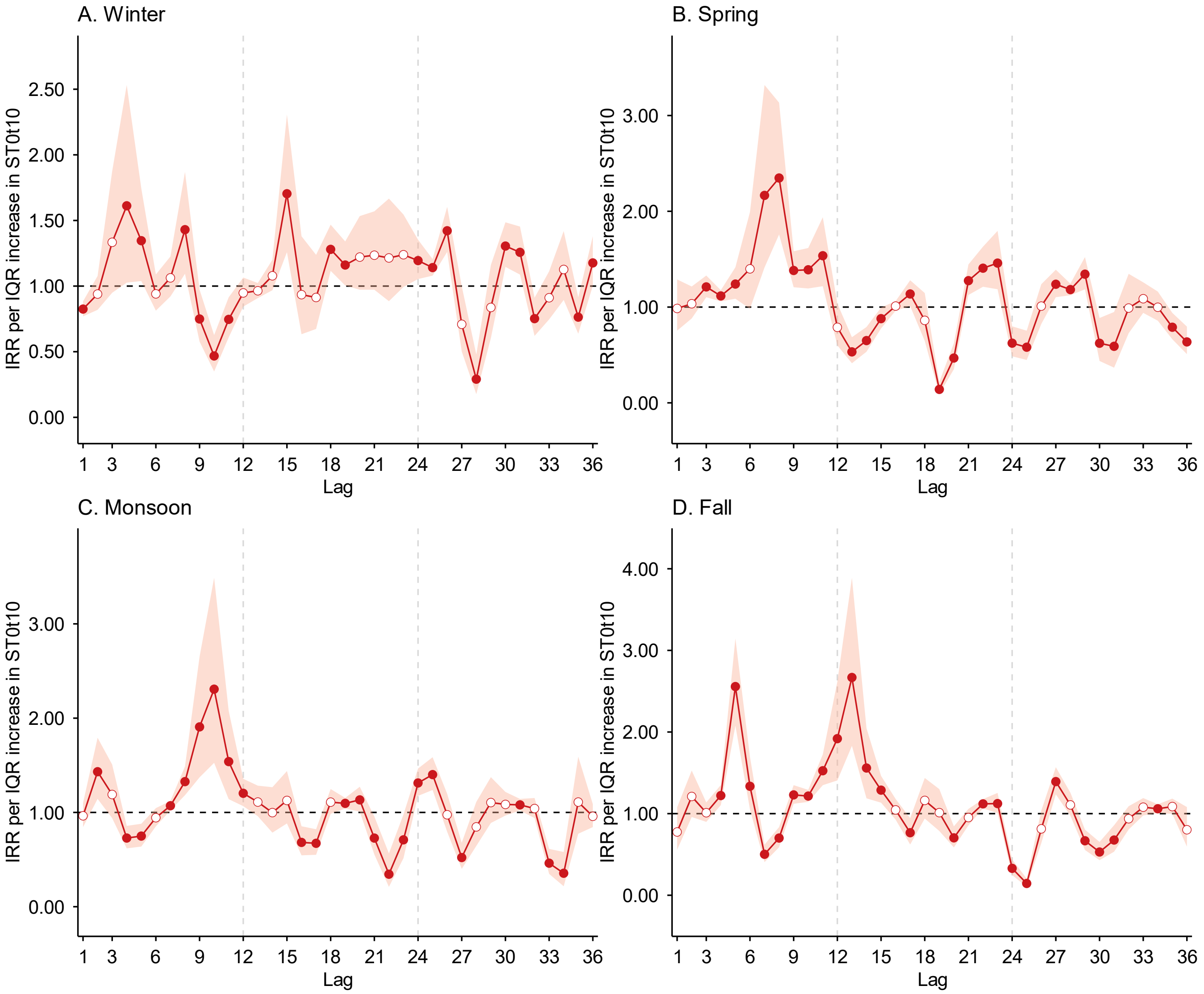

Log-transformed IRRs and 95% CIs (shaded areas) per IQR increase were estimated using DLNMs across lag months 1–36 preceding disease incidence, stratified by season (A: Winter; B: Spring; C: Monsoon; D: Fall). Models were adjusted for spatiotemporal trends and surveillance-related changes in reporting and laboratory testing. Layer 3 soil moisture and soil temperature were modeled simultaneously in a single DLNM with season-specific effects. The horizontal dashed line indicates null association (IRR = 1). Solid points indicate statistically significant associations; hollow points indicate non-significant associations. Corresponding numerical data are reported in Table S3.

**Figure S9.** Associations between coccidioidomycosis incidence and lagged below-ground soil moisture (Layer 4: 10–40 cm; SM10t40) across seasons.

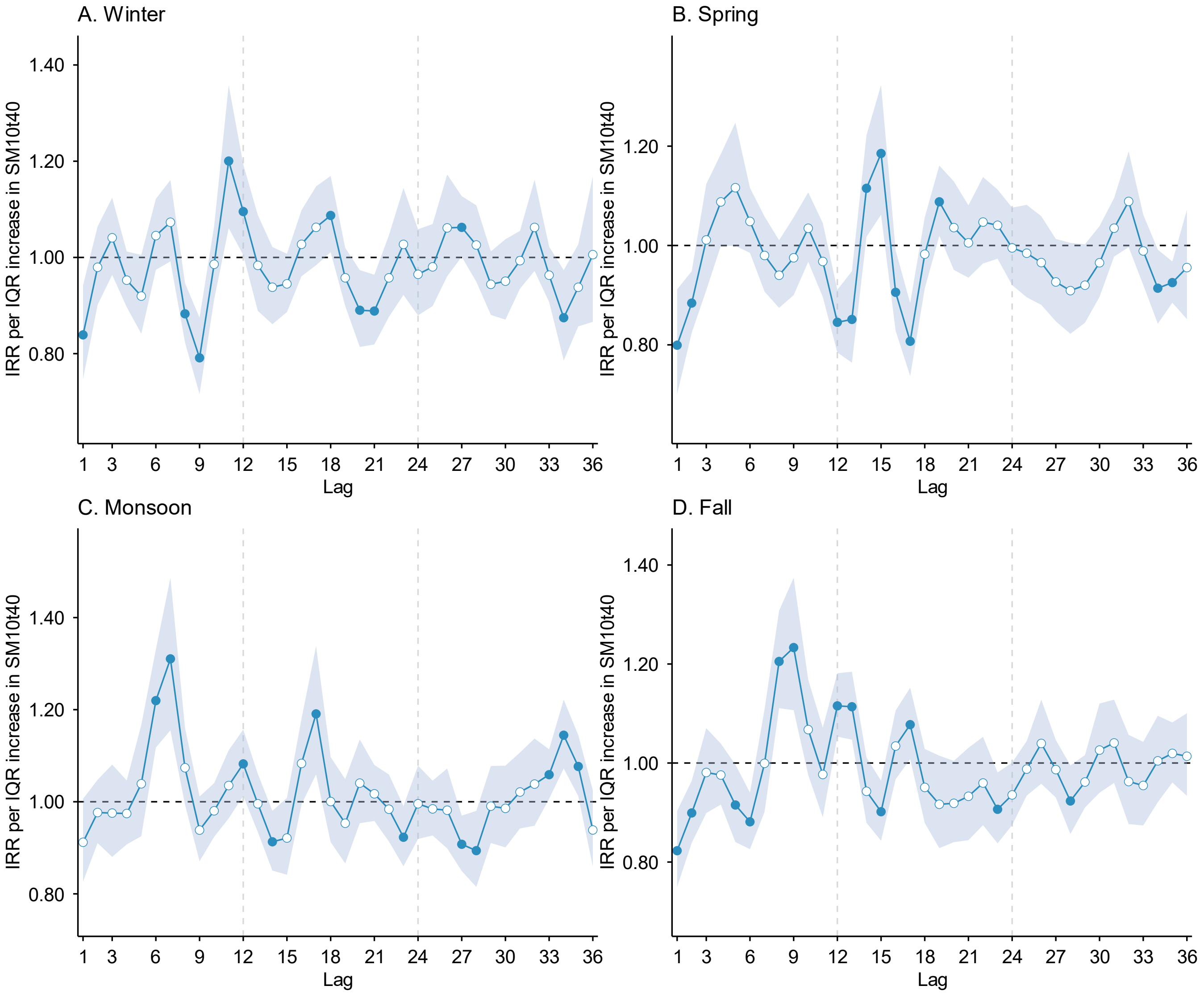

IRRs and 95% CIs (shaded areas) per IQR increase were estimated using DLNMs across lag months 1–36 preceding disease incidence, stratified by season (A: Winter; B: Spring; C: Monsoon; D: Fall). Models were adjusted for spatiotemporal trends and surveillance-related changes in reporting and laboratory testing. Layer 4 soil moisture and soil temperature were modeled simultaneously in a single DLNM with season-specific effects. The horizontal dashed line indicates null association (IRR = 1). Solid points indicate statistically significant associations; hollow points indicate non-significant associations. Corresponding numerical data are reported in Table S4.

**Figure S10.** Associations between coccidioidomycosis incidence and lagged below-ground soil temperature (Layer 4: 10–40 cm; ST10t40) across seasons.

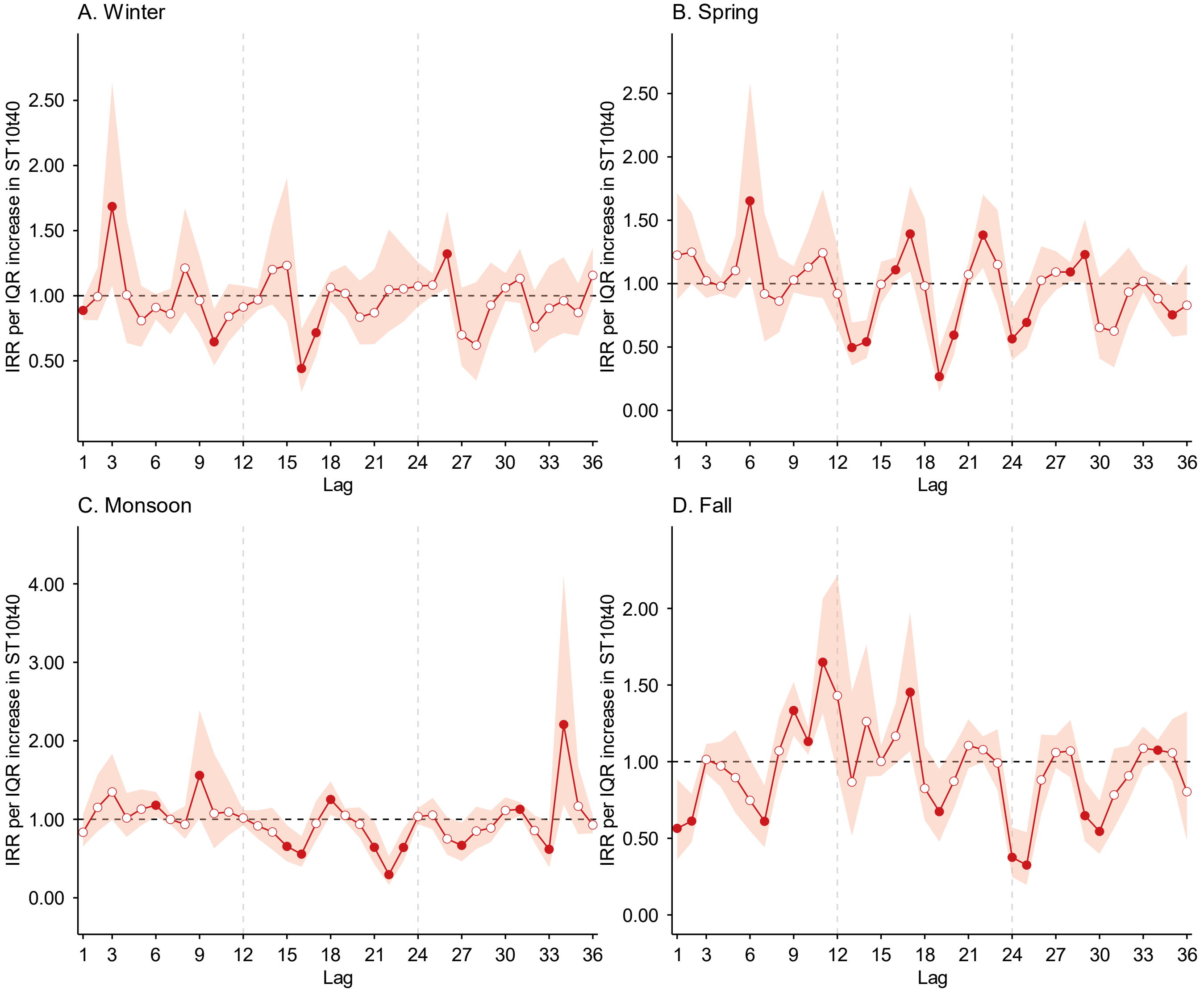

Log-transformed IRRs and 95% CIs (shaded areas) per IQR increase were estimated using DLNMs across lag months 1–36 preceding disease incidence, stratified by season (A: Winter; B: Spring; C: Monsoon; D: Fall). Models were adjusted for spatiotemporal trends and surveillance-related changes in reporting and laboratory testing. Layer 4 soil moisture and soil temperature were modeled simultaneously in a single DLNM with season-specific effects. The horizontal dashed line indicates null association (IRR = 1). Solid points indicate statistically significant associations; hollow points indicate non-significant associations. Corresponding numerical data are reported in Table S4.

**Figure S11.** Associations between coccidioidomycosis incidence and lagged below-ground soil moisture (Layer 5: 40–100 cm; SM40t100) across seasons.

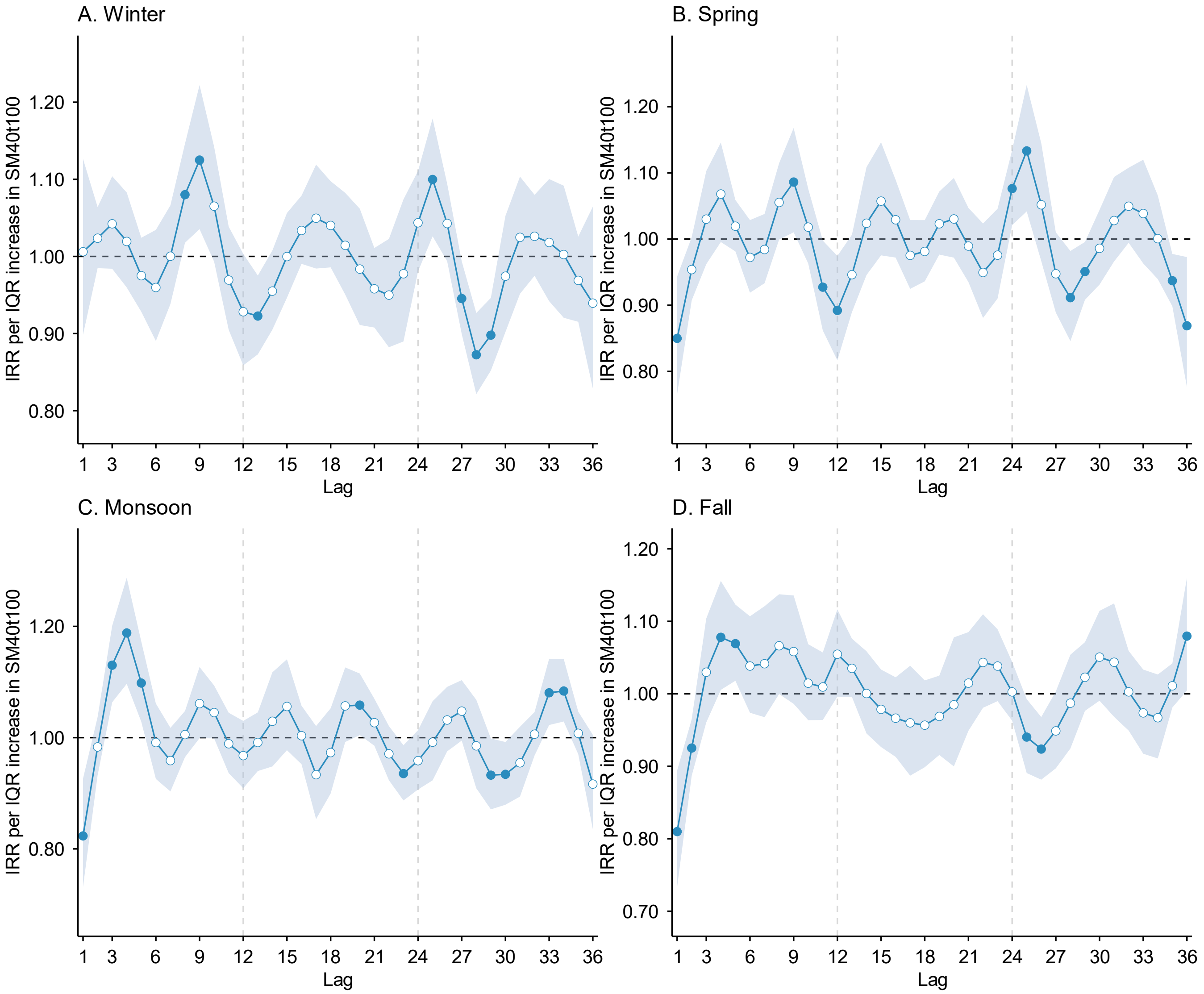

IRRs and 95% CIs (shaded areas) per IQR increase were estimated using DLNMs across lag months 1–36 preceding disease incidence, stratified by season (A: Winter; B: Spring; C: Monsoon; D: Fall). Models were adjusted for spatiotemporal trends and surveillance-related changes in reporting and laboratory testing. Layer 5 soil moisture and soil temperature were modeled simultaneously in a single DLNM with season-specific effects. The horizontal dashed line indicates null association (IRR = 1). Solid points indicate statistically significant associations; hollow points indicate non-significant associations. Corresponding numerical data are reported in Table S5.

**Figure S12.** Associations between coccidioidomycosis incidence and lagged below-ground soil temperature (Layer 5: 40–100 cm; ST40t100) across seasons.

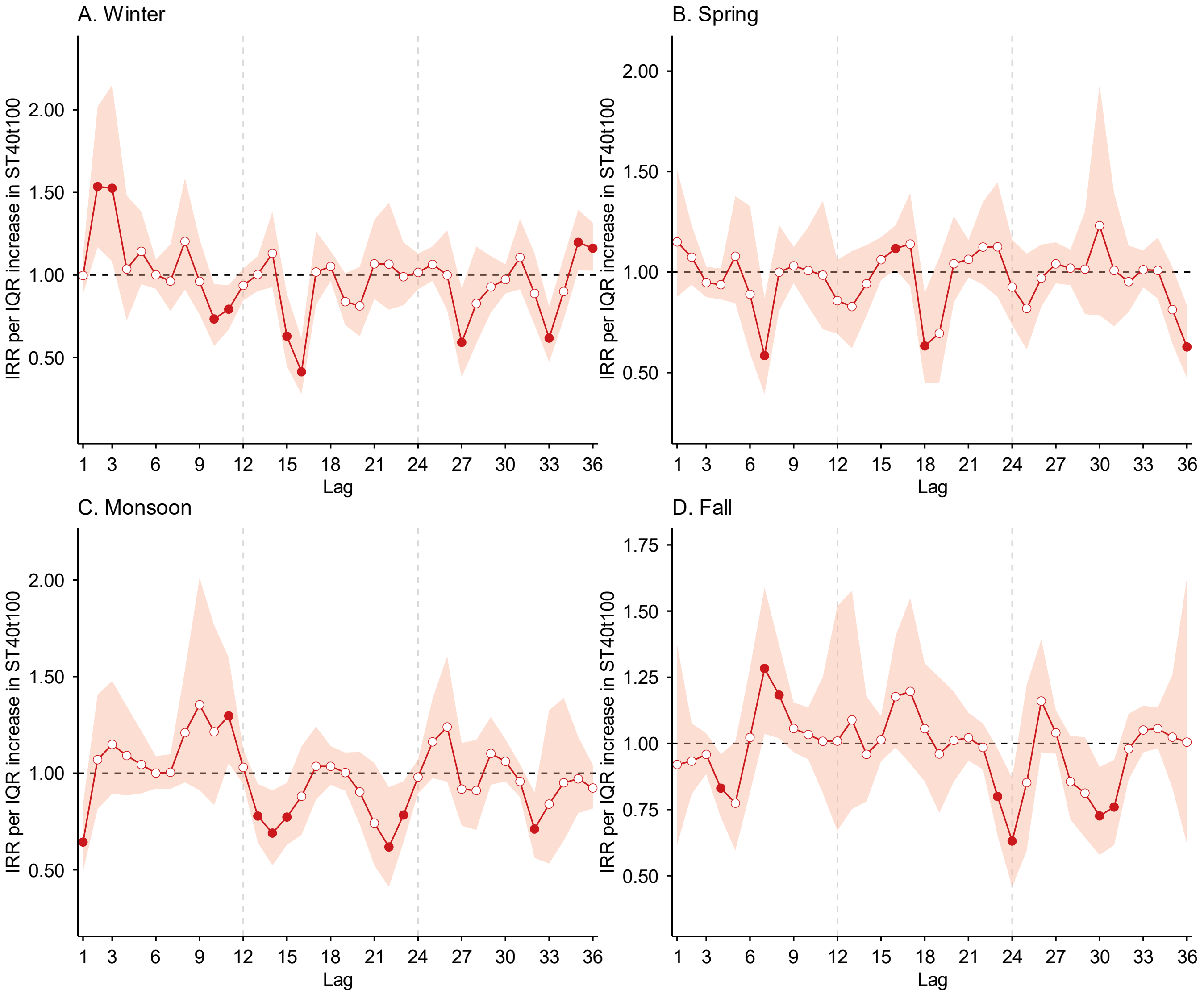

Log-transformed IRRs and 95% CIs (shaded areas) per IQR increase were estimated using DLNMs across lag months 1–36 preceding disease incidence, stratified by season (A: Winter; B: Spring; C: Monsoon; D: Fall). Models were adjusted for spatiotemporal trends and surveillance-related changes in reporting and laboratory testing. Layer 5 soil moisture and soil temperature were modeled simultaneously in a single DLNM with season-specific effects. The horizontal dashed line indicates null association (IRR = 1). Solid points indicate statistically significant associations; hollow points indicate non-significant associations. Corresponding numerical data are reported in Table S5.

**Figure S13.** Associations between coccidioidomycosis incidence and lagged above-ground precipitation (Layer 2: meteorological layer) across seasons.

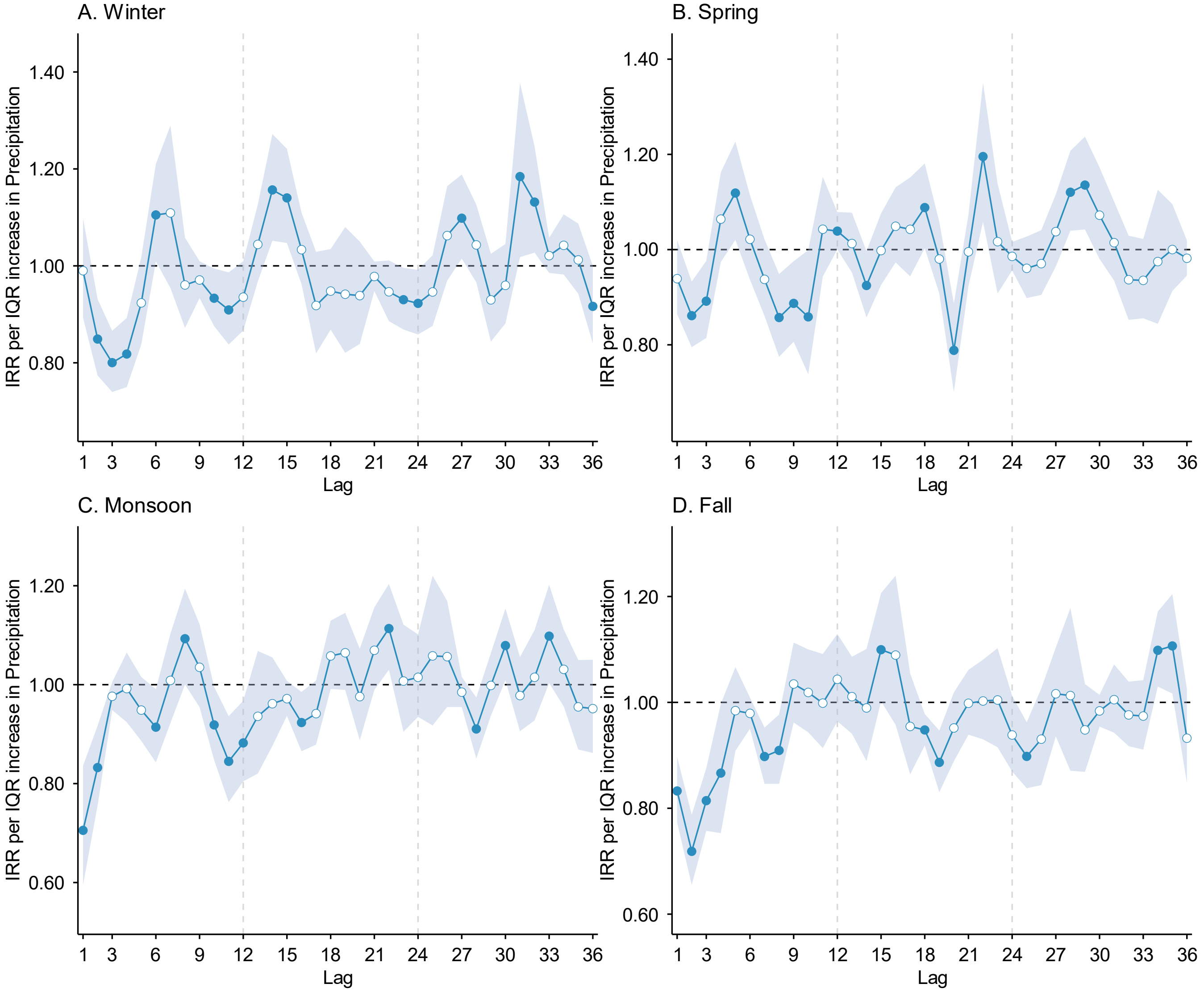

IRRs and 95% CIs (shaded areas) per IQR increase were estimated using DLNMs across lag months 1–36 preceding disease incidence, stratified by season (A: Winter; B: Spring; C: Monsoon; D: Fall). Models were adjusted for spatiotemporal trends and surveillance-related changes in reporting and laboratory testing. Layer 2 meteorological conditions (precipitation and air temperature) were modeled simultaneously in a single DLNM with season-specific effects. The horizontal dashed line indicates null association (IRR = 1). Solid points indicate statistically significant associations; hollow points indicate non-significant associations. Corresponding numerical data are reported in Table S6.

**Figure S14.** Associations between coccidioidomycosis incidence and lagged above-ground air temperature (Layer 2: meteorological layer; AT) across seasons.

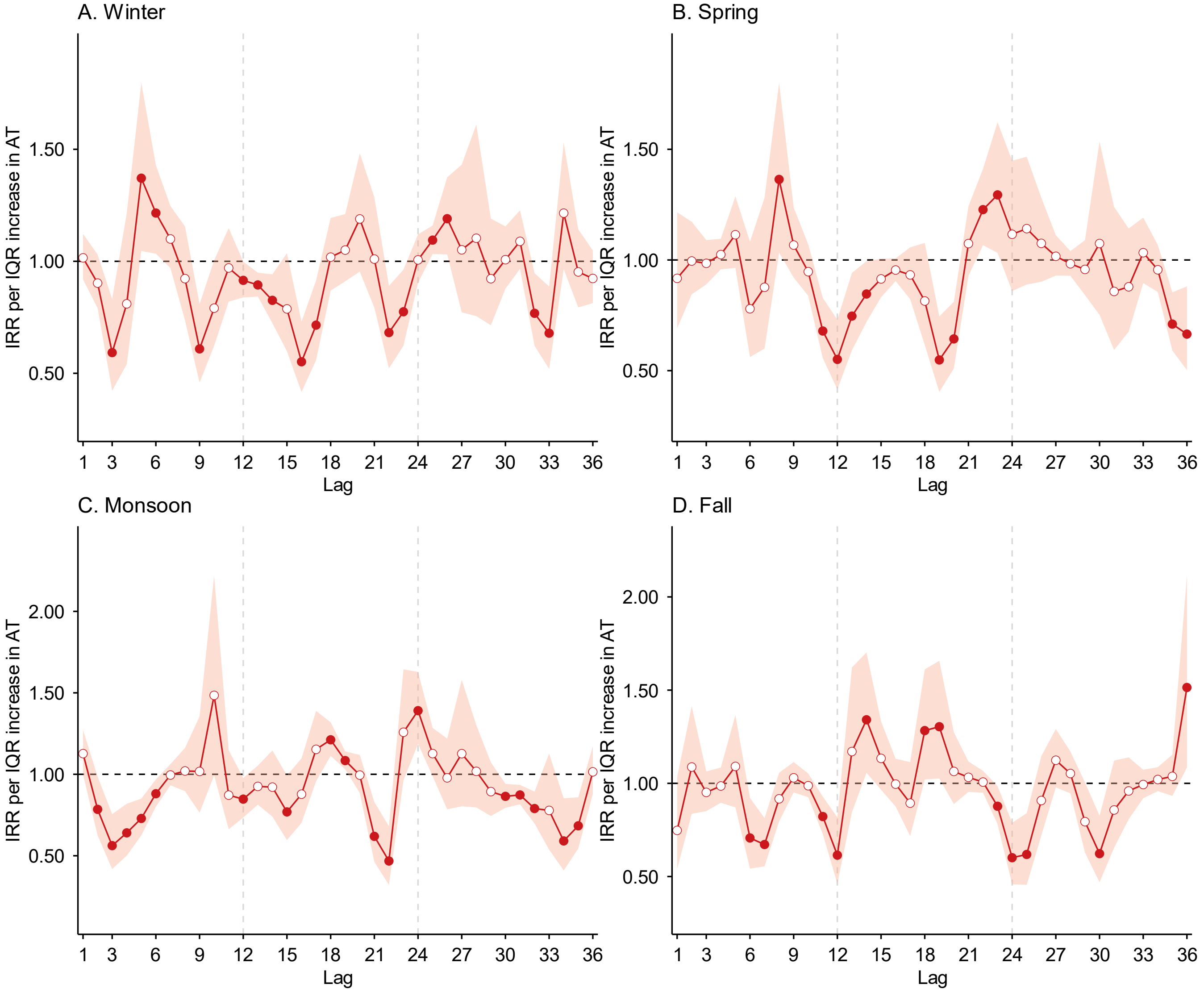

Log-transformed IRRs and 95% CIs (shaded areas) per IQR increase were estimated using DLNMs across lag months 1–36 preceding disease incidence, stratified by season (A: Winter; B: Spring; C: Monsoon; D: Fall). Models were adjusted for spatiotemporal trends and surveillance-related changes in reporting and laboratory testing. Layer 2 meteorological conditions (precipitation and AT) were modeled simultaneously in a single DLNM with season-specific effects. The horizontal dashed line indicates null association (IRR = 1). Solid points indicate statistically significant associations; hollow points indicate non-significant associations. Corresponding numerical data are reported in Table S6.

**Figure S15.** Associations between coccidioidomycosis incidence and lagged above-ground $PM_{10}$ (Layer 1: dust dispersion layer) across seasons.

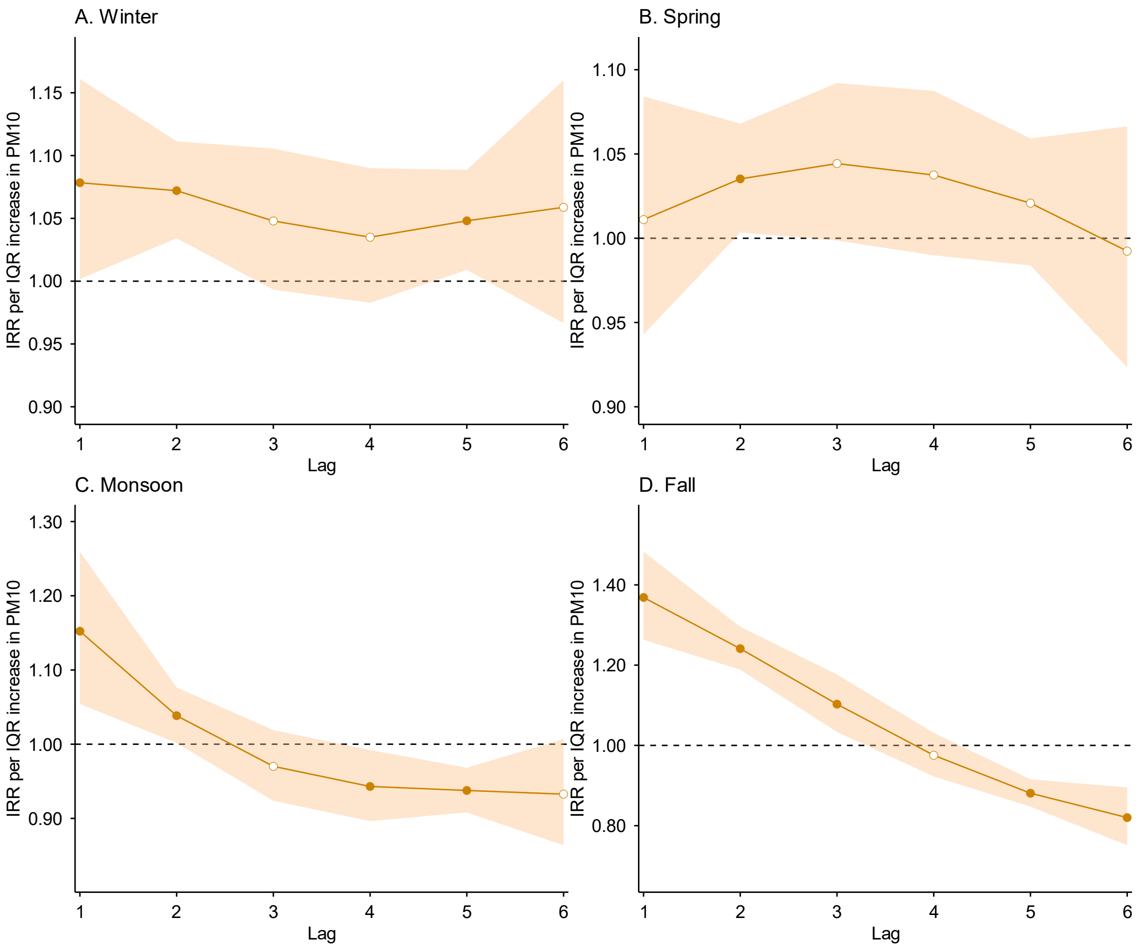

IRRs and 95% CIs (shaded areas) per IQR increase were estimated using DLNMs across lag months 1–6 preceding disease incidence, stratified by season (A: Winter; B: Spring; C: Monsoon; D: Fall). Models were adjusted for spatiotemporal trends and surveillance-related changes in reporting and laboratory testing. Layer 1 dust dispersion conditions ($PM_{10}$ and wind speed) were modeled simultaneously in a single DLNM with season-specific effects. The horizontal dashed line indicates null association (IRR = 1). Solid points indicate statistically significant associations; hollow points indicate non-significant associations. Corresponding numerical data are reported in Table S7.

**Figure S16.** Associations between coccidioidomycosis incidence and lagged above-ground wind speed (Layer 1: dust dispersion layer) across seasons.

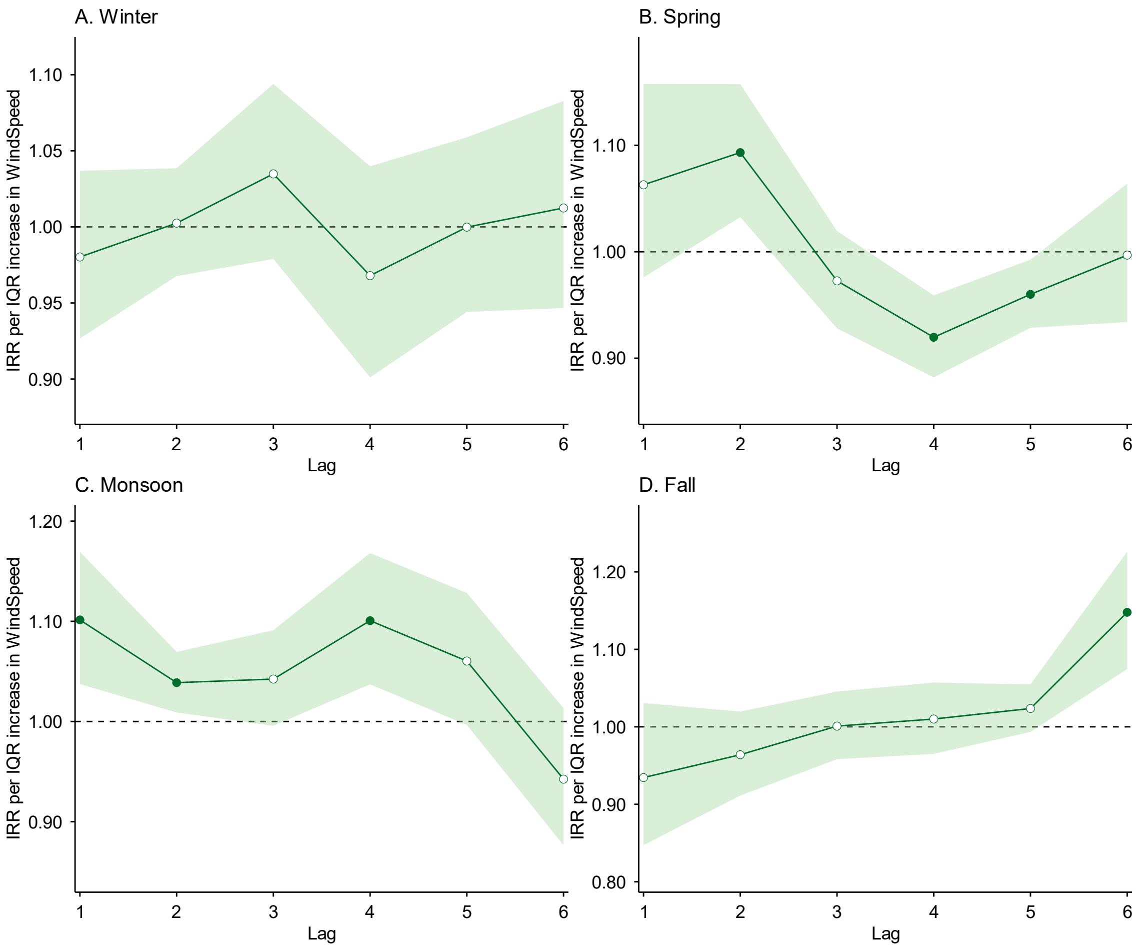

Log-transformed IRRs and 95% CIs (shaded areas) per IQR increase were estimated using DLNMs across lag months 1–36 preceding disease incidence, stratified by season (A: Winter; B: Spring; C: Monsoon; D: Fall). Models were adjusted for spatiotemporal trends and surveillance-related changes in reporting and laboratory testing. Layer 1 dust dispersion conditions ($PM_{10}$ and wind speed) were modeled simultaneously in a single DLNM with season-specific effects. The horizontal dashed line indicates null association (IRR = 1). Solid points indicate statistically significant associations; hollow points indicate non-significant associations. Corresponding numerical data are reported in Table S7.

**Figure S17.** Lag-specific exposure–response relationships between winter (January–March) coccidioidomycosis incidence and below-ground soil moisture (Layer 3: 0–10 cm; topsoil; SM0t10) at lags 1–36 months.

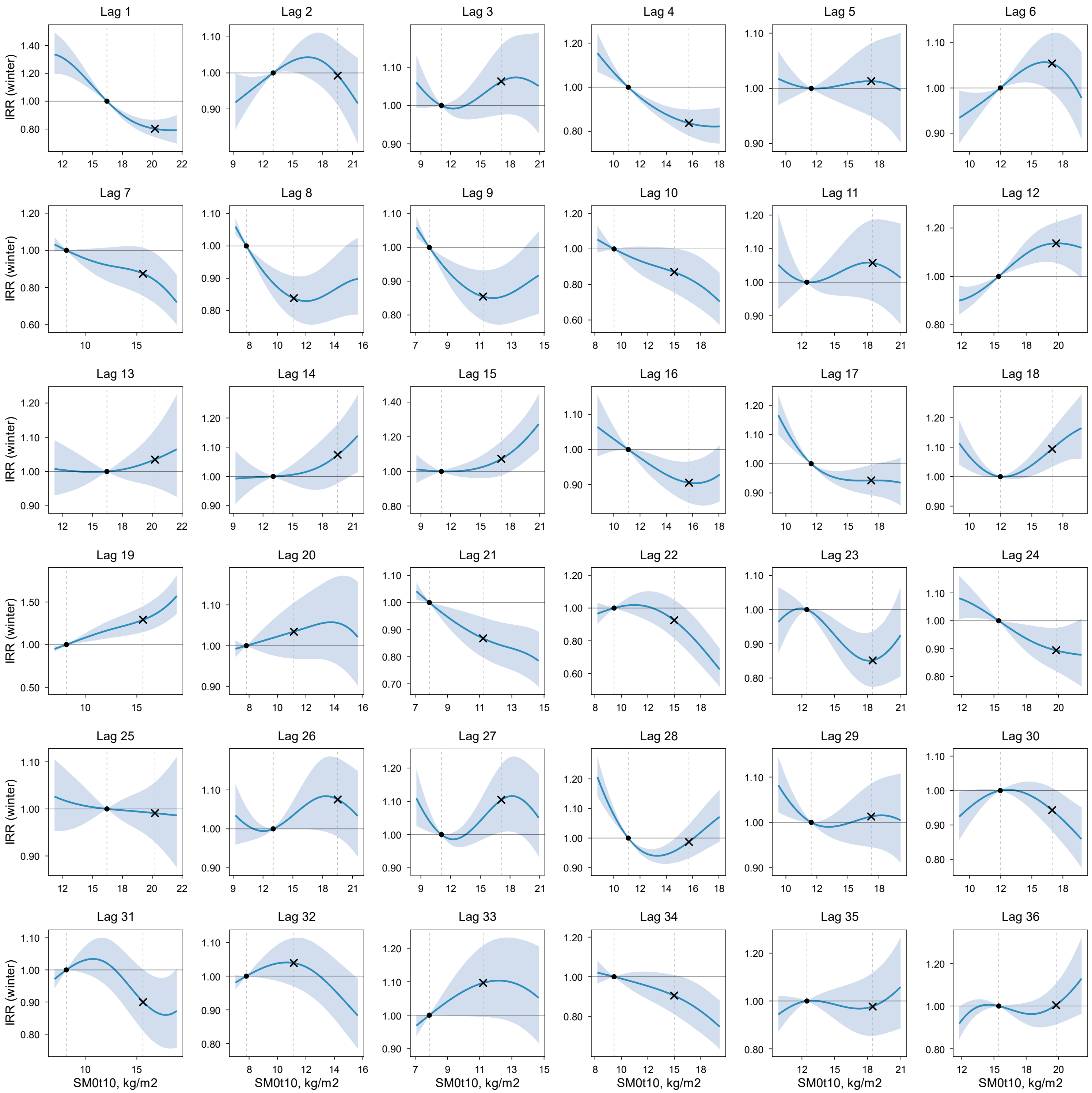

IRR (solid line) and 95% CI (shaded area) were estimated using a DLNM. Within each panel (lag month), IRR is expressed relative to the 25th percentile of the exposure variable in the study region (reference; IRR = 1), indicated by the left vertical dashed line and the dot on the IRR = 1 line. The right vertical dashed line indicates the 75th percentile, with the "×" symbol marking the estimated IRR at that level. Models were adjusted for spatiotemporal trends and surveillance-related changes in reporting and laboratory testing. Layer 3 soil moisture and soil temperature were modeled simultaneously in a single DLNM with season-specific effects.

**Figure S18.** Lag-specific exposure–response relationships between winter (January–March) coccidioidomycosis incidence and below-ground soil temperature (Layer 3: 0–10 cm; topsoil; ST0t10) at lags 1–36 months.

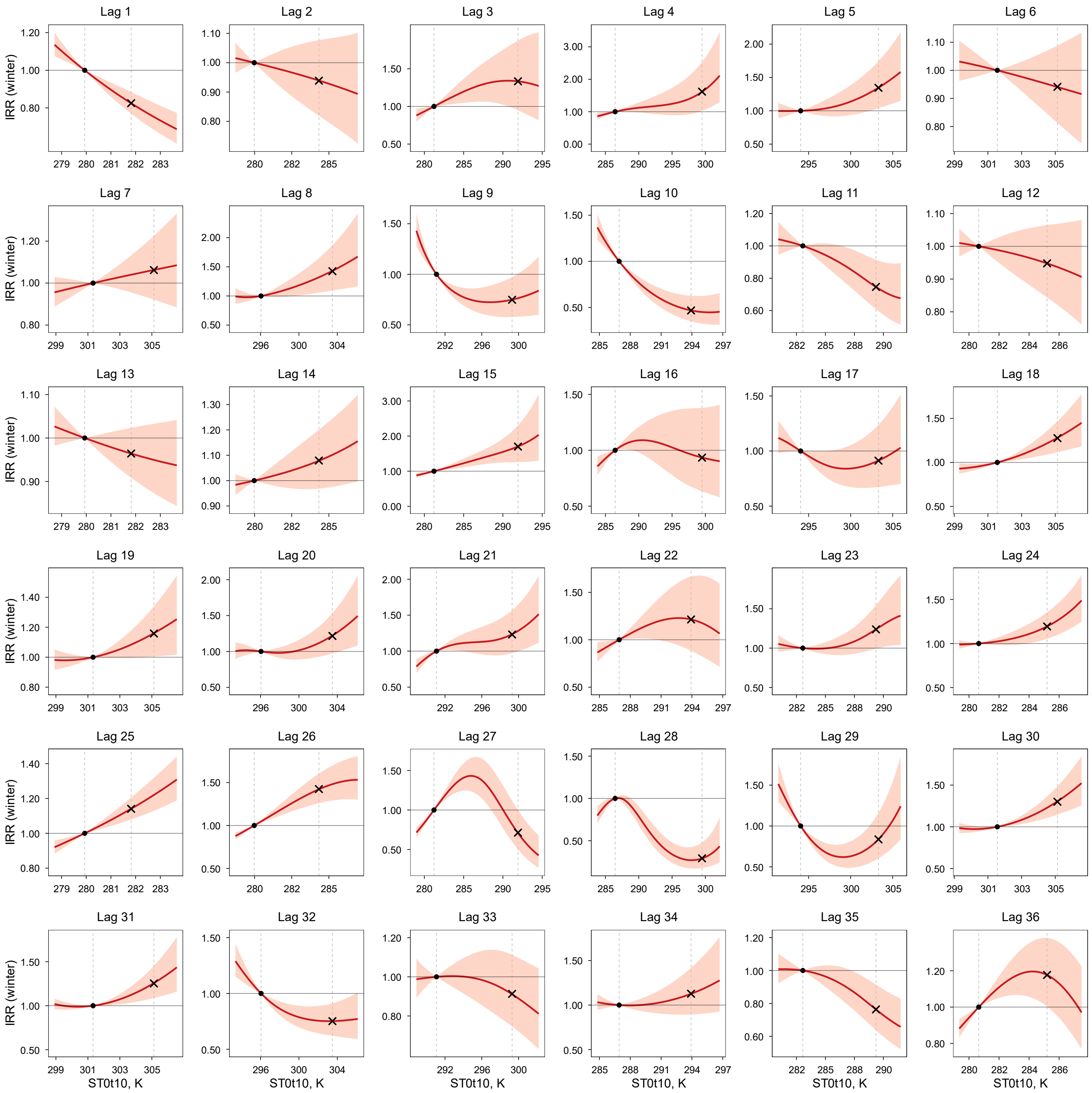

IRR (solid line) and 95% CI (shaded area) were estimated using a DLNM. Within each panel (lag month), IRR is expressed relative to the 25th percentile of the exposure variable in the study region (reference; IRR = 1), indicated by the left vertical dashed line and the dot on the IRR = 1 line. The right vertical dashed line indicates the 75th percentile, with the "×" symbol marking the estimated IRR at that level. Models were adjusted for spatiotemporal trends and surveillance-related changes in reporting and laboratory testing. Layer 3 soil moisture and soil temperature were modeled simultaneously in a single DLNM with season-specific effects.

**Figure S19.** Lag-specific exposure–response relationships between winter (January–March) coccidioidomycosis incidence and below-ground soil moisture (Layer 4: 10–40 cm; SM10t40) at lags 1–36 months.

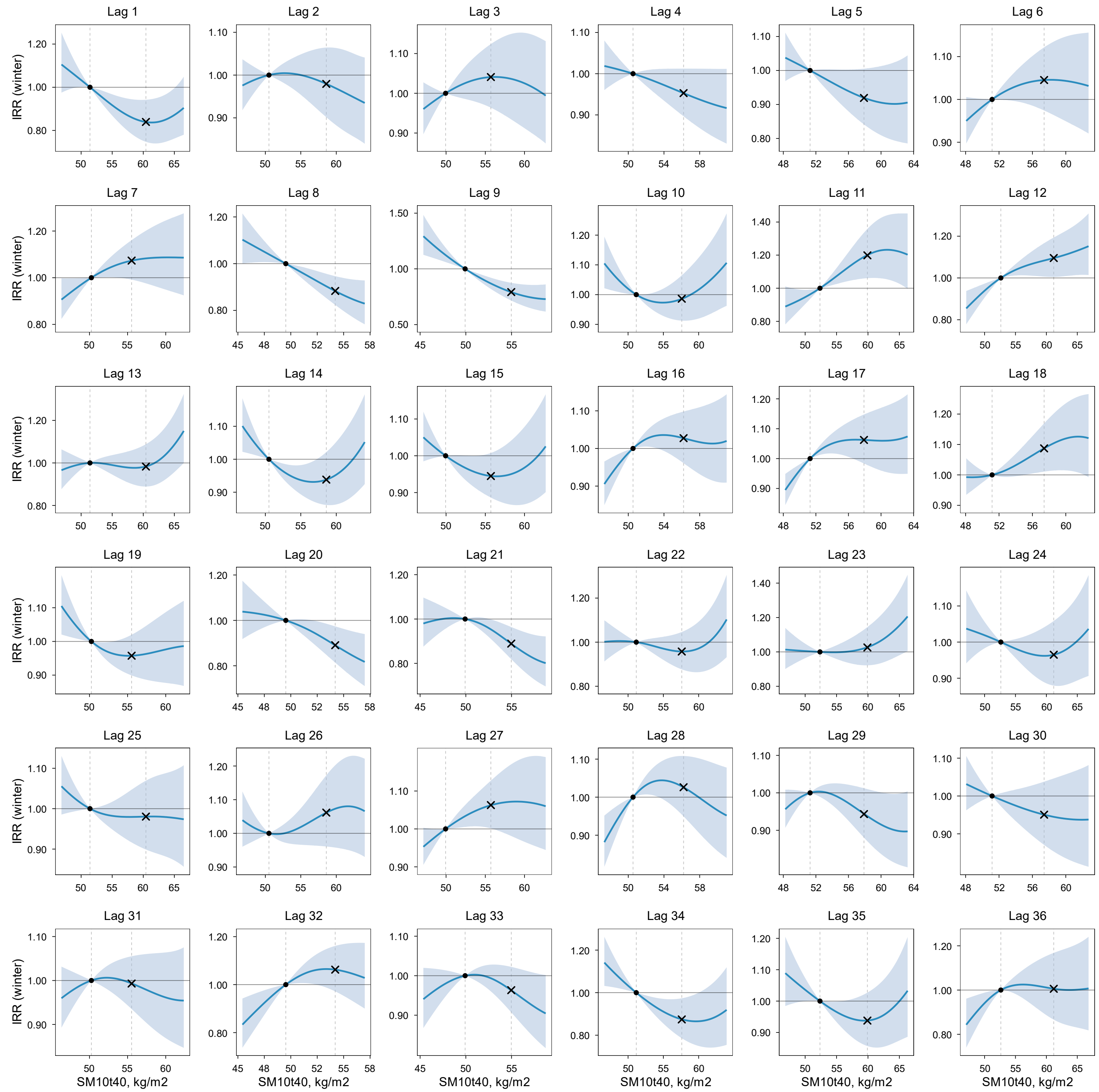

IRR (solid line) and 95% CI (shaded area) were estimated using a DLNM. Within each panel (lag month), IRR is expressed relative to the 25th percentile of the exposure variable in the study region (reference; IRR = 1), indicated by the left vertical dashed line and the dot on the IRR = 1 line. The right vertical dashed line indicates the 75th percentile, with the "×" symbol marking the estimated IRR at that level. Models were adjusted for spatiotemporal trends and surveillance-related changes in reporting and laboratory testing. Layer 4 soil moisture and soil temperature were modeled simultaneously in a single DLNM with season-specific effects.

**Figure S20.** Lag-specific exposure–response relationships between winter (January–March) coccidioidomycosis incidence and below-ground soil temperature (Layer 4: 10–40 cm; ST10t40) at lags 1–36 months.

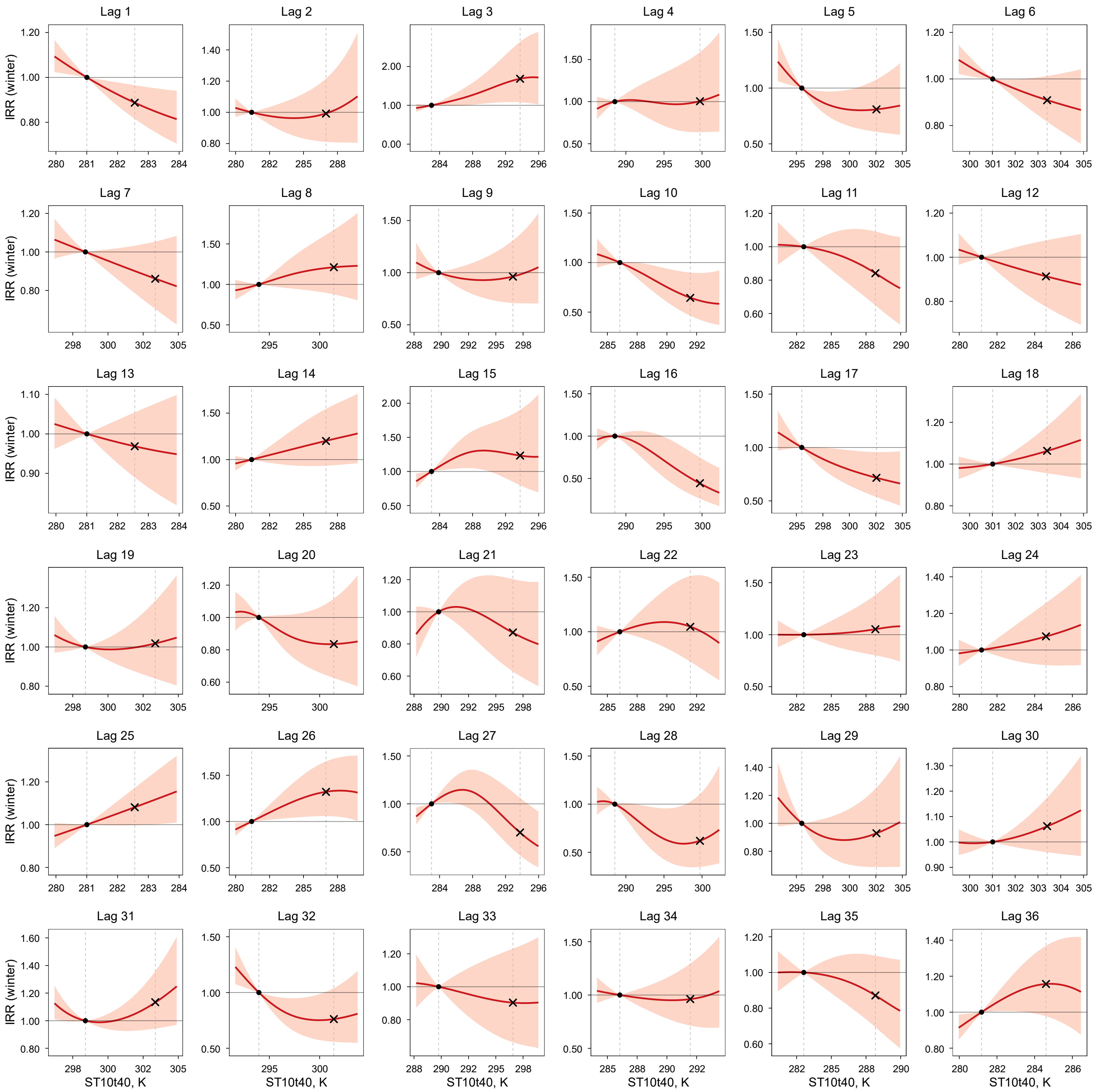

IRR (solid line) and 95% CI (shaded area) were estimated using a DLNM. Within each panel (lag month), IRR is expressed relative to the 25th percentile of the exposure variable in the study region (reference; IRR = 1), indicated by the left vertical dashed line and the dot on the IRR = 1 line. The right vertical dashed line indicates the 75th percentile, with the "×" symbol marking the estimated IRR at that level. Models were adjusted for spatiotemporal trends and surveillance-related changes in reporting and laboratory testing. Layer 4 soil moisture and soil temperature were modeled simultaneously in a single DLNM with season-specific effects.

**Figure S21.** Lag-specific exposure–response relationships between winter (January–March) coccidioidomycosis incidence and below-ground soil moisture (Layer 5: 40–100 cm; SM40t100) at lags 1–36 months.

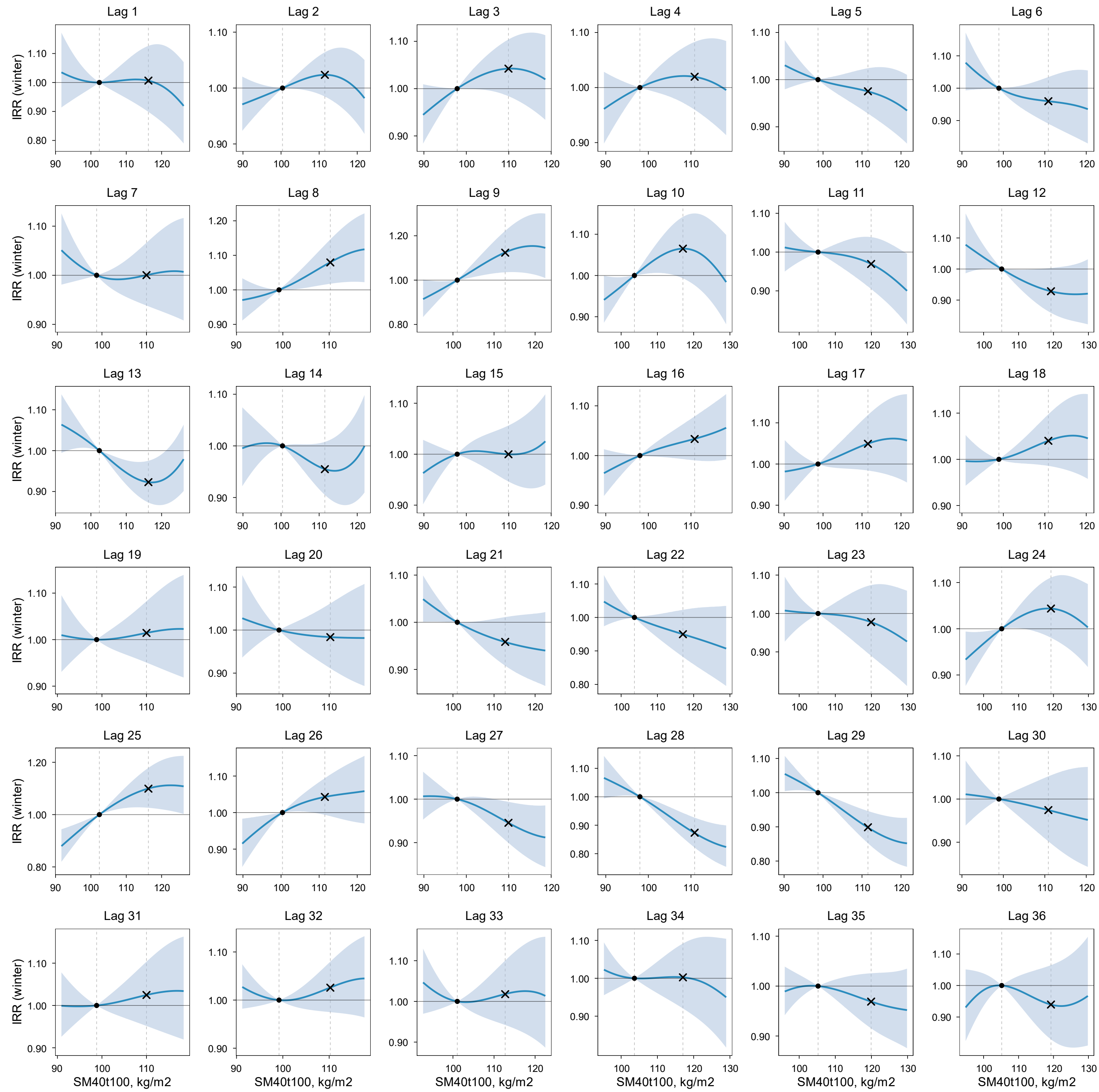

IRR (solid line) and 95% CI (shaded area) were estimated using a DLNM. Within each panel (lag month), IRR is expressed relative to the 25th percentile of the exposure variable in the study region (reference; IRR = 1), indicated by the left vertical dashed line and the dot on the IRR = 1 line. The right vertical dashed line indicates the 75th percentile, with the "×" symbol marking the estimated IRR at that level. Models were adjusted for spatiotemporal trends and surveillance-related changes in reporting and laboratory testing. Layer 5 soil moisture and soil temperature were modeled simultaneously in a single DLNM with season-specific effects.

**Figure S22.** Lag-specific exposure–response relationships between winter (January–March) coccidioidomycosis incidence and below-ground soil temperature (Layer 5: 40–100 cm; ST40t100) at lags 1–36 months.

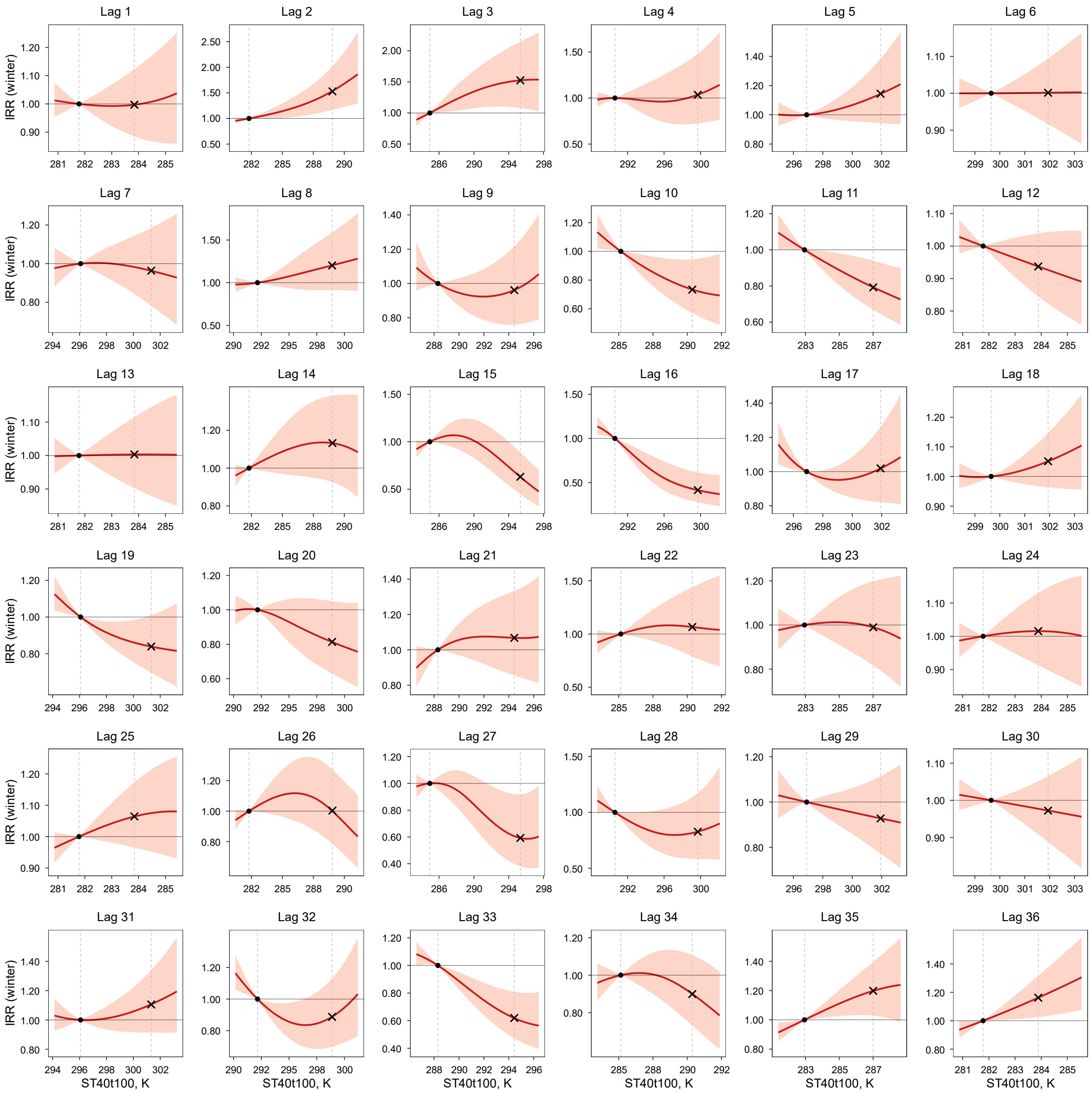

IRR (solid line) and 95% CI (shaded area) were estimated using a DLNM. Within each panel (lag month), IRR is expressed relative to the 25th percentile of the exposure variable in the study region (reference; IRR = 1), indicated by the left vertical dashed line and the dot on the IRR = 1 line. The right vertical dashed line indicates the 75th percentile, with the "×" symbol marking the estimated IRR at that level. Models were adjusted for spatiotemporal trends and surveillance-related changes in reporting and laboratory testing. Layer 5 soil moisture and soil temperature were modeled simultaneously in a single DLNM with season-specific effects.

**Figure S23.** Lag-specific exposure–response relationships between winter (January–March) coccidioidomycosis incidence and above-ground precipitation (Layer 2: meteorological layer) at lags 1–36 months.

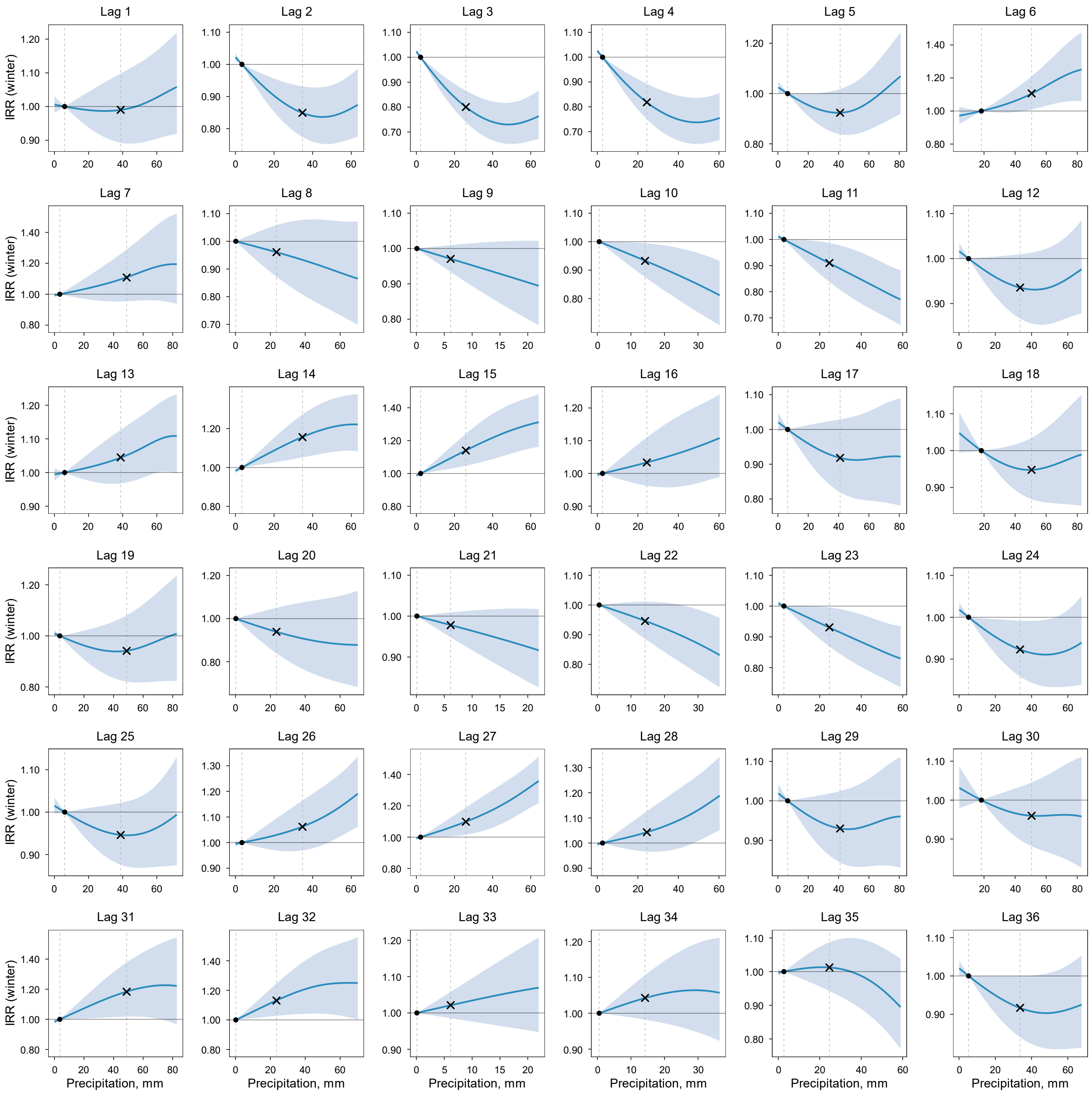

IRR (solid line) and 95% CI (shaded area) were estimated using a DLNM. Within each panel (lag month), IRR is expressed relative to the 25th percentile of the exposure variable in the study region (reference; IRR = 1), indicated by the left vertical dashed line and the dot on the IRR = 1 line. The right vertical dashed line indicates the 75th percentile, with the "×" symbol marking the estimated IRR at that level. Models were adjusted for spatiotemporal trends and surveillance-related changes in reporting and laboratory testing. Layer 2 meteorological conditions (precipitation and air temperature) were modeled simultaneously in a single DLNM with season-specific effects.

**Figure S24.** Lag-specific exposure–response relationships between winter (January–March) coccidioidomycosis incidence and above-ground air temperature (Layer 2: meteorological layer; AT) at lags 1–36 months.

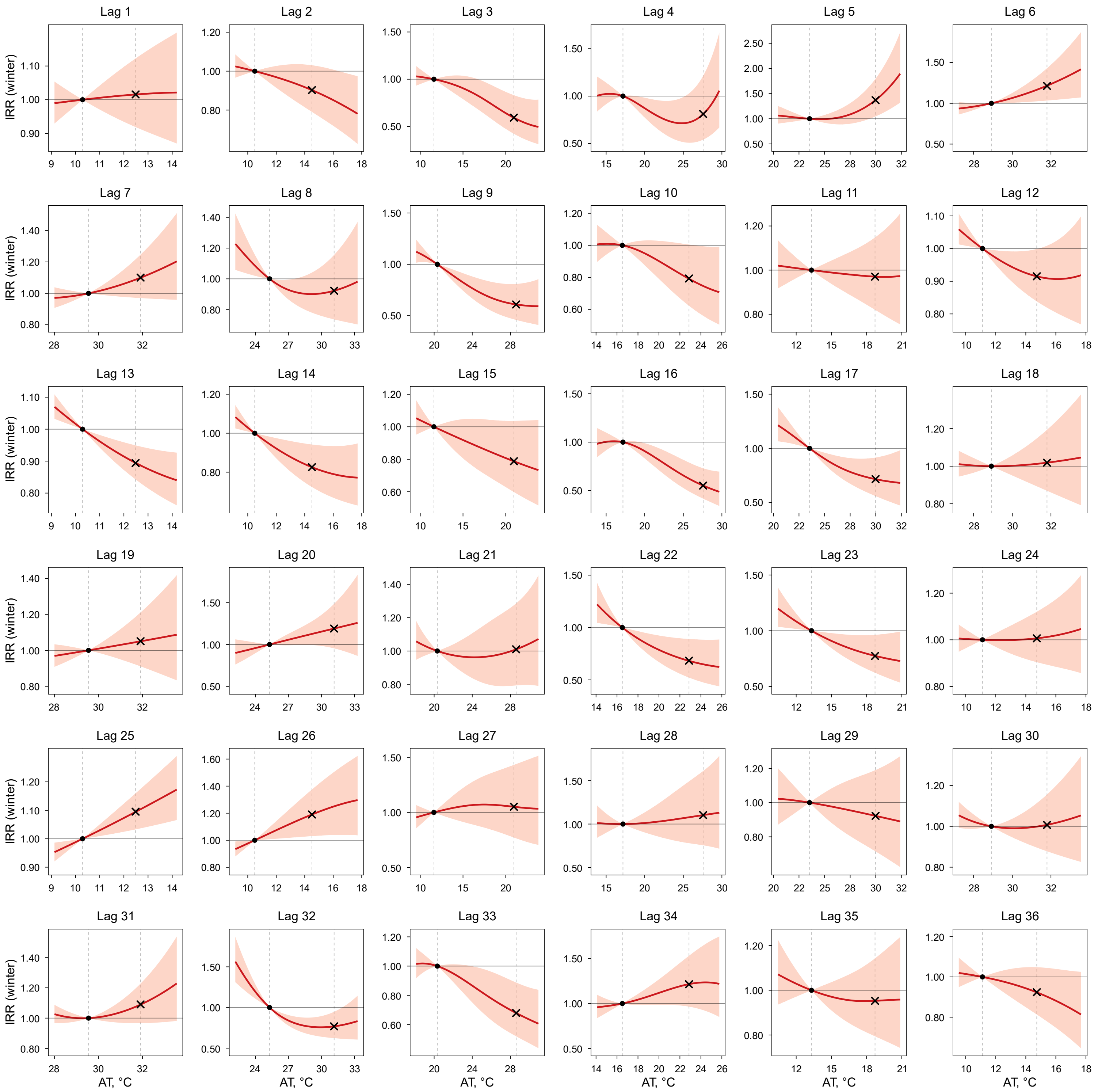

IRR (solid line) and 95% CI (shaded area) were estimated using a DLNM. Within each panel (lag month), IRR is expressed relative to the 25th percentile of the exposure variable in the study region (reference; IRR = 1), indicated by the left vertical dashed line and the dot on the IRR = 1 line. The right vertical dashed line indicates the 75th percentile, with the "×" symbol marking the estimated IRR at that level. Models were adjusted for spatiotemporal trends and surveillance-related changes in reporting and laboratory testing. Layer 2 meteorological conditions (precipitation and AT) were modeled simultaneously in a single DLNM with season-specific effects.

**Figure S25.** Lag-specific exposure–response relationships between winter (January–March) coccidioidomycosis incidence and above-ground $PM_{10}$ (Layer 1: dust dispersion layer) at lags 1–36 months.

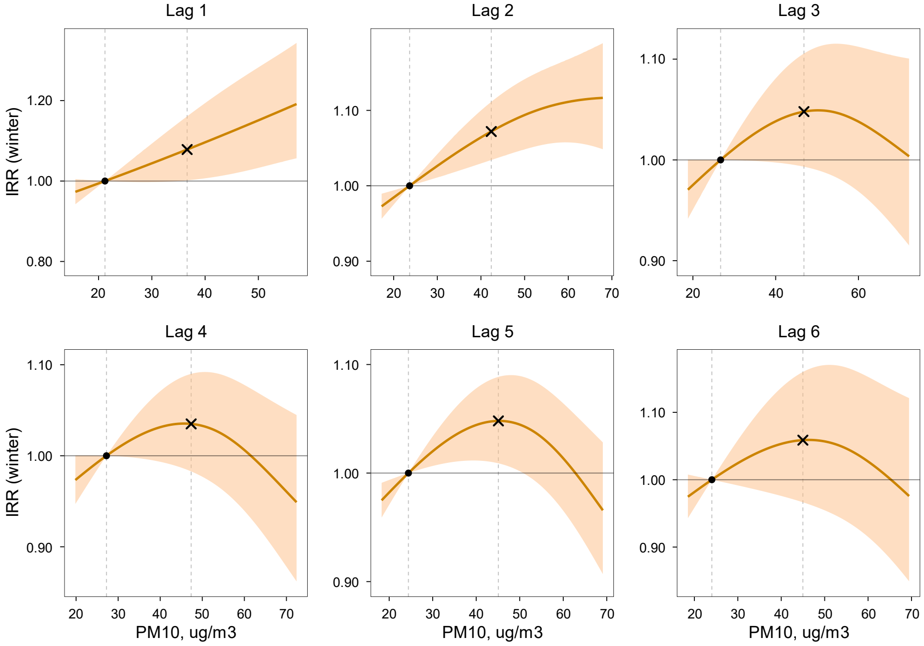

IRR (solid line) and 95% CI (shaded area) were estimated using a DLNM. Within each panel (lag month), IRR is expressed relative to the 25th percentile of the exposure variable in the study region (reference; IRR = 1), indicated by the left vertical dashed line and the dot on the IRR = 1 line. The right vertical dashed line indicates the 75th percentile, with the "×" symbol marking the estimated IRR at that level. Models were adjusted for spatiotemporal trends and surveillance-related changes in reporting and laboratory testing. Layer 1 dust dispersion conditions ($PM_{10}$ and wind speed) were modeled simultaneously in a single DLNM with season-specific effects.

**Figure S26.** Lag-specific exposure–response relationships between winter (January–March) coccidioidomycosis incidence and above-ground wind speed (Layer 1: dust dispersion layer) at lags 1–36 months.

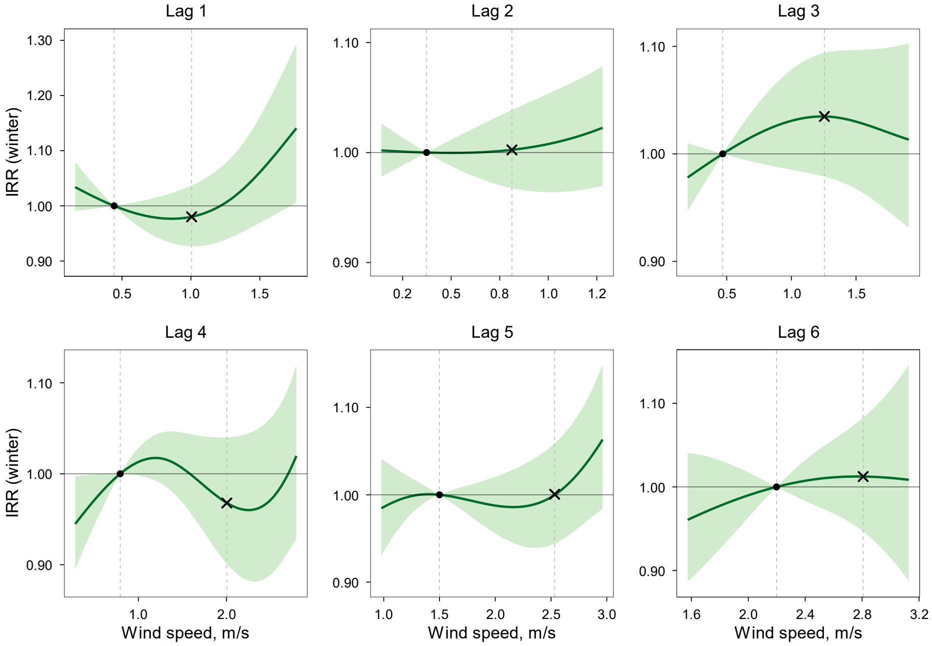

IRR (solid line) and 95% CI (shaded area) were estimated using a DLNM. Within each panel (lag month), IRR is expressed relative to the 25th percentile of the exposure variable in the study region (reference; IRR = 1), indicated by the left vertical dashed line and the dot on the IRR = 1 line. The right vertical dashed line indicates the 75th percentile, with the "×" symbol marking the estimated IRR at that level. Models were adjusted for spatiotemporal trends and surveillance-related changes in reporting and laboratory testing. Layer 1 dust dispersion conditions ($PM_{10}$ and wind speed) were modeled simultaneously in a single DLNM with season-specific effects.

**Figure S27.** Lag-specific exposure–response relationships between spring (April–June) coccidioidomycosis incidence and below-ground soil moisture (Layer 3: 0–10 cm; topsoil; SM0t10) at lags 1–36 months.

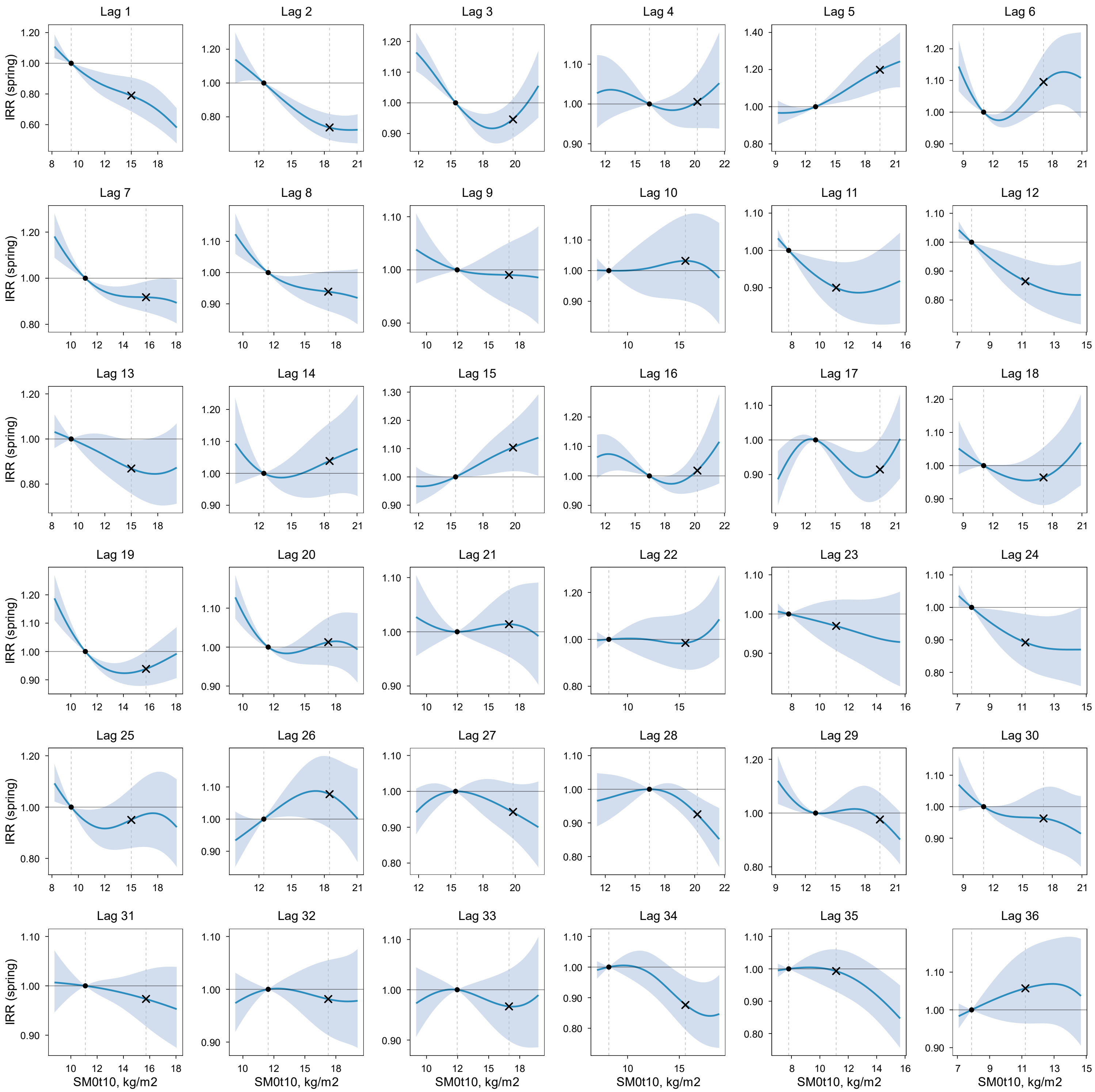

IRR (solid line) and 95% CI (shaded area) were estimated using a DLNM. Within each panel (lag month), IRR is expressed relative to the 25th percentile of the exposure variable in the study region (reference; IRR = 1), indicated by the left vertical dashed line and the dot on the IRR = 1 line. The right vertical dashed line indicates the 75th percentile, with the "×" symbol marking the estimated IRR at that level. Models were adjusted for spatiotemporal trends and surveillance-related changes in reporting and laboratory testing. Layer 3 soil moisture and soil temperature were modeled simultaneously in a single DLNM with season-specific effects.

**Figure S28.** Lag-specific exposure–response relationships between spring (April–June) coccidioidomycosis incidence and below-ground soil temperature (Layer 3: 0–10 cm; topsoil; ST0t10) at lags 1–36 months.

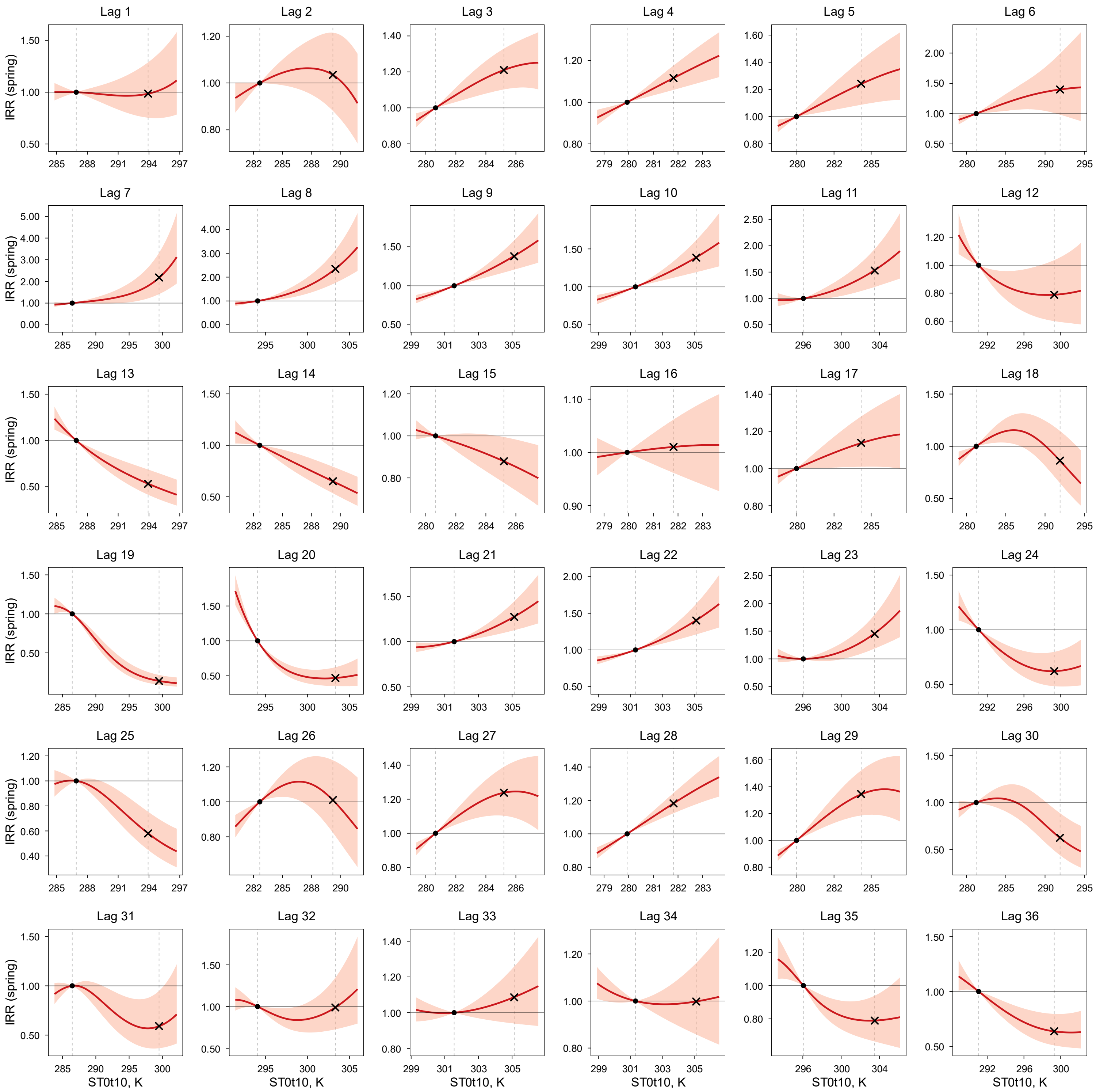

IRR (solid line) and 95% CI (shaded area) were estimated using a DLNM. Within each panel (lag month), IRR is expressed relative to the 25th percentile of the exposure variable in the study region (reference; IRR = 1), indicated by the left vertical dashed line and the dot on the IRR = 1 line. The right vertical dashed line indicates the 75th percentile, with the "×" symbol marking the estimated IRR at that level. Models were adjusted for spatiotemporal trends and surveillance-related changes in reporting and laboratory testing. Layer 3 soil moisture and soil temperature were modeled simultaneously in a single DLNM with season-specific effects.

**Figure S29.** Lag-specific exposure–response relationships between spring (April–June) coccidioidomycosis incidence and below-ground soil moisture (Layer 4: 10–40 cm; SM10t40) at lags 1–36 months.

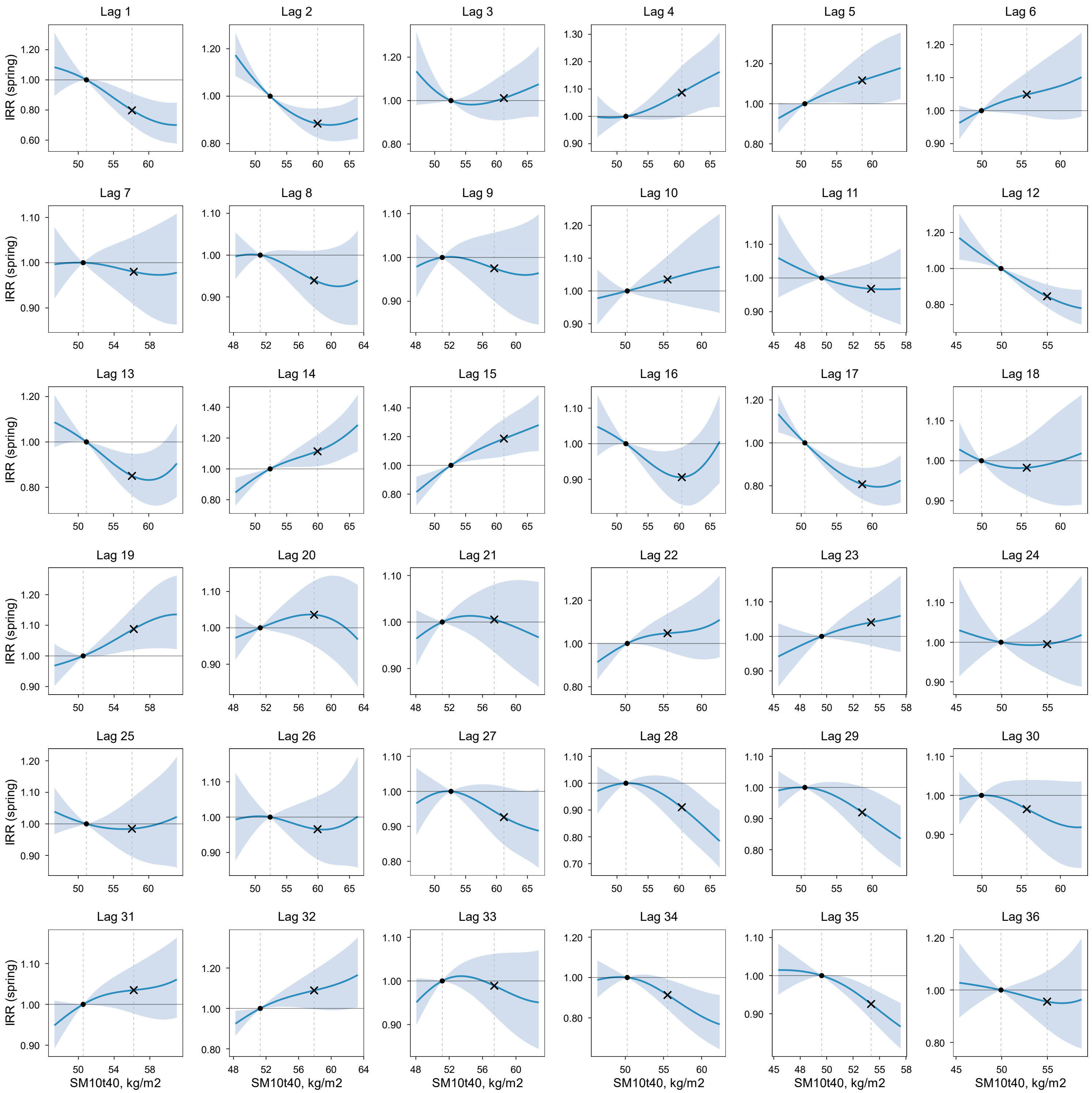

IRR (solid line) and 95% CI (shaded area) were estimated using a DLNM. Within each panel (lag month), IRR is expressed relative to the 25th percentile of the exposure variable in the study region (reference; IRR = 1), indicated by the left vertical dashed line and the dot on the IRR = 1 line. The right vertical dashed line indicates the 75th percentile, with the "×" symbol marking the estimated IRR at that level. Models were adjusted for spatiotemporal trends and surveillance-related changes in reporting and laboratory testing. Layer 4 soil moisture and soil temperature were modeled simultaneously in a single DLNM with season-specific effects.

**Figure S30.** Lag-specific exposure–response relationships between spring (April–June) coccidioidomycosis incidence and below-ground soil temperature (Layer 4: 10–40 cm; ST10t40) at lags 1–36 months.

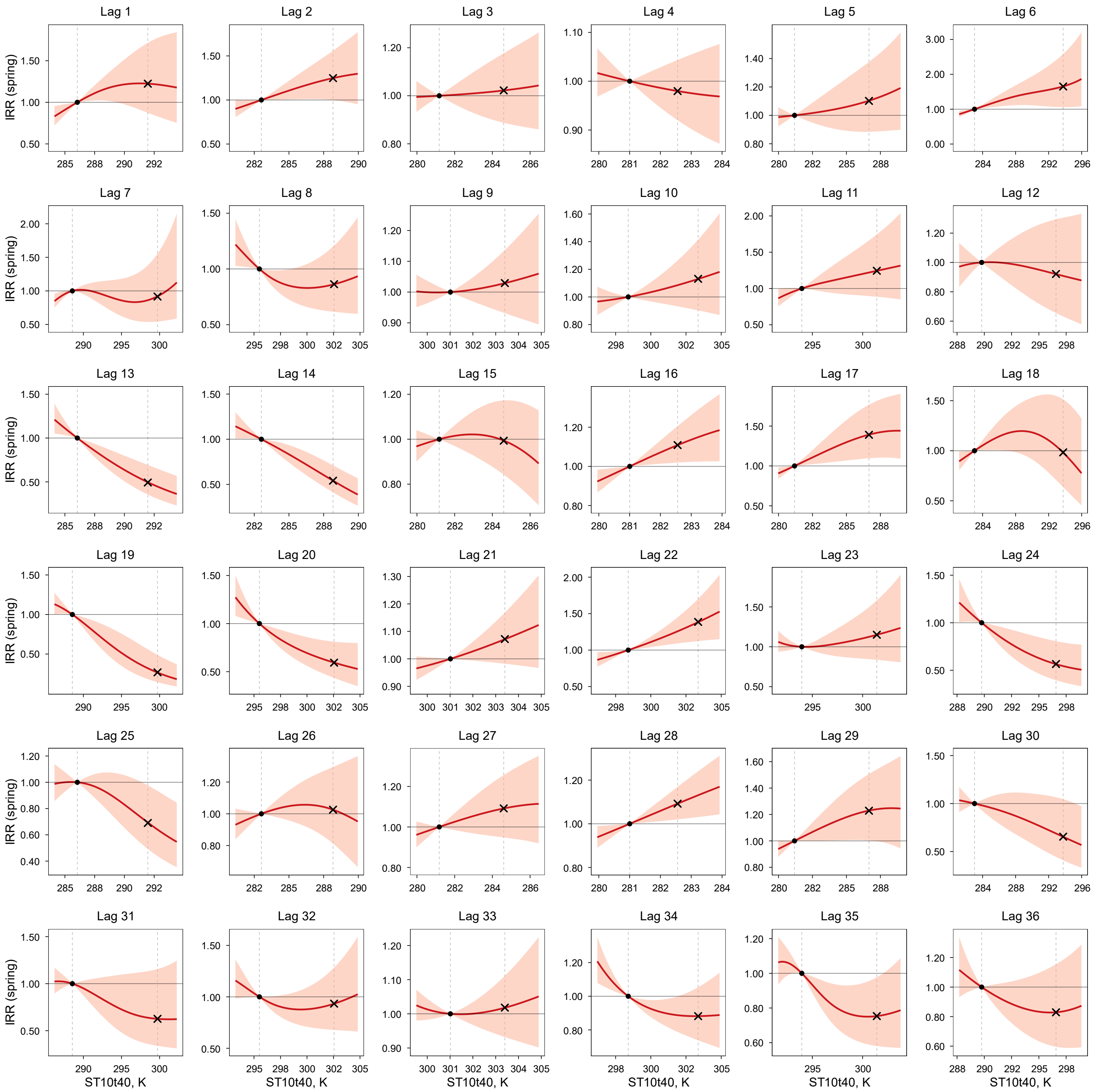

IRR (solid line) and 95% CI (shaded area) were estimated using a DLNM. Within each panel (lag month), IRR is expressed relative to the 25th percentile of the exposure variable in the study region (reference; IRR = 1), indicated by the left vertical dashed line and the dot on the IRR = 1 line. The right vertical dashed line indicates the 75th percentile, with the "×" symbol marking the estimated IRR at that level. Models were adjusted for spatiotemporal trends and surveillance-related changes in reporting and laboratory testing. Layer 4 soil moisture and soil temperature were modeled simultaneously in a single DLNM with season-specific effects.

**Figure S31.** Lag-specific exposure–response relationships between spring (April–June) coccidioidomycosis incidence and below-ground soil moisture (Layer 5: 40–100 cm; SM40t100) at lags 1–36 months.

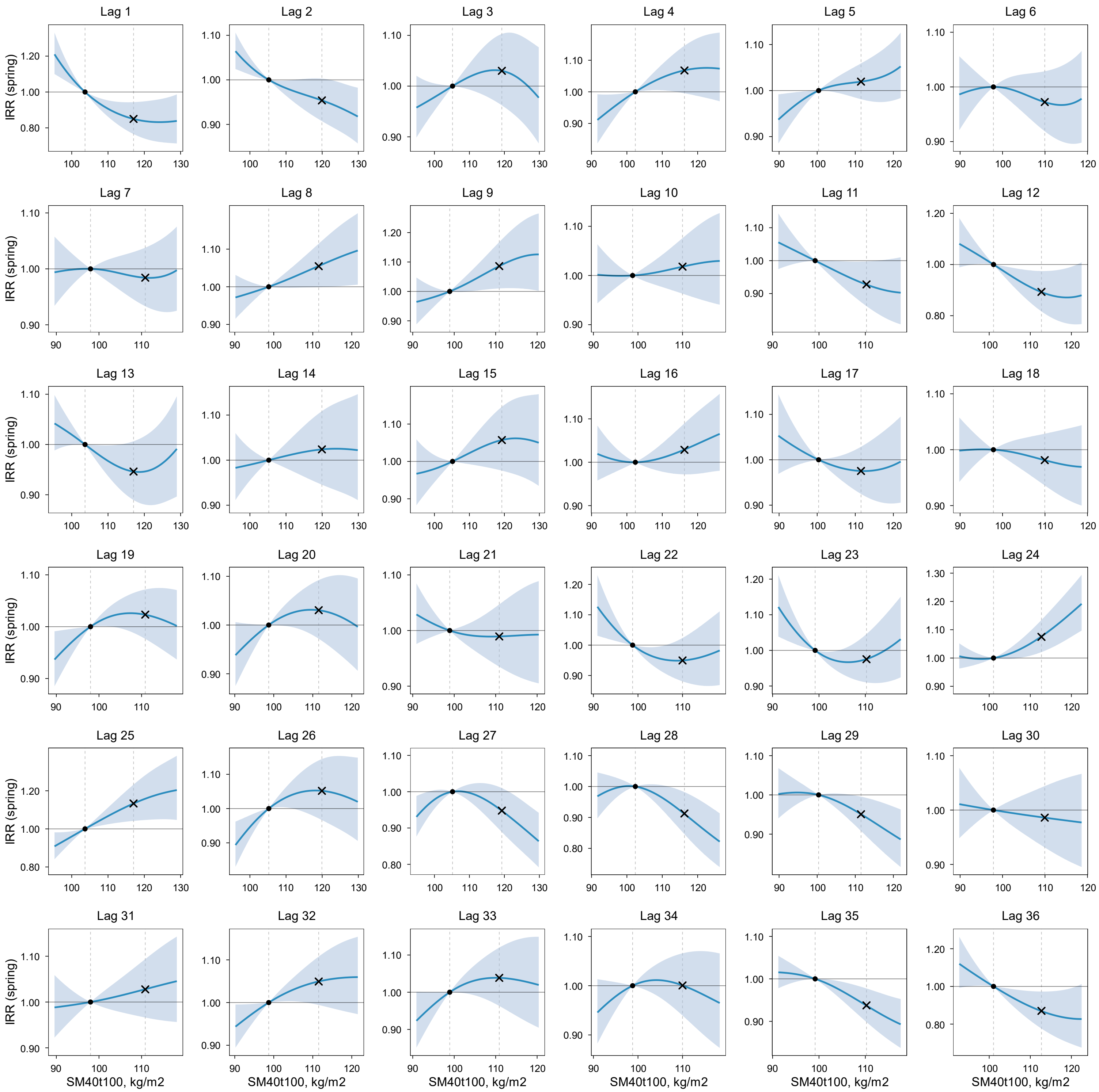

IRR (solid line) and 95% CI (shaded area) were estimated using a DLNM. Within each panel (lag month), IRR is expressed relative to the 25th percentile of the exposure variable in the study region (reference; IRR = 1), indicated by the left vertical dashed line and the dot on the IRR = 1 line. The right vertical dashed line indicates the 75th percentile, with the "×" symbol marking the estimated IRR at that level. Models were adjusted for spatiotemporal trends and surveillance-related changes in reporting and laboratory testing. Layer 5 soil moisture and soil temperature were modeled simultaneously in a single DLNM with season-specific effects.

**Figure S32.** Lag-specific exposure–response relationships between spring (April–June) coccidioidomycosis incidence and below-ground soil temperature (Layer 5: 40–100 cm; ST40t100) at lags 1–36 months.

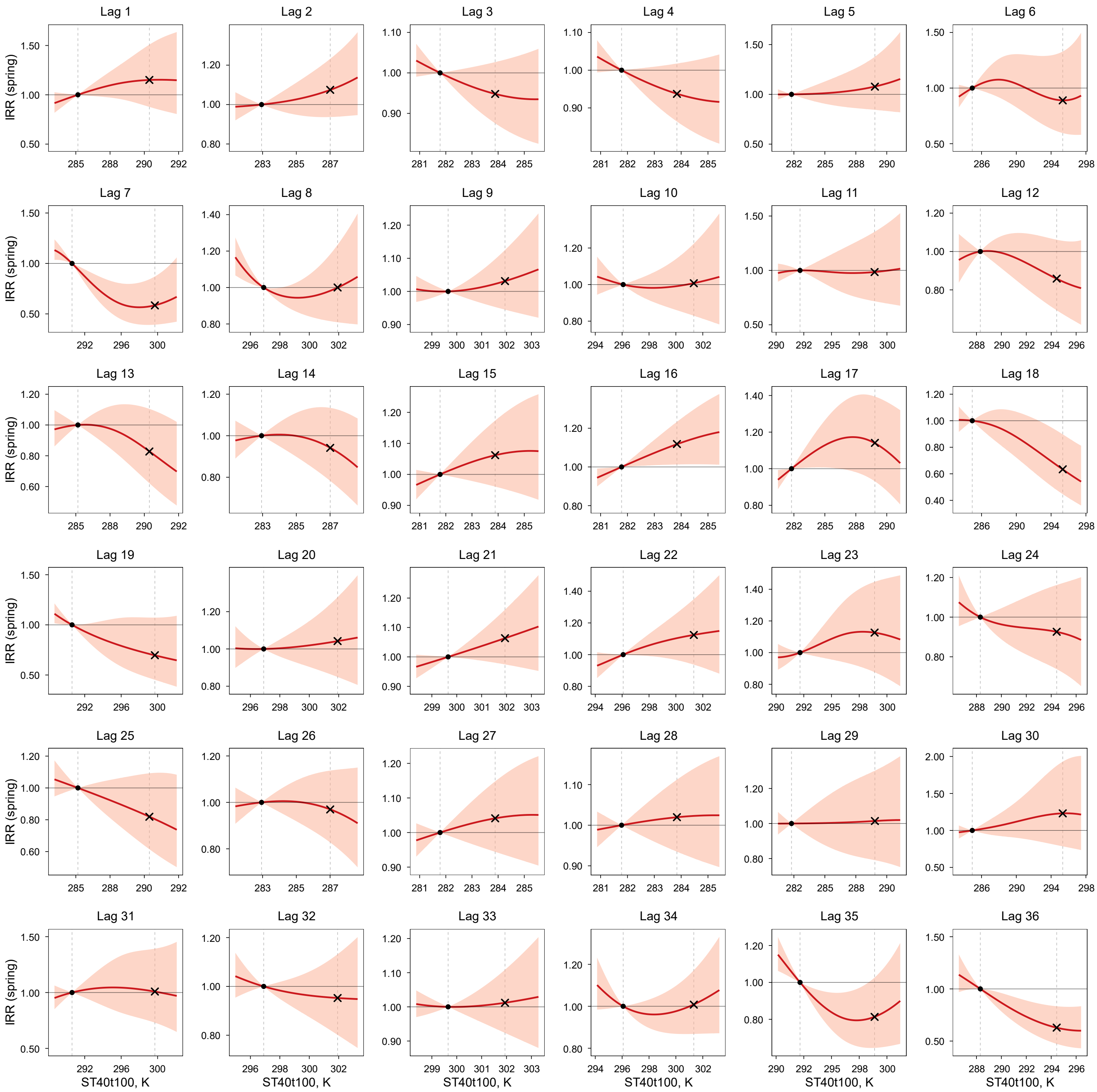

IRR (solid line) and 95% CI (shaded area) were estimated using a DLNM. Within each panel (lag month), IRR is expressed relative to the 25th percentile of the exposure variable in the study region (reference; IRR = 1), indicated by the left vertical dashed line and the dot on the IRR = 1 line. The right vertical dashed line indicates the 75th percentile, with the "×" symbol marking the estimated IRR at that level. Models were adjusted for spatiotemporal trends and surveillance-related changes in reporting and laboratory testing. Layer 5 soil moisture and soil temperature were modeled simultaneously in a single DLNM with season-specific effects.

**Figure S33.** Lag-specific exposure–response relationships between spring (April–June) coccidioidomycosis incidence and above-ground precipitation (Layer 2: meteorological layer) at lags 1–36 months.

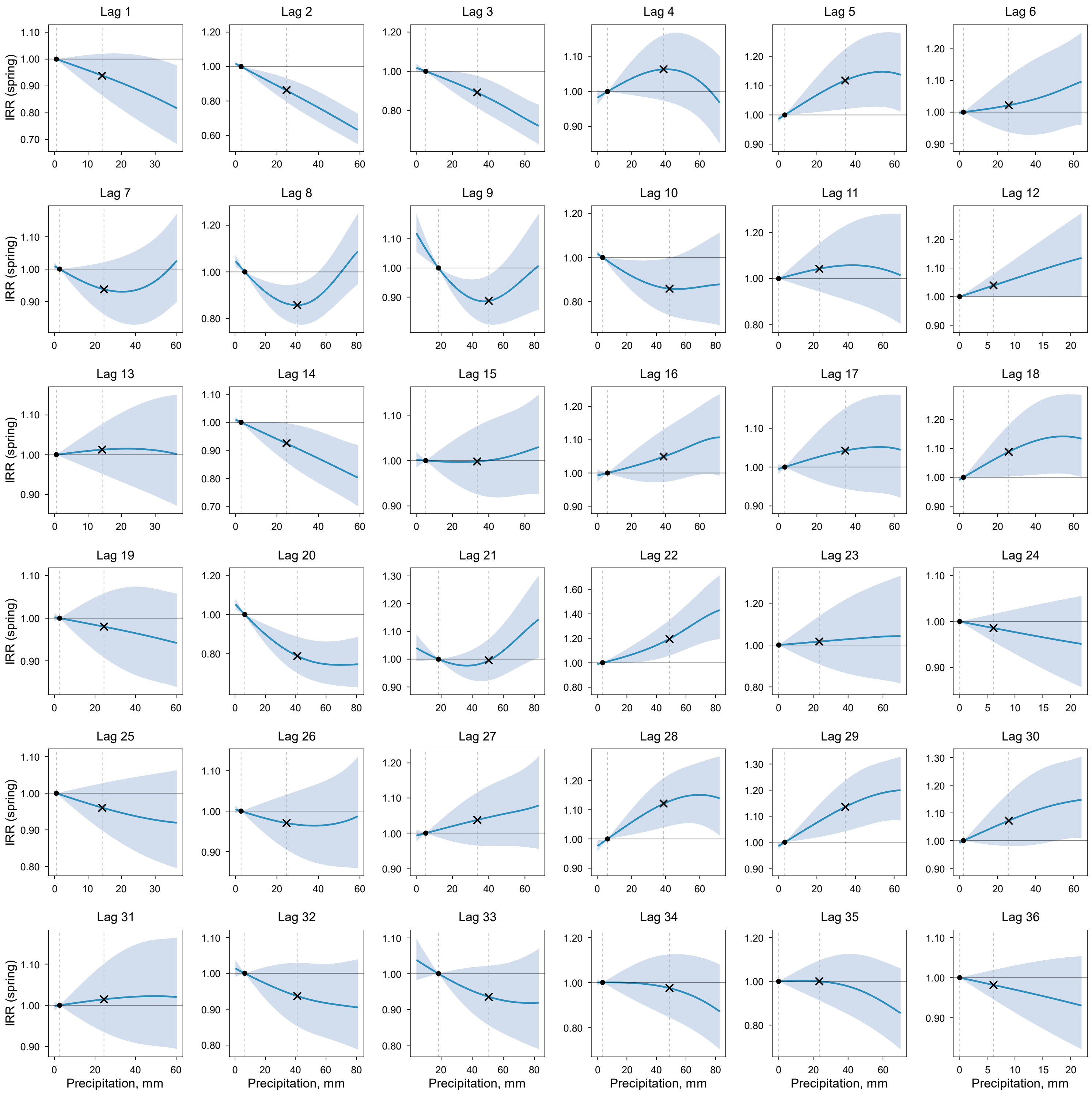

IRR (solid line) and 95% CI (shaded area) were estimated using a DLNM. Within each panel (lag month), IRR is expressed relative to the 25th percentile of the exposure variable in the study region (reference; IRR = 1), indicated by the left vertical dashed line and the dot on the IRR = 1 line. The right vertical dashed line indicates the 75th percentile, with the "×" symbol marking the estimated IRR at that level. Models were adjusted for spatiotemporal trends and surveillance-related changes in reporting and laboratory testing. Layer 2 meteorological conditions (precipitation and air temperature) were modeled simultaneously in a single DLNM with season-specific effects.

**Figure S34.** Lag-specific exposure–response relationships between spring (April–June) coccidioidomycosis incidence and above-ground air temperature (Layer 2: meteorological layer; AT) at lags 1–36 months.

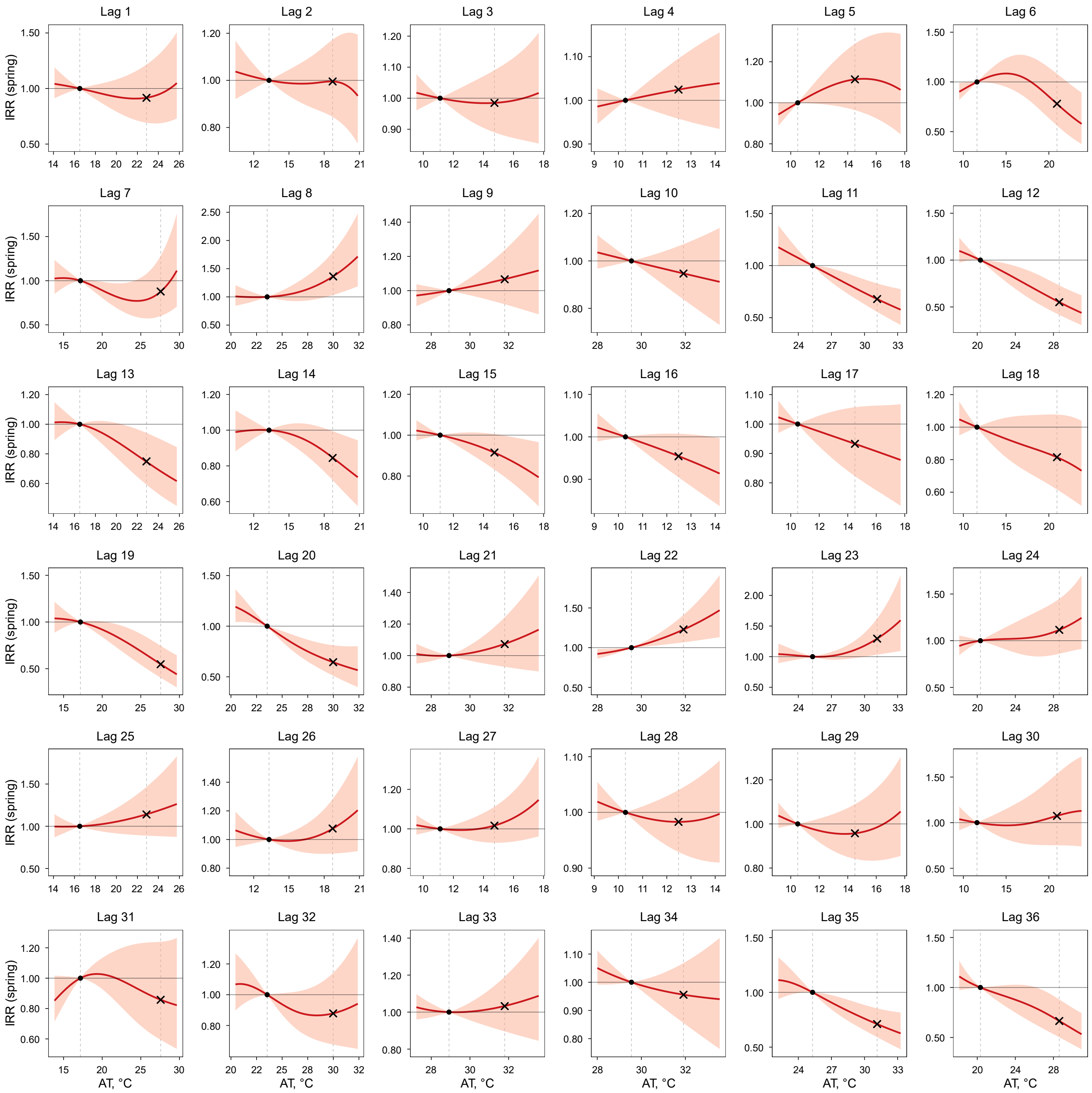

IRR (solid line) and 95% CI (shaded area) were estimated using a DLNM. Within each panel (lag month), IRR is expressed relative to the 25th percentile of the exposure variable in the study region (reference; IRR = 1), indicated by the left vertical dashed line and the dot on the IRR = 1 line. The right vertical dashed line indicates the 75th percentile, with the "×" symbol marking the estimated IRR at that level. Models were adjusted for spatiotemporal trends and surveillance-related changes in reporting and laboratory testing. Layer 2 meteorological conditions (precipitation and AT) were modeled simultaneously in a single DLNM with season-specific effects.

**Figure S35.** Lag-specific exposure–response relationships between spring (April–June) coccidioidomycosis incidence and above-ground $PM_{10}$ (Layer 1: dust dispersion layer) at lags 1–36 months.

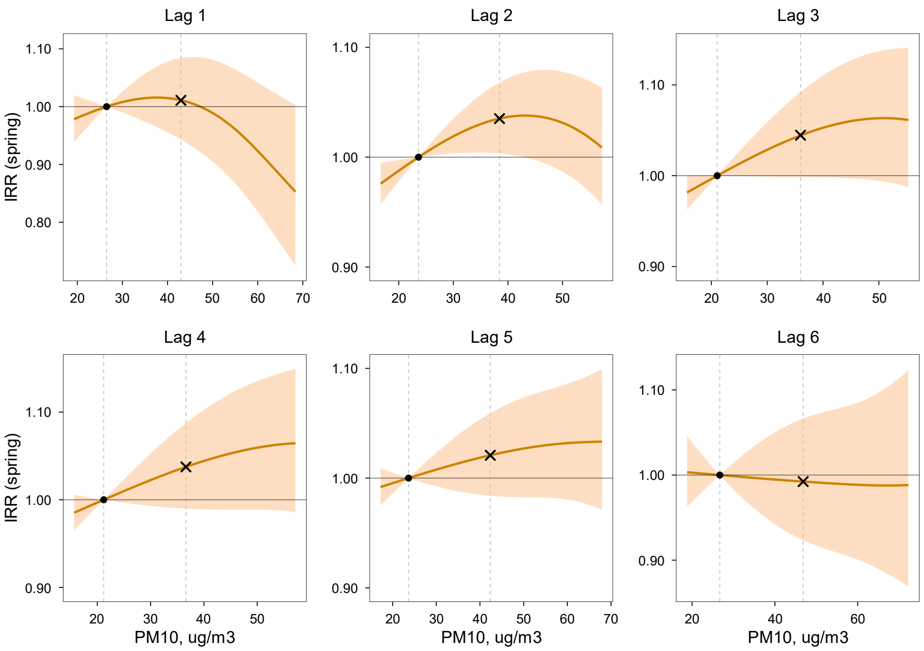

IRR (solid line) and 95% CI (shaded area) were estimated using a DLNM. Within each panel (lag month), IRR is expressed relative to the 25th percentile of the exposure variable in the study region (reference; IRR = 1), indicated by the left vertical dashed line and the dot on the IRR = 1 line. The right vertical dashed line indicates the 75th percentile, with the "×" symbol marking the estimated IRR at that level. Models were adjusted for spatiotemporal trends and surveillance-related changes in reporting and laboratory testing. Layer 1 dust dispersion conditions ($PM_{10}$ and wind speed) were modeled simultaneously in a single DLNM with season-specific effects.

**Figure S36.** Lag-specific exposure–response relationships between spring (April–June) coccidioidomycosis incidence and above-ground wind speed (Layer 1: dust dispersion layer) at lags 1–36 months.

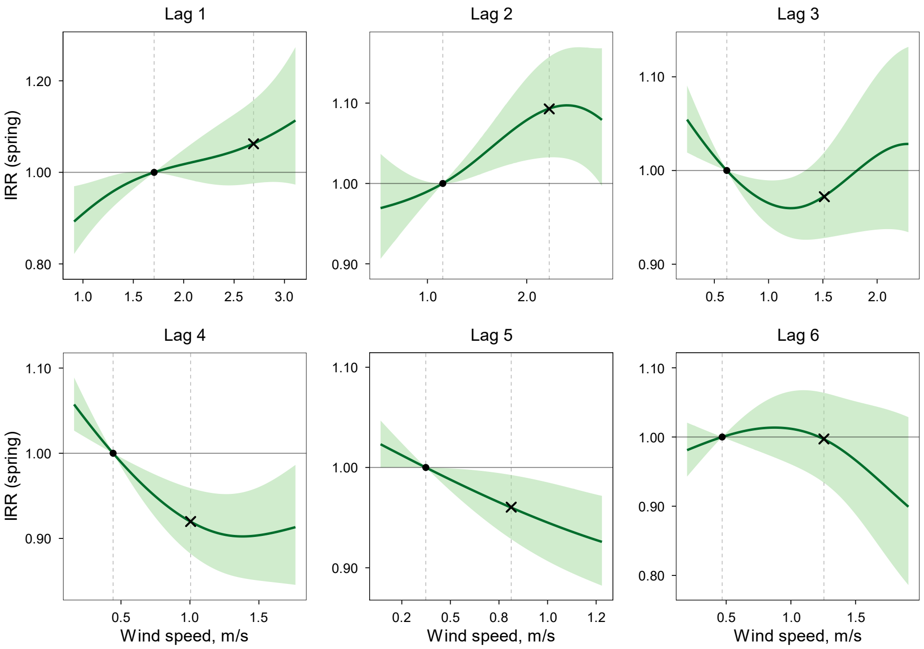

IRR (solid line) and 95% CI (shaded area) were estimated using a DLNM. Within each panel (lag month), IRR is expressed relative to the 25th percentile of the exposure variable in the study region (reference; IRR = 1), indicated by the left vertical dashed line and the dot on the IRR = 1 line. The right vertical dashed line indicates the 75th percentile, with the "×" symbol marking the estimated IRR at that level. Models were adjusted for spatiotemporal trends and surveillance-related changes in reporting and laboratory testing. Layer 1 dust dispersion conditions ($PM_{10}$ and wind speed) were modeled simultaneously in a single DLNM with season-specific effects.

**Figure S37.** Lag-specific exposure–response relationships between monsoon (July–September) coccidioidomycosis incidence and below-ground soil moisture (Layer 3: 0–10 cm; topsoil; SM0t10) at lags 1–36 months.

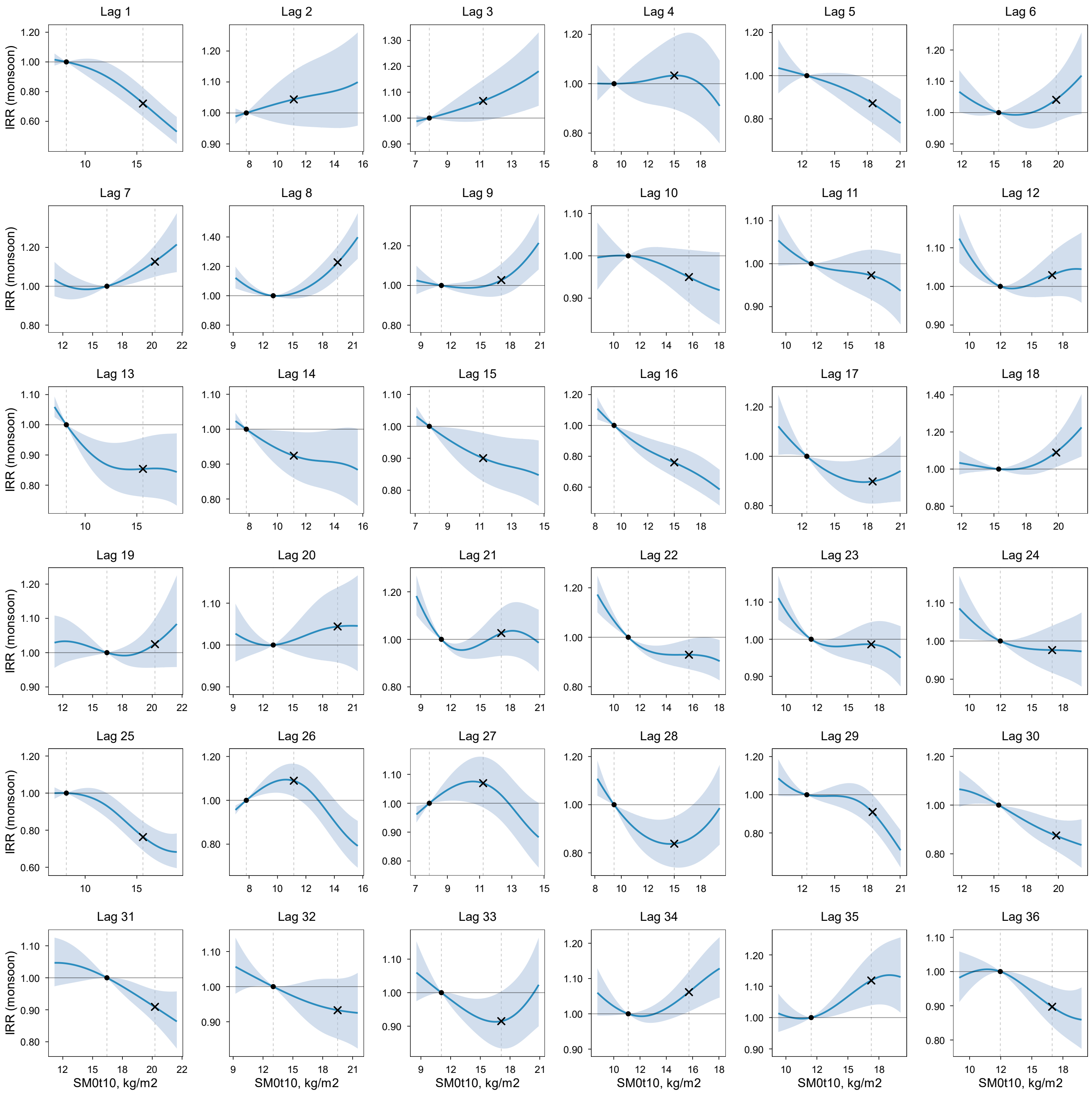

IRR (solid line) and 95% CI (shaded area) were estimated using a DLNM. Within each panel (lag month), IRR is expressed relative to the 25th percentile of the exposure variable in the study region (reference; IRR = 1), indicated by the left vertical dashed line and the dot on the IRR = 1 line. The right vertical dashed line indicates the 75th percentile, with the "×" symbol marking the estimated IRR at that level. Models were adjusted for spatiotemporal trends and surveillance-related changes in reporting and laboratory testing. Layer 3 soil moisture and soil temperature were modeled simultaneously in a single DLNM with season-specific effects.

**Figure S38.** Lag-specific exposure–response relationships between monsoon (July–September) coccidioidomycosis incidence and below-ground soil temperature (Layer 3: 0–10 cm; topsoil; ST0t10) at lags 1–36 months.

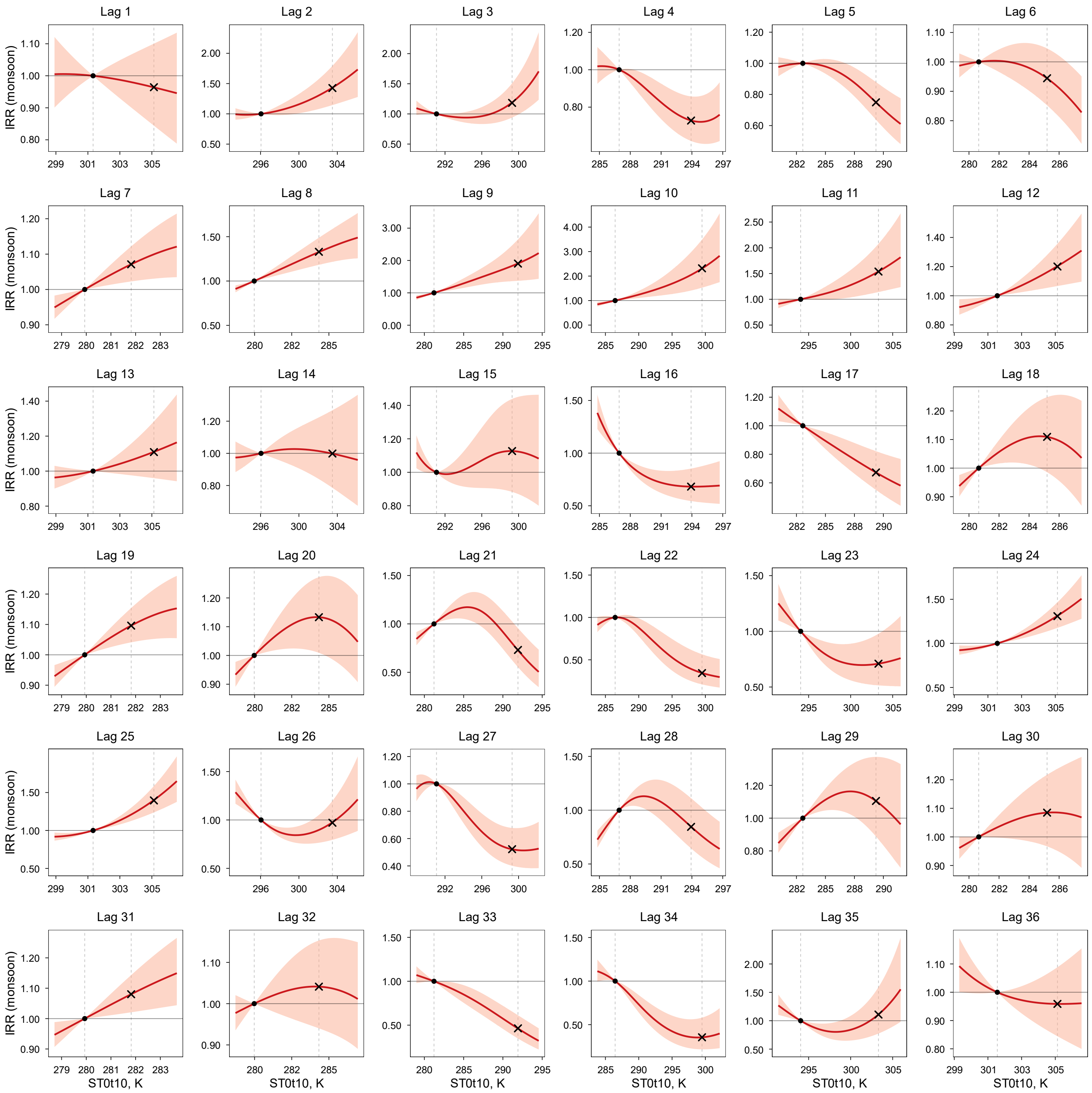

IRR (solid line) and 95% CI (shaded area) were estimated using a DLNM. Within each panel (lag month), IRR is expressed relative to the 25th percentile of the exposure variable in the study region (reference; IRR = 1), indicated by the left vertical dashed line and the dot on the IRR = 1 line. The right vertical dashed line indicates the 75th percentile, with the "×" symbol marking the estimated IRR at that level. Models were adjusted for spatiotemporal trends and surveillance-related changes in reporting and laboratory testing. Layer 3 soil moisture and soil temperature were modeled simultaneously in a single DLNM with season-specific effects.

**Figure S39.** Lag-specific exposure–response relationships between monsoon (July–September) coccidioidomycosis incidence and below-ground soil moisture (Layer 4: 10–40 cm; SM10t40) at lags 1–36 months.

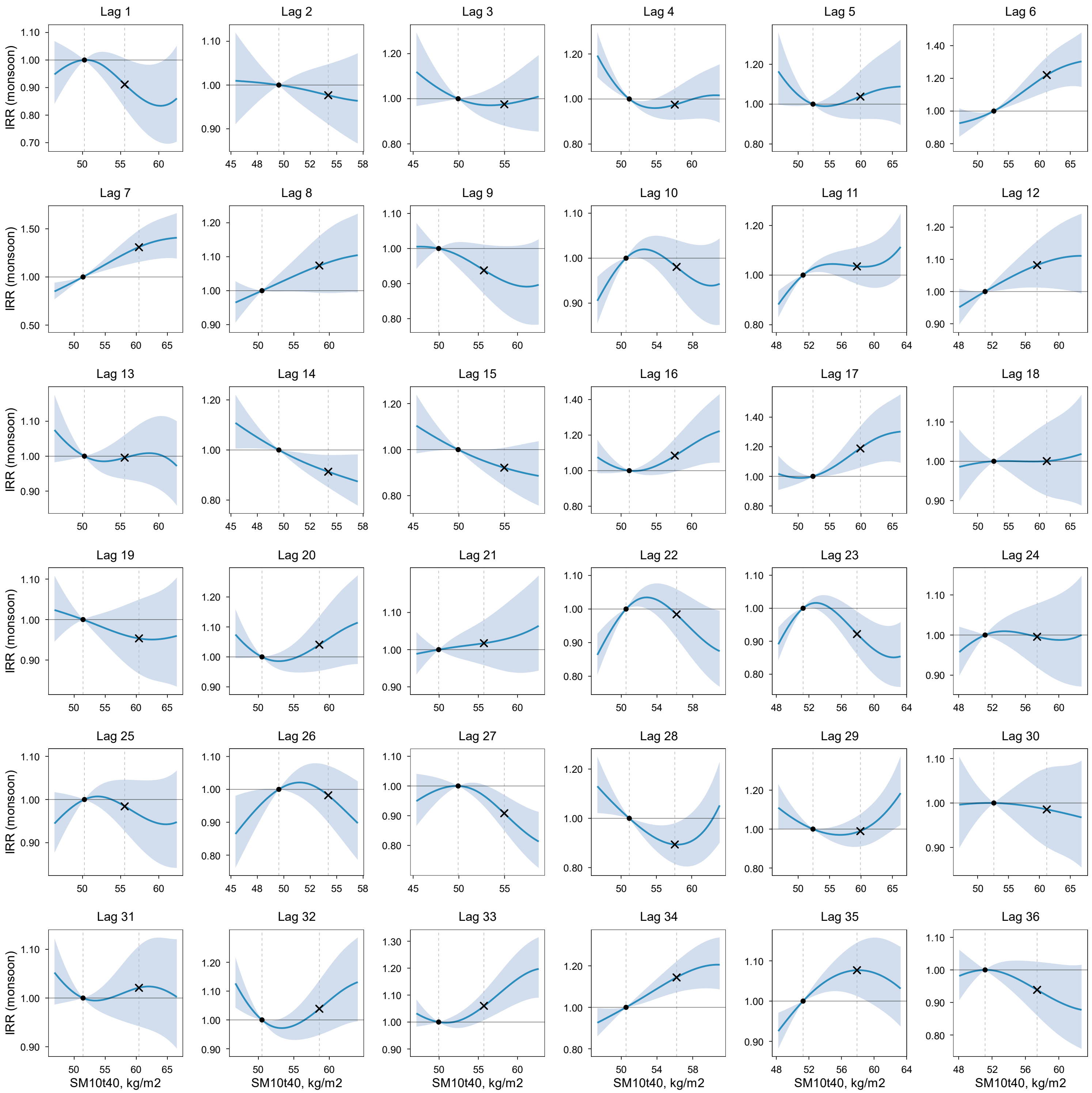

IRR (solid line) and 95% CI (shaded area) were estimated using a DLNM. Within each panel (lag month), IRR is expressed relative to the 25th percentile of the exposure variable in the study region (reference; IRR = 1), indicated by the left vertical dashed line and the dot on the IRR = 1 line. The right vertical dashed line indicates the 75th percentile, with the "×" symbol marking the estimated IRR at that level. Models were adjusted for spatiotemporal trends and surveillance-related changes in reporting and laboratory testing. Layer 4 soil moisture and soil temperature were modeled simultaneously in a single DLNM with season-specific effects.

**Figure S40.** Lag-specific exposure–response relationships between monsoon (July–September) coccidioidomycosis incidence and below-ground soil temperature (Layer 4: 10–40 cm; ST10t40) at lags 1–36 months.

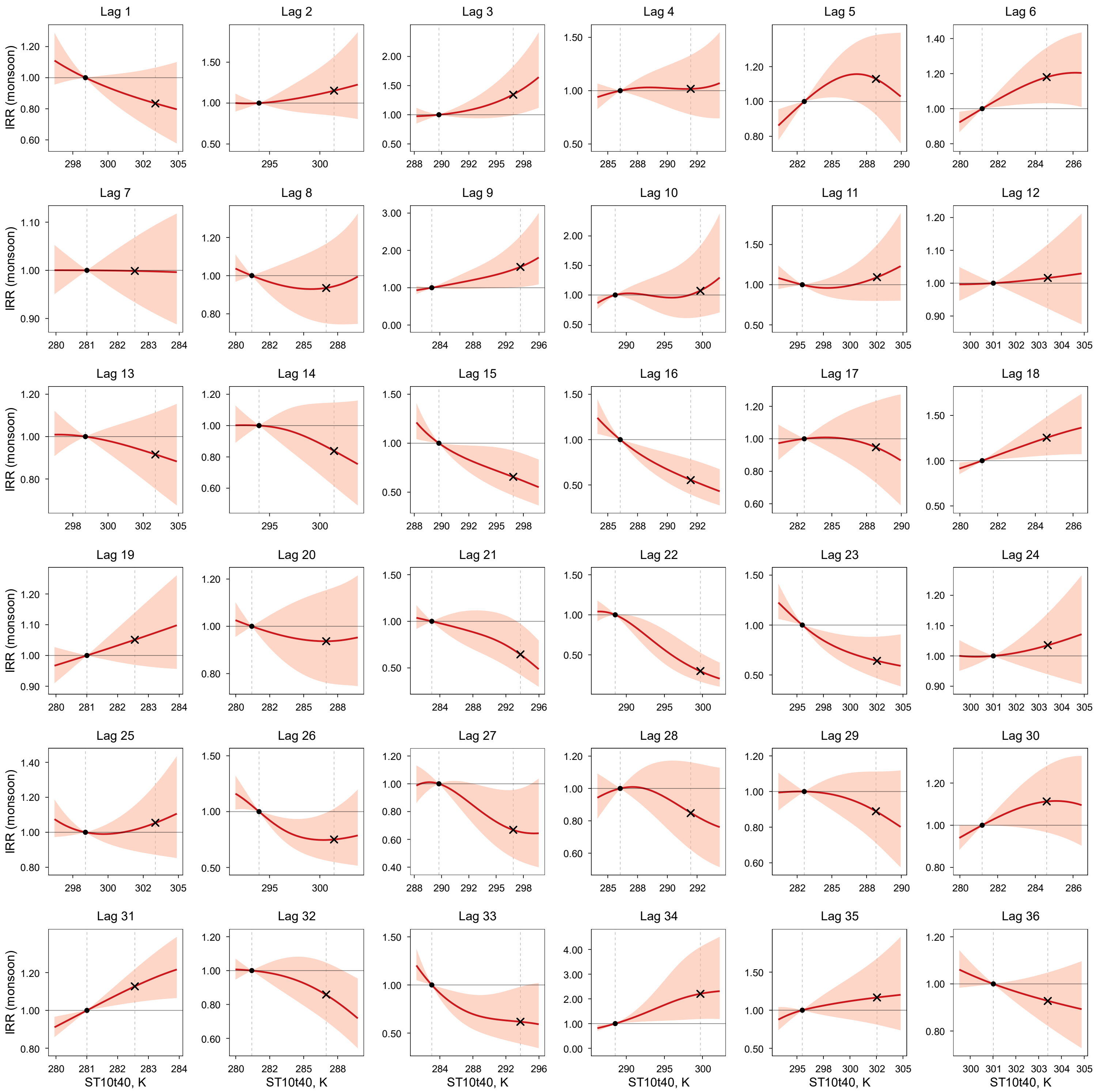

IRR (solid line) and 95% CI (shaded area) were estimated using a DLNM. Within each panel (lag month), IRR is expressed relative to the 25th percentile of the exposure variable in the study region (reference; IRR = 1), indicated by the left vertical dashed line and the dot on the IRR = 1 line. The right vertical dashed line indicates the 75th percentile, with the "×" symbol marking the estimated IRR at that level. Models were adjusted for spatiotemporal trends and surveillance-related changes in reporting and laboratory testing. Layer 4 soil moisture and soil temperature were modeled simultaneously in a single DLNM with season-specific effects.

**Figure S41.** Lag-specific exposure–response relationships between monsoon (July–September) coccidioidomycosis incidence and below-ground soil moisture (Layer 5: 40–100 cm; SM40t100) at lags 1–36 months.

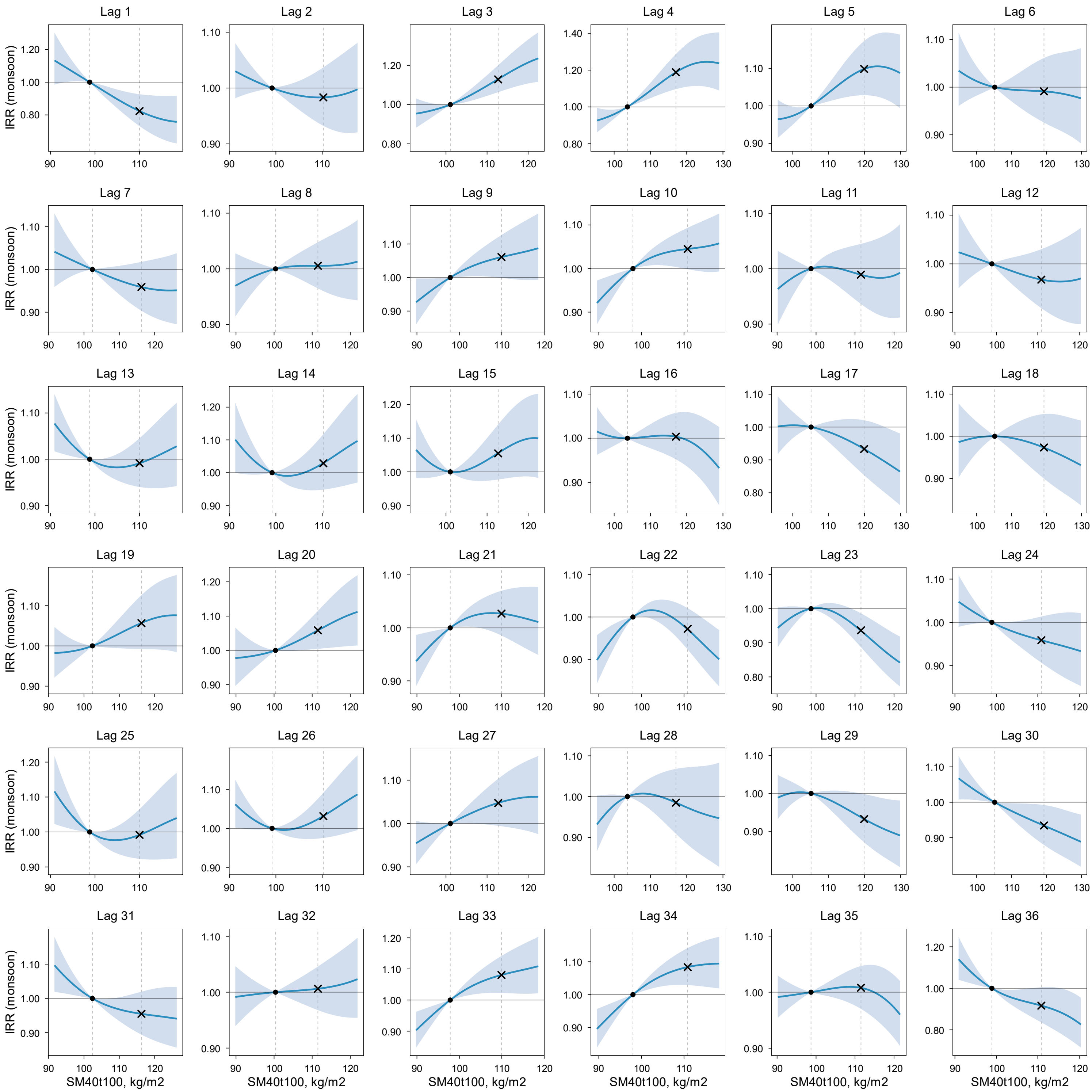

IRR (solid line) and 95% CI (shaded area) were estimated using a DLNM. Within each panel (lag month), IRR is expressed relative to the 25th percentile of the exposure variable in the study region (reference; IRR = 1), indicated by the left vertical dashed line and the dot on the IRR = 1 line. The right vertical dashed line indicates the 75th percentile, with the "×" symbol marking the estimated IRR at that level. Models were adjusted for spatiotemporal trends and surveillance-related changes in reporting and laboratory testing. Layer 5 soil moisture and soil temperature were modeled simultaneously in a single DLNM with season-specific effects.

**Figure S42.** Lag-specific exposure–response relationships between monsoon (July–September) coccidioidomycosis incidence and below-ground soil temperature (Layer 5: 40–100 cm; ST40t100) at lags 1–36 months.

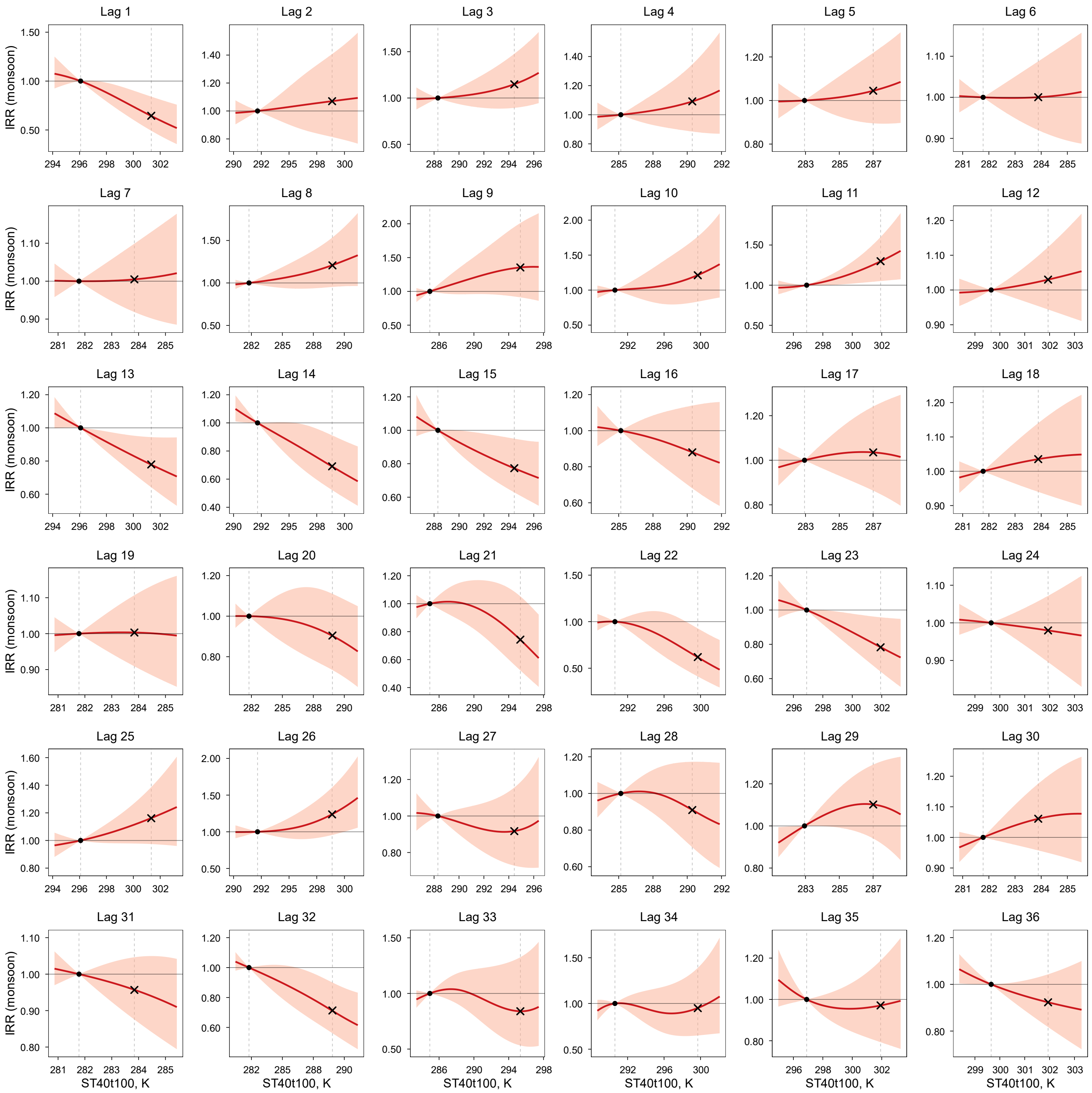

IRR (solid line) and 95% CI (shaded area) were estimated using a DLNM. Within each panel (lag month), IRR is expressed relative to the 25th percentile of the exposure variable in the study region (reference; IRR = 1), indicated by the left vertical dashed line and the dot on the IRR = 1 line. The right vertical dashed line indicates the 75th percentile, with the "×" symbol marking the estimated IRR at that level. Models were adjusted for spatiotemporal trends and surveillance-related changes in reporting and laboratory testing. Layer 5 soil moisture and soil temperature were modeled simultaneously in a single DLNM with season-specific effects.

**Figure S43.** Lag-specific exposure–response relationships between monsoon (July–September) coccidioidomycosis incidence and above-ground precipitation (Layer 2: meteorological layer) at lags 1–36 months.

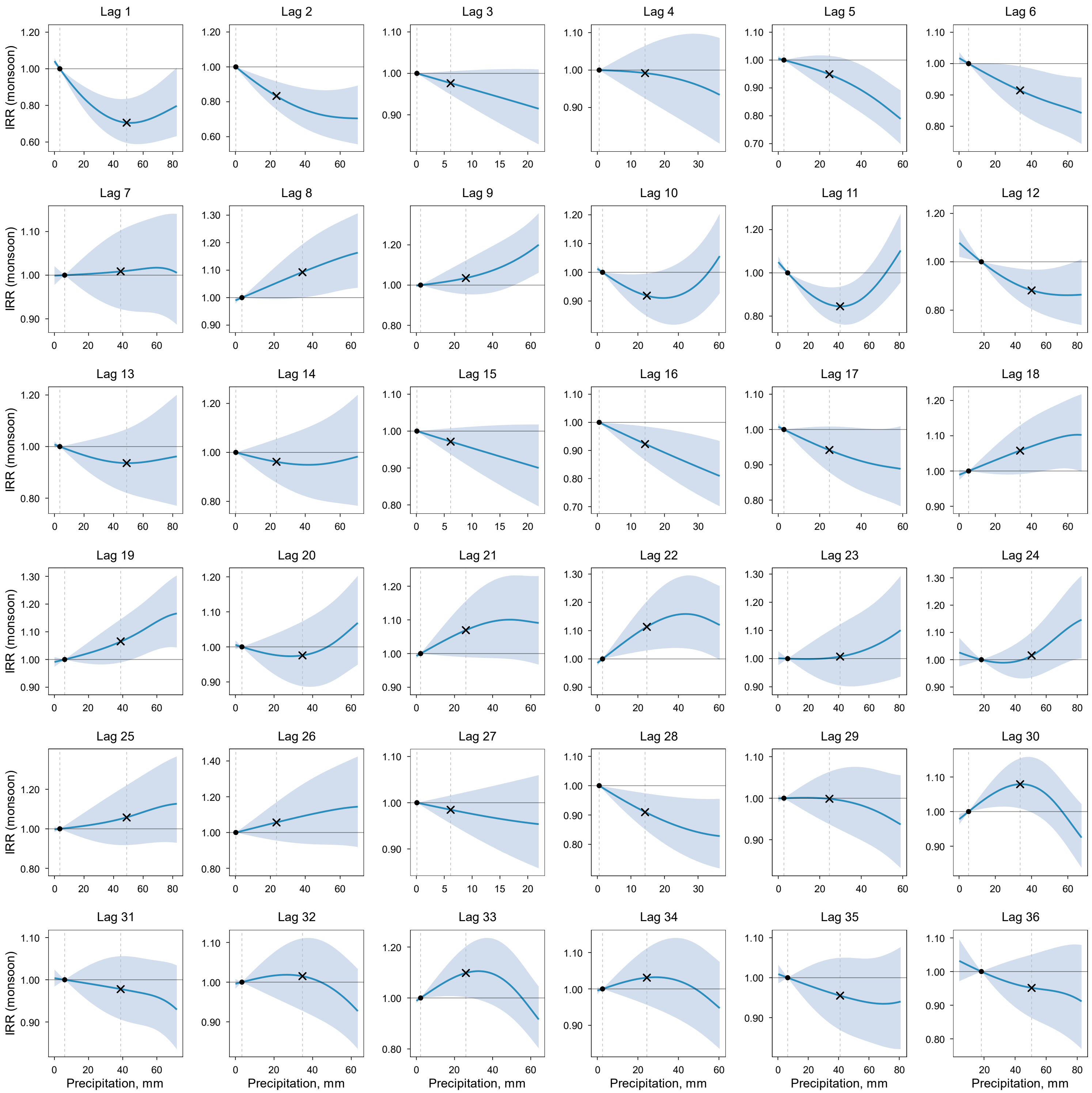

IRR (solid line) and 95% CI (shaded area) were estimated using a DLNM. Within each panel (lag month), IRR is expressed relative to the 25th percentile of the exposure variable in the study region (reference; IRR = 1), indicated by the left vertical dashed line and the dot on the IRR = 1 line. The right vertical dashed line indicates the 75th percentile, with the "×" symbol marking the estimated IRR at that level. Models were adjusted for spatiotemporal trends and surveillance-related changes in reporting and laboratory testing. Layer 2 meteorological conditions (precipitation and air temperature) were modeled simultaneously in a single DLNM with season-specific effects.

**Figure S44.** Lag-specific exposure–response relationships between monsoon (July–September) coccidioidomycosis incidence and above-ground air temperature (Layer 2: meteorological layer; AT) at lags 1–36 months.

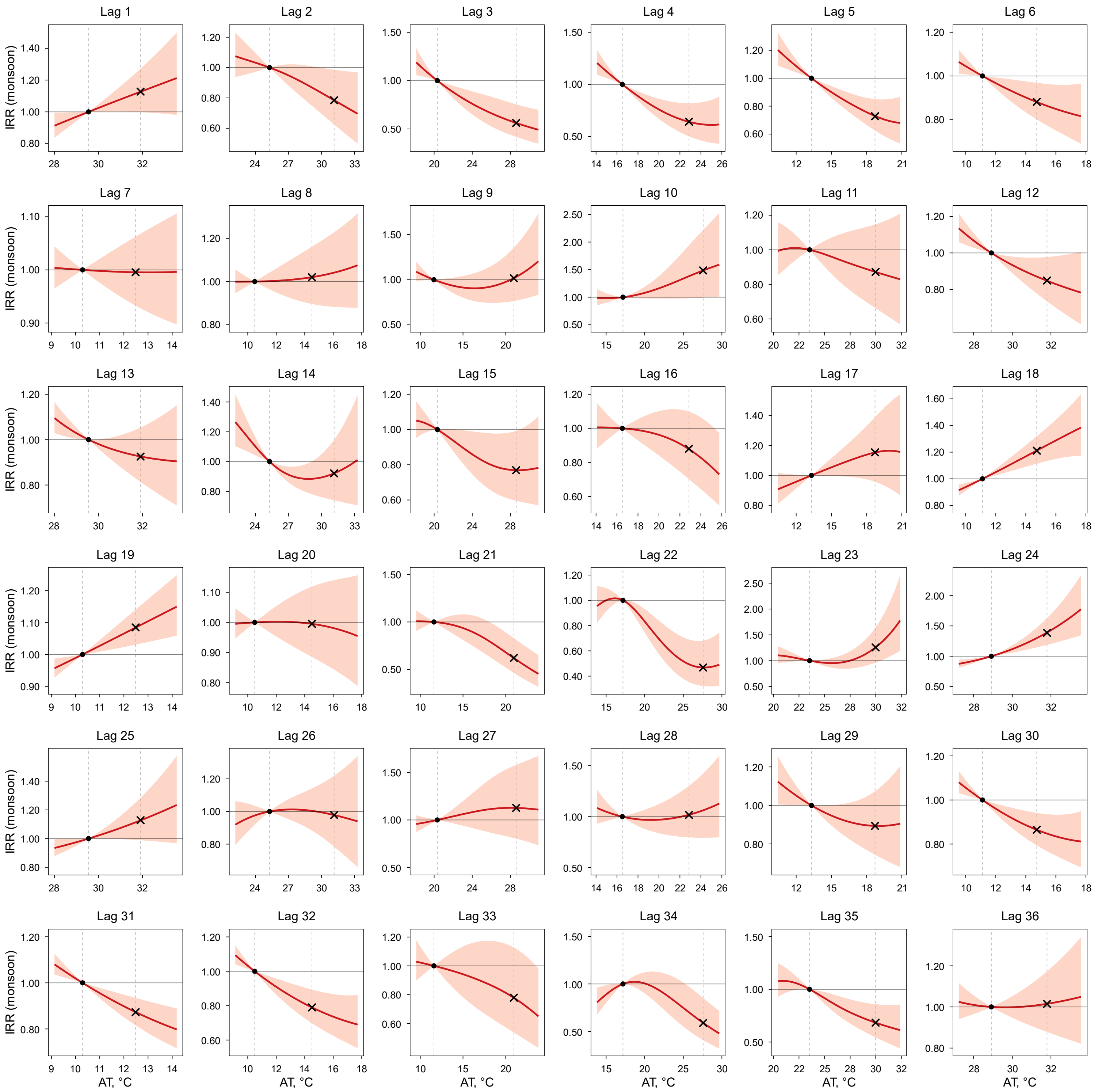

IRR (solid line) and 95% CI (shaded area) were estimated using a DLNM. Within each panel (lag month), IRR is expressed relative to the 25th percentile of the exposure variable in the study region (reference; IRR = 1), indicated by the left vertical dashed line and the dot on the IRR = 1 line. The right vertical dashed line indicates the 75th percentile, with the "×" symbol marking the estimated IRR at that level. Models were adjusted for spatiotemporal trends and surveillance-related changes in reporting and laboratory testing. Layer 2 meteorological conditions (precipitation and AT) were modeled simultaneously in a single DLNM with season-specific effects.

**Figure S45.** Lag-specific exposure–response relationships between monsoon (July–September) coccidioidomycosis incidence and above-ground $PM_{10}$ (Layer 1: dust dispersion layer) at lags 1–36 months.

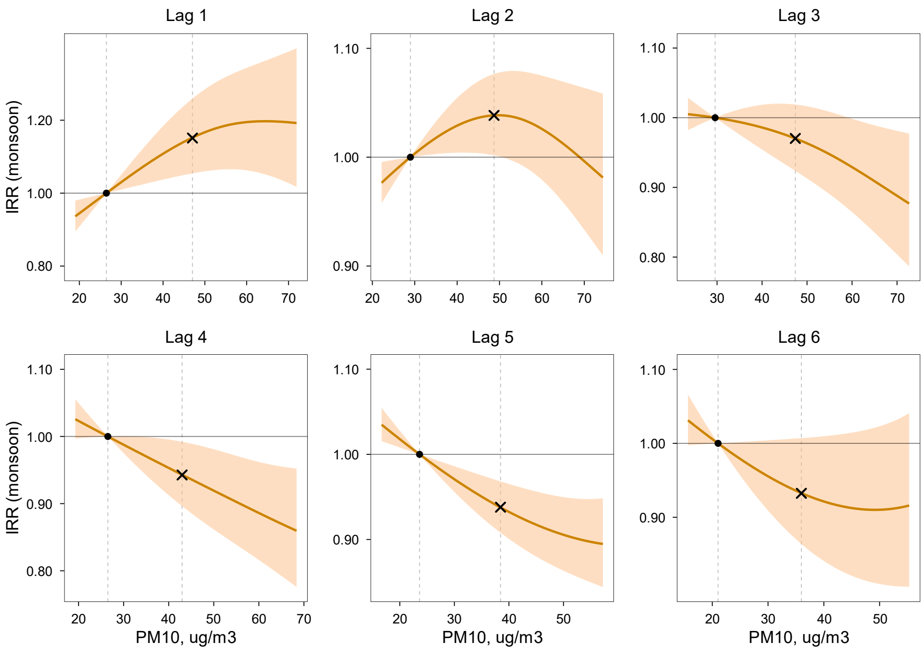

IRR (solid line) and 95% CI (shaded area) were estimated using a DLNM. Within each panel (lag month), IRR is expressed relative to the 25th percentile of the exposure variable in the study region (reference; IRR = 1), indicated by the left vertical dashed line and the dot on the IRR = 1 line. The right vertical dashed line indicates the 75th percentile, with the "×" symbol marking the estimated IRR at that level. Models were adjusted for spatiotemporal trends and surveillance-related changes in reporting and laboratory testing. Layer 1 dust dispersion conditions ($PM_{10}$ and wind speed) were modeled simultaneously in a single DLNM with season-specific effects.

**Figure S46.** Lag-specific exposure–response relationships between monsoon (July–September) coccidioidomycosis incidence and above-ground wind speed (Layer 1: dust dispersion layer) at lags 1–36 months.

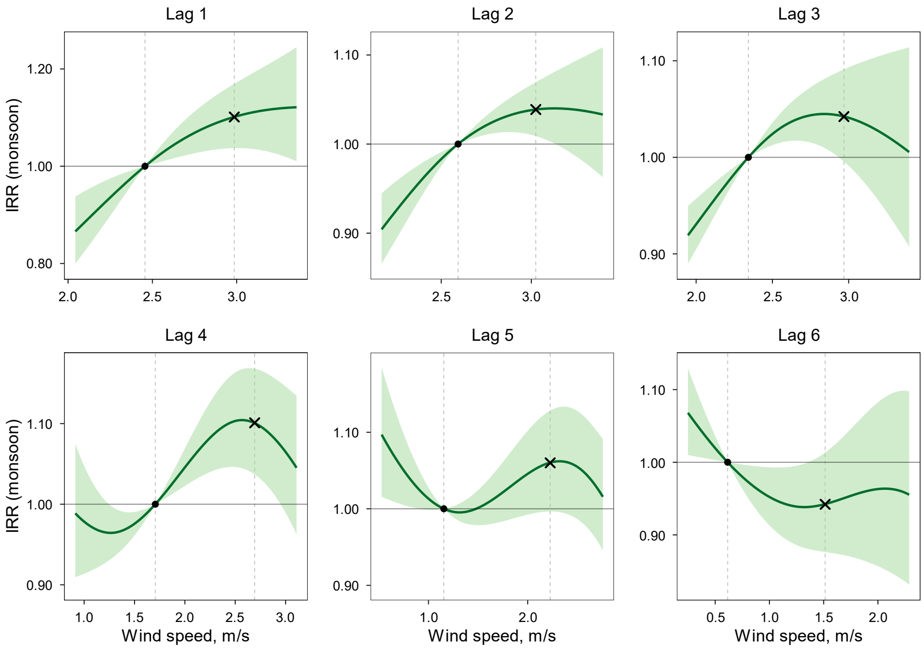

IRR (solid line) and 95% CI (shaded area) were estimated using a DLNM. Within each panel (lag month), IRR is expressed relative to the 25th percentile of the exposure variable in the study region (reference; IRR = 1), indicated by the left vertical dashed line and the dot on the IRR = 1 line. The right vertical dashed line indicates the 75th percentile, with the "×" symbol marking the estimated IRR at that level. Models were adjusted for spatiotemporal trends and surveillance-related changes in reporting and laboratory testing. Layer 1 dust dispersion conditions ($PM_{10}$ and wind speed) were modeled simultaneously in a single DLNM with season-specific effects.

**Figure S47.** Lag-specific exposure–response relationships between fall (October–December) coccidioidomycosis incidence and below-ground soil moisture (Layer 3: 0–10 cm; topsoil; SM0t10) at lags 1–36 months.

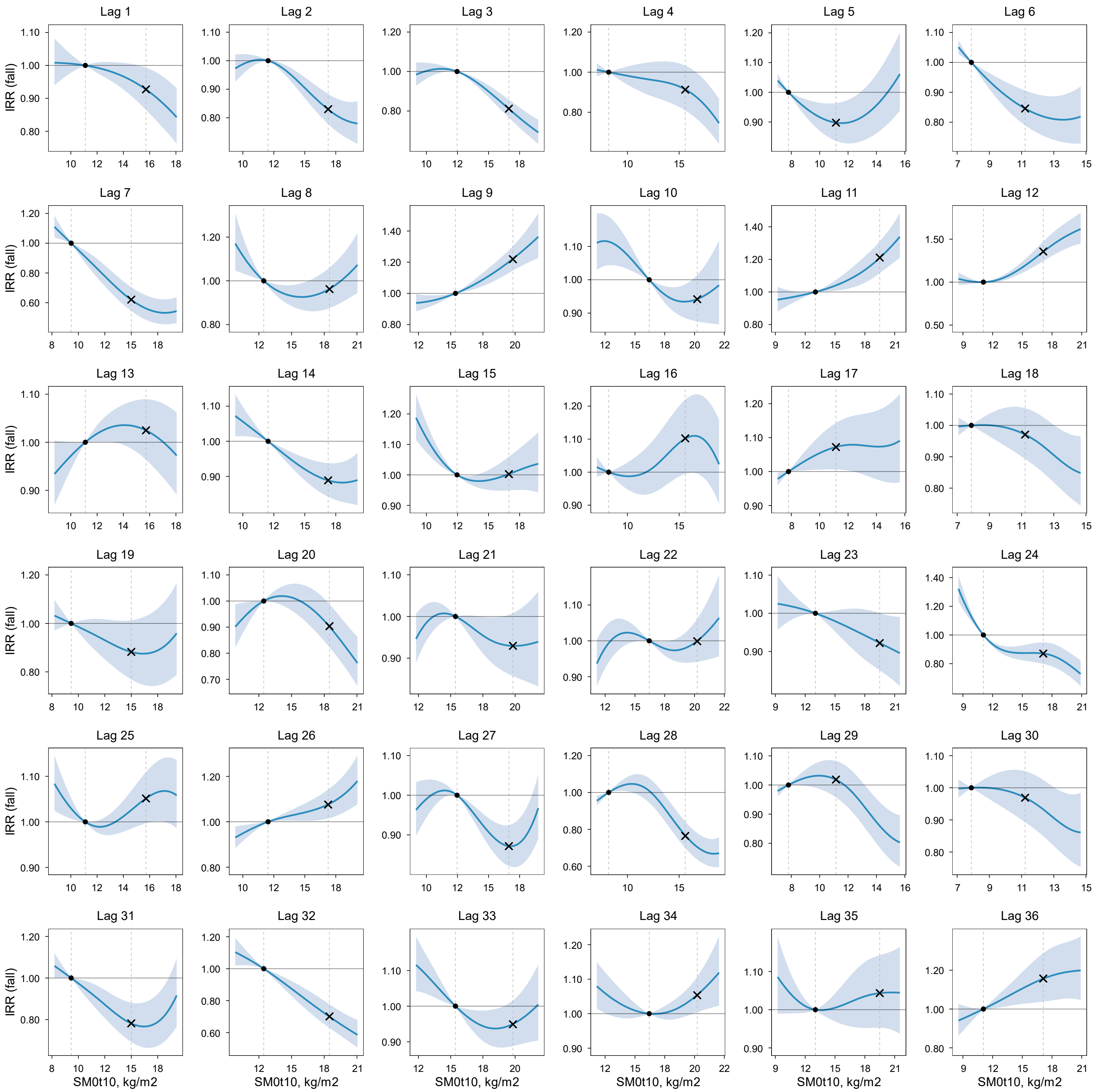

IRR (solid line) and 95% CI (shaded area) were estimated using a DLNM. Within each panel (lag month), IRR is expressed relative to the 25th percentile of the exposure variable in the study region (reference; IRR = 1), indicated by the left vertical dashed line and the dot on the IRR = 1 line. The right vertical dashed line indicates the 75th percentile, with the "×" symbol marking the estimated IRR at that level. Models were adjusted for spatiotemporal trends and surveillance-related changes in reporting and laboratory testing. Layer 3 soil moisture and soil temperature were modeled simultaneously in a single DLNM with season-specific effects.

**Figure S48.** Lag-specific exposure–response relationships between fall (October–December) coccidioidomycosis incidence and below-ground soil temperature (Layer 3: 0–10 cm; topsoil; ST0t10) at lags 1–36 months.

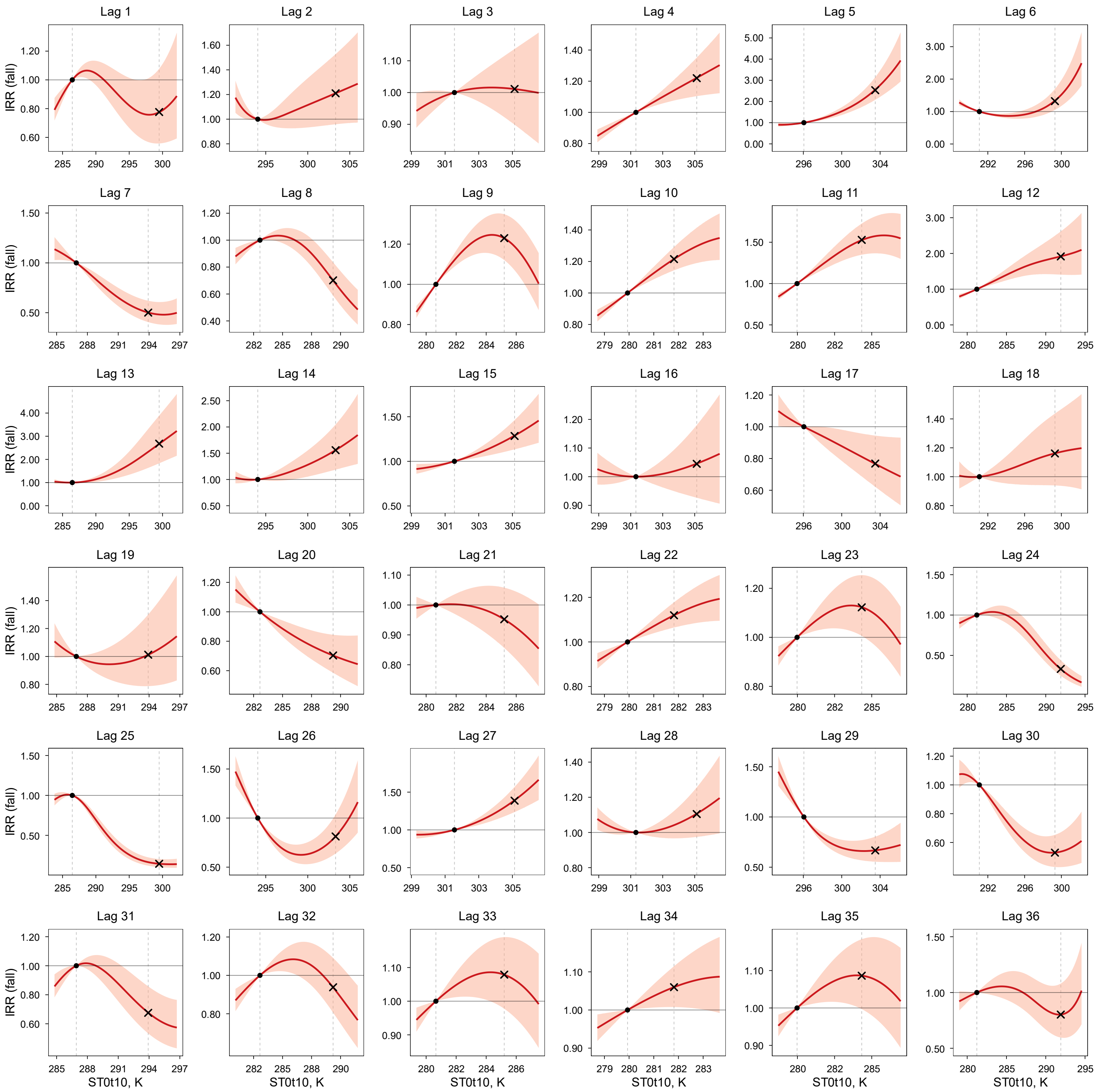

IRR (solid line) and 95% CI (shaded area) were estimated using a DLNM. Within each panel (lag month), IRR is expressed relative to the 25th percentile of the exposure variable in the study region (reference; IRR = 1), indicated by the left vertical dashed line and the dot on the IRR = 1 line. The right vertical dashed line indicates the 75th percentile, with the "×" symbol marking the estimated IRR at that level. Models were adjusted for spatiotemporal trends and surveillance-related changes in reporting and laboratory testing. Layer 3 soil moisture and soil temperature were modeled simultaneously in a single DLNM with season-specific effects.

**Figure S49.** Lag-specific exposure–response relationships between fall (October–December) coccidioidomycosis incidence and below-ground soil moisture (Layer 4: 10–40 cm; SM10t40) at lags 1–36 months.

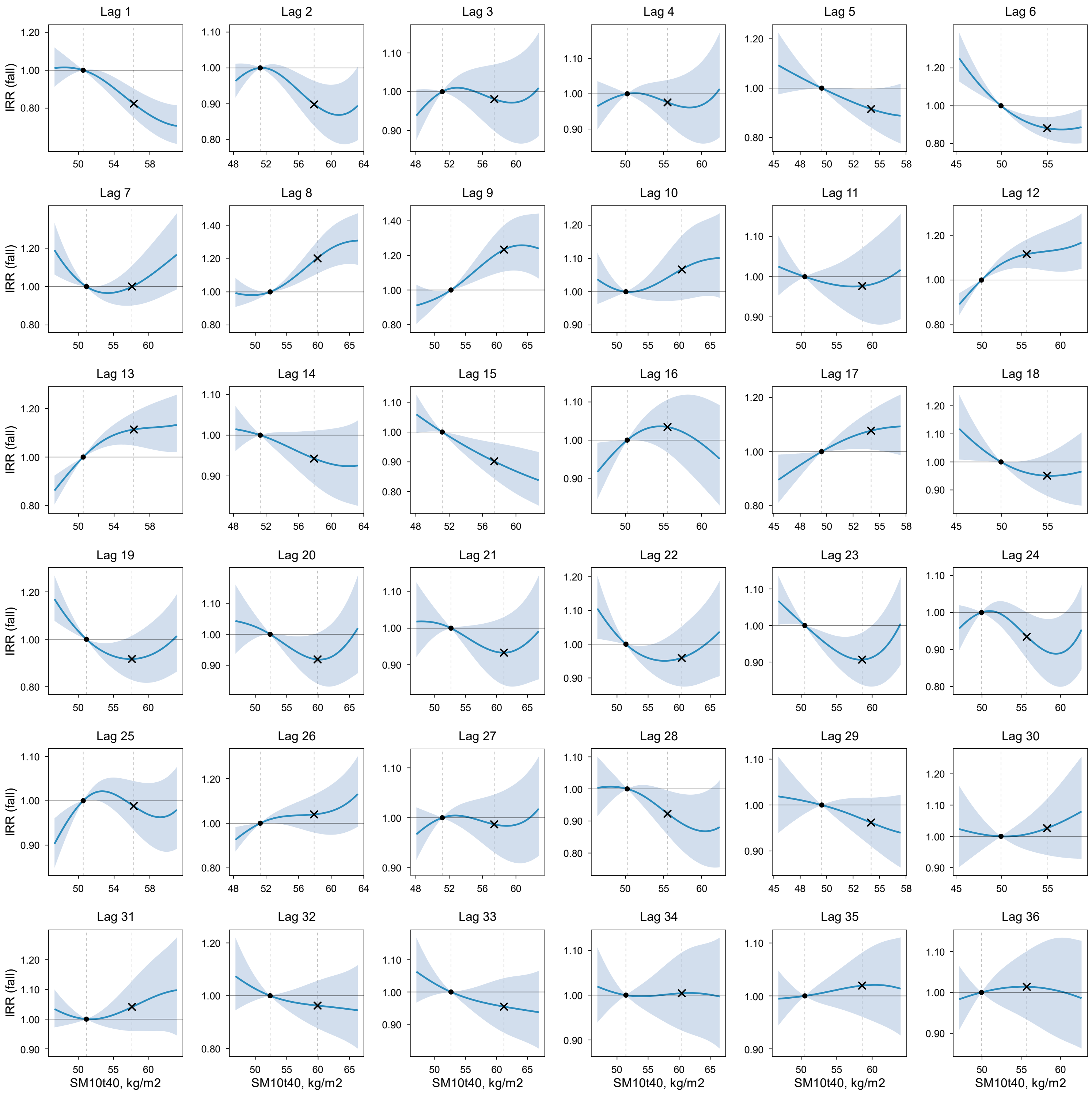

IRR (solid line) and 95% CI (shaded area) were estimated using a DLNM. Within each panel (lag month), IRR is expressed relative to the 25th percentile of the exposure variable in the study region (reference; IRR = 1), indicated by the left vertical dashed line and the dot on the IRR = 1 line. The right vertical dashed line indicates the 75th percentile, with the "×" symbol marking the estimated IRR at that level. Models were adjusted for spatiotemporal trends and surveillance-related changes in reporting and laboratory testing. Layer 4 soil moisture and soil temperature were modeled simultaneously in a single DLNM with season-specific effects.

**Figure S50.** Lag-specific exposure–response relationships between fall (October–December) coccidioidomycosis incidence and below-ground soil temperature (Layer 4: 10–40 cm; ST10t40) at lags 1–36 months.

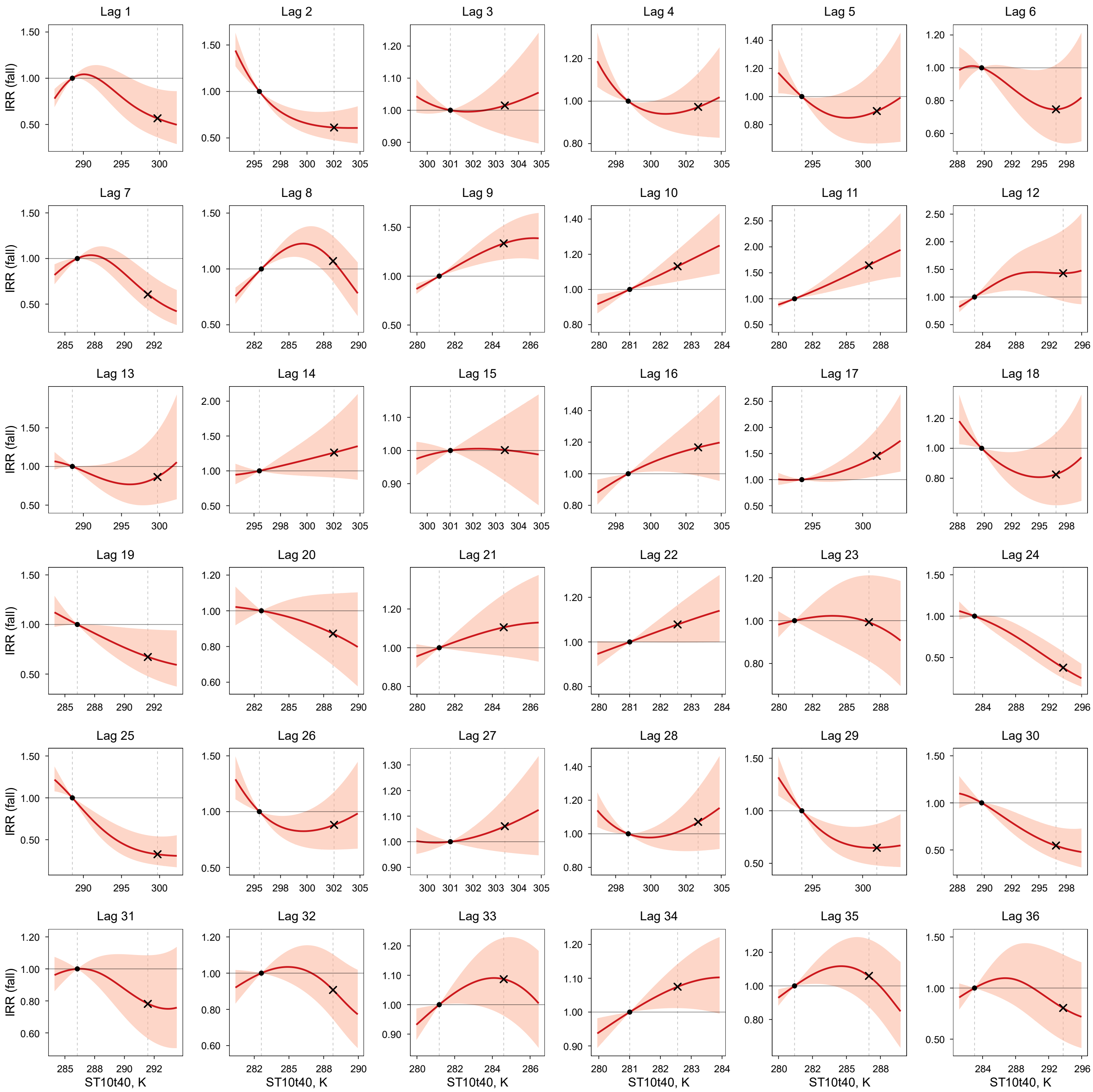

IRR (solid line) and 95% CI (shaded area) were estimated using a DLNM. Within each panel (lag month), IRR is expressed relative to the 25th percentile of the exposure variable in the study region (reference; IRR = 1), indicated by the left vertical dashed line and the dot on the IRR = 1 line. The right vertical dashed line indicates the 75th percentile, with the "×" symbol marking the estimated IRR at that level. Models were adjusted for spatiotemporal trends and surveillance-related changes in reporting and laboratory testing. Layer 4 soil moisture and soil temperature were modeled simultaneously in a single DLNM with season-specific effects.

**Figure S51.** Lag-specific exposure–response relationships between fall (October–December) coccidioidomycosis incidence and below-ground soil moisture (Layer 5: 40–100 cm; SM40t100) at lags 1–36 months.

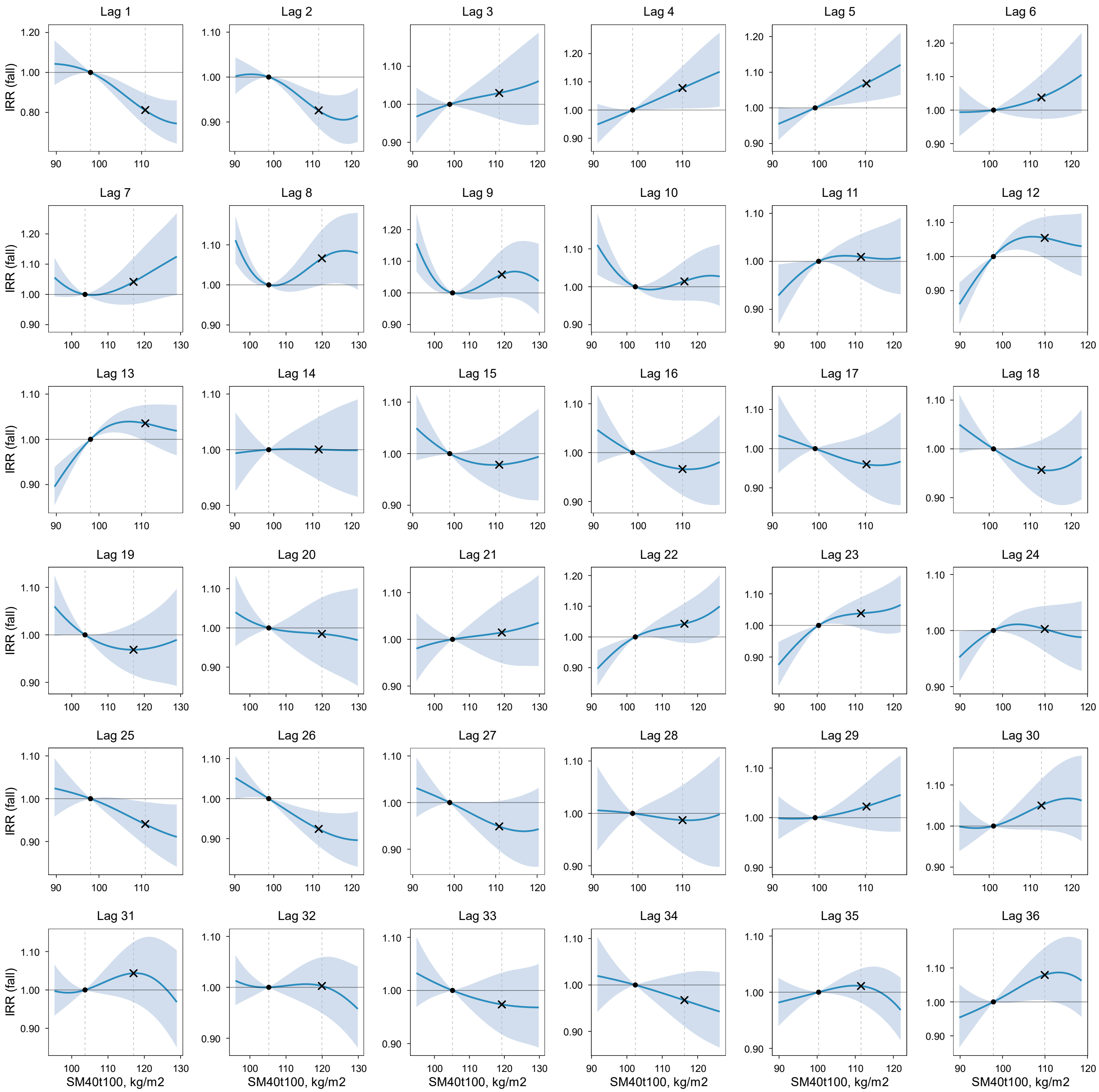

IRR (solid line) and 95% CI (shaded area) were estimated using a DLNM. Within each panel (lag month), IRR is expressed relative to the 25th percentile of the exposure variable in the study region (reference; IRR = 1), indicated by the left vertical dashed line and the dot on the IRR = 1 line. The right vertical dashed line indicates the 75th percentile, with the "×" symbol marking the estimated IRR at that level. Models were adjusted for spatiotemporal trends and surveillance-related changes in reporting and laboratory testing. Layer 5 soil moisture and soil temperature were modeled simultaneously in a single DLNM with season-specific effects.

**Figure S52.** Lag-specific exposure–response relationships between fall (October–December) coccidioidomycosis incidence and below-ground soil temperature (Layer 5: 40–100 cm; ST40t100) at lags 1–36 months.

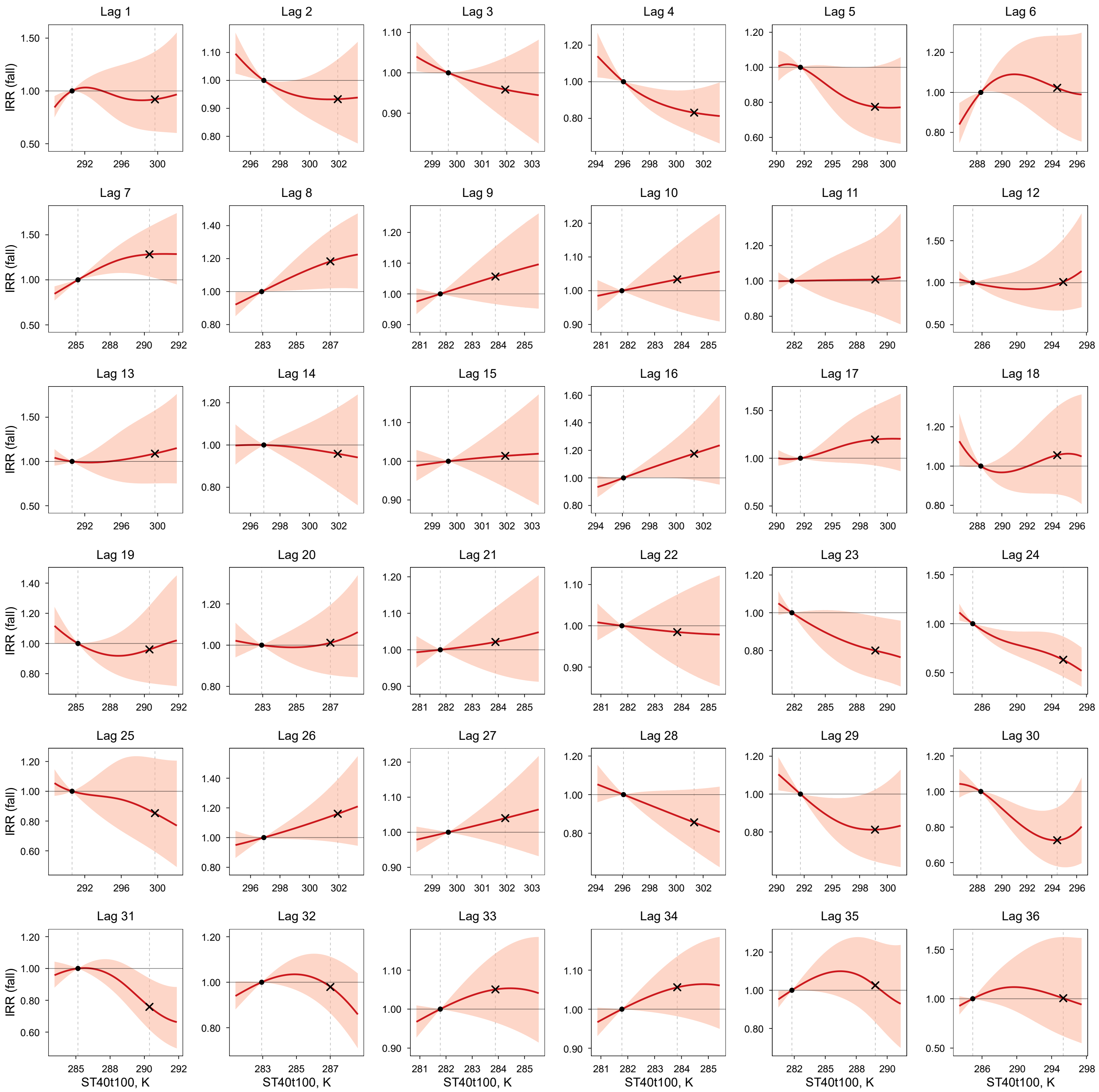

IRR (solid line) and 95% CI (shaded area) were estimated using a DLNM. Within each panel (lag month), IRR is expressed relative to the 25th percentile of the exposure variable in the study region (reference; IRR = 1), indicated by the left vertical dashed line and the dot on the IRR = 1 line. The right vertical dashed line indicates the 75th percentile, with the "×" symbol marking the estimated IRR at that level. Models were adjusted for spatiotemporal trends and surveillance-related changes in reporting and laboratory testing. Layer 5 soil moisture and soil temperature were modeled simultaneously in a single DLNM with season-specific effects.

**Figure S53.** Lag-specific exposure–response relationships between fall (October–December) coccidioidomycosis incidence and above-ground precipitation (Layer 2: meteorological layer) at lags 1–36 months.

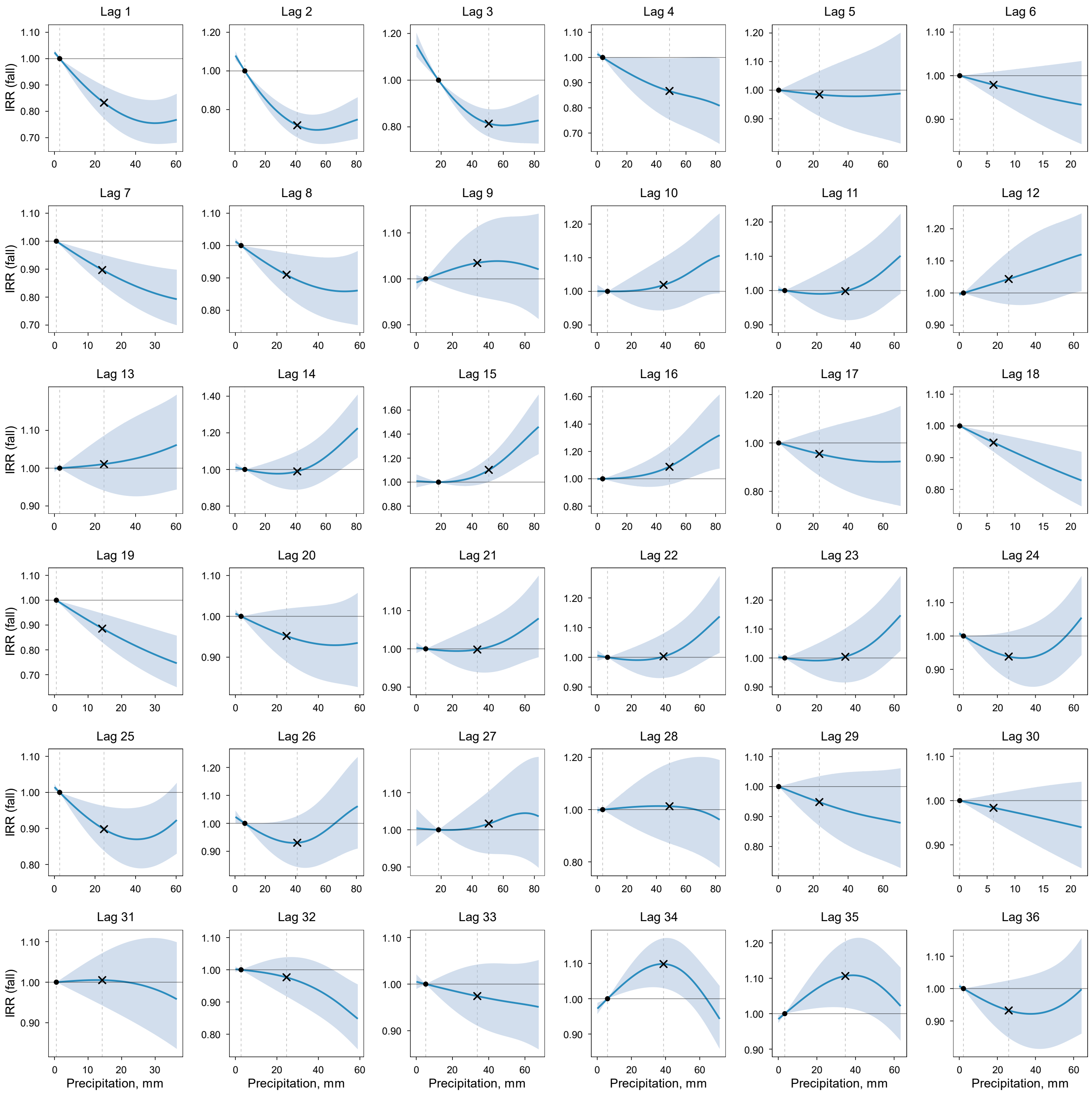

IRR (solid line) and 95% CI (shaded area) were estimated using a DLNM. Within each panel (lag month), IRR is expressed relative to the 25th percentile of the exposure variable in the study region (reference; IRR = 1), indicated by the left vertical dashed line and the dot on the IRR = 1 line. The right vertical dashed line indicates the 75th percentile, with the "×" symbol marking the estimated IRR at that level. Models were adjusted for spatiotemporal trends and surveillance-related changes in reporting and laboratory testing. Layer 2 meteorological conditions (precipitation and air temperature) were modeled simultaneously in a single DLNM with season-specific effects.

**Figure S54.** Lag-specific exposure–response relationships between fall (October–December) coccidioidomycosis incidence and above-ground air temperature (Layer 2: meteorological layer; AT) at lags 1–36 months.

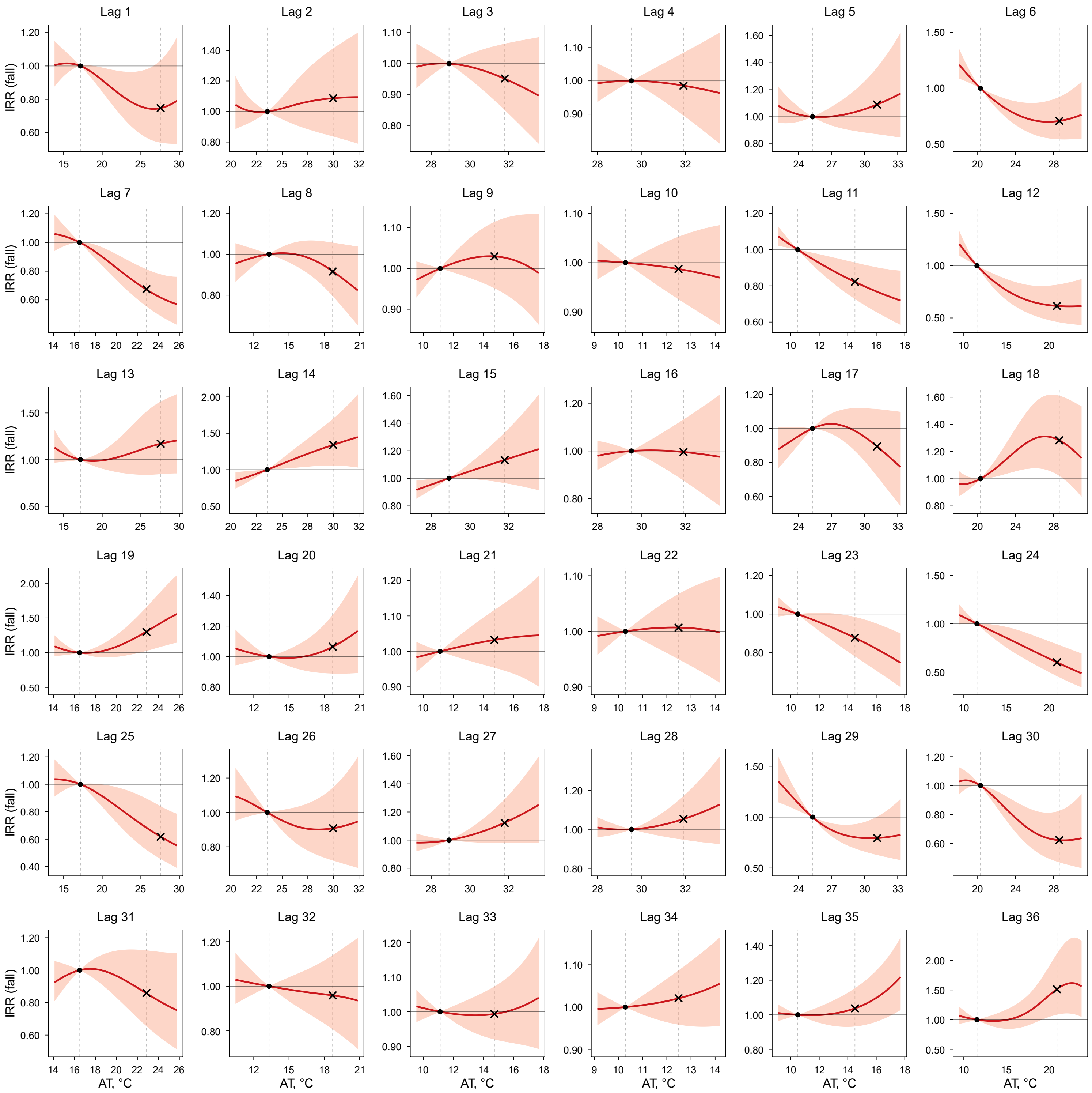

IRR (solid line) and 95% CI (shaded area) were estimated using a DLNM. Within each panel (lag month), IRR is expressed relative to the 25th percentile of the exposure variable in the study region (reference; IRR = 1), indicated by the left vertical dashed line and the dot on the IRR = 1 line. The right vertical dashed line indicates the 75th percentile, with the "×" symbol marking the estimated IRR at that level. Models were adjusted for spatiotemporal trends and surveillance-related changes in reporting and laboratory testing. Layer 2 meteorological conditions (precipitation and AT) were modeled simultaneously in a single DLNM with season-specific effects.

**Figure S55.** Lag-specific exposure–response relationships between fall (October–December) coccidioidomycosis incidence and above-ground $PM_{10}$ (Layer 1: dust dispersion layer) at lags 1–36 months.

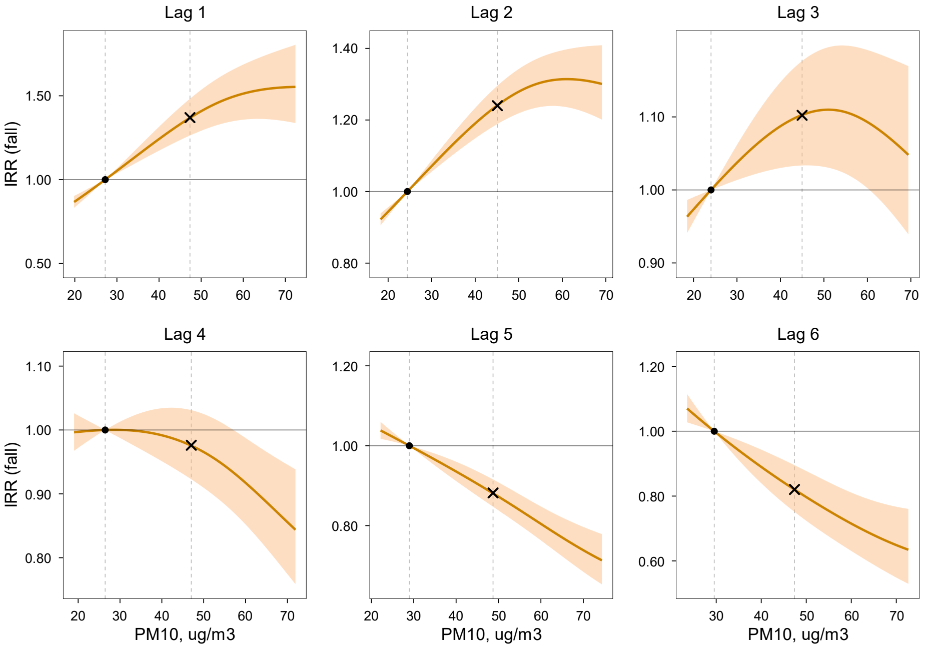

IRR (solid line) and 95% CI (shaded area) were estimated using a DLNM. Within each panel (lag month), IRR is expressed relative to the 25th percentile of the exposure variable in the study region (reference; IRR = 1), indicated by the left vertical dashed line and the dot on the IRR = 1 line. The right vertical dashed line indicates the 75th percentile, with the "×" symbol marking the estimated IRR at that level. Models were adjusted for spatiotemporal trends and surveillance-related changes in reporting and laboratory testing. Layer 1 dust dispersion conditions ($PM_{10}$ and wind speed) were modeled simultaneously in a single DLNM with season-specific effects.

**Figure S56.** Lag-specific exposure–response relationships between fall (October–December) coccidioidomycosis incidence and above-ground wind speed (Layer 1: dust dispersion layer) at lags 1–36 months.

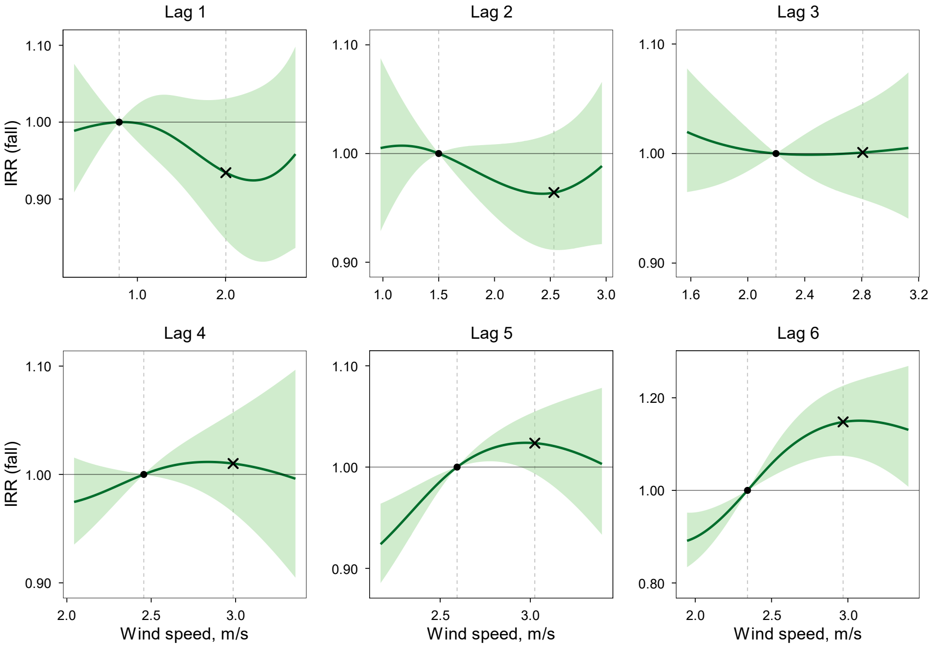

IRR (solid line) and 95% CI (shaded area) were estimated using a DLNM. Within each panel (lag month), IRR is expressed relative to the 25th percentile of the exposure variable in the study region (reference; IRR = 1), indicated by the left vertical dashed line and the dot on the IRR = 1 line. The right vertical dashed line indicates the 75th percentile, with the "×" symbol marking the estimated IRR at that level. Models were adjusted for spatiotemporal trends and surveillance-related changes in reporting and laboratory testing. Layer 1 dust dispersion conditions ($PM_{10}$ and wind speed) were modeled simultaneously in a single DLNM with season-specific effects.

**Figure S57.** Two-stage multilayer stacked ensemble machine learning framework for forecasting coccidioidomycosis incidence.

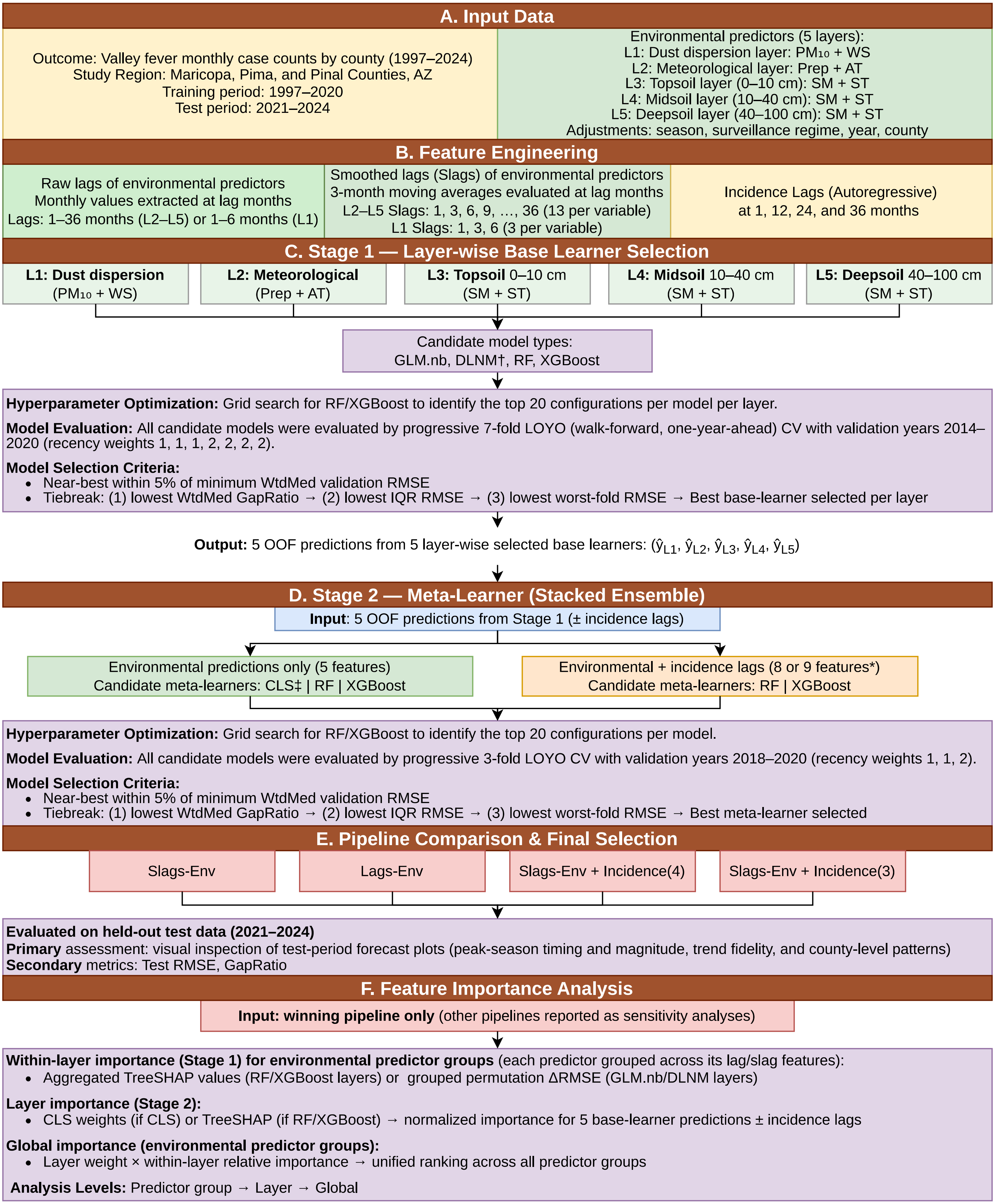

**Abbreviations**: AZ, Arizona; WS, wind speed; Prep, precipitation; AT, air temperature; SM, soil moisture; ST, soil temperature; Env, environmental; Slags, smoothed lags (3-month moving averages); GLM.nb, generalized linear model with negative binomial distribution; DLNM, distributed lag non-linear model (see †); RF, random forest; XGBoost, extreme gradient boosting; CLS, constrained least squares (see ‡); LOYO, leave-one-year-out; CV, cross-validation; OOF, out-of-fold; WtdMed, recency-weighted median; RMSE, root mean square error; GapRatio, gap ratio (measure of overfitting); TreeSHAP, Tree Shapley Additive Explanations; ΔRMSE, change in RMSE (used for grouped permutation importance).

† DLNM was applicable only in the Raw Lags (Lags-Env) pipeline configuration because its cross-basis function requires a contiguous time series to simultaneously estimate bi-dimensional exposure–response and lag–response surfaces. In the Slags pipelines, environmental predictors are pre-aggregated into 3-month moving averages at 13 sparse lag positions. This aggregation collapses the temporal granularity necessary to construct the internal lag matrix and estimate a smooth, continuous lag–response structure.

‡ CLS finds a convex combination of the K base-learner OOF predictions that minimizes the sum of squared errors: $min \sum_t \sum_{i=1}^{n} (Y_{i,t} - \sum_{k=1}^{5} \alpha_k \hat{Y}_{i,t,k})^2$ subject to $\sum_{k=1}^{5} \alpha_k = 1$ and all $\alpha_k \geq 0$ (where $\hat{Y}_{i,t,k}$ denotes the prediction of $Y_{i,t}$ from $k$th layer's model), solved via quadratic programming. The resulting weights are directly interpretable as each base learner's proportional contribution to the final ensemble forecast. CLS was excluded from pipeline configurations that included incidence lags because the sum-to-1 constraint loses its interpretive meaning when the feature space is mixed: forcing ensemble weights and autoregressive coefficients onto the same simplex conflates two conceptually distinct quantities. RF and XGBoost impose no such constraint and can naturally accommodate mixed feature types.

* (Stage 2 incidence lags): Autoregressive components introduced at the meta-learner level to capture temporal dependencies alongside environmental drivers. The number of meta-learner input features varies by configuration: 8 features for "Slags-Env + Incidence(3)" (5 OOF predictions + incidence lags at 12, 24, and 36 months) and 9 features for "Slags-Env + Incidence(4)" (5 OOF predictions + incidence lags at 1, 12, 24, and 36 months).

**Pipleline configurations**: Four end-to-end pipeline configurations were compared. (1) Slags-Env: Stage 1 base learners use smoothed-lag (Slag) environmental features; the Stage 2 meta-learner uses the 5 OOF predictions from Stage 1 only (environmental signal only, no autoregressive terms); candidate meta-learners: CLS, RF, XGBoost. (2) Lags-Env: Stage 1 base learners use raw-lag environmental features; the Stage 2 meta-learner uses the 5 OOF predictions from Stage 1 only; candidate base learners include DLNM in addition to GLM.nb, RF, and XGBoost; candidate meta-learners: CLS, RF, XGBoost. (3) Slags-Env + Incidence(4): Stage 1 identical to Slags-Env; the Stage 2 meta-learner additionally incorporates autoregressive incidence lags at 1, 12, 24, and 36 months (9 features total); candidate meta-learners: RF and XGBoost only (CLS excluded). (4) Slags-Env + Incidence(3): Stage 1 identical to Slags-Env; the Stage 2 meta-learner incorporates autoregressive incidence lags at 12, 24, and 36 months only (8 features total); candidate meta-learners: RF and XGBoost only (CLS excluded).

**Pipeline evaluation**: *Primary criterion*: visual inspection of held-out test-period forecast plots, specifically peak-season timing and magnitude, trend fidelity, and consistency across the combined study region and individual counties. *Secondary criterion*: if visual inspection does not clearly separate pipelines, test RMSE and GapRatio are used to determine the winning configuration.

**WtdMed RMSE**: Recency-weighted median of fold-level validation RMSE computed in two stages: (1) Seed Aggregation: For RF and XGBoost, the fold-level RMSE is the median across five independent random seeds; GLM.nb, DLNM, and CLS are deterministic and require only a single run per fold; (2) Fold Aggregation: Fold-level RMSEs are summarized as a recency-weighted median, assigning higher weights to more recent validation years (Stage 1 weights 1, 1, 1, 2, 2, 2, 2 for 2014–2020; Stage 2 weights 1, 1, 2 for 2018–2020).

**WtdMed GapRatio** measures a model's tendency to overfit. For each CV fold, GapRatio = max(0, (validation RMSE − training RMSE) / validation RMSE): a value of 0 indicates perfect generalization (no gap between training and validation performance), while a value approaching 1 indicates severe overfitting (validation error far exceeds training error). WtdMed GapRatio is the recency-weighted median of fold-level GapRatio values across all CV folds, using the same recency weights as WtdMed RMSE. Among configurations that are near-best on WtdMed RMSE (within 5% of the minimum), the configuration with the lowest WtdMed GapRatio is preferred, as it is the least prone to overfitting.

**IQR of validation RMSE** (interquartile range of fold-level validation RMSE) measures the year-to-year consistency of a model's forecast accuracy. It is computed as Q3 − Q1 of the fold-level validation RMSE values across all CV folds, without recency weighting. A low IQR indicates that the model performs steadily across all validation years; a high IQR indicates that performance varies substantially—the model may forecast some years well but fail in others. IQR of validation RMSE is used as the second tiebreak, after WtdMed GapRatio.

**Worst-fold validation RMSE** is the single highest fold-level validation RMSE observed across all CV folds—that is, the model's performance in its worst forecasting year. While WtdMed RMSE and IQR summarize central tendency and spread, worst-fold RMSE captures tail risk: a model with an acceptable average but a catastrophically poor single year would be penalized here. Worst-fold validation RMSE is used as the third tiebreak, after IQR of validation RMSE.

**Figure S58.** Observed and forecast monthly coccidioidomycosis incidence from the two-stage multilayer ensemble learning model in Maricopa (smoothed-lag environmental-only pipeline)**.**

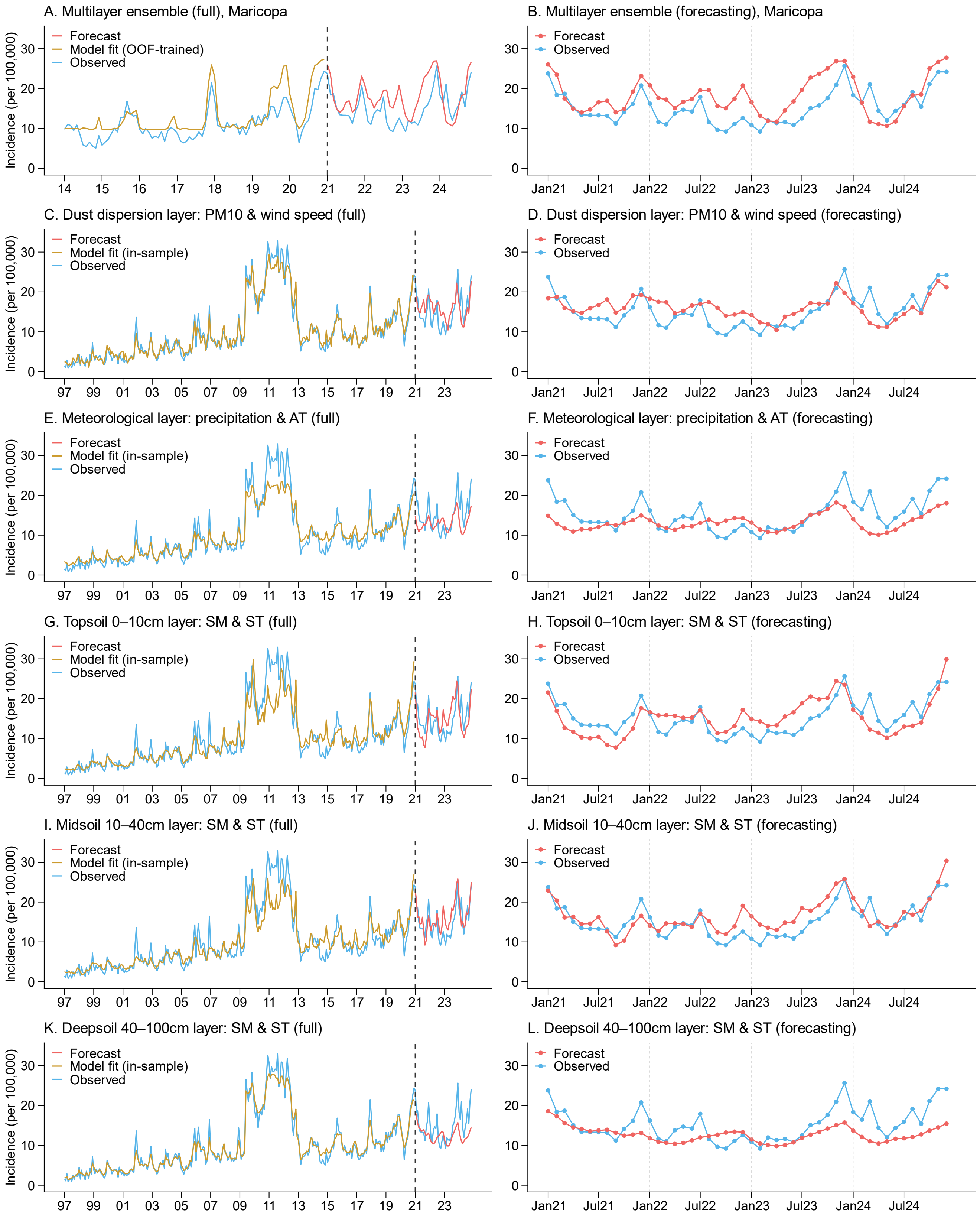

The base learners (Stage 1) were trained on 1997–2020 using smoothed-lag environmental predictors, and the meta-learner (Stage 2) was trained on out-of-fold (OOF) predictions from the base learners during 2014–2020. The model was forecast over a strictly held-out period of 2021–2024. Left panels (A, C, E, G, I, K) show the full study period; right panels (B, D, F, H, J, L) show the forecasting period only. Panels A–B show the multilayer ensemble (meta-learner) combining OOF forecasts from all base learners. Panels C–L show individual base learners (five environmental layers): dust dispersion ($PM_{10}$ and wind speed; C–D), meteorological conditions (precipitation and air temperature [AT]; E–F), topsoil 0–10 cm (soil moisture [SM] and soil temperature [ST]; G–H), middle soil 10–40 cm (I–J), and deep soil 40–100 cm (K–L).

**Figure S59.** Observed and forecast monthly coccidioidomycosis incidence from the two-stage multilayer ensemble learning model in Pima (smoothed-lag environmental-only pipeline)**.**

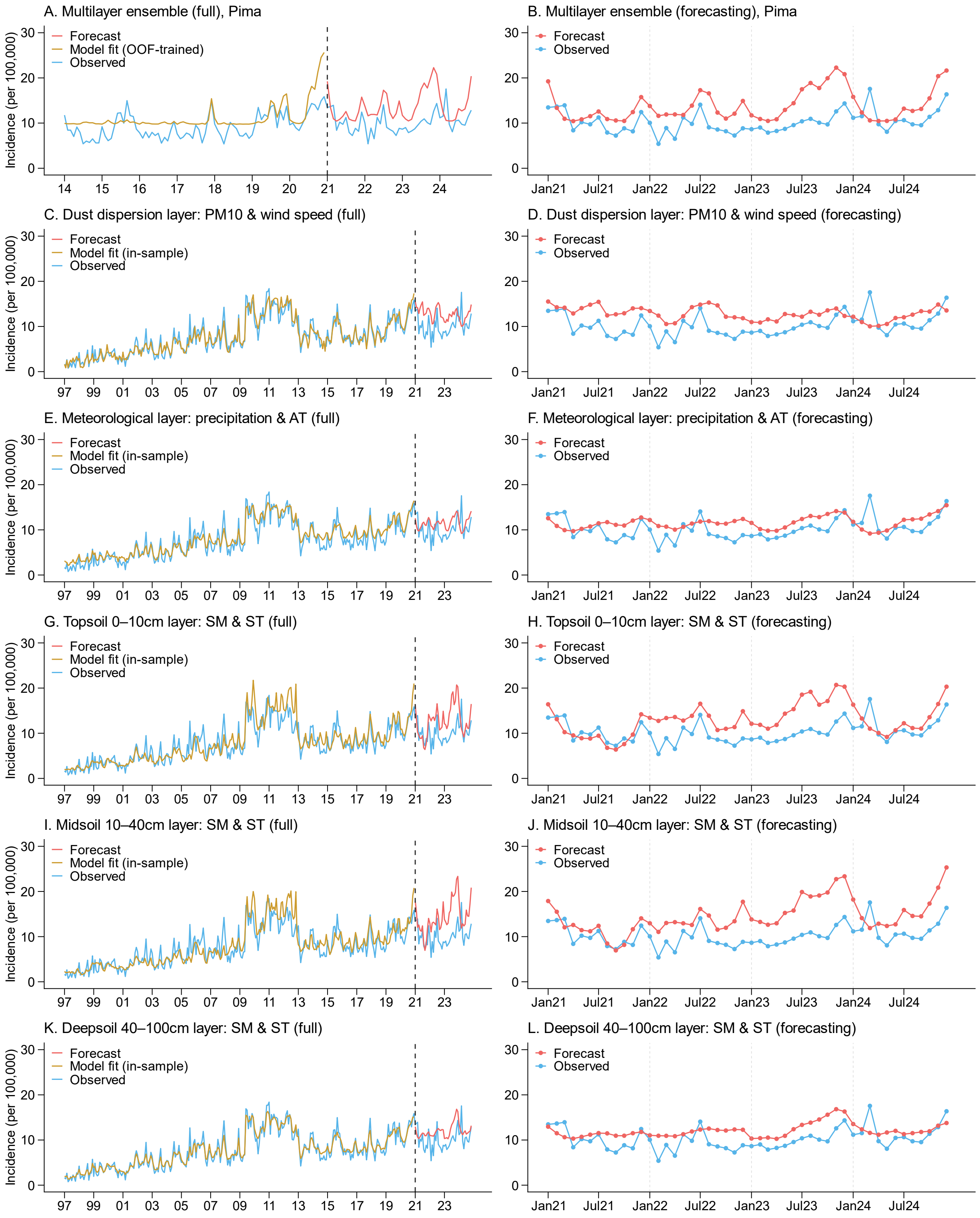

The base learners (Stage 1) were trained on 1997–2020 using smoothed-lag environmental predictors, and the meta-learner (Stage 2) was trained on out-of-fold predictions from the base learners during 2014–2020. The model was forecast over a strictly held-out period of 2021–2024. Left panels (A, C, E, G, I, K) show the full study period; right panels (B, D, F, H, J, L) show the forecasting period only. Panels A–B show the multilayer ensemble (meta-learner) combining OOF forecasts from all base learners. Panels C–L show individual base learners: dust dispersion ($PM_{10}$ and wind speed; C–D), meteorological conditions (precipitation and air temperature; E–F), topsoil 0–10 cm (soil moisture and soil temperature; G–H), midsoil 10–40 cm (I–J), and deepsoil 40–100 cm (K–L).

**Figure S60.** Observed and forecast monthly coccidioidomycosis incidence from the two-stage multilayer ensemble learning model in Pinal (smoothed-lag environmental-only pipeline)**.**

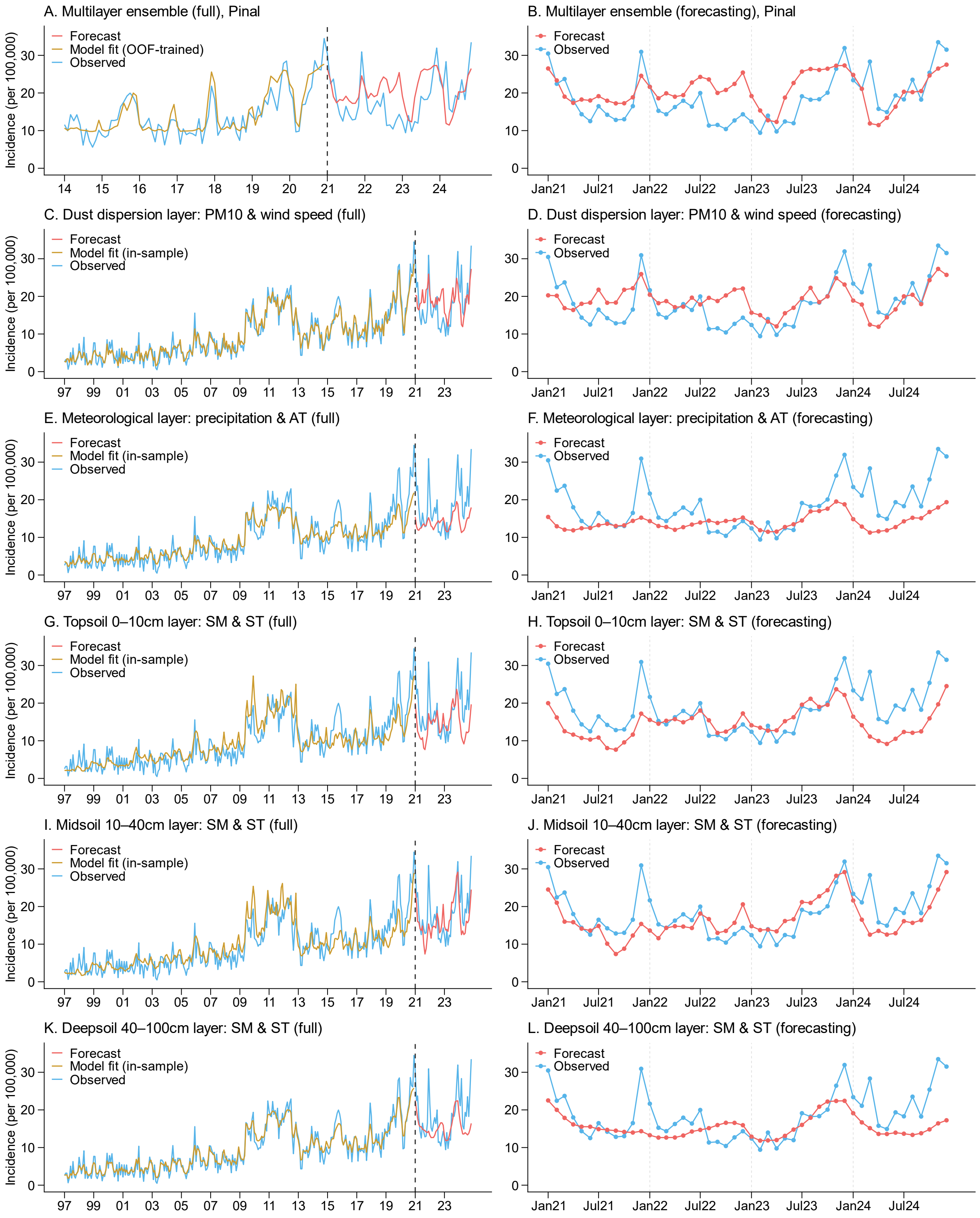

The base learners (Stage 1) were trained on 1997–2020 using smoothed-lag environmental predictors, and the meta-learner (Stage 2) was trained on out-of-fold predictions from the base learners during 2014–2020. The model was forecast over a strictly held-out period of 2021–2024. Left panels (A, C, E, G, I, K) show the full study period; right panels (B, D, F, H, J, L) show the forecasting period only. Panels A–B show the multilayer ensemble (meta-learner) combining OOF forecasts from all base learners. Panels C–L show individual base learners: dust dispersion ($PM_{10}$ and wind speed; C–D), meteorological conditions (precipitation and air temperature; E–F), topsoil 0–10 cm (soil moisture and soil temperature; G–H), midsoil 10–40 cm (I–J), and deepsoil 40–100 cm (K–L).

**Figure S61.** Within-layer predictive importance of environmental predictor groups versus adjustment covariates across five base learner layers in the stage-1 winner models (smoothed-lag environmental-only pipeline).

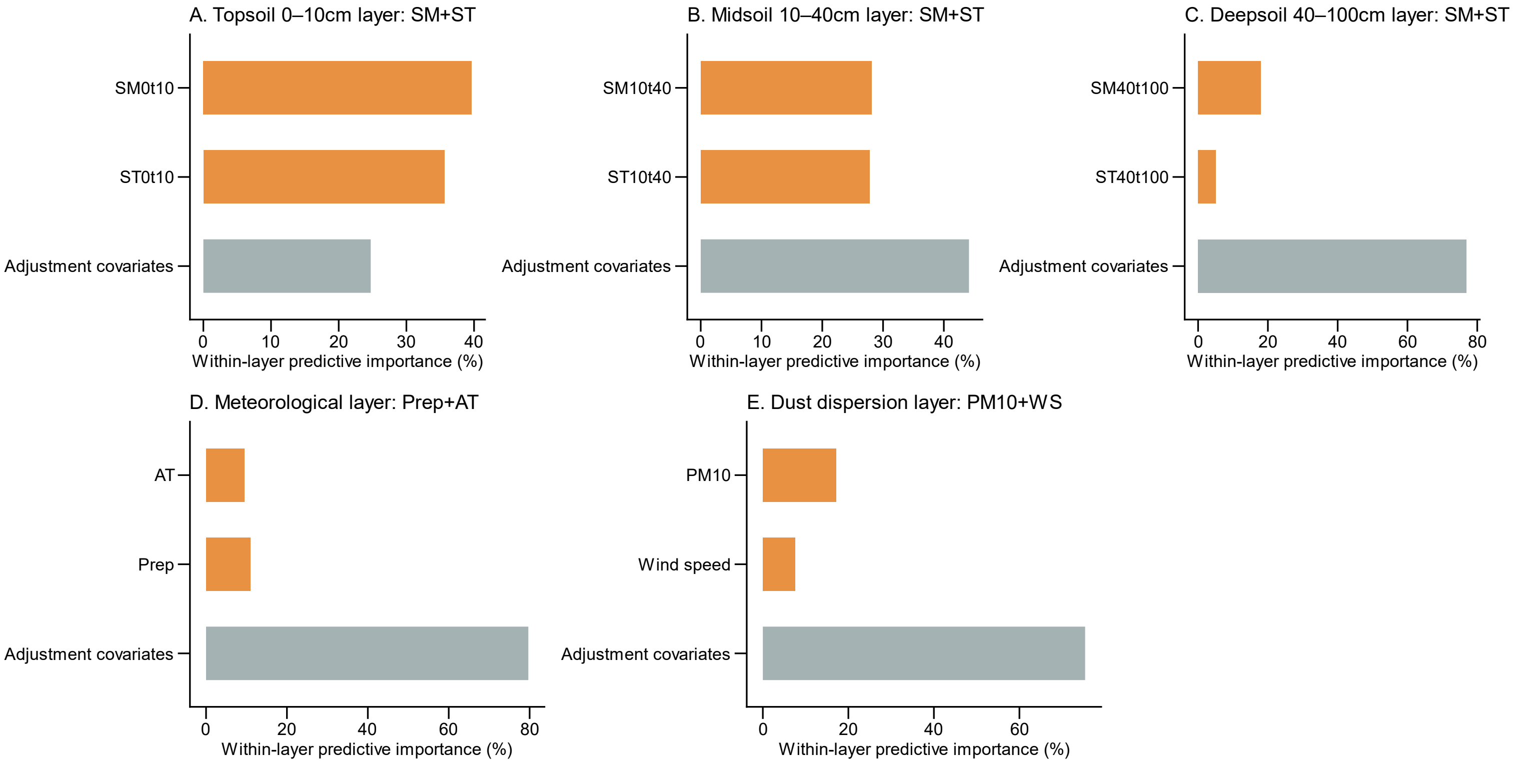

Results are shown for the winning pipeline configuration—the smoothed-lag environmental-only (Slags-Env) pipeline—which uses smoothed-lag environmental predictors. Bars show the relative predictive importance (%) of each predictor group within a given layer, normalized to sum to 100% per panel. Each environmental predictor group comprises smoothed lagged features (3-month moving averages) at lag positions 1, 3, 6, 9, 12, 15, 18, 21, 24, 27, 30, 33, and 36 months (13 features per group) for soil and meteorological layers (A–D), and at lag positions 1, 3, and 6 months (3 features per group) for the dust layer (E). Environmental predictor groups (orange) include (A–C) soil moisture (SM) and soil temperature (ST) at three depth intervals, (D) precipitation (Prep) and air temperature (AT), and (E) $PM_{10}$ and wind speed (WS). Adjustment covariates (grouped; grey) include surveillance regime, calendar year, season, and county. Predictive importance was quantified using grouped permutation-based ΔRMSE for layers with generalized negative binomial base learners (A–B) and aggregated TreeSHAP values for layers with tree-based base learners (random forest: C–D; gradient-boosted trees: E). All importance estimates were computed on the strictly held-out test period (2021–2024).

**Figure S62**. Layer-level and global importance of environmental predictor groups in the two-stage ensemble model (smoothed-lag environmental-only pipeline).

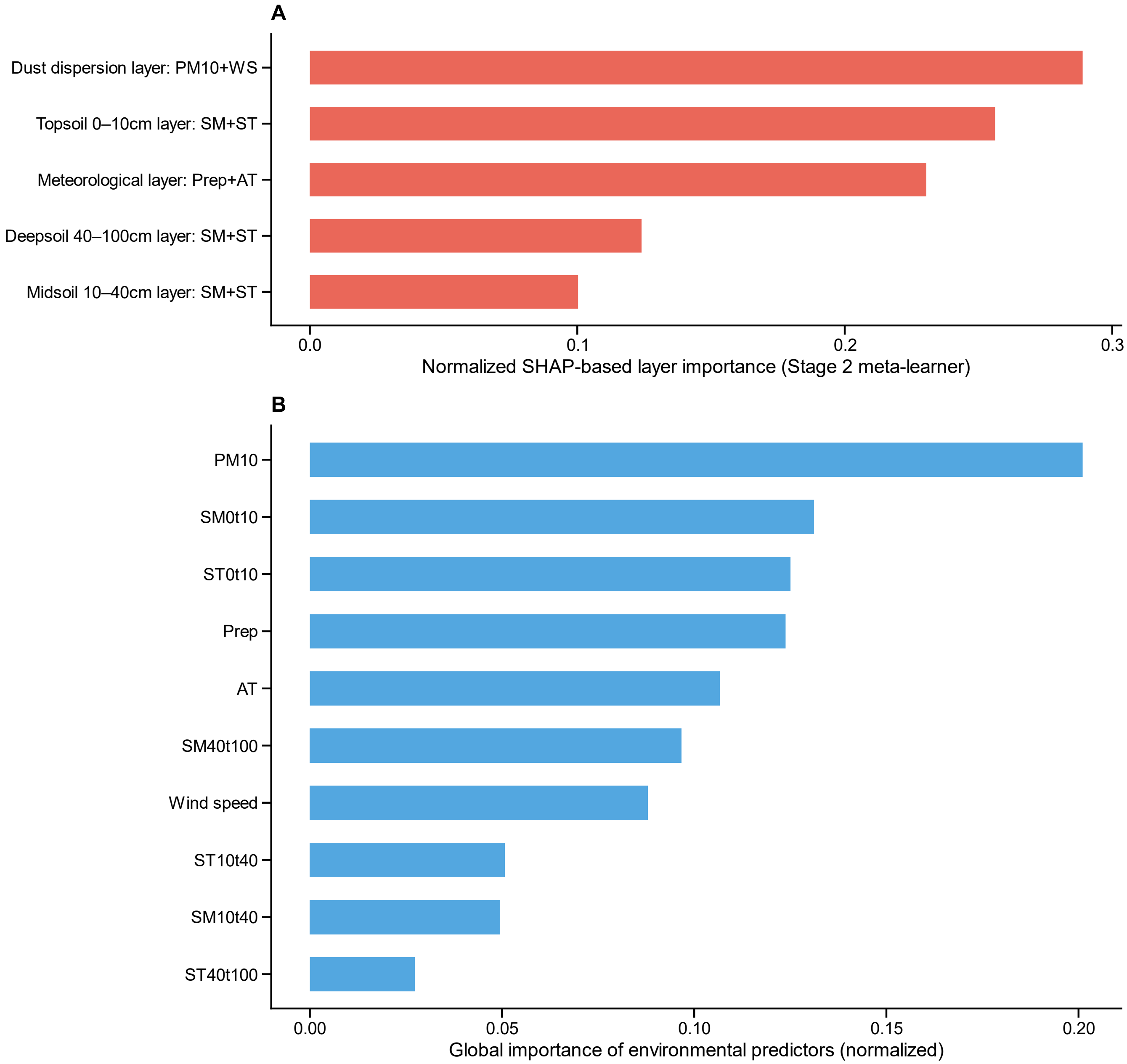

Results are shown for the winning pipeline configuration—the smoothed-lag environmental-only (Slags-Env) pipeline—which uses smoothed-lag environmental predictors. (A) Relative importance of the five base learner layers to the Stage 2 meta-learner, quantified as normalized mean absolute TreeSHAP values (summing to 1.0) from the random forest meta-learner. (B) Global importance of environmental predictor groups across all layers. Global importance was computed as the product of each layer's weight from panel A and the within-layer relative importance of each environmental predictor group (excluding adjustment covariates), normalized to sum to 1.0. Each predictor group comprises smoothed lagged features (3-month moving averages) at lag positions 1, 3, 6, 9, 12, 15, 18, 21, 24, 27, 30, 33, and 36 months (13 features per group) for soil and meteorological layers, and at lag positions 1, 3, and 6 months (3 features per group) for the dust layer. All importance estimates were computed on the strictly held-out test period (2021–2024).

**Figure S63.** Sensitivity analysis comparing multilayer ensemble forecast performance with and without incidence lag features.

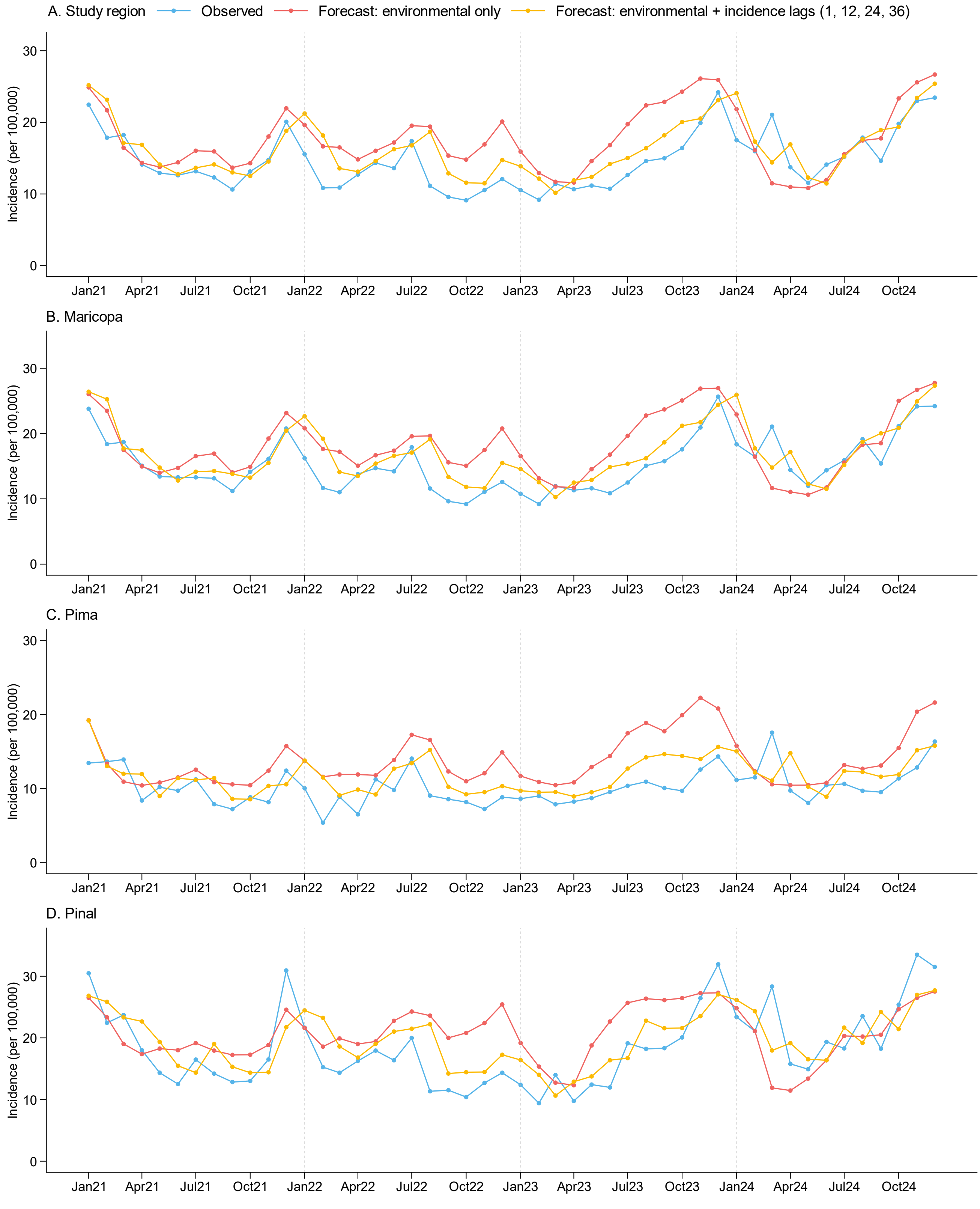

Observed and forecast monthly coccidioidomycosis incidence (per 100,000 population) from the two-stage multilayer ensemble learning model for the study region (A) and Maricopa (B), Pima (C), and Pinal (D) counties. Forecasts are compared between (1) a model using environmental predictors only (no prior-incidence inputs) and (2) a model additionally incorporating incidence lag features at 1, 12, 24, and 36 months (i.e., observed incidence at *t*–1, *t*–12, *t*–24, and *t*–36). Both models use concurrent environmental data, which are available within approximately one week of each target month; therefore, incidence estimates can be generated well before finalized surveillance data become available, providing a practical surveillance lead time. The lag-inclusive model additionally requires surveillance data from at least 1 month prior, whereas the environmental-only model eliminates the need for surveillance data entirely during the forecasting period (2021–2024).

**Figure S64.** Sensitivity analysis comparing multilayer ensemble forecasts with and without annual-cycle incidence lag features.

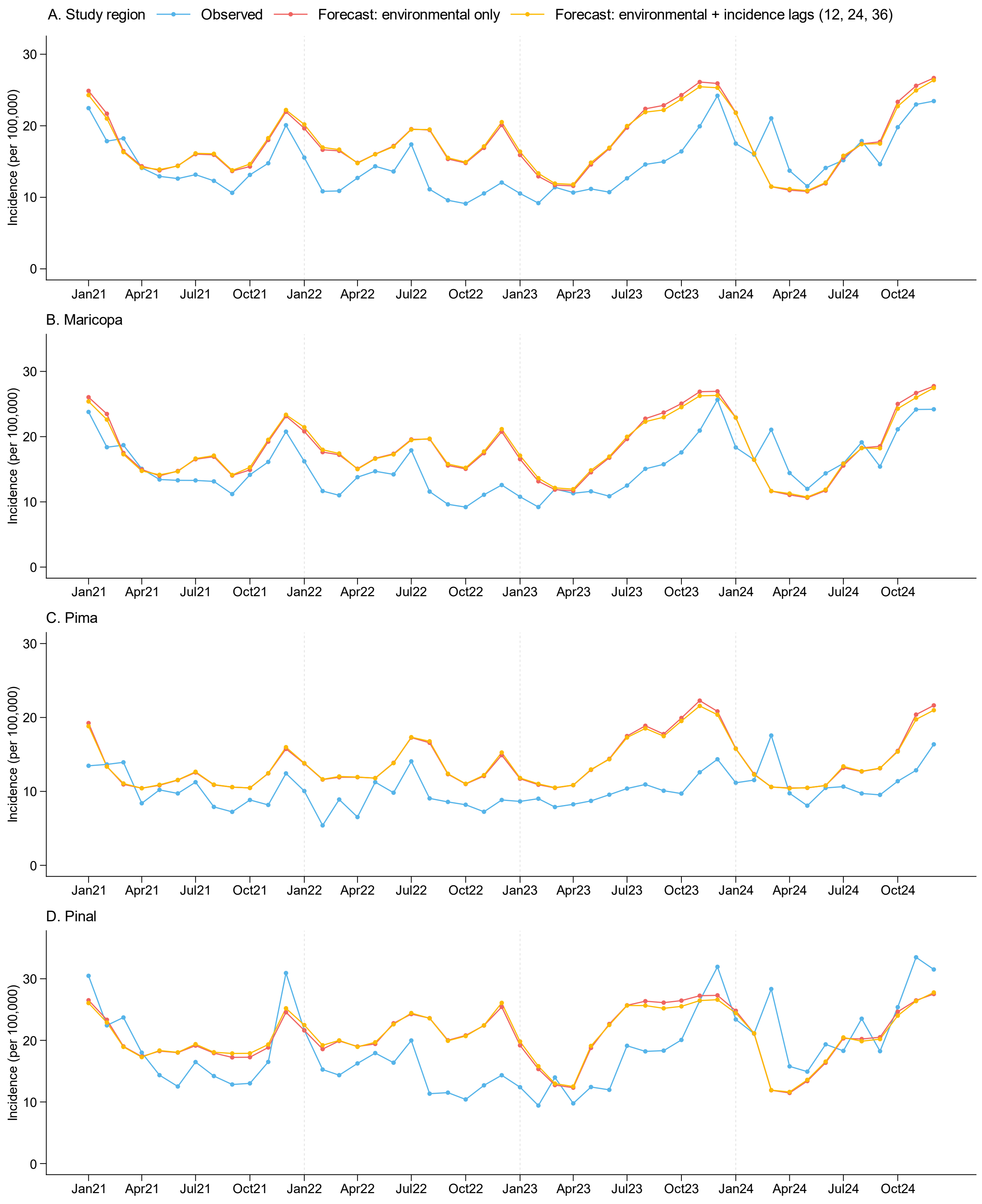

Observed and forecast monthly coccidioidomycosis incidence from the two-stage multilayer ensemble learning model for the study region (A) and Maricopa (B), Pima (C), and Pinal (D) counties. Forecasts are compared between (1) a model using environmental predictors only (no prior-incidence inputs) and (2) a model additionally incorporating incidence lag features at 12, 24, and 36 months (ie, observed incidence at *t–12*, *t–24*, and *t–36*), thereby capturing annual-cycle information without using the most recent incidence (t–1). The comparison is evaluated over the out-of-sample forecasting period (2021–2024). Lines denote observed incidence and the two forecast series, as labelled in each panel.

**Figure S65.** Sensitivity analysis comparing multilayer ensemble forecast performance using smoothed versus raw lags of environmental predictors.

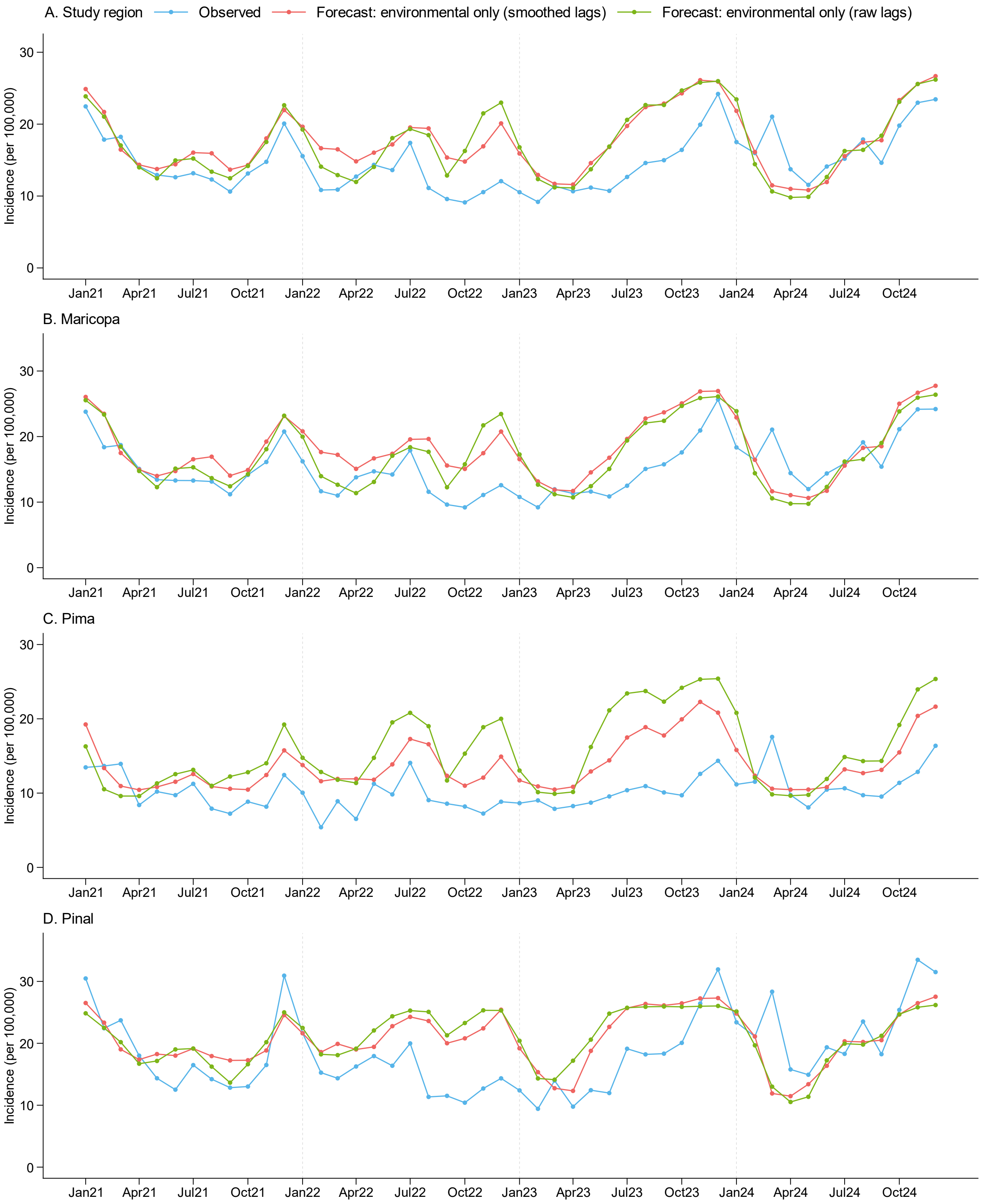

Observed and forecast monthly coccidioidomycosis incidence (per 100,000 population) from the two-stage multilayer ensemble learning model for the study region (A) and Maricopa (B), Pima (C), and Pinal (D) counties over a strictly held-out forecasting period of 2021–2024. Forecasts are compared between two models that share the same ensemble structure and environmental predictors but differ in the construction of lagged features: (1) raw individual monthly lag values (green) and (2) smoothed lag features derived from three-month moving averages (red).

**Figure S66.** Sensitivity analysis of forecasting performance using raw (unsmoothed) lags of environmental predictors in the two-stage multilayer ensemble learning framework.

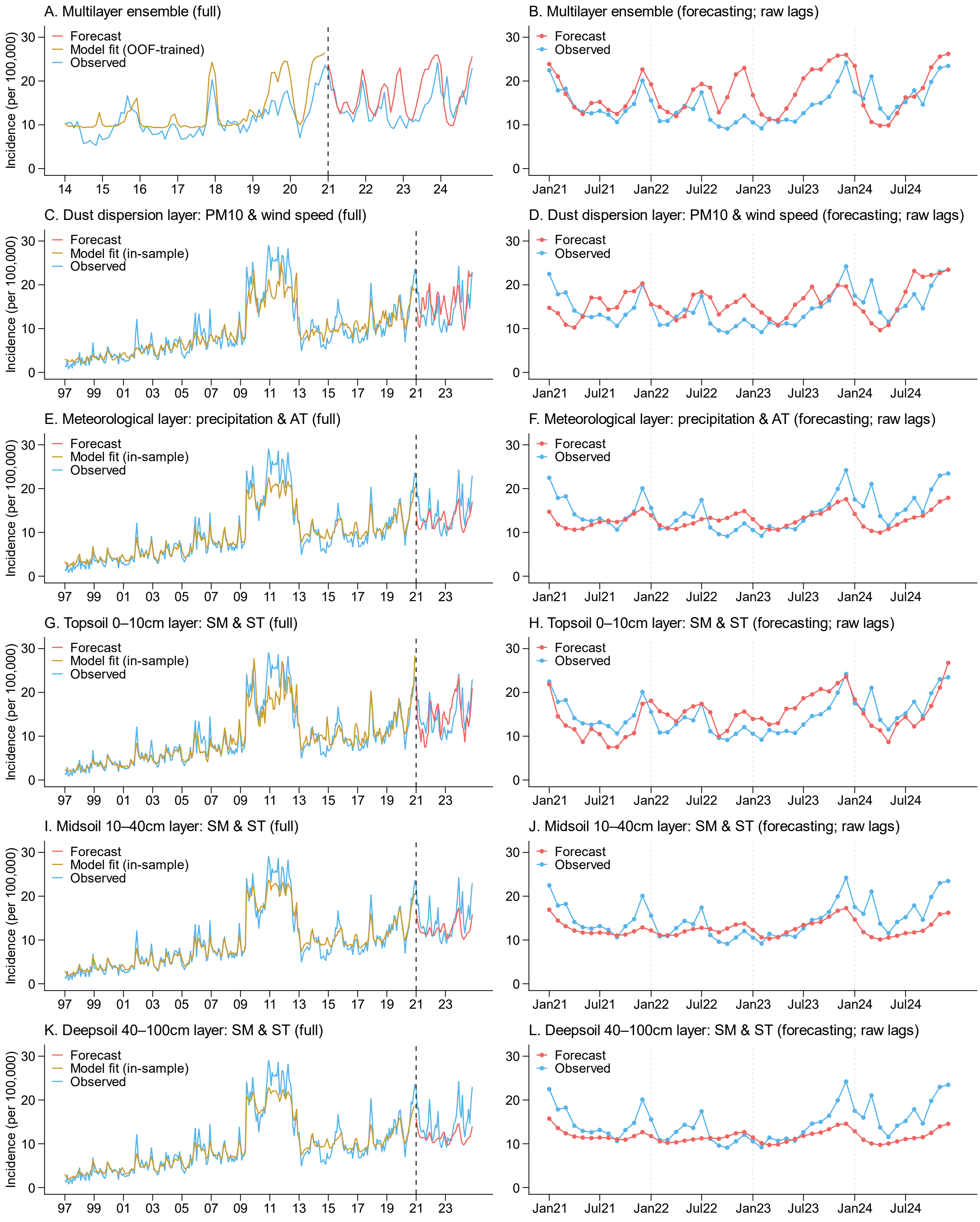

The model structure is identical to Figure 4 but uses raw individual monthly lag values in place of smoothed lag features. The base learners (Stage 1) were trained on 1997–2020, and the meta-learner (Stage 2) was trained on OOF predictions from the base learners during 2014–2020. The model was forecast over a strictly held-out period of 2021–2024. Left panels (A, C, E, G, I, K) show the full study period; right panels (B, D, F, H, J, L) show the forecasting period only. Panels A–B show the multilayer ensemble (meta-learner) combining OOF forecasts from all base learners. Panels C–L show the corresponding Stage 1 base learners (five environmental layers) fitted using raw monthly lag values of environmental predictors: dust dispersion ($PM_{10}$ and wind speed; C–D), meteorological conditions (precipitation and air temperature; E–F), topsoil 0–10 cm (soil moisture and soil temperature; G–H), midsoil 10–40 cm (I–J), and deep soil 40–100 cm (K–L).
